# Whole-genome sequencing identifies variants in *ANK1*, *LRRN1*, *HAS1,* and other genes and regulatory regions for stroke in type 1 diabetes

**DOI:** 10.1101/2022.11.30.22282752

**Authors:** Anni A. Antikainen, Jani K. Haukka, Anmol Kumar, Anna Syreeni, Stefanie Hägg-Holmberg, Anni Ylinen, Elina Kilpeläinen, Anastasia Kytölä, Aarno Palotie, Jukka Putaala, Lena M. Thorn, Valma Harjutsalo, Per-Henrik Groop, Niina Sandholm, the FinnDiane Study Group

## Abstract

**Aims:** Individuals with type 1 diabetes (T1D) carry a markedly increased risk of stroke, with distinct clinical and neuroimaging characteristics as compared to those without diabetes. Using whole-genome sequencing (WGS) and whole-exome sequencing (WES), we aimed to find rare and low-frequency genomic variants associated with stroke in T1D. The lead findings were followed up in various datasets to replicate the findings and to assess their specificity to diabetes.

**Methods and Results:** We studied stroke genetics in 1,051 individuals with T1D using WGS or WES. We analysed the genome with single-variant analyses, gene aggregate analyses, and aggregate analyses on genomic windows, enhancers and promoters. Furthermore, we attempted replication in T1D using a genome-wide association study (N=3,945) and direct genotyping (N=3,600), and in the general population from the FinnGen project and UK Biobank summary statistics. We identified a rare missense mutation on *SREBF1* associated with hemorrhagic stroke (rs114001633, p.Pro227Leu, *p*-value=8.96×10^-9^), which further replicated in T1D. Using gene aggregate analysis with protein altering or protein truncating variants, we identified exome-wide significant genes: *ANK1* and *LRRN1* displayed replication evidence in T1D, while *LRRN1*, *HAS1* and *UACA* replicated in the general population (UK Biobank). Furthermore, we performed sliding-window analyses and identified 14 genome-wide significant windows for stroke on 4q33-34.1, of which two replicated in T1D, and a suggestive genomic window on *LINC01500*, which replicated in T1D. Finally, with the regulatory region aggregate analyses, we identified a stroke-associated *TRPM2-AS* promoter (*p*-value=5.78×10^-6^), which we validated with an in vitro cell-based assay. *TRPM2* has been previously linked to ischemic stroke.

**Conclusions:** Here, we report the first genome-wide analysis on stroke in individuals with diabetes. We identified multiple stroke risk loci with evidence of replication: 4q33-34.1, *SREBF1*, and *ANK1* for stroke in T1D; and *HAS1*, *UACA*, *LRRN1*, *LINC01500*, and *TRPM2-AS* promoter for stroke potentially generalizable to the non-diabetic population.

## 1. Introduction

Stroke is a notable cause of mortality and long-term disability worldwide, with diabetes among the most important risk factors. Of note, the standardized incidence ratio is roughly 3-fold among individuals with type 1 diabetes (T1D) compared to the general population^1^. Furthermore, 537 million adults live with diabetes today and the prevalence is rising (IDF Diabetes Atlas, 2021)^2^. Even though much of this trend is driven by an increase in obesity and insulin-resistant type 2 diabetes (T2D), the incidence of insulin-dependent T1D has increased as well^3^. T1D is a lifelong condition caused by an autoimmune reaction towards the pancreas and treated with daily insulin injections. The strokes themselves may be of hemorrhagic (20%) or ischemic (80%) origin and classified into even more specific subtypes. Interestingly, the two diabetes types affect stroke risk differentially: T1D increases the risk of both ischemic- and hemorrhagic stroke^4, 5^, while the risk imposed by T2D has been estimated more modest for hemorrhagic strokes^5^. Importantly, T1D predisposes individuals towards cerebral small-vessel disease and strokes of microvascular origin^6^. Diabetes causes also other complications, of which diabetic kidney disease (DKD) and severe retinopathy predict cerebrovascular disease in T1D^7^. Understanding stroke pathophysiology in diabetes is important for improving treatment and quality of life for individuals with T1D.

Stroke heritability has been estimated to vary between 30% and 40% in the general population^8^. Stroke heritability varies greatly depending on the subtype, with the largest heritabilities estimated for large artery atherosclerotic stroke and lobar intracranial hemorrhage, and the lowest for small vessel disease^8^. To date, 126 common genomic loci have been associated with stroke with genome-wide significance, although partly lacking external replication: 65 common genomic loci had been associated with stroke, a stroke subtype, or small vessel disease^9^, while recently, a large cross-ancestry genome-wide association study (GWAS) meta-analysis proposed 61 novel genomic loci for stroke^10^. Associations at many of the known common stroke loci overlap with other cardiovascular phenotypes, e.g., coronary artery disease (CAD)^8^. Our previously study suggested a heritable component of stroke in individuals with T1D as a history of maternal stroke was associated with hemorrhagic stroke in T1D^11^. However, very few studies have investigated genetic risk factors for stroke in diabetes^12–14^, and no genome-wide studies in individuals with diabetes yet exist. On the other hand, genetic studies on CAD in diabetes have identified a few diabetes-specific loci^15, 16^, although still pending external replication, and have replicated three known general population CAD risk loci in diabetes: *CDKN2B-AS1*, *PSRC1* and *LPA*^14, 15, 17^.

A substantial proportion of heritability remains unexplained for stroke^8^. Rare genetic variants with minor allele frequency (MAF) of ≤1% may significantly contribute to stroke heritability. In fact, some rare monogenic disorders have stroke as one of their manifestations^8, 9, 18^. In GWASs, the imputation accuracy of rare variants may be limited, and largely depends on the minor allele count (MAC) in the reference sample^19^. Rare variants can be reliably studied with next-generation sequencing-based techniques such as whole-genome sequencing (WGS) and whole-exome sequencing (WES). We have previously used WES to identify protein coding variants associated in lipid and apolipoprotein traits in T1D^20^. In the general population, novel stroke risk loci have been identified with WGS^21^. However, UK Biobank WES analysis for cardiometabolic traits did not discover exome-wide significant stroke risk genes^22^.

Historically, the Finnish population has been isolated and, thus, represents a unique genetic background with enrichment of low-frequency deleterious variants^23^, which may in part enable the discovery of rare disease-associated mutations. Here we studied genetics of stroke and its subtypes with WGS and WES in Finnish individuals with T1D with multiple statistical approaches by focusing on rare and low-frequency genomic variants. We aimed both to find stroke-risk loci specific to individuals with T1D, and to identify risk loci generalizable to the non-diabetic population, since discovery of rare variants is more probable in a high-risk Finnish diabetic population. Finally, we performed cell-based *in vitro* experiments to further validate a discovered promoter region. Altogether, here we report the first genome-wide study on stroke genetics in diabetes.

## 2. Methods

### 2.1 Ethical statement

The study protocol has been approved by the ethics committee of the Helsinki and Uusimaa Hospital District (491/E5/2006, 238/13/03/00/2015, and HUS-3313-2018), and performed in accordance with the Declaration of Helsinki. All participants gave informed consent before participation.

### 2.2 Materials

The study is part of the Finnish Diabetic Nephropathy (FinnDiane) Study^24^. We studied WGS in 571 and WES in 480 non-related individuals with T1D, entailing 112 and 74 stroke cases, respectively (**Table 1**, **Table S1**, **Figure S1** and **S2**). Of note, patient selection from the large FinnDiane cohort to next-generation sequencing were originally designed for DKD. We collected stroke phenotypes from Finnish registries until the end of 2017 (**Table S2**). The identified cases were verified and classified into ischemic- and hemorrhagic strokes by trained neurologists using medical files and brain imaging data. For individuals without data verified by neurologists available (N_WGS_=27, N_WES_=2), we considered only the registry data, and excluded controls with intermediate stroke phenotypes. Importantly, we required stroke to have occurred after T1D diagnosis, and controls to have >35 years of age and >20 years of diabetes duration. We attempted replication in individuals with T1D within the FinnDiane GWAS data set (N=3,945, **Table S3 and S4, Figure S3**), restricted to high imputation quality variants (*r*^2^>0.80), and by directly genotyping twelve variants for replication (N=3,263, **Table S5, Figure S4**). Stroke phenotype within replication in T1D was defined accordingly.

**Table 1:** Clinical characteristics of study participants in the next-generation sequencing data sets.

|  | WES |  |  | WGS |  |  |
| --- | --- | --- | --- | --- | --- | --- |
|  | Cases | Controls | *p*-value | Cases | Controls | *p*-value |
| N | 74 | 406 |  | 112 | 459 |  |
| Hemorrhagic/Ischemic | 22/49 |  |  | 26/64 |  |  |
| CVD death <2017<br>(yes/no, % yes) | 35/39 (47%) | 66/340 (16%) | $2.92 \times 10^{-8}$ | 58/54 (52%) | 83/376<br>(18%) | $2.61 \times 10^{-12}$ |
| Sex (male/female, %<br>males) | 39/35 (53%) | 182/224<br>(45%) | 0.25 | 80/32 (71%) | 236/223<br>(51%) | 0.00013 |
| Age (years) | 48.39 (11.89) | 60.69 (11.13) | $7.08 \times 10^{-13}$ | 51.08<br>(10.29) | 58.70 (9.61) | $3.52 \times 10^{-11}$ |
| T1D duration (years) | 33.6 (10.45) | 47.49 (9.84) | $5.89 \times 10^{-18}$ | 36.16 (9.81) | 46.00 (8.14) | $6.34 \times 10^{-18}$ |
| Calendar year of T1D<br>onset* | 1969.5<br>(11.75) | 1968 (11) | 0.29 | 1968<br>(10.25) | 1969 (9) | 0.80 |
| T1D onset age (years) | 14.79 (6.64) | 13.21 (7.27) | 0.066 | 14.92 (8.93) | 12.69 (7.75) | 0.016 |
| Weighted mean HbA<sub>1c</sub><br>(%)* | 9.00 (2) | 8.51 (1.4) | 0.0037 | 8.86 (1.76) | 8.34 (1.57) | 0.00097 |
| HbA<sub>1c</sub> count* | 19 (28.5) | 29 (26) | 0.0038 | 19 (30) | 29 (30) | 0.011 |
| DKD (yes/no, yes-%) | 52/22 (70%) | 201/205<br>(50%) | 0.00098 | 87/25 (78%) | 206/253<br>(45%) | $2.49 \times 10^{-10}$ |

*Weighted mean HbA<sub>1c</sub> is calculated until the stroke event or the end of follow-up. DKD: diabetic kidney disease, defined as end-stage renal disease (ESRD), macro- or microalbuminuria. Mean (standard deviation, SD), \*Median (interquartile range, IQR). Student's t-test, Wilcoxon signed rank test or Fisher's exact test.*

### 2.3 Study Design

We performed single variant and variant aggregate analyses by meta-analysing WES and WGS whenever possible (**Figure 1**); and conducted stroke subtype association analyses for the lead findings. In addition, we conducted gene aggregate analyses with the minimal adjustment separately with protein altering variants (PAVs) and protein truncating variants (PTVs); and repeated the analyses with an additional DKD adjustment. Finally, we conducted minimally adjusted intergenic aggregate analyses within genomic windows by statistically up-weighting functionally important and rare variants; and within established enhancers and promoters by weighting variants according to rarity.

**Figure 1:**
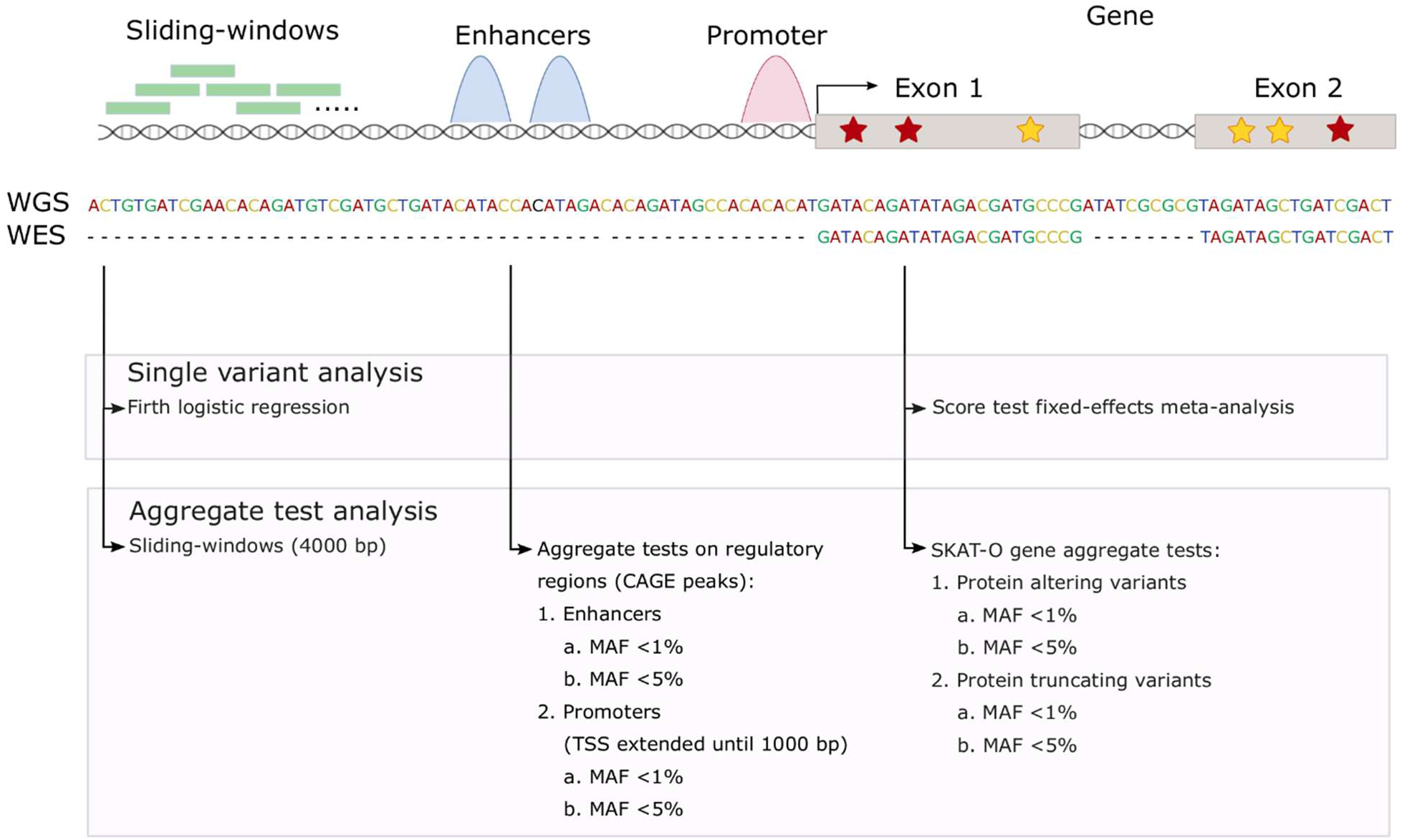
Study design.

### 2.4 Single variant analyses

First, we analyzed the genome with an additive inheritance model. For variants available in WES and WGS data, we performed score test fixed-effect inverse variance based meta-analysis (MAC≥5, WES and WGS: MAC≥2) using rvtests (version 20190205)^25^ and metal (version 20110325)^26^. For variants available only in one data set we utilized Firth regression (MAC≥5)^25^. Recessive variants can have high mutation severity in comparison to e.g., autosomal dominant variants^27^. Thus, we further analyzed the genome with a recessive inheritance model by exploiting a similar scheme (total homozygote minor allele carriers ≥5; WES and WGS homozygote carriers ≥2). Autosomal recessive Firth regression was performed with plink2 (version 20210420)^28^. The additive and recessive single variant analyses were adjusted for the calendar year of diabetes onset, sex, and two first genomic data principal components (i.e., minimal adjustment setting), and additionally for DKD.

### 2.5 Gene aggregate analyses

In order to improve statistical power for rare (MAF≤1%) and low-frequency (MAF≤5%) variants, we performed gene aggregate analyses with an optimal unified sequence kernel association test (SKAT-O) meta-analysis with MetaSKAT (version 0.81)^29^, separately within two distinct classes (**Table S6**): protein altering variants and protein truncating variants i.e., the more severe putative loss-of-function variants^30^. Only variable sites (MAC≥1) were accepted into gene aggregate analysis, and the aggregate tests were required to entail at least two variants (N_variant_≥2), with a cumulative MAC (CMAC) ≥5. Multiple testing correction, based on number of tested genes, resulted in significance thresholds of *p*-value<4×10^-6^ for PAVs (MAF≤1% and MAF≤5%), *p*-value<7×10^-5^ for PTVs with MAF≤1%, and *p*-value<5×10^-5^ for PTVs with MAF≤5%. For the known Mendelian stroke risk genes^18^, we report results regardless of variant number or CMAC.

### 2.6 Sliding-window and regulatory region aggregate analyses with whole-genome sequencing

To increase statistical power for low-frequency and rare variants on intergenic regions, we performed minimally adjusted and functionally informed sliding-window analyses, i.e., aggregate analyses within 4,000 base pair (bp) regions – separated by 2,000 bps – with variants statistically weighted according to rarity and functional importance using STAAR-O (STAAR R package 0.9.6)^31, 32^. Functional importance was defined with Combined Annotation-Dependent Depletion (CADD) data^32^ using variant MAF (to up-weight rarer variants), pre-computed CADD score, and the first annotation principal component from seven annotation classes (**Figure S5**, **Table S7**), calculated following guidelines^31^.

With the minimal adjustment setting, we studied established regulatory regions i.e., enhancers and promoters (N_variant_≥2, CMAC≥5), as defined in FANTOM5 cap analysis of gene expression (CAGE) human data^33^, with promoters defined as the transcription start site (TSS) extended to 1,000 bp. However, we utilized only allele frequencies as variant annotation, allowing us to include more variants. With low-frequency variants, multiple testing corrected significance thresholds were *p*-value<2.9×10^-7^ and *p*-value<2.6×10^-6^ for promoters and enhancers, respectively. For rare variants, the thresholds were *p*-value<3.5×10^-7^ and *p*-value<4.3×10^-6^, respectively.

### 2.4 Replication

Within the FinnDiane GWAS data, we attempted replication of genetic variants (rvtests 20190205^25^) and had good statistical power (>80%) to detect a nominal association with an odds ratio (OR) ≥2.5 for additive low-frequency variants (MAF=1%) (**Figure S6**)^34^. However, for rare variants with MAF=0.1% and OR<9, we had only limited power to detect an association even with nominal significance (*p*-value<0.05). We attempted direct genotyping for replication for twelve variants, although minor allele carriers were observed only for seven of them (**Table S8**). Most variants within the aggregate discoveries were rare or ultra-rare (MAF≈0.1%), making replication with imputed genomic data problematic. Nevertheless, we attempted replication within the FinnDiane GWAS data by including also the directly genotyped variants (SKAT-O, STAAR-O). We performed SKAT-O using GMMAT R package 1.3.2 by imputing missing genotypes to mean^35^, while intergenic aggregate analyses were performed similarly with STAAR R package^31^. Of note, relatedness in replication was accounted for with relatedness matrices instead of genomic principal components^25, 36^. We attempted replication in the general population for genetic variants from the large-scale population-wide FinnGen project GWAS data (https://www.finngen.fi/en) (**Table S9**), and for the gene aggregate discoveries from UK Biobank summary statistics^22, 37^.

### 2.5 Detailed Materials and Methods

Detailed Materials and Methods are available in the Online Supplemental Material.

## 3. Results

### 3.1 Single variant analyses

We sought for genetic variants associated with stroke using WES and WGS data, and discovered a suggestive stroke-risk locus, 4q33-34.1, with minimally adjusted additive inheritance model (4:170787127, *p*-value=8.83×10^-8^, MAF=3.7%, **Table 2**, **Figure S7**). We attempted replication, however, the variant 4:170787127 was unavailable for replication in the T1D specific GWAS and in the FinnGen general population GWAS summary statistics. A variant with the third lowest *p*-value on 4q33-34.1 did not replicate in T1D nor the general population (**Table 2**).

**Table 2:**
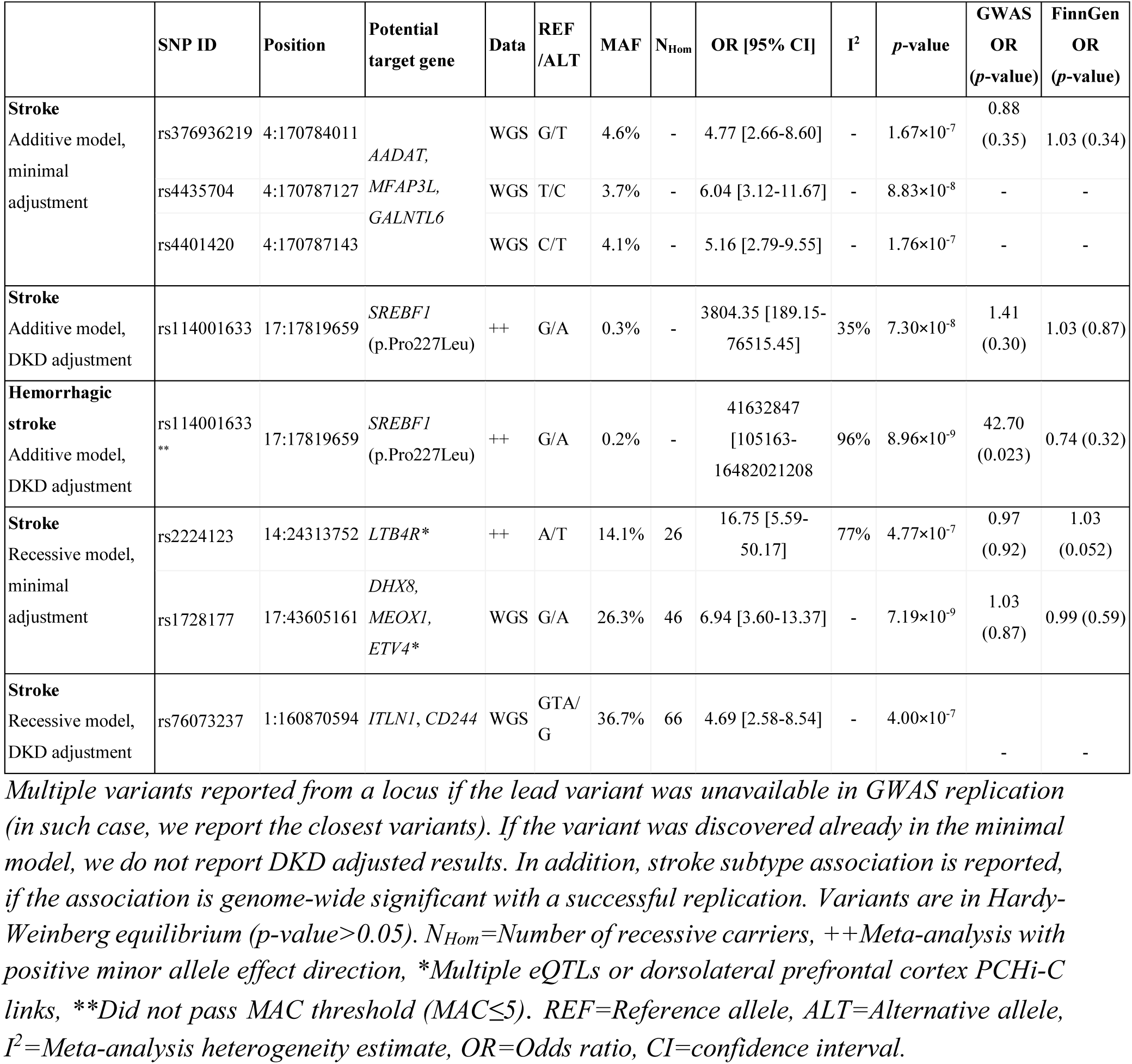
Lead variants discovered with single variant association analyses.

|  | SNP ID | Position | Potential target gene | Data | REF /ALT | MAF | N<sub>Hom</sub> | OR [95% CI] | I<sup>2</sup> | p-value | GWAS OR (p-value) | FinnGen OR (p-value) |
| --- | --- | --- | --- | --- | --- | --- | --- | --- | --- | --- | --- | --- |
| **Stroke**<br>Additive model,<br>minimal<br>adjustment | rs376936219 | 4:170784011 | *AADAT*,<br>*MFAP3L*,<br>*GALNTL6* | WGS | G/T | 4.6% | - | 4.77 [2.66-8.60] | - | 1.67×10<sup>-7</sup> | 0.88<br>(0.35) | 1.03 (0.34) |
|  | rs4435704 | 4:170787127 |  | WGS | T/C | 3.7% | - | 6.04 [3.12-11.67] | - | 8.83×10<sup>-8</sup> | - | - |
|  | rs4401420 | 4:170787143 |  | WGS | C/T | 4.1% | - | 5.16 [2.79-9.55] | - | 1.76×10<sup>-7</sup> | - | - |
| **Stroke**<br>Additive model,<br>DKD adjustment | rs114001633 | 17:17819659 | *SREBF1*<br>(p.Pro227Leu) | ++ | G/A | 0.3% | - | 3804.35 [189.15-76515.45] | 35% | 7.30×10<sup>-8</sup> | 1.41<br>(0.30) | 1.03 (0.87) |
| **Hemorrhagic stroke**<br>Additive model,<br>DKD adjustment | rs114001633<br>** | 17:17819659 | *SREBF1*<br>(p.Pro227Leu) | ++ | G/A | 0.2% | - | 41632847<br>[105163-16482021208] | 96% | 8.96×10<sup>-9</sup> | 42.70<br>(0.023) | 0.74 (0.32) |
| **Stroke**<br>Recessive model,<br>minimal<br>adjustment | rs2224123 | 14:24313752 | *LTB4R\** | ++ | A/T | 14.1% | 26 | 16.75 [5.59-50.17] | 77% | 4.77×10<sup>-7</sup> | 0.97<br>(0.92) | 1.03<br>(0.052) |
|  | rs1728177 | 17:43605161 | *DHX8*,<br>*MEOX1*,<br>*ETV4\** | WGS | G/A | 26.3% | 46 | 6.94 [3.60-13.37] | - | 7.19×10<sup>-9</sup> | 1.03<br>(0.87) | 0.99 (0.59) |
| **Stroke**<br>Recessive model,<br>DKD adjustment | rs76073237 | 1:160870594 | *ITLN1*, *CD244* | WGS | GTA/<br>G | 36.7% | 66 | 4.69 [2.58-8.54] | - | 4.00×10<sup>-7</sup> | - | - |

Multiple variants reported from a locus if the lead variant was unavailable in GWAS replication (in such case, we report the closest variants). If the variant was discovered already in the minimal model, we do not report DKD adjusted results. In addition, stroke subtype association is reported, if the association is genome-wide significant with a successful replication. Variants are in Hardy-Weinberg equilibrium ( $p\text{-value} > 0.05$ ). $N_{Hom}$ =Number of recessive carriers, ++Meta-analysis with positive minor allele effect direction, \*Multiple eQTLs or dorsolateral prefrontal cortex PCHi-C links, \*\*Did not pass MAC threshold ( $MAC \leq 5$ ). REF=Reference allele, ALT=Alternative allele, $I^2$ =Meta-analysis heterogeneity estimate, OR=Odds ratio, CI=confidence interval.

As DKD is a common diabetic complication that has been reported to predict incident stroke in T1D^7^, we performed additional analyses adjusted for DKD, and discovered a rare missense mutation on *SREBF1* exome-wide significantly (*p*-value<3×10^-7^) associated with stroke (rs114001633, p.Pro227Leu, *p*-value=7.30×10^-8^, MAF=0.26%) (**Table 2**, **Figure S8**). In the stroke subtype analysis for the lead findings, this variant was genome-wide significant for hemorrhagic stroke (*p*-value=8.96×10^-9^, MAC=3, **Table 2**, **Table S10**). However, rs114001633 did not pass the MAC threshold in the stroke subtype analysis (MAC≥5), thus, the result must be interpreted with caution. Due to the rarity of the variant, we performed additional genotyping for replication, whereby the variant replicated for hemorrhagic stroke (*p*-value=0.02, N=3,263).

We further considered a recessive inheritance model and identified a genome-wide significant variant on *DHX8* intron with the minimal adjustment (rs1728177, *p*-value=7.19×10^-9^, MAF=26%) (**Table 2**, **Figure S9**), which however did not replicate. Furthermore, we found two suggestive recessive loci, including one on 5’ untranslated region of *LTB4R* (rs2224123, *p*-value=4.77×10^-7^, MAF=14%). rs2224123 did not replicate in T1D but was borderline significant in the general population with an additive model (FinnGen GWAS: *p*-value=0.052). In the sequencing data, significance with an additive model was nominal (*p*-value=0.019), suggesting that rs2224123 acts recessively, but recessive data were not available for the replication look-up in the general population.

### 3.2 Gene aggregate analyses

To improve statistical power for rare and low-frequency variants, we performed the gene aggregate analyses. In the minimally adjusted model, low-frequency PAVs on *ANK1* were significantly associated with stroke (*p*-value=2.23×10^-6^, CMAC=247), and even more strongly with ischemic stroke (*p*-value=1.31×10^-6^, CMAC=225) (**Figure 2A**, **Figure S10, Tables S11** and **S12**). Furthermore, the aggregate of low-frequency or rare PAVs was suggestively associated with stroke in nine genes (**Figure 2A**). Of these, *TARBP2* associated significantly with ischemic stroke (*p*-value=1.71×10^-7^, CMAC=5, MAF≤1%), and *CLEC4M* with hemorrhagic stroke (*p*-value=4.74×10^-15^, CMAC=11, MAF≤1%).

**Figure 2:**
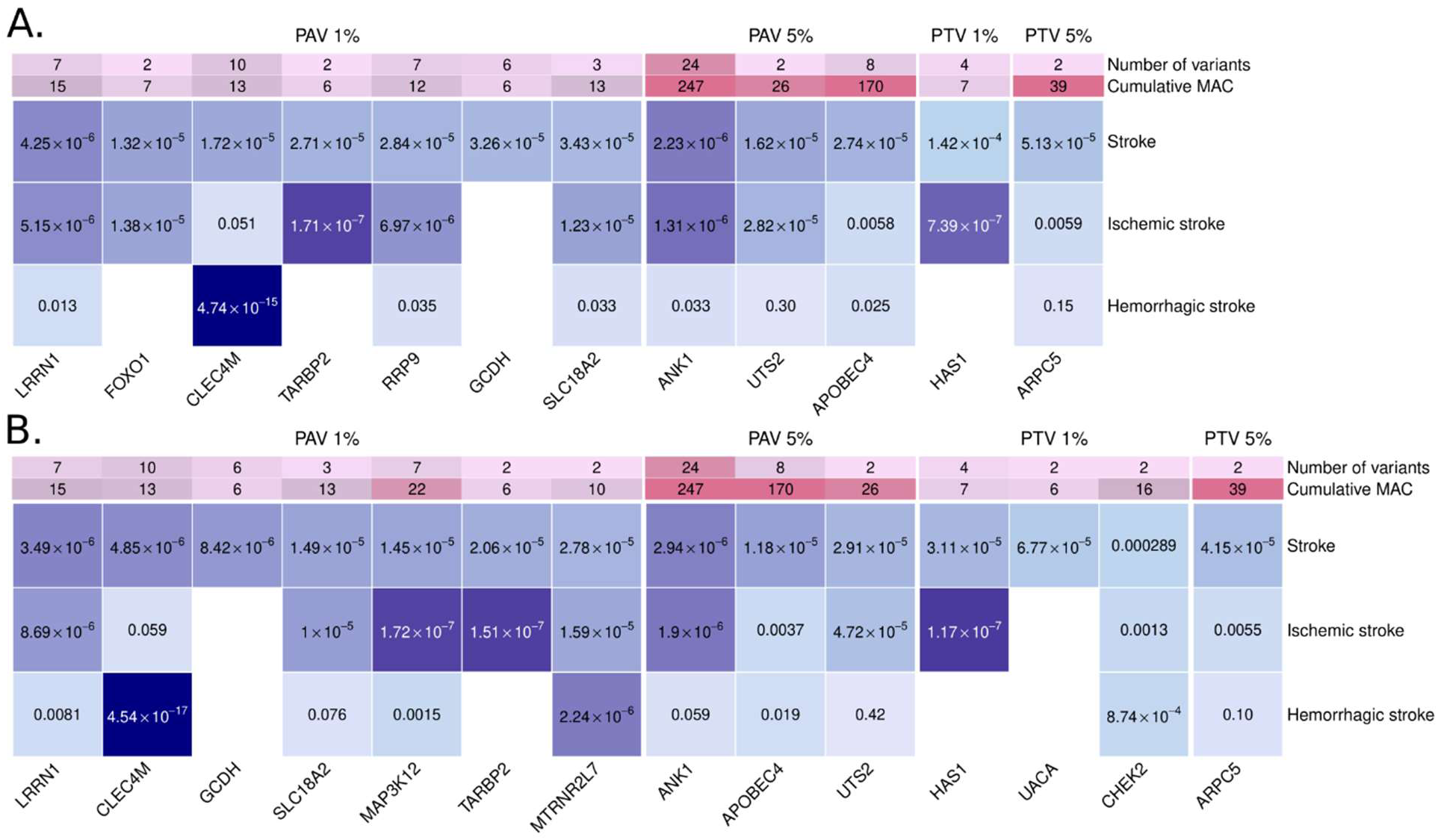
Discovered genes with the SKAT-O gene aggregate tests. **A.** Minimal adjustment, **B.** Additional adjustment for diabetic kidney disease. The color indicates the -log_10_(*p*-value), with darker color indicating more significant finding. Only the rare variant model (MAF≤1%) is reported, if no low-frequency variants (1%<MAF≤5%) were available in the gene. Bonferroni corrected significance thresholds: 4×10^-6^ (protein altering variant, PAV≤1%), 4×10^-6^ (PAV≤5%), 5×10^-5^ (protein truncating variant, PTV≤5%), and 7×10^-5^ (PTV≤1%). Number of variants and CMAC given based on the combined stroke phenotype.

After additional DKD adjustment, rare PAVs in *LRRN1* were significantly associated with stroke (*p*-value=3.49×10^-6^, CMAC=15), more strongly with ischemic stroke (*p*-value=8.69×10^-6^, CMAC=12; **Figure 2B**, **Figure S11**, **Tables S11** and **S13**). Furthermore, we identified suggestive genes, missed in the minimally adjusted model: *MAP3K12* and *MTRNR2L7*. In the stroke subtype analysis, rare PAVs in *MAP3K12* were significantly associated with ischemic stroke (*p*-value=1.72×10^-7^, CMAC=17), and in *MTRNR2L7* with hemorrhagic stroke (*p*-value=2.24×10^-6^, CMAC=6). *MAP3K12* and *TARBP2* are located close to each other on the genome, thus, they may represent the same association signal through linkage disequilibrium (LD) or modifier effects onto the causal gene (**Figure S12**).

The aggregate of PTVs was suggestively associated with stroke in two genes including hyaluronan synthase 1 (*HAS1;* **Figure 2A**, **Table S12**). In analysis for stroke subtypes, *HAS1* was significantly associated with ischemic stroke (*p*-value=7.39×10^-7^, CMAC=7). With additional DKD adjustment, rare PTVs in *HAS1* (*p*-value=3.11×10^-5^) and *UACA* (*p*-value=6.77×10^-5^, CMAC=6), and low-frequency PTVs in *ARPC5* (*p*-value=4.15×10^-5^, CMAC=39), were significantly associated with stroke (**Figure 2B**, **Table S13**).

### 3.3 Replication of gene aggregate findings

We attempted T1D specific replication within the FinnDiane GWAS data, by including also five directly genotyped variants, using the gene aggregate approach and by inspecting the exonic variants individually. Despite the uncertainty of genotype imputation and our limited statistical power for rare variants, *ANK1* and *LRRN1* showcased weak evidence of replication in T1D: Although *ANK1* did not reach significance with SKAT-O (**Table S14**), one of the available fifteen variants was significant for stroke (rs779805849, *p*-value=0.01) (**Table 3**, **Figure 3**), and two additional variants replicated for hemorrhagic stroke (rs146416859 and rs61753679, *p*-value<0.05) (**Tables S12**). *LRRN1* did not replicate in FinnDiane with rare PAVs (*p*-value=0.50, N_variant_=4) (**Table S15**). However, when we extended the model to low-frequency PAVs (**Table S15**), thus improved statistical power and imputation quality, *LRRN1* replicated for ischemic stroke (*p*-value=0.039, N_variant_=6). *UACA* contained two rare PTVs associated with stroke, of which one replicated through genotyping (*p*-value=0.0030, **Table S13**). However, the variant was ultra-rare, and replication thus uncertain. We were unable to replicate *HAS1* in T1D due to missing data; we directly genotyped one variant but found no rare allele carriers.

**Table 3.**
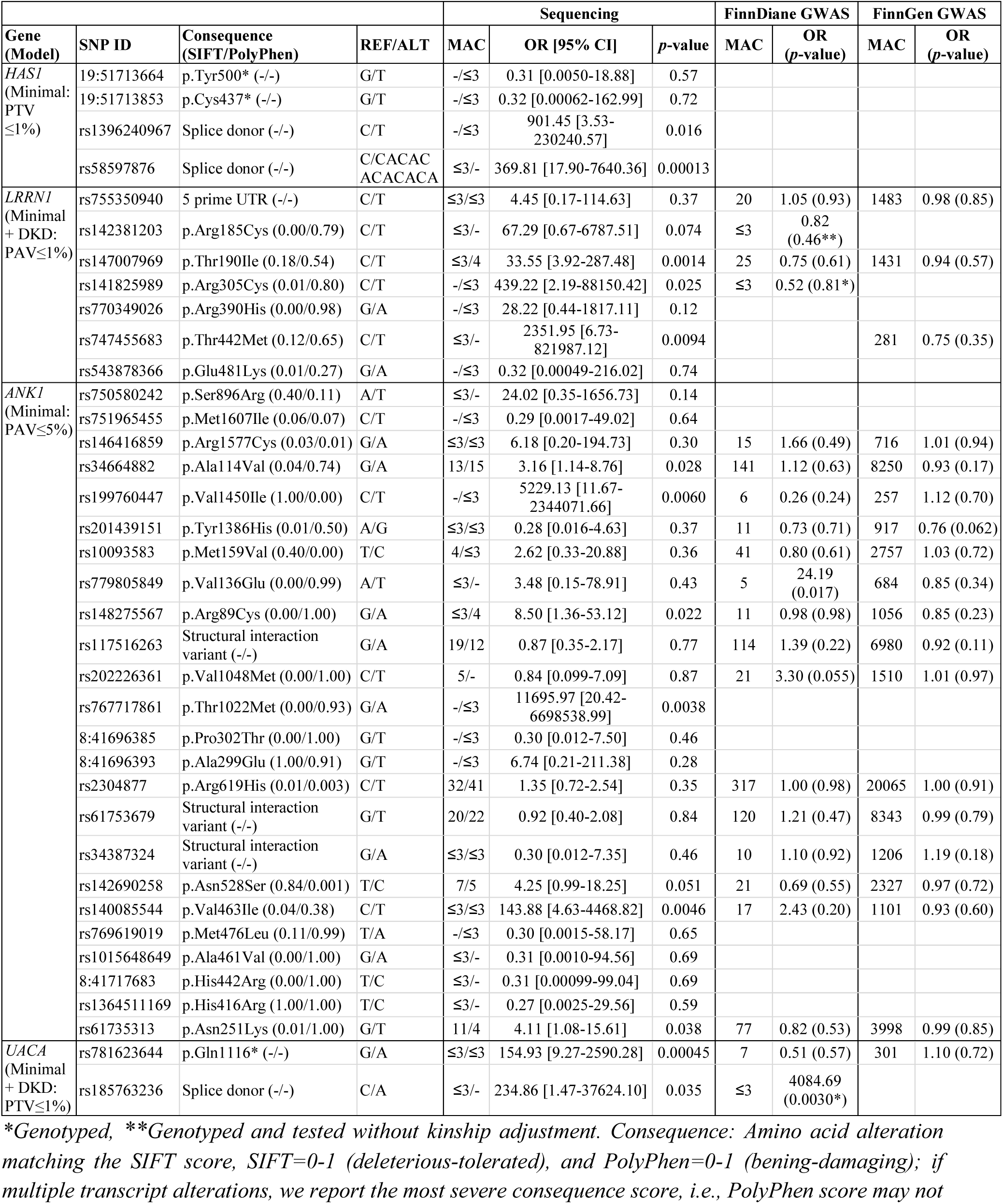
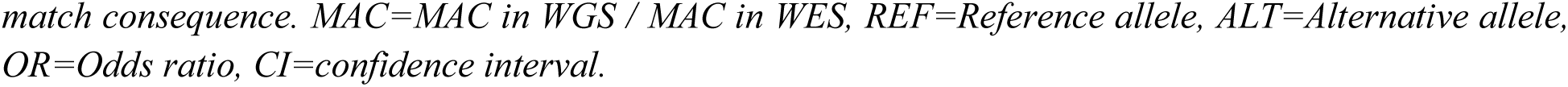
Protein altering- and protein truncating variants in significant stroke risk genes according to significance thresholds corrected by the number of genes within designated models.

**Figure 3:**
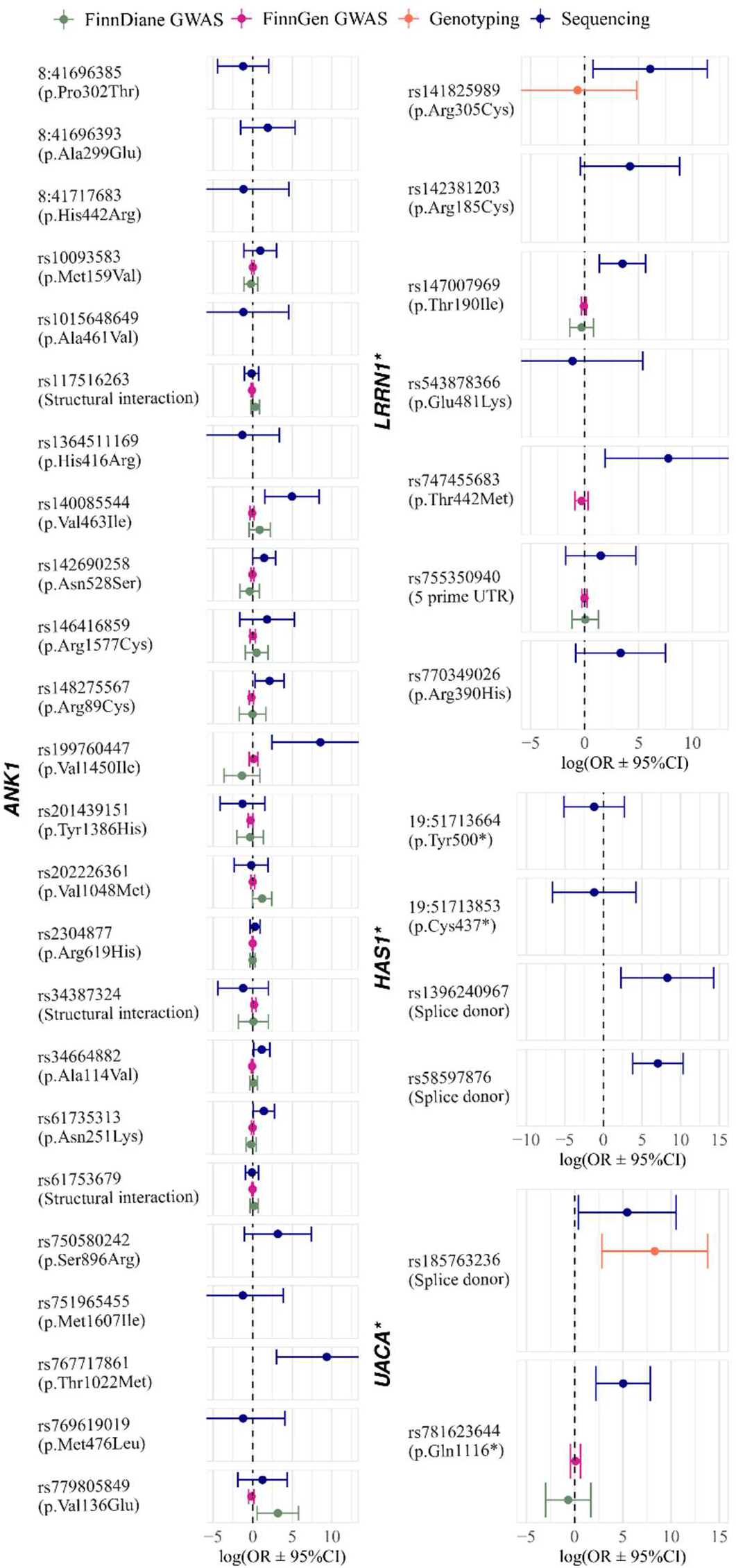
Variants in significant stroke-associated genes after multiple testing correction (SKAT-O) showcasing evidence of replication. *Diabetic kidney disease adjusted stroke model.

We further attempted replication in the general population by look-ups from two UK Biobank WES studies^22, 37^ (**Tables S16** and **S17**). Importantly, *HAS1* replicated for stroke in both studies with ultra-rare loss-of-function variants (MAF≤0.1%: Jurgens et al. *p*-value=0.039^22^; Backman et al. *p*-value=0.035^37^), while *UACA* replicated only in the latter study (MAF≤0.01%: Backman et al. *p*-value=0.035^37^). Finally, *LRRN1* replicated for stroke with an ultra-rare missense variant model (MAF≤0.001%: Backman et al. *p*-value=0.026^37^), although not for ischemic stroke. Of note, *ANK1* did not replicate in the general population.

Out of the suggestive genes, *FOXO1*, *TARBP2*, and *MAP3K12* showcased weak replication in T1D (**Tables S12, S13, S14** and **S15**). One variant within *FOXO1* replicated for hemorrhagic stroke (*p*-value=0.012), two within *MAP3K12* for hemorrhagic stroke (*p*-value=0.013); and *TARBP2* replicated for hemorrhagic stroke with SKAT-O (*p*-value=2.59×10^-4^). UK Biobank general population gene burden WES analysis look-ups supported stroke associations for *UTS2*, *MAP3K12*, and *FOXO1* (**Table S17**)^22, 37^.

### 3.4 Known Mendelian stroke genes in T1D

Mutations on Mendelian stroke risk genes may for instance cause small vessel disease or cerebral cavernous malformations, which can eventually lead to stroke^8^. We inspected the association of 17 autosomal genes previously linked to stroke through nonsynonymous mutations^18^ (**Figure S13**). Rare PAVs on *KRIT1* associated with stroke (*p*-value=0.018) and ischemic stroke (*p*-value=0.0092). Furthermore, rare PAVs on *ADA2* and on *TREX1* associated with hemorrhagic stroke (*p*-value=0.027 and *p*-value=0.010, respectively). Loss-of-function mutations on *KRIT1* cause vascular malformations, while *ADA2* has been linked to autoinflammatory small vessel vasculitis and *TREX1* to small vessel disease^8, 18^.

### 3.5 Sliding window analyses

To increase statistical power for low-frequency and rare variants on non-coding regulatory regions, we performed genome-wide sliding-window aggregate analyses. We found further evidence for the 4q33-34.1 genomic region as we discovered fourteen windows within the region, with a genome-wide significant association between an aggregate of low-frequency variants and stroke (MAF≤5%; **Figure 4A**, **Table S18**). Importantly, two of these windows (4:170782001-170786000, *p*-value=3.40×10^-8^, CMAC=934; and 4:170784001-170786000, *p*-value=1.10×10^-8^, CMAC=1190) and ten individual variants within the 4q33-34.1 genomic region replicated for stroke in T1D (FinnDiane GWAS: *p*-value<0.05; **Table S19**). To identify the most likely effector genes for the 4q33-34.1, we inspected variant expression quantitative trait loci (eQTL) from GTEx Portal and eQTLGen Consortium^38^, and functional genomics from the 3D Genome Browser^39^. 4q33-34.1 is located in the same topologically associating domain with distal promoters of *GALNTL6*, *MFAP3L* and *AADAT* in the frontal lobe and hippocampus (**Figure S14**). In addition, promoter capture high-throughput chromosome conformation capture (PCHi-C) links could be identified for a few individual variants, e.g., for *GALNTL6* in the hippocampus, and *AADAT* and *MFAP3L* in the dorsolateral prefrontal cortex. Several variants were eQTLs of *LINC02431* in testis, and one variant was an eQTL of *AADAT* in esophagus (normalized effect size [NES] =-0.55).

**Figure 4:**
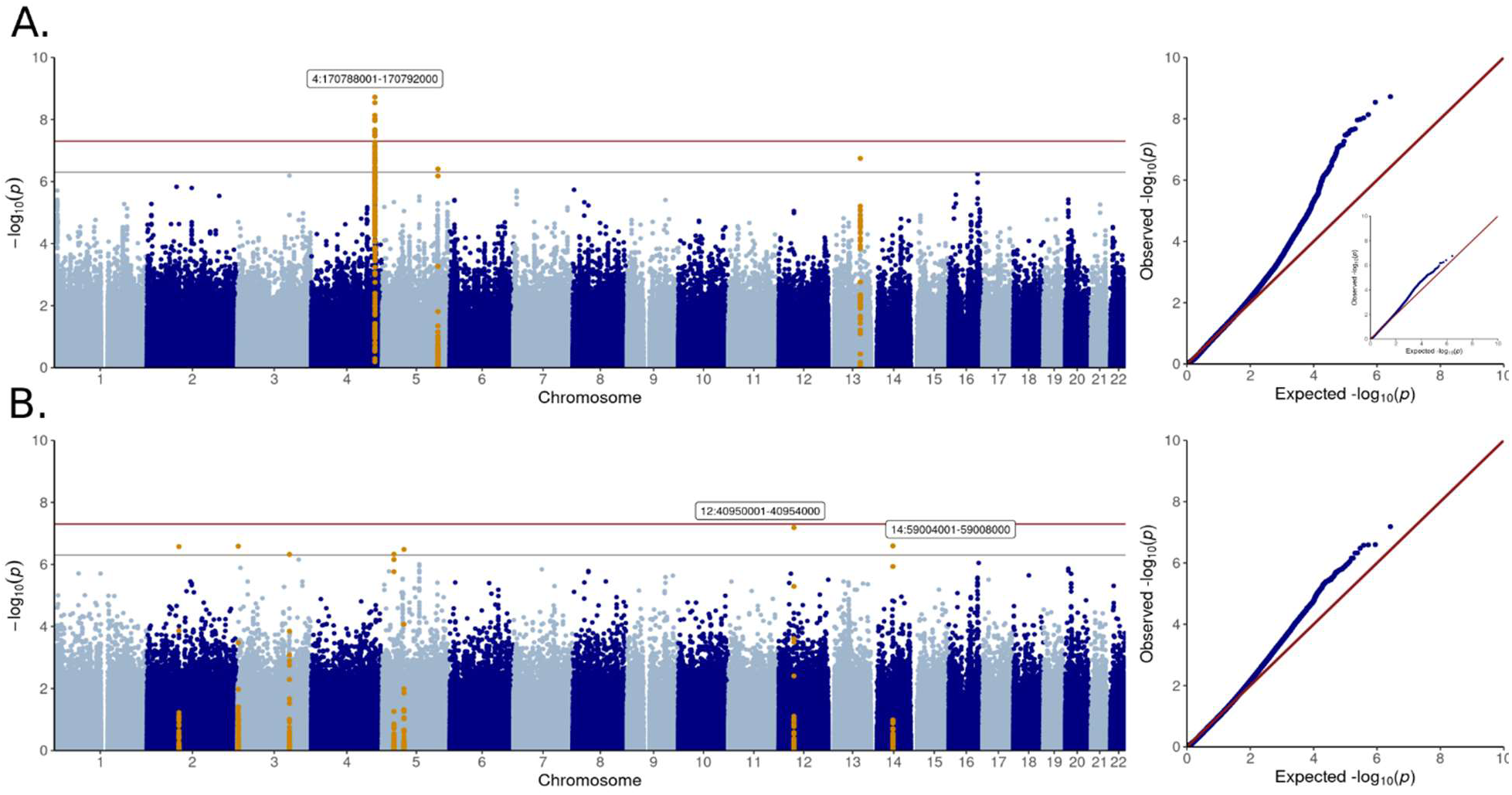
STAAR sliding-window analyses. **A.** MAF≤5% (inserted QQ-plot without the 4q33-34.1 region), and **B.** MAF≤1%.

When we inspected rare variants (MAF≤1%), we discovered multiple suggestive windows, e.g., close to or within the *CNTN1*, *CNTN4*, *LINC01500*, and *TGOLN2* genes (**Figure 4B**, **Table S18**). In stroke subtype analysis, the *CNTN1* window was genome-wide significant for hemorrhagic stroke (12:40950001-40954000: *p*-value=2.10×10^-8^, CMAC=24). Interestingly, *CNTN1* and *CNTN4* are located on different chromosomes, but belong to the same contactin protein family; however, replication is pending. The suggestive window near *LINC01500* (14:59004001-59008000: *p*-value=2.53×10^-7^, CMAC=19) replicated for stroke in T1D (FinnDiane GWAS: *p*-value=0.01, CMAC=24). Four variants within the window were available in the FinnGen general population GWAS, and one replicated (rs1281241634, *p*-value=0.03) (**Table S19**). According to PCHi-C, the *LINC01500* intronic window looped to the *DACT1* promoter on the dorsolateral prefrontal cortex (**Figure S15**). Finally, the *TGOLN2* window replicated for hemorrhagic stroke in T1D (FinnDiane GWAS: *p*-value=0.037).

### 3.6 Promoters and enhancers

As a more targeted approach to explore the non-coding genome, we studied rare and low-frequency variants on established regulatory regions. We discovered three enhancers with suggestive stroke-associated enrichment of rare or low-frequency variants within intronic regions of *TRPM3*, *LOC105378983,* and *BDNF*, encoding brain-derived neurotrophic factor (**Tables S20** and **S21, Figure S16**). The *BDNF* enhancer was significant after multiple testing correction for ischemic stroke (*p*-value=1.01×10^-6^). Regional aggregate replications were not possible in the T1D specific GWAS (N_variant_<2), and individual variants were missing or did not replicate. PCHi-C linked the *BDNF* enhancer to its promoter on specific brain regions (**Figure S15**).

We did not identify stroke-associated promoters after multiple testing correction (*p*-value<3×10^-7^, **Figure S17**). The strongest associations were two *TGOLN2* promoters (*p*-value=5.60×10^-6^, CMAC=9, MAF≤1%), located on the previously mentioned *TGOLN2* window, and a *TRPM2-AS* promoter (*p*-value=5.78×10^-6^, CMAC=33, MAF≤1%; **Tables S22** and **S23**). *TGOLN2* promoters did not replicate in T1D. *TRPM2-AS* promoter nearly replicated in T1D (FinnDiane GWAS: *p*-value=0.053). When we inspected variants individually, one out of nine available variants replicated in the general population for ischemic stroke (FinnGen GWAS: *p*-value=0.038). In GTEx, rs762428 within the *TRPM2-AS* promoter associated significantly to *TRPM2* level in whole blood (NES=-0.63) and lungs (NES=-0.41, *p*<0.001), also nominally in other tissues such as the hypothalamus (NES=-0.42). *TRPM2* encodes a calcium-permeable and non-selective cation channel expressed mainly in the brain. The gene has been linked to ischemic stroke^40^, and belongs to the same protein subfamily as the above mentioned *TRPM3*. *TRPM2* inhibitors have been proposed as a drug target for central nervous system diseases^41^, thus, our results suggested that these inhibitors could be beneficial also for stroke in T1D.

Our cell-based experimental research on the *TRPM2-AS* promoter detected *TRPM2-AS* and *TRPM2* transcripts specifically in HELA cells among tested cell-lines (**Figure S18**). Luciferase promoter analysis of the *TRPM2-AS* promoter in HELA cells indicated strong promoter activity; although, the most strongly stroke-associated variant, rs753589764, did not significantly affect luciferase activity under normal cell culture conditions (*p*-value=0.27, 22 technical repeats). However, we cannot rule out a variant effect under cellular stress, e.g., oxidative stress.

## 4. Discussion

Stroke heritability has been estimated to range between 30% and 40%, but the genomic loci identified thus far explain only a small fraction of heritability^8^. One potential explanation underlying the missing heritability are rare variants missed by GWAS. Therefore, we performed WES and WGS in a total of 1,051 Finnish individuals with T1D to discover rare and low-frequency variants associated with stroke and its major subtypes, either specific for T1D, or generalizable to the non-diabetic population. We identified multiple significant loci with evidence of replication, including protein altering or truncating variants on *ANK1*, *HAS1*, *UACA*, and *LRRN1*, as well as a 4q33-34.1 intergenic region.

With single variant analyses, we identified a missense mutation on *SREBF1* (rs114001633, p.Pro227Leu) which was genome-wide significantly associated with hemorrhagic stroke, and further replicated. As the variant was ultra-rare, and we had a relatively small number of hemorrhagic stroke cases, further replication is needed in T1D to conform this finding. *SREBF1* encodes a transcription factor involved in lipid metabolism and insulin signaling^42^.

Gene aggregate tests (SKAT-O) detected four genes with significant stroke-associated burden of PAVs (*ANK1* and *LRRN1*) or PTVs (*HAS1* and *UACA*) and evidence of replication; *LRRN1*, *HAS1*, and *UACA* after adjustment for DKD. *ANK1* did not replicate in T1D with SKAT-O, however, one out of the fifteen available variants replicated for stroke in T1D (rs779805849, p.Val136Glu). Of note, SIFT and PolyPhen predicted many *ANK1* variants as deleterious^43, 44^. *ANK1* encodes ankyrin-1, within which mutations cause hereditary spherocytosis^45^. Previous genome-wide association studies have linked the gene to T2D^46^, while another gene from the ankyrin protein family, *ANK2*, is a previously identified stroke risk locus^47^.

Rare PAVs in *LRRN1* associated with ischemic stroke. *LRRN1* did not replicate with the corresponding model in T1D, however; with a model extended to low-frequency PAVs, *LRRN1* replicated for ischemic stroke. Rare variant replication is problematic with GWAS data due to the uncertainty of the imputation, which may explain the need of increasing the allele frequency threshold to observe a successful replication. Furthermore, *LRRN1* replicated with ultra-rare variants in the general population^37^. *LRRN1* encodes leucine rich repeat neuronal protein 1, with a brain-enriched expression profile.

*HAS1* consistently replicated in the general population^22, 37^, while *UACA* replicated in one study with ultra-rare variants^37^. *HAS1* encodes an enzyme producing hyaluronan and with expression induced by inflammation and glycemic stress^48^. Of note, an increased hyaluronan turnover has been suggested to follow ischemic stroke^49^. *HAS1* replication was not feasible in T1D due to absent mutation carriers, thus, a diabetes-specific replication is pending. Nevertheless, *HAS1* PTVs may be of particular importance in T1D, as dysregulation of endothelial glycocalyx hyaluronan has been suggested to contribute to diabetic complications^50^. Finally, it must be noted that PTVs have not been functionally confirmed as loss-of-function, but the annotations are predictions; PTV at the beginning of a gene is likely more severe than at the end, and in fact, PTVs closer to the *HAS1* transcription start site were more strongly associated with stroke.

To increase statistical power on regulatory regions, we performed statistical aggregate tests in genomic windows, enhancers and promoters^31, 33^. Of note, we extended genomic window length from the default to increase statistical power, which however also reduced precision as the causal region might be narrower. We found fourteen genome-wide significant stroke-associated windows with low-frequency variants on 4q33-34.1, of which two replicated for stroke in T1D. According to eQTLs and PCHi-C interactions, 4q33-34.1 variants most likely target *GALNTL6*, *MFAP3L* or *AADAT*. We also discovered a suggestive stroke-associated window within *LINC01500*, which replicated for stroke in T1D. According to PCHi-C, the *LINC01500* window targets a promoter of *DACT1*. Finally, we identified a suggestive stroke-associated promoter of *TRPM2-AS*, which nearly replicated in T1D (*p*-value=0.053). Importantly, transient receptor melastatin 2 (*TRPM2*) has been previously associated with ischemic stroke^40, 41^. Our functional cell-based assay validated the *TRPM2-AS* region promoter activity. However, the most strongly stroke-associated variant, rs753589764, did not associate with *TRPM2-AS* promoter activity in normal cell culture conditions.

Limitations of the study include the limited statistical power at the discovery stage, especially for the stroke subtypes, and replication of rare variants with imputed GWAS data. We were able to improve the statistical power on exomes by meta-analyzing WES and WGS, and we performed stroke-subtype specific analyses only for a limited number of suggestive findings to avoid spurious signals due to unstable statistical estimates. To further improve statistical power, we performed statistical aggregate tests on gene exons and on intergenic regions, i.e., enhancers, promoters, and genomic windows. Of note, we studied only transcribed enhancers, and thus, some enhancers could have been missed. We defined promoters with an arbitrarily selected 1,000 bp extension downstream TSS, which may not have always been optimal as the promoter lengths vary. A further limitation is the lack of sequencing-based replication data in individuals with T1D. Instead, we sought for replication by combining available data sources, i.e., FinnGen (Finnish general population GWAS), UK Biobank (general population WES), and FinnDiane (GWAS and genotyping in Finnish individuals with T1D).

The strengths of this study include a well characterized cohort and comprehensively performed single variant and aggregate analyses both for the coding and non-coding regions of the genome.

Stroke is a challenging phenotype to address with ICD codes and many loci associated with rare stroke phenotypes may go unnoticed even with large population-wide genetic studies. We performed analyses for well-defined stroke phenotypes verified by trained neurologists. Furthermore, as we conducted the analyses in specific high-risk individuals from an isolated population, thus with lesser genetic and phenotypic diversity, we had improved statistical opportunities to identify genetic risk loci.

In conclusion, we studied rare and low-frequency stroke-associated variants with WES and WGS in individuals with T1D and report the first genome-wide study on stroke genetics in diabetes. The results highlight 4q33-34.1, *SREBF1*, and *ANK1* for stroke in T1D; and *HAS1*, *UACA*, *LRRN1*, *LINC01500*, and *TRPM2-AS* promoter as stroke risk loci that likely generalize to the non-diabetic population.

## Data availability

Gene aggregate test stroke summary statistics are provided in the Supplementary Data. The sequencing data supporting the current study cannot be deposited in a public repository because of restrictions due to the study consent. The Readers may propose collaboration to research the individual level data with correspondence with the lead investigator.

## Author’s contributions

A.A.A. analyzed the data, wrote the manuscript, and contributed to interpretation of the data, conception and study design, and pre-processing of whole exome sequencing data. J.H. pre-processed the whole-genome sequencing data and contributed to computational analyses and conception and study design. N.S. contributed to acquisition of phenotypic and genotypic data, conception and study design, manuscript writing and interpretation of data. A.Ku. performed lab experiments. A.S, E.K., A.Ky., and A.P. contributed to acquisition and data processing of genetic data. S.H.-H. and A.Y. contributed to acquisition of phenotypic data. J.P., L.M.T., V.H. and P.-H.G. contributed to interpretation of data, acquisition of phenotypic data, and to conception and study design. J.H., A.Ku., A.S., S.H.-H., A.Y., E.K., A.Ky., A.P., J.P., L.M.T, V.H., P.-H.G., and N.S. revised the manuscript critically for important intellectual content. All authors gave final approval of the version to be submitted and any revised version.

## Supporting information

STROBE Guideline

## Acknowledgements

We are indebted to the late Carol Forsblom (1964–2022), the international coordinator of the FinnDiane Study Group, for his considerable contribution. The skilled technical assistance of Heli Krigsman, Hanna Olanne, Maikki Parkkonen, Mira Rahkonen, Anna Sandelin, and Jaana Tuomikangas is gratefully acknowledged. We also want to acknowledge all the physicians and nurses at each FinnDiane center participating in the recruitment and characterization of the individuals with T1D (**Table S24**) and the FinnDiane participants. In addition, we acknowledge the participants and investigators of the FinnGen study. We acknowledge that the ELIXIR Finland node, hosted at the CSC – IT Center for Science for ICT resources, enabling the WES and WGS data processing. Finally, we want to acknowledge Bert Vogelstein and Jukka Kallijärvi for material provided for the *in vitro* promoter experiments: pBV-Luc plasmids (Bert Vogelstein) and renilla control plasmid (Kallijärvi lab, Folkhälsan Research Center).

We utilized data provided by GTEx. The Genotype-Tissue Expression (GTEx) Project was supported by the Common Fund of the Office of the Director of the National Institutes of Health, and by NCI, NHGRI, NHLBI, NIDA, NIMH, and NINDS. The data reported here were obtained from the GTEx Portal on 01/04/2022: https://gtexportal.org/home/.

## References

1. Harjutsalo V, Barlovic DP, Gordin D, Forsblom C, King G, Groop P-H. Presence and Determinants of Cardiovascular Disease and Mortality in Individuals With Type 1 Diabetes of Long Duration: The FinnDiane 50 Years of Diabetes Study. Diabetes Care. 2021;44:1885–1893.

2. Sun H, Saeedi P, Karuranga S, Pinkepank M, Ogurtsova K, Duncan BB, Stein C, Basit A, Chan JCN, Mbanya JC, et al. IDF Diabetes Atlas: Global, regional and country-level diabetes prevalence estimates for 2021 and projections for 2045. Diabetes Res. Clin. Pract. 2022;183.

3. Forlenza GP, Rewers M. The epidemic of type 1 diabetes: what is it telling us? Curr. Opin. Endocrinol. Diabetes Obes. 2011;18:248–251.

4. Ståhl CH, Lind M, Svensson A, Gudbjörnsdottir S, Mårtensson A, Rosengren A. Glycaemic control and excess risk of ischaemic and haemorrhagic stroke in patients with type 1 diabetes: a cohort study of 33 453 patients. J. Intern. Med. 2017;281:261–272.

5. Janghorbani M, Hu FB, Willett WC, Li TY, Manson JE, Logroscino G, Rexrode KM. Prospective study of type 1 and type 2 diabetes and risk of stroke subtypes: the Nurses’ Health Study. Diabetes Care. 2007;30:1730–1735.

6. Thorn LM, Shams S, Gordin D, Liebkind R, Forsblom C, Summanen P, Hägg-Holmberg S, Tatlisumak T, Salonen O, Putaala J, et al. Clinical and MRI Features of Cerebral Small-Vessel Disease in Type 1 Diabetes. Diabetes Care. 2019;42:327–330.

7. Hägg S, Thorn LM, Putaala J, Liebkind R, Harjutsalo V, Forsblom CM, Gordin D, Tatlisumak T, Groop P-H, FinnDiane Study Group. Incidence of stroke according to presence of diabetic nephropathy and severe diabetic retinopathy in patients with type 1 diabetes. Diabetes Care. 2013;36:4140– 4146.

8. Dichgans M, Pulit SL, Rosand J. Stroke genetics: discovery, biology, and clinical applications. Lancet Neurol. 2019;18:587–599.

9. Debette S, Markus HS. Stroke Genetics: Discovery, Insight Into Mechanisms, and Clinical Perspectives. Circ. Res. 2022;130:1095–1111.

10. Mishra A, Malik R, Hachiya T, Jürgenson T, Namba S, Posner DC, Kamanu FK, Koido M, Le Grand Q, Shi M, et al. Stroke genetics informs drug discovery and risk prediction across ancestries. Nature. 2022;611:115–123.

11. Ylinen A, Hägg-Holmberg S, Eriksson MI, Forsblom C, Harjutsalo V, Putaala J, Groop P-H, Thorn LM. The impact of parental risk factors on the risk of stroke in type 1 diabetes. Acta Diabetol. 2021;58:911–917.

12. Syreeni A, Dahlström EH, Hägg-Holmberg S, Forsblom C, Eriksson MI, Harjutsalo V, Putaala J, Groop P-H, Sandholm N, Thorn LM. Haptoglobin Genotype Does Not Confer a Risk of Stroke in Type 1 Diabetes. Diabetes. 2022;71:2728–2738.

13. Dahlström EH, Saksi J, Forsblom C, Uglebjerg N, Mars N, Thorn LM, Harjutsalo V, Rossing P, Ahluwalia TS, Lindsberg PJ, et al. The Low-Expression Variant of FABP4 Is Associated With Cardiovascular Disease in Type 1 Diabetes. Diabetes. 2021;70:2391–2401.

14. Vujkovic M, Keaton JM, Lynch JA, Miller DR, Zhou J, Tcheandjieu C, Huffman JE, Assimes TL, Lorenz K, Zhu X, et al. Discovery of 318 new risk loci for type 2 diabetes and related vascular outcomes among 1.4 million participants in a multi-ancestry meta-analysis. Nat. Genet. 2020;52:680–691.

15. Antikainen AAV, Sandholm N, Trégouët D-A, Charmet R, McKnight AJ, Ahluwalia TS, Syreeni A, Valo E, Forsblom C, Gordin D, et al. Genome-wide association study on coronary artery disease in type 1 diabetes suggests beta-defensin 127 as a risk locus. Cardiovasc. Res. 2021;117:600–612.

16. Qi L, Qi Q, Prudente S, Mendonca C, Andreozzi F, di Pietro N, Sturma M, Novelli V, Mannino GC, Formoso G, et al. Association Between a Genetic Variant Related to Glutamic Acid Metabolism and Coronary Heart Disease in Individuals With Type 2 Diabetes. JAMA. 2013;310:821–828.

17. Fall T, Gustafsson S, Orho-Melander M, Ingelsson E. Genome-wide association study of coronary artery disease among individuals with diabetes: the UK Biobank. Diabetologia. 2018;61:2174– 2179.

18. Grami N, Chong M, Lali R, Mohammadi-Shemirani P, Henshall DE, Rannikmäe K, Paré G. Global assessment of Mendelian stroke genetic prevalence in 101 635 individuals from 7 ethnic groups. Stroke. 2020;51:1290–1293.

19. Si Y, Vanderwerff B, Zöllner S. Why are rare variants hard to impute? Coalescent models reveal theoretical limits in existing algorithms. Genetics. 2021;217:iyab011.

20. Sandholm N, Hotakainen R, Haukka JK, Jansson Sigfrids F, Dahlström EH, Antikainen AA, Valo E, Syreeni A, Kilpeläinen E, Kytölä A, et al. Whole-exome sequencing identifies novel protein-altering variants associated with serum apolipoprotein and lipid concentrations. Genome Med. 2022;14:132.

21. Hu Y, Haessler JW, Manansala R, Wiggins KL, Moscati A, Beiser A, Heard-Costa NL, Sarnowski C, Raffield LM, Chung J, et al. Whole-Genome Sequencing Association Analyses of Stroke and Its Subtypes in Ancestrally Diverse Populations From Trans-Omics for Precision Medicine Project. Stroke. 2022;53:875–885.

22. Jurgens SJ, Choi SH, Morrill VN, Chaffin M, Pirruccello JP, Halford JL, Weng L-C, Nauffal V, Roselli C, Hall AW, et al. Analysis of rare genetic variation underlying cardiometabolic diseases and traits among 200,000 individuals in the UK Biobank. Nat. Genet. 2022;54:240–250.

23. Locke AE, Steinberg KM, Chiang CWK, Service SK, Havulinna AS, Stell L, Pirinen M, Abel HJ, Chiang CC, Fulton RS, et al. Exome sequencing of Finnish isolates enhances rare-variant association power. Nature. 2019;572:323–328.

24. Thorn LM, Forsblom C, Fagerudd J, Thomas MC, Pettersson-Fernholm K, Saraheimo M, Wadén J, Rönnback M, Rosengård-Bärlund M, Björkesten C-G af, et al. Metabolic Syndrome in Type 1 Diabetes: Association with diabetic nephropathy and glycemic control (the FinnDiane study). Diabetes Care. 2005;28:2019–2024.

25. Zhan X, Hu Y, Li B, Abecasis GR, Liu DJ. RVTESTS: an efficient and comprehensive tool for rare variant association analysis using sequence data. Bioinformatics. 2016;32:1423–1426.

26. Willer CJ, Li Y, Abecasis GR. METAL: fast and efficient meta-analysis of genomewide association scans. Bioinformatics. 2010;26:2190–2191.

27. Pei J, Kinch LN, Otwinowski Z, Grishin NV. Mutation severity spectrum of rare alleles in the human genome is predictive of disease type. PLoS Comput. Biol. 2020;16:e1007775.

28. Chang CC, Chow CC, Tellier LC, Vattikuti S, Purcell SM, Lee JJ. Second-generation PLINK: rising to the challenge of larger and richer datasets. Gigascience. 2015;4:s13742–015.

29. Lee S, Teslovich TM, Boehnke M, Lin X. General framework for meta-analysis of rare variants in sequencing association studies. Am. J. Hum. Genet. 2013;93:42–53.

30. Rivas MA, Pirinen M, Conrad DF, Lek M, Tsang EK, Karczewski KJ, Maller JB, Kukurba KR, DeLuca DS, Fromer M, et al. Effect of predicted protein-truncating genetic variants on the human transcriptome. Science. 2015;348:666–669.

31. Li X, Li Z, Zhou H, Gaynor SM, Liu Y, Chen H, Sun R, Dey R, Arnett DK, Aslibekyan S, et al. Dynamic incorporation of multiple in silico functional annotations empowers rare variant association analysis of large whole-genome sequencing studies at scale. Nat. Genet. 2020;52:969–983.

32. Rentzsch P, Witten D, Cooper GM, Shendure J, Kircher M. CADD: predicting the deleteriousness of variants throughout the human genome. Nucleic Acids Res. 2019;47:D886–D894.

33. Abugessaisa I, Noguchi S, Hasegawa A, Harshbarger J, Kondo A, Lizio M, Severin J, Carninci P, Kawaji H, Kasukawa T. FANTOM5 CAGE profiles of human and mouse reprocessed for GRCh38 and GRCm38 genome assemblies. Sci. Data. 2017;4:1–10.

34. Moore CM, Jacobson SA, Fingerlin TE. Power and Sample Size Calculations for Genetic Association Studies in the Presence of Genetic Model Misspecification. Hum. Hered. 2019;84:256–271.

35. Chen H, Huffman JE, Brody JA, Wang C, Lee S, Li Z, Gogarten SM, Sofer T, Bielak LF, Bis JC, et al. Efficient Variant Set Mixed Model Association Tests for Continuous and Binary Traits in Large-Scale Whole-Genome Sequencing Studies. Am. J. Hum. Genet. 2019;104:260–274.

36. Zhou X, Stephens M. Genome-wide efficient mixed-model analysis for association studies. Nat. Genet. 2012;44:821–824.

37. Backman JD, Li AH, Marcketta A, Sun D, Mbatchou J, Kessler MD, Benner C, Liu D, Locke AE, Balasubramanian S, et al. Exome sequencing and analysis of 454,787 UK Biobank participants. Nature. 2021;599:628–634.

38. Võsa U, Claringbould A, Westra H-J, Bonder MJ, Deelen P, Zeng B, Kirsten H, Saha A, Kreuzhuber R, Yazar S, et al. Large-scale cis- and trans-eQTL analyses identify thousands of genetic loci and polygenic scores that regulate blood gene expression. Nat. Genet. 2021;53:1300–1310.

39. Wang Y, Song F, Zhang B, Zhang L, Xu J, Kuang D, Li D, Choudhary MNK, Li Y, Hu M, et al. The 3D Genome Browser: a web-based browser for visualizing 3D genome organization and long-range chromatin interactions. Genome Biol. 2018;19:151.

40. Zong P, Lin Q, Feng J, Yue L. A Systemic Review of the Integral Role of TRPM2 in Ischemic Stroke: From Upstream Risk Factors to Ultimate Neuronal Death. Cells. 2022;11:491.

41. Belrose JC, Jackson MF. TRPM2: a candidate therapeutic target for treating neurological diseases. Acta Pharmacol. Sin. 2018;39:722–732.

42. DeBose-Boyd RA, Ye J. SREBPs in Lipid Metabolism, Insulin Signaling, and Beyond. Trends Biochem. Sci. 2018;43:358–368.

43. Adzhubei IA, Schmidt S, Peshkin L, Ramensky VE, Gerasimova A, Bork P, Kondrashov AS, Sunyaev SR. A method and server for predicting damaging missense mutations. Nat. Methods. 2010;7:248– 249.

44. Kumar P, Henikoff S, Ng PC. Predicting the effects of coding non-synonymous variants on protein function using the SIFT algorithm. Nat. Protoc. 2009;4:1073–1081.

45. Park J, Jeong D-C, Yoo J, Jang W, Chae H, Kim J, Kwon A, Choi H, Lee JW, Chung N-G, et al. Mutational characteristics of ANK1 and SPTB genes in hereditary spherocytosis. Clin. Genet. 2016;90:69–78.

46. Yan R, Lai S, Yang Y, Shi H, Cai Z, Sorrentino V, Du H, Chen H. A novel type 2 diabetes risk allele increases the promoter activity of the muscle-specific small ankyrin 1 gene. Sci. Rep. 2016;6:25105.

47. Malik R, Chauhan G, Traylor M, Sargurupremraj M, Okada Y, Mishra A, Rutten-Jacobs L, Giese A-K, van der Laan SW, Gretarsdottir S, et al. Multiancestry genome-wide association study of 520,000 subjects identifies 32 loci associated with stroke and stroke subtypes. Nat. Genet. 2018;50:524– 537.

48. Siiskonen H, Oikari S, Pasonen-Seppänen S, Rilla K. Hyaluronan Synthase 1: A Mysterious Enzyme with Unexpected Functions. Front. Immunol. 2015;6:1–11.

49. Katarzyna Greda A, Nowicka D. Hyaluronidase inhibition accelerates functional recovery from stroke in the mouse brain. J. Neurochem. 2021;157:781–801.

50. Wang G, Tiemeier GL, van den Berg BM, Rabelink TJ. Endothelial Glycocalyx Hyaluronan: Regulation and Role in Prevention of Diabetic Complications. Am. J. Pathol. 2020;190:781–790.

