## Supplementary material for "Whole-genome sequencing identifies variants in *ANK1*, *LRRN1*, *HAS1,* and other genes and regulatory regions for stroke in type 1 diabetes": STROBE Guideline

### STROBE checklist

| Item | STROBE guideline | Extension for Genetic Association Studies | Page No |
|---|---|---|---|
| **Title and Abstract** | (a) Indicate the study's design with a commonly used term in the title or the abstract | | Title page, 1 |
| | (b) Provide in the abstract an informative and balanced summary of what was done and what was found | | 1 |
| **Introduction** | | | |
| **Background rationale** | Explain the scientific background and rationale for the investigation being reported | | 3-5 |
| **Objectives** | State specific objectives, including any pre-specified hypotheses | *State if the study is the first report of a genetic association, a replication effort, or both* | 4-5 |
| **Methods** | | | |
| **Study design** | Present key elements of study design early in the paper | | 6-7, Fig. 1 |
| **Setting** | Describe the setting, locations and relevant dates, including periods of recruitment, exposure, follow-up, and data collection | | 6, Sup. Material pages 1-2 |
| **Participants** | (a) *Cohort study:* give the eligibility criteria, and the sources and methods of selection of participants. Describe methods of follow-up | *Give information on the criteria and methods for selection of subsets of participants from a larger study, when relevant* | 6, Sup. Material pages 1-2, Table S2, Table S8 |
| | *Case–control study*: give the eligibility criteria, and the sources and methods of case ascertainment and control selection. Give the rationale for the choice of cases and controls | | |
| | *Cross-sectional study:* give the eligibility criteria, and the sources and methods of selection of participants | | |
| | (b) *Cohort study:* for matched studies, give matching criteria and number of exposed and unexposed | | |
| | *Case–control study*: for matched studies, give matching criteria and the number of controls per case | | |
| **Variables** | (a) Clearly define all outcomes, exposures, predictors, potential confounders, and effect modifiers. Give diagnostic criteria, if applicable | *(b) Clearly define genetic exposures (genetic variants) using a widely-used nomenclature system. Identify variables likely to be associated with population stratification (confounding by ethnic origin)* | 6, Sup. Material pages 1-2, Table 1, Figures S1-S4, Tables S1, S3-S5 |
| **Data sources/ measurement** | (a) For each variable of interest, give sources of data and details of methods of assessment (measurement). Describe comparability of assessment methods if there is more than one group | *(b) Describe laboratory methods, including source and storage of DNA, genotyping methods and platforms (including the allele calling algorithm used, and its version), error rates and call rates. State the laboratory/center where genotyping was done. Describe comparability of laboratory methods if there is more than one group. Specify whether genotypes were assigned using all of the data from the study simultaneously or in smaller batches* | Sup. Material pages 1-4, Figures S18-S20 |
| **Bias** | (a) Describe any efforts to address potential sources of bias | *(b) For quantitative outcome variables, specify if any investigation of potential bias resulting from pharmacotherapy was undertaken. If relevant, describe the nature and magnitude of the potential bias, and explain what approach was used to deal with this* | 7, Sup. Material pages 2-4 |
| **Study size** | Explain how the study size was arrived at | | 6, Sup. Material pages 1-2, Figures S19-S20, Table S8 |
| **Quantitative variables** | Explain how quantitative variables were handled in the analyses. If applicable, describe which groupings were chosen, and why | *If applicable, describe how effects of treatment were dealt with* | 6-7, Sup. Material pages 1-2, Table S2 |
| **Statistical methods** | (a) Describe all statistical methods, including those used to control for confounding | *State software version used and options (or settings) chosen* | 7-9, Sup. Material pages 2-4 |
| | (b) Describe any methods used to examine subgroups and interactions | | |
| | (c) Explain how missing data were addressed | | |
| | *Cohort study*: if applicable, explain how loss to follow-up was addressed | | |
| | *Case–control study*: if applicable, explain how matching of cases and controls was addressed | | |
| | *Cross-sectional study*: if applicable, describe analytical methods taking account of sampling strategy | | |
| | (e) Describe any sensitivity analyses | | |
| | | *(f) State whether Hardy–Weinberg equilibrium was considered and, if so, how* | Sup. Material page 1, Table 2, Figures S19-S20 |
| | | *(g) Describe any methods used for inferring genotypes or haplotypes* | Sup. Material pages 1-2, Figures S19-S20 |
| | | *(h) Describe any methods used to assess or address population stratification* | 7, 9, Sup. Material pages 2-4 |

| | | | |
|---|---|---|---|
| | | *(i) Describe any methods used to address multiple comparisons or to control risk of false-positive findings* | Sup. Material pages 2-4, Figure 2 |
| | | *(j) Describe any methods used to address and correct for relatedness among subjects* | 8-9, Sup. Material pages 2-3 |
| **Results** | | | |
| **Participants** | (a) Report the numbers of individuals at each stage of the study—e.g., numbers potentially eligible, examined for eligibility, confirmed eligible, included in the study, completing follow-up, and analyzed | *Report numbers of individuals in whom genotyping was attempted and numbers of individuals in whom genotyping was successful* | 6, Sup. Material pages 1-2, Table S2, Table S8, Figures S19-S20 |
| | (b) Give reasons for non-participation at each stage | | |
| | (c) Consider use of a flow diagram | | |
| **Descriptive data** | (a) Give characteristics of study participants (e.g., demographic, clinical, social) and information on exposures and potential confounders | *Consider giving information by genotype* | Table 1, Table S1, Tables S3-S5, Figures S1-S4 |
| | (b) Indicate the number of participants with missing data for each variable of interest | | |
| | (c) *Cohort study*: summarize follow-up time, e.g., average and total amount | | |
| **Outcome data** | *Cohort study*: report numbers of outcome events or summary measures over time | *Report outcomes (phenotypes) for each genotype category over time* | Table 1, Table S1, Tables S3-S5, Figures S1-S4 |
| | *Case–control study*: report numbers in each exposure category, or summary measures of exposure | *Report numbers in each genotype category* | |
| | *Cross-sectional study*: report numbers of outcome events or summary measures | *Report outcomes (phenotypes) for each genotype category* | |
| **Main results** | (a) Give unadjusted estimates and, if applicable, confounder-adjusted estimates and their precision (e.g., 95% confidence intervals). Make clear which confounders were adjusted for and why they were included | | 10-16, Tables 2-3, Figure 3, Figure S18, Tables S10-S23 |
| | (b) Report category boundaries when continuous variables were categorized | | |
| | (c) If relevant, consider translating estimates of relative risk into absolute risk for a meaningful time period | | |
| | | *(d) Report results of any adjustments for multiple comparisons* | |
| **Other analyses** | (a) Report other analyses done—e.g., analyses of subgroups and interactions, and sensitivity analyses | | 10-16 Figures 2 and 4 |
| | | *(b) If numerous genetic exposures (genetic variants) were examined, summarize results from all analyses undertaken* | |
| | | *(c) If detailed results are available elsewhere, state how they can be accessed* | 20, Sup. Material page 4 |
| **Discussion** | | | |
| **Key results** | Summarize key results with reference to study objectives | | 17-19 |
| **Limitations** | Discuss limitations of the study, taking into account sources of potential bias or imprecision. Discuss both direction and magnitude of any potential bias | | 19 |
| **Interpretation** | Give a cautious overall interpretation of results considering objectives, limitations, multiplicity of analyses, results from similar studies, and other relevant evidence | | 17-20 |
| **Generalizability** | Discuss the generalizability (external validity) of the study results | | 20 |
| **Other information** | | | |
| **Funding** | Give the source of funding and the role of the funders for the present study and, if applicable, for the original study on which the present article is based | | 20-23 |
