## Supplementary Information for "Whole-genome sequencing identifies variants in *ANK1*, *LRRN1*, *HAS1,* and other genes and regulatory regions for stroke in type 1 diabetes"

#### Supplementary Material

Detailed Methods ... 1
- Participants and stroke definitions ... 1
- Sequencing material ... 1
- Replication material ... 2
- Statistical methods ... 2
  - Single variant analyses ... 2
  - Gene aggregate analyses ... 2
  - Sliding-window analyses and regulatory regions ... 3
- Regional plots and functional characterization ... 3
- Functional research on *TRPM2-AS* promoter ... 3
- Single variant replication ... 4
- Gene aggregate replication ... 4
- Sliding-window and regulatory region replication ... 4

Supplemental Figures ... 5
- Figure S1: Clinical characteristics of individuals in WGS ... 5
- Figure S2: Clinical characteristics of individuals in WES ... 6
- Figure S3: Clinical characteristics of individuals in FinnDiane GWAS ... 7
- Figure S4: Clinical characteristics of individuals in genotyping ... 8
- Figure S5: Annotation PCA within functional classes (CADD) ... 9
- Figure S6: Statistical power in replication ... 10
- Figure S7: Additive single variant analysis with minimal adjustment. ... 11
- Figure S8: Single variant analysis with DKD adjustment ... 12
- Figure S9: Recessive single variant analyses ... 13
- Figure S10: SKAT-O minimal model Manhattan plot ... 14
- Figure S11: SKAT-O with additional DKD adjustment Manhattan plot ... 15
- Figure S12: *MAP3K12* and *TARBP2* regional plot ... 16
- Figure S13: Known Mendelian stroke-risk genes in T1D ... 17
- Figure S14: Topologically associating domain (TAD) on 4q33-34.1 ... 18
- Figure S15: Enhancer variant PCHi-C links ... 19

### Detailed Methods

#### Participants and stroke definitions

The study is part of the FinnDiane Study established to investigate complications of T1D. We have access to 490 and 583 non-related individuals with T1D with WES and WGS, respectively. Individuals in the present study were diagnosed with T1D by their attending physician and had diabetes onset age <40 and insulin initiated within one calendar year from the diabetes diagnosis. Stroke phenotypes were identified from the Finnish Death Registry, Statistics Finland, and the Care Register for Health Care, Finnish Institute for Health and Welfare until the end of 2017 and were verified by trained neurologists from medical files and brain imaging data. We required acute stroke events to have occurred after the T1D diagnosis. For individuals without data verified by neurologists available ($N_{WGS}$=27, $N_{WES}$=2), we considered only the registry data, and excluded five controls with t*ransient ischemic attacks or other intermediate stroke phenotypes* due to potential for misclassification (**Table S2**). Furthermore, we classified stroke events into ischemic- and hemorrhagic strokes whenever stroke subtypes verified by neurologists were available. Ischemic strokes entailed lacunar-, non-lacunar-, and unclear infarctions, while hemorrhagic strokes comprised intracerebral and subarachnoid hemorrhages. We limited controls to individuals with >35 years of age and >20 years of diabetes duration, because only ~5% of cases had experienced their first stroke event before these limits in sequencing data (**Table 1, Table S1, Figure S1** and **S2**).

#### Sequencing material

In total, 599 individuals were whole genome sequenced at Macrogen Inc. using the Illumina HiSeq X platform (Macrogen Inc., Rockville, MD, USA) with at least 30× average coverage (1-8 lanes). Furthermore, 502 participants were whole-exome sequenced at the University of Oxford, UK, as described earlier[1]. In short, libraries were multiplexed and captured with Illumina TruSeq™ Exome Enrichment Kit and sequenced on an Illumina HiSeq2000 with 100 bp paired end reads (1-2 lanes). Seven samples did not pass the initial quality control (QC) or showed discrepancies with other genetic data. For this work, the pre-processed aligned reads of 495 individuals were converted back to unaligned FASTQ-reads and re-processed with comparable pipeline to WGS.

First, we trimmed the WES and WGS sequencing reads[2]. We processed 495 WES and 599 WGS samples further according to Broad Institute's best practices guidelines with Genome Analysis Toolkit 4 (GATK4)[3] (**Figure S19**): We aligned reads by lane to GRCh38 reference genome with Burrows-Wheeler Aligner, sorted and marked duplicate reads, recalibrated bases by chromosome, and called variants by sample. WES and WGS were joint called separately. We filtered variants according to excess heterozygosity threshold of 54.69, truth sensitivity level 99.7%, and GATK's recommended tranche thresholds for SNPs and indels. We assessed sample concordance with FinnDiane GWAS data whenever possible. Finally, 490 and 583 individuals passed QC within WES and WGS, respectively. In variant QC, for autosomal variants, we required Hardy-Weinberg equilibrium (HWE) *p*-value $>10^{-10}$ and variant call rate >98%; and for X chromosome variants, only variant call rate >98%. We annotated variants with SNPEff v.5 software[4]. WES and WGS comprised 324,817 and 21.92 million variants, respectively.

#### Replication material

In FinnDiane, we have GWAS data for 6,458 Finnish individuals with T1D and their relatives. Genomes were genotyped at the University of Virginia, and previously processed to GRCh37 reference genome[5]. We shifted the genotyping positions to GRCh38, re-imputed the data to SISu v3 reference panel, and annotated with SNPEff v.5 software[4] (**Figure S20**). We restricted the data to 15,026,805 high imputation quality variants ($r^2$>0.80), and individuals to those with T1D; age at onset <40 years and insulin treatment initiated within two years from diagnosis, if insulin initiation year known, and to those not in the sequencing data. Controls were required to have diabetes duration >20 years and age >35 years. GWAS replication entailed 367 cases and 3,578 controls (**Figure S3, Tables S3** and **S4**). Instead of principal component adjustment, we utilized rvtests kinship matrix with Balding-Nichol's approximation in single variant analysis and GEMMA relatedness matrix in aggregate analyses[6,7].

Twelve lead variants were selected for replication by variant genotyping in 3,600 FinnDiane participants with T1D on one Agena iPlex multiplexing assay at the Institute for Molecular Medicine Finland, Helsinki, Finland (**Table S8**). Variants were prioritized onto to the same multiplexing design based on statistical significance, and one heterozygous variant carrier within sequencing data included as the positive control. Replication was limited to individuals within GWAS data to perform relatedness adjustment with kinship matrix (N=3,263, **Table S5, Figure S4**).

#### Statistical methods

##### Single variant analyses

We analyzed variants available in WES and WGS data with score test fixed-effect inverse variance based meta-analysis (CMAC≥5, WES and WGS: MAC≥2), and variants only available in one of the data sets with Firth regression (MAC≥5). Furthermore, autosomal variants were analyzed recessively according to similar scheme (cumulative homozygote minor allele carriers ≥5; WES and WGS homozygote carriers ≥2). Analyses were carried out with rvtests (version 20190205)[6] and metal (version 20110325)[8], except autosomal recessive Firth regression with plink2 (version 20210420)[9,10].

##### Gene aggregate analyses

We performed autosomal gene aggregate tests with SKAT-O, combining burden and variance-component tests, with the aim to increase statistical power and stability[11]. We performed SKAT-O meta-analysis, separately with PAVs and PTVs, between WES and WGS using MetaSKAT (version 0.81)[12], which exploits single variant score statistics. PTVs were here defined as predicted loss-of-function mutations (**Table S6**). PAV analyses entail, in addition to the PTVs, mutations that are predicted to alter amino acid sequence. Only variable sites (MAC≥1) were accepted into gene aggregate ($N_{variant}$≥2, CMAC≥5). We did not report genes with all variants in perfect LD, and inspected individual variant stroke-associations within the genes using score test fixed-effects meta-analysis[6,8]. Multiple testing correction based on number of included genes resulted in significance thresholds of $p$-value<$4\times10^{-6}$ for PAVs (MAF≤1% and MAF≤5%), $p$-value<$7\times10^{-5}$ for PTVs with MAF≤1%, and $p$-value<$5\times10^{-5}$ for PTVs with MAF≤5%. For the known Mendelian stroke risk genes[13], we reported results regardless of variant number or CMAC.

#### Sliding-window analyses and regulatory regions

We performed functional annotation weighted sliding window analyses on the WGS data with the STAAR R package 0.9.6[14]. For variant annotations we utilized CADD v1.6 GRCh38 data[15,16], more specifically; variant MAF (to up-weight rarer variants), pre-computed CADD score, and the first annotation principal components from seven annotation classes (**Figure S5, Table S7**). Following guidelines[14], missing annotations were imputed to default and distributions were standardized before principal component analysis. Annotations were transformed to PHRED scale with appropriate direction. We utilized 4,000 bp windows ($N_{variant}\geq2$, CMAC≥5) separated by 2,000 bp skips. We studied 20.7 million autosomal variants with available functional annotations.

As enhancers and promoters, we considered FANTOM5 CAGE profiles reprocessed to the GRCh38 reference genome (https://fantom.gsc.riken.jp/5/)[17–19]; however, we extended CAGE TSSs to form full-length promoters (1,000 bp). We analyzed regulatory regions ($N_{variant}\geq2$, CMAC≥5) with the STAAR R package[14], using only allele frequencies as variant annotations, which allowed us to include more autosomal variants. With low-frequency variants, 184,192 promoters and 24,045 enhancers were analyzed, resulting in multiple testing corrected significance thresholds $p$-value<$2.9\times10^{-7}$ and $p$-value<$2.6\times10^{-6}$, respectively. For rare variants, the thresholds were $p$-value<$3.5\times10^{-7}$ and $p$-value<$4.3\times10^{-6}$, respectively. We did not report regions with all variants in perfect LD.

#### Regional plots and functional characterization

We produced regional association plots with LocusZoom[20] and Gviz R package 1.38.3[21]; and inspected variant characteristics from GTEx Portal, eQTLGen Consortium ($p$-value<0.05)[22], RegulomeDB[23], YUE Lab (http://3dgenome.fsm.northwestern.edu/)[24], and the Ensemble Variant Effect Predictor[25–27].

#### Functional research on *TRPM2-AS* promoter

HELA, HEK-293 and HUVEC cells were cultured as per standard protocol. Total RNA was extracted using TRIzol™ Reagent (Cat. No. 15596026, Thermo scientific.), cDNA was synthesized using SuperScript™ III Reverse Transcriptase (Cat. No. 18080044, Invitrogen), and qPCR was done using iTaq™ Universal SYBR® Green Supermix (Cat. No. 1725121, Bio-Rad) as per manufacturer's protocol. Promoter sequence was PCR amplified using Phusion™ High-Fidelity DNA Polymerase (F530L, Thermo scientific.) from genomic DNA of one FinnDiane participant carrying rs753589764 minor allele, and one participant carrying none of the identified *TRPM2-AS* rare minor alleles; and cloned on pBV-Luc plasmid (Addgene plasmid #16539, gift from Bert Vogelstein). Transfection of promoter sequence carrying pBV-Luc reporter and renilla luciferase control plasmid was done using FuGENE® HD Transfection Reagent (Cat. No. E2311, Promega): Four technical repeats each transfection. Dual-Luciferase Reporter Assay (Cat. No. E1910, promega) was performed as per manufacturer's recommendations.

#### Single variant replication

We performed GWAS in individuals with T1D using score test (rvtests version 20190205)[6], and calculated statistical power with the genpwr R package 1.0.4[28]. Genotyping was successful for a varying number of individuals depending on the variant. We observed mutation carriers for six variants (**Table S8**) and performed single variant analyses similarly with score test. We analyzed one *LRRN1* variant with linear regression (stats R package 4.2.1) and without relatedness adjustment, because no mutation carriers were observed among individuals in the GWAS data i.e., kinship matrix.

We attempted general population replication from the FinnGen project release 6 (https://www.finngen.fi/en), and selected stroke phenotypes that best matched our definitions (**Table S9**).

#### Gene aggregate replication

We performed SKAT-O analyses in FinnDiane GWAS ($r^2$>0.80) for T1D specific replication with the GMMAT R package 1.3.2[29], by including also genotyped variants, and imputed missing data to mean (i.e., individuals with missing genotype at a variant). We attempted general population replication from UK Biobank WES studies[30,31].

#### Sliding-window and regulatory region replication

We attempted T1D specific replication with GWAS data ($r^2$>0.80) using STAAR R package[14].

### Supplemental Figures

#### Figure S1: Clinical characteristics of individuals in WGS

**A.** Age, **B.** Diabetes duration, **C.** Age at diabetes onset, **D.** Calendar year of diabetes onset, **E.** Weighted mean HbA1c, **F.** DKD status.

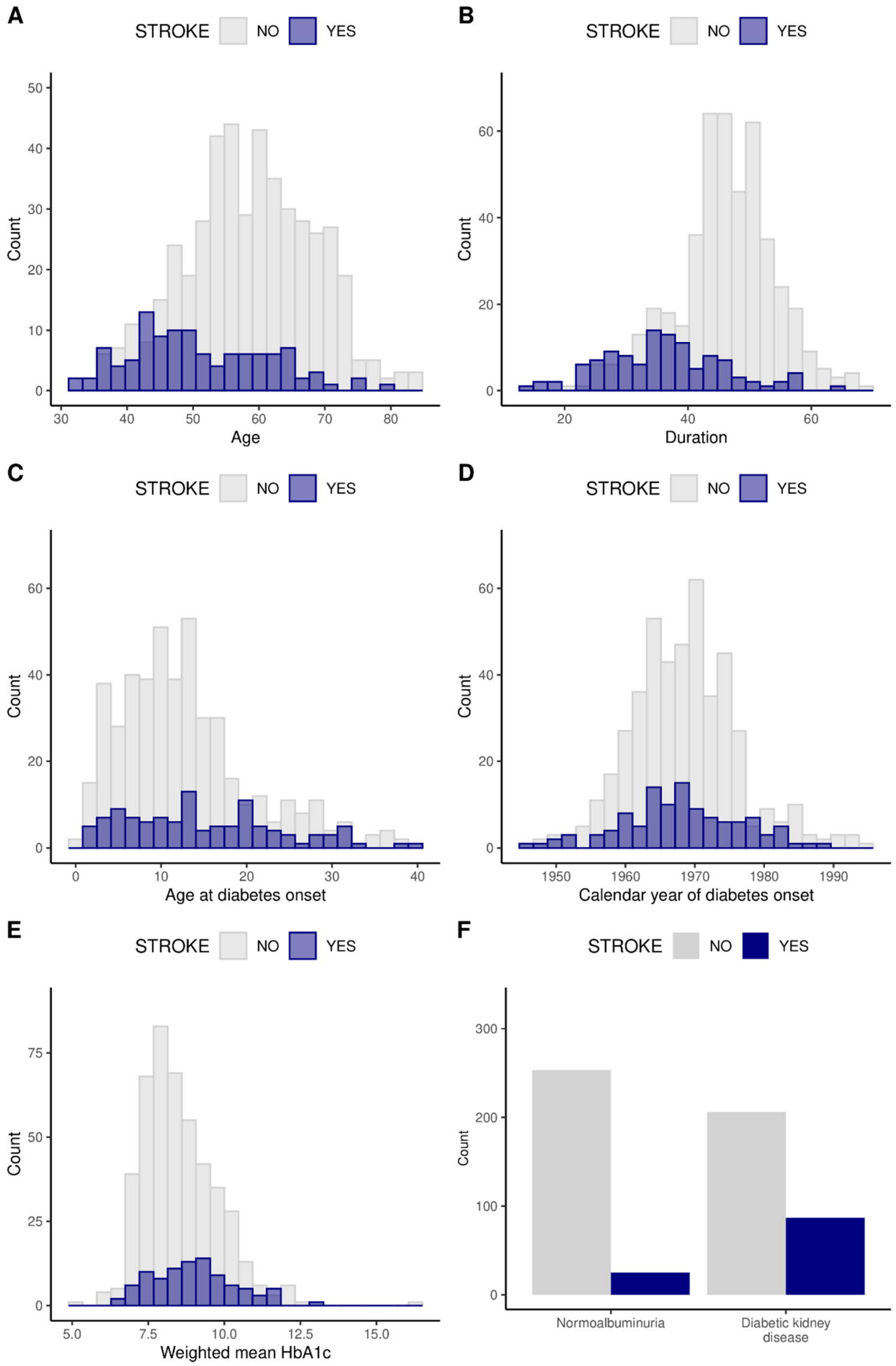

Figure S2: Clinical characteristics of individuals in WES

**A.** Age, **B.** Diabetes duration, **C.** Age at diabetes onset, **D.** Calendar year of diabetes onset, **E.** Weighted mean HbA1c, **F.** DKD status.

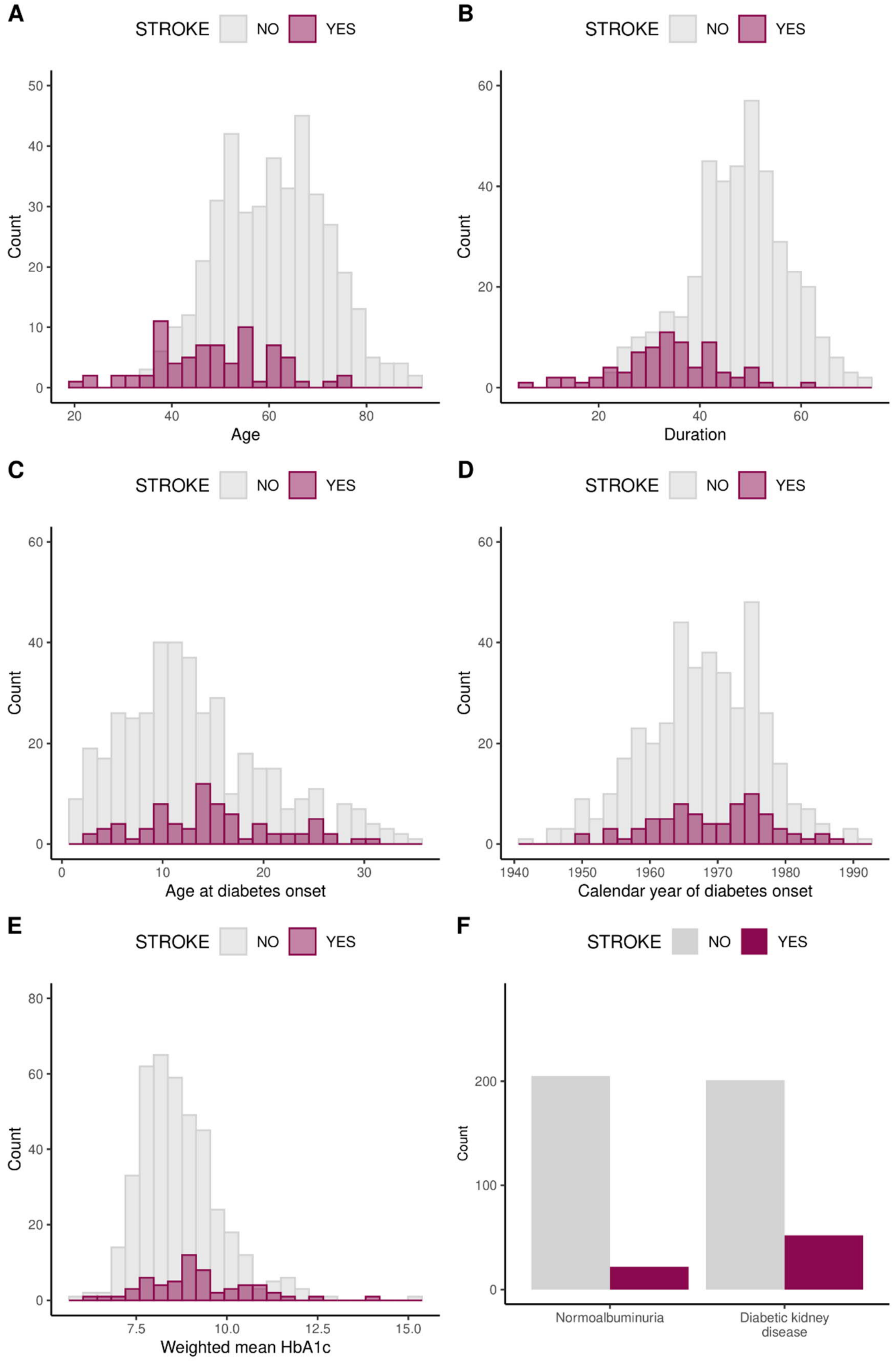

Figure S3: Clinical characteristics of individuals in FinnDiane GWAS

GWAS replication within FinnDiane. **A.** Age, **B.** Diabetes duration, **C.** Age at diabetes onset, **D.** Calendar year of diabetes onset, **E.** Weighted mean HbA1c, **F.** DKD status.

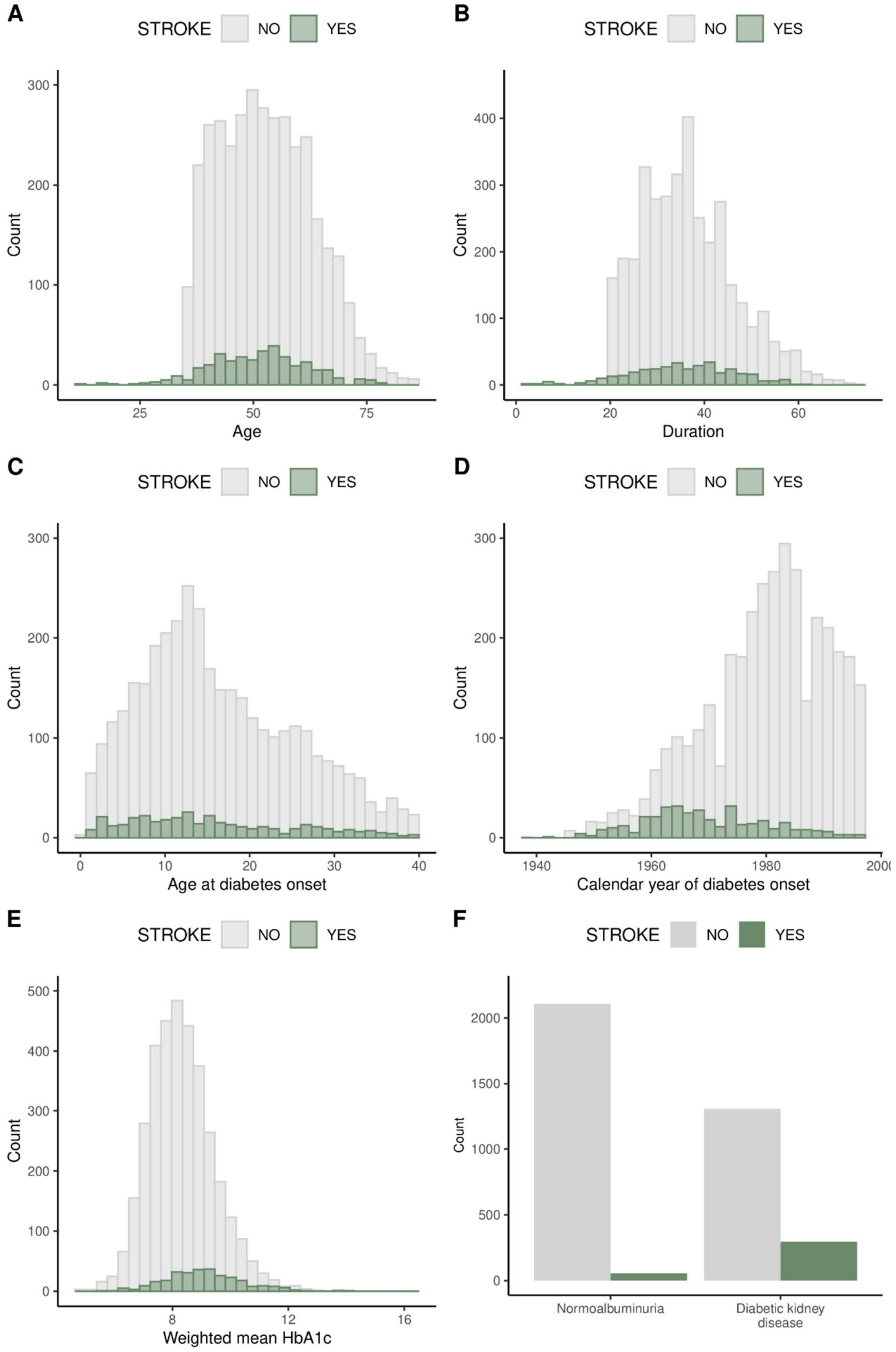

Figure S4: Clinical characteristics of individuals in genotyping

Genotyping replication within FinnDiane. **A.** Age, **B.** Diabetes duration, **C.** Age at diabetes onset, **D.** Calendar year of diabetes onset, **E.** Weighted mean HbA1c, **F.** DKD status.

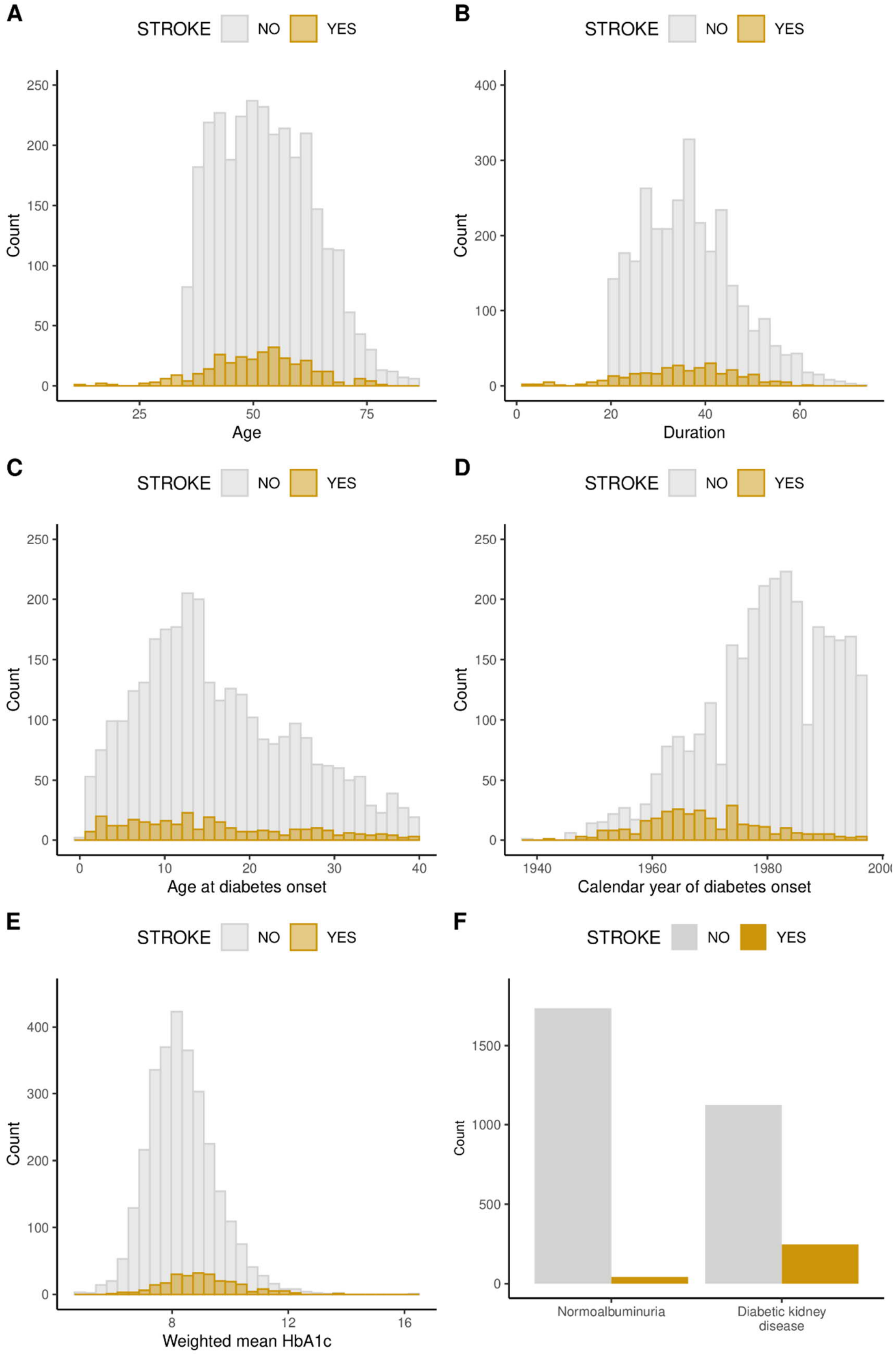

#### Figure S5: Annotation PCA within functional classes (CADD)

**A.** Conservation, **B.** Epigenetics, **C.** microRNA, **D.** Mutation density, **E.** Protein function, **F.** Transcription factor, **G.** Proximity to transcription start- and end sites. Variance explained by the corresponding annotation principal component (aPC) is presented at Dimension 1.

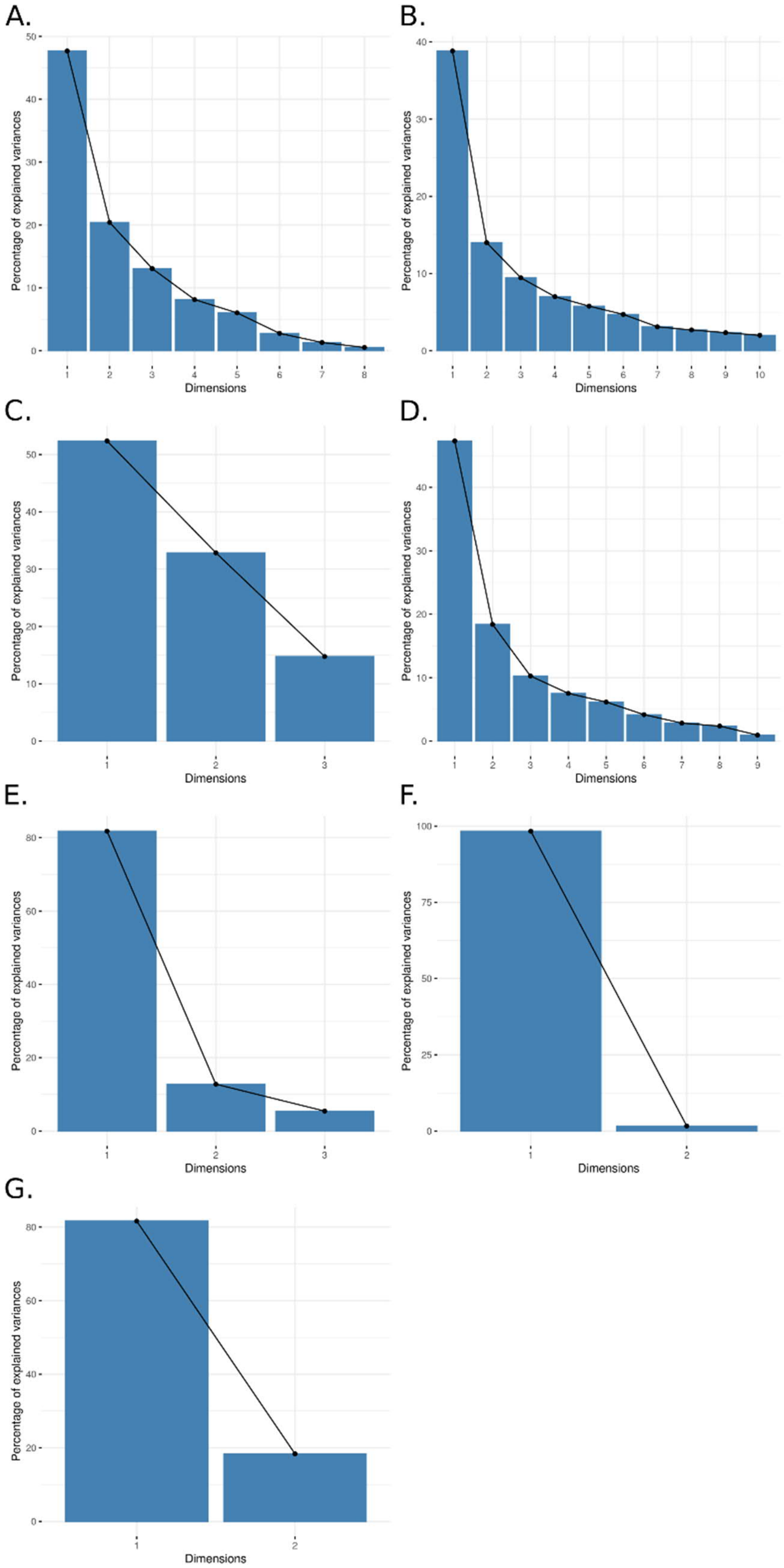

#### Figure S6: Statistical power in replication

Statistical power to replicate low-frequency (MAF=5%, MAF=1%) and rare (MAF=0.5%, MAF=0.1%) variants with the number of patients in FinnDiane GWAS replication (N=3,945, $N_{cases}$=367) using different alpha levels: **A.** Nominal significance (0.05), **B.** Multiple testing corrected threshold (0.01), **C.** Multiple testing corrected threshold (0.005), **D.** Multiple testing corrected threshold (0.0005).

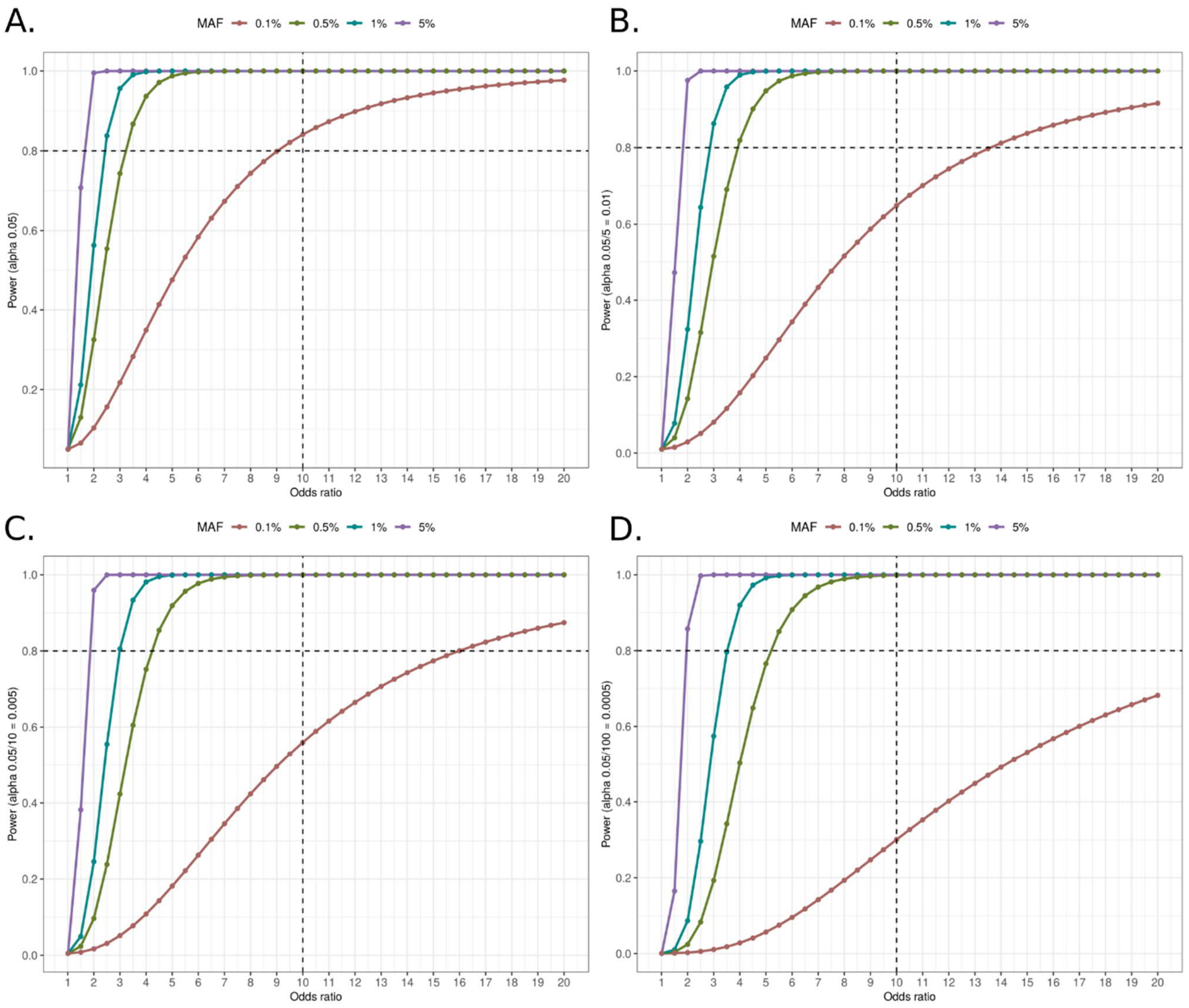

Figure S7: Additive single variant analysis with minimal adjustment.

**A.** Manhattan plot, **B.** LocusZoom plot of 4q33.1-34 (LD structure according to a nearby variant, rs4386563).

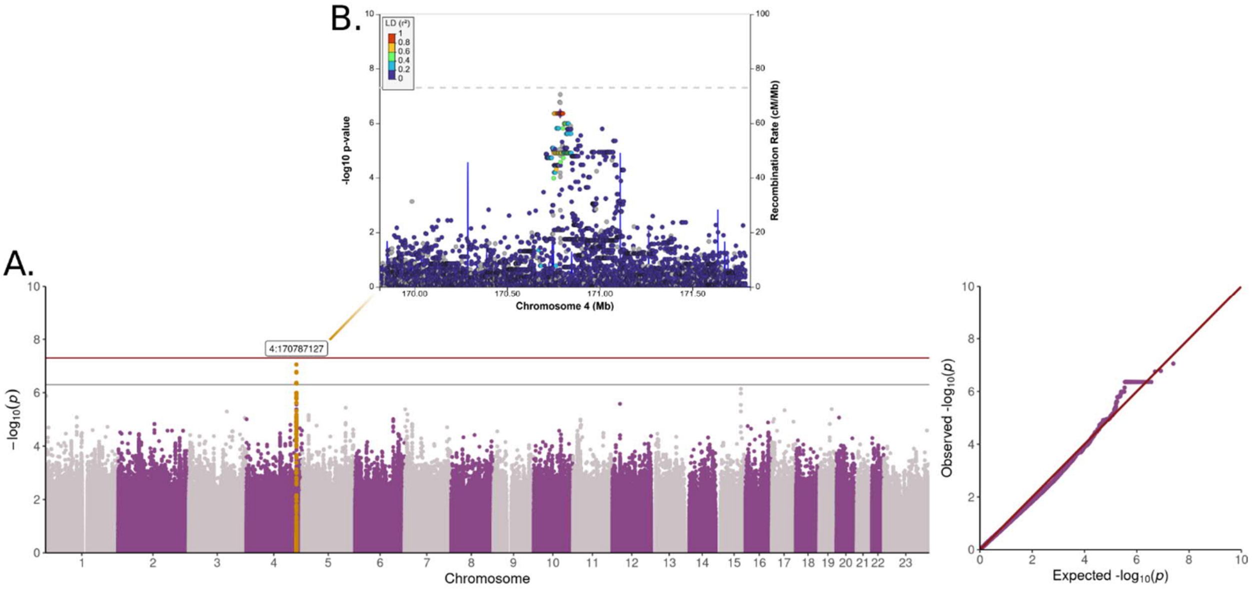

#### Figure S8: Single variant analysis with DKD adjustment

Stroke single variant analysis Manhattan plot (Firth regression or score test fixe-effects meta-analysis).

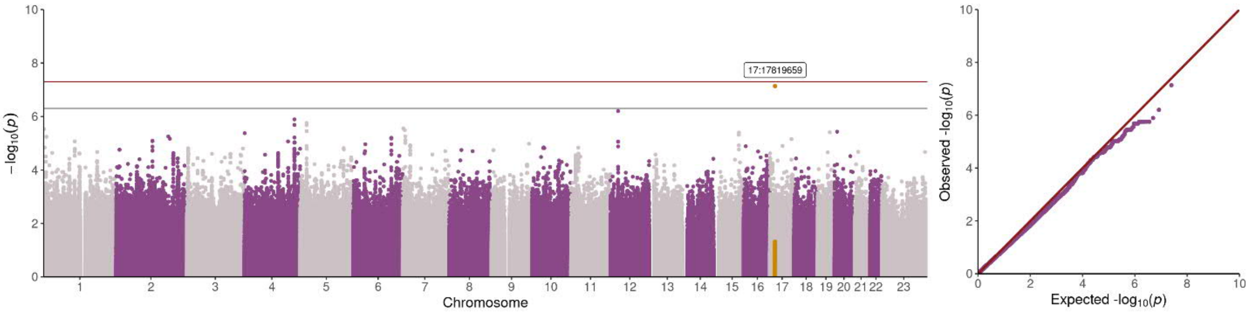

### Figure S9: Recessive single variant analyses

Stroke recessive autosomal single variant analysis Manhattan plots (Firth regression or score test fixed-effects meta-analysis): **A.** Minimal adjustment, and **B.** An additional DKD adjustment.

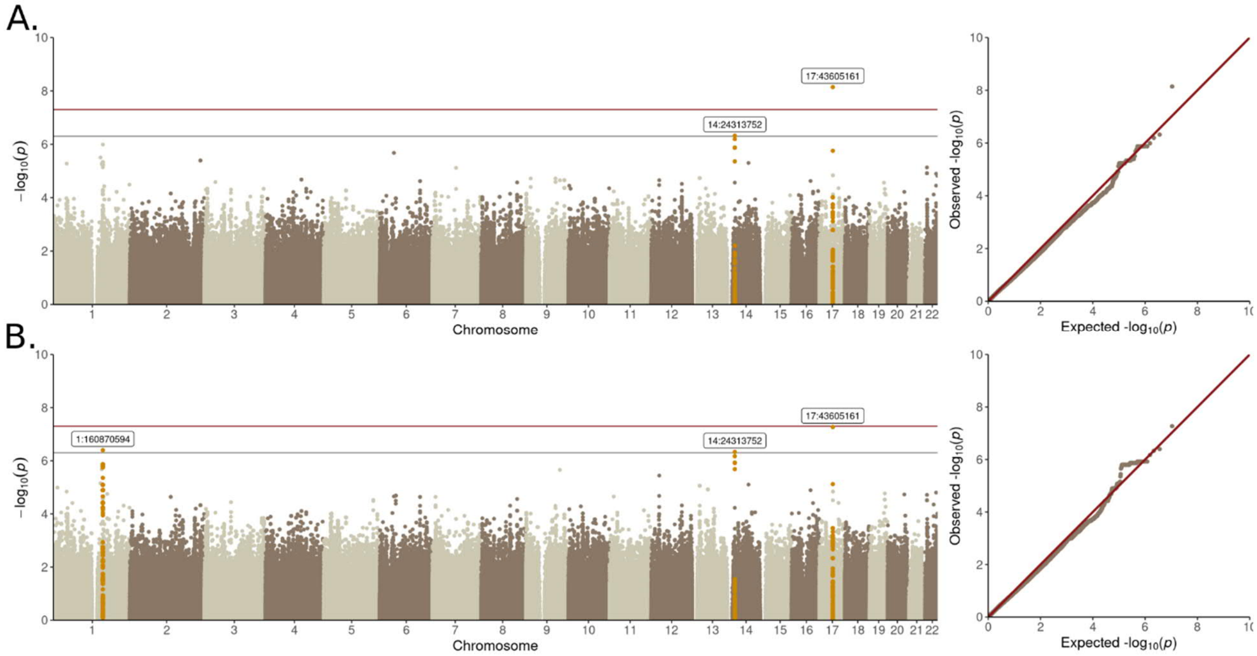

Figure S10: SKAT-O minimal model Manhattan plot

**A.** Protein altering variant (PAV) ≤1%, **B.** PAV≤5%, **C.** Protein truncating variant (PTV) ≤1%, **D.** PTV≤5%.

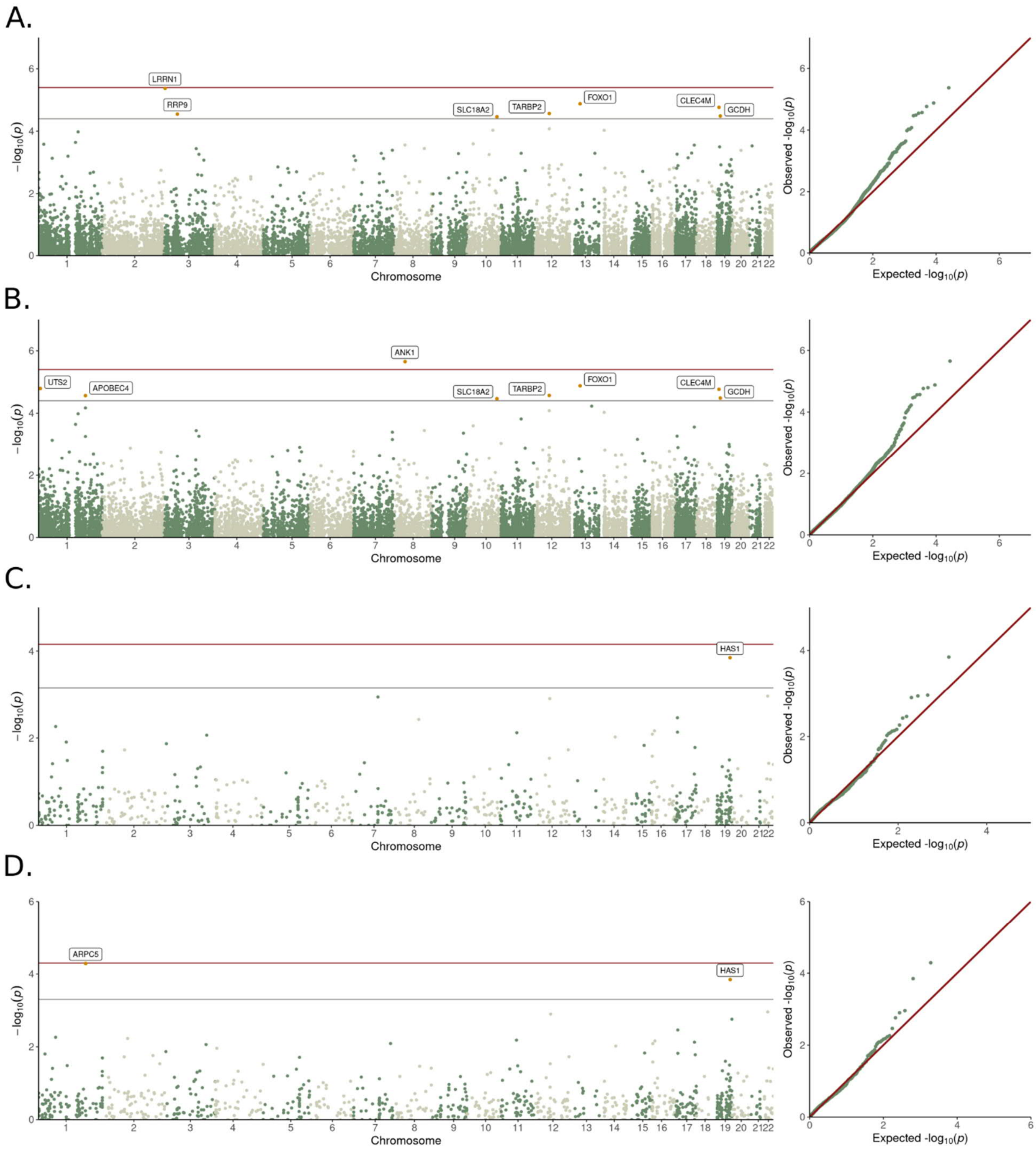

Figure S11: SKAT-O with additional DKD adjustment Manhattan plot

**A.** Protein altering variant (PAV) ≤1%, **B.** PAV≤5%, **C.** Protein truncating variant (PTV) ≤1%, **D.** PTV≤5%.

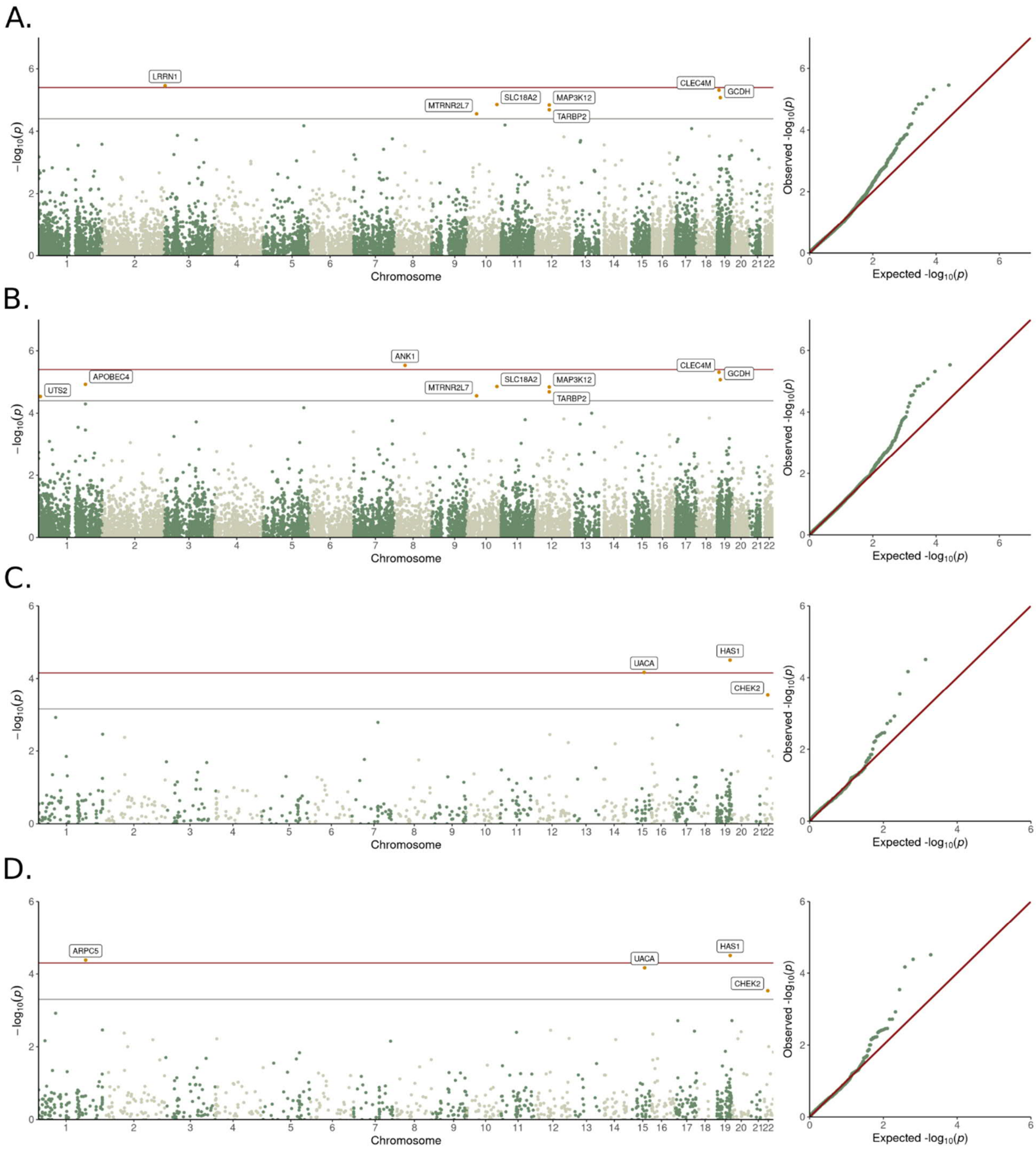

### Figure S12: *MAP3K12* and *TARBP2* regional plot

Chromosomal positions, genes and gene transcripts. Protein altering variants within SKAT-O tests are highlighted, and statistical significance are presented with the additional DKD adjustment.

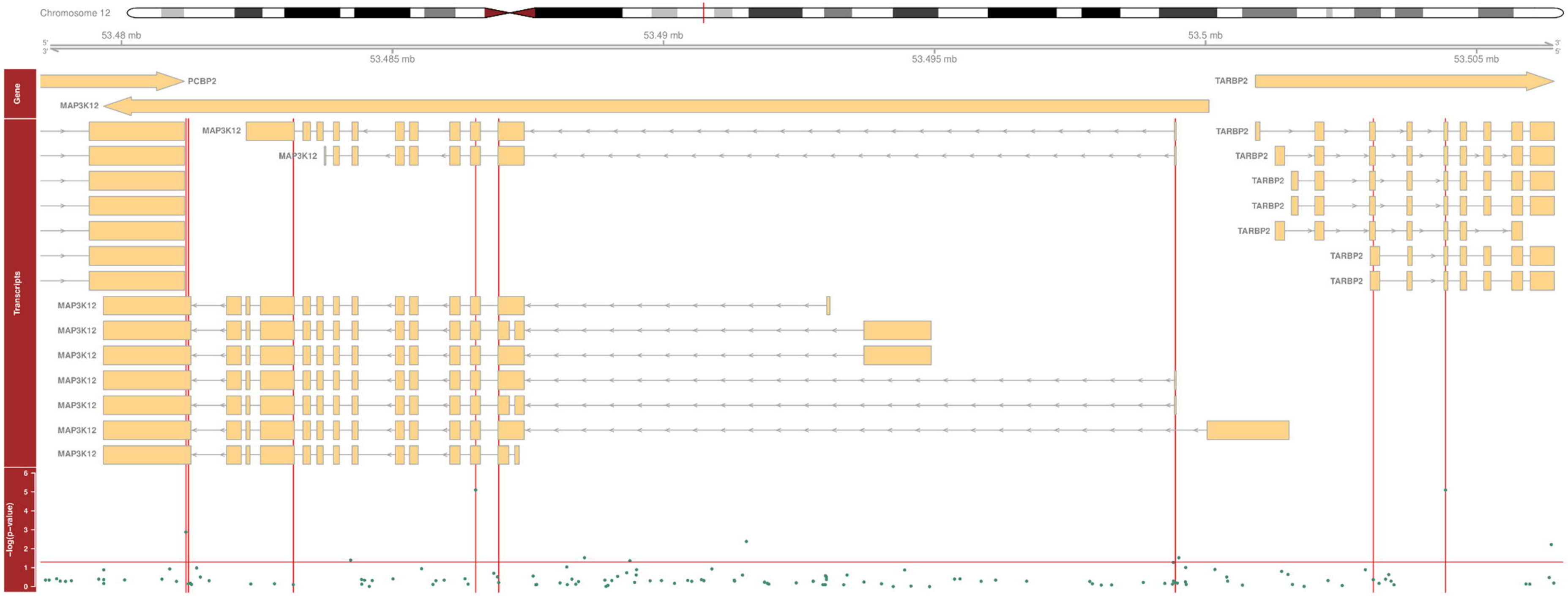

Figure S13: Known Mendelian stroke-risk genes in T1D

**A.** SKAT-O *p*-value, **B.** $N_{variant}$ (MAC) in SKAT-O. Results presented with minimal adjustment.

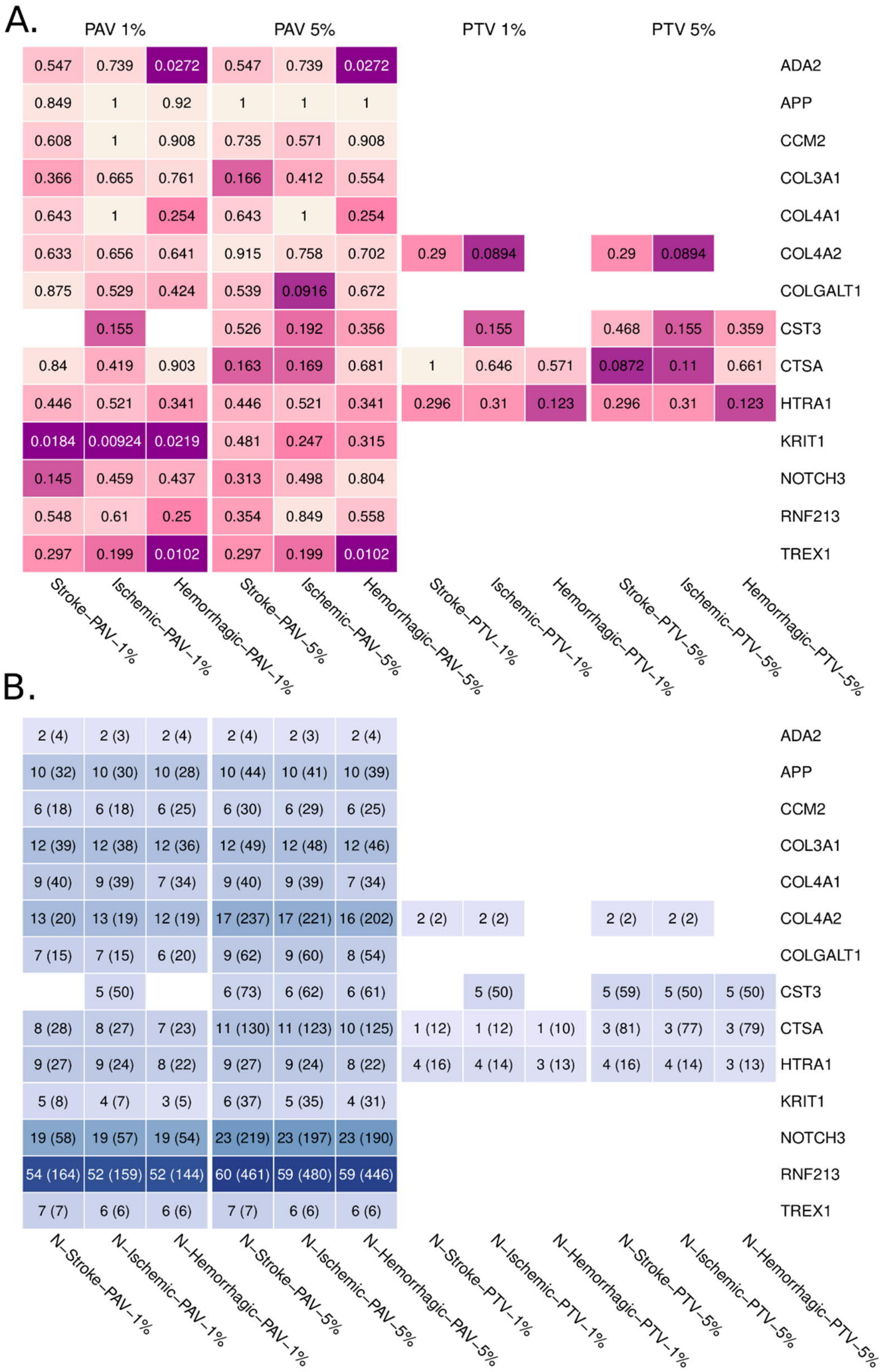

### Figure S14: Topologically associating domain (TAD) on 4q33-34.1

**A.** Frontal lobe, **B.** Hippocampus. The region has been predicted to locate on the same TAD with *GALNTL6* promoter as well as *AADAT* and *MFAP3L* distal promoters. The identified windows are within 170,752,001-171,082,000 (highlighted with light blue); and the top variant is 4:170787127.

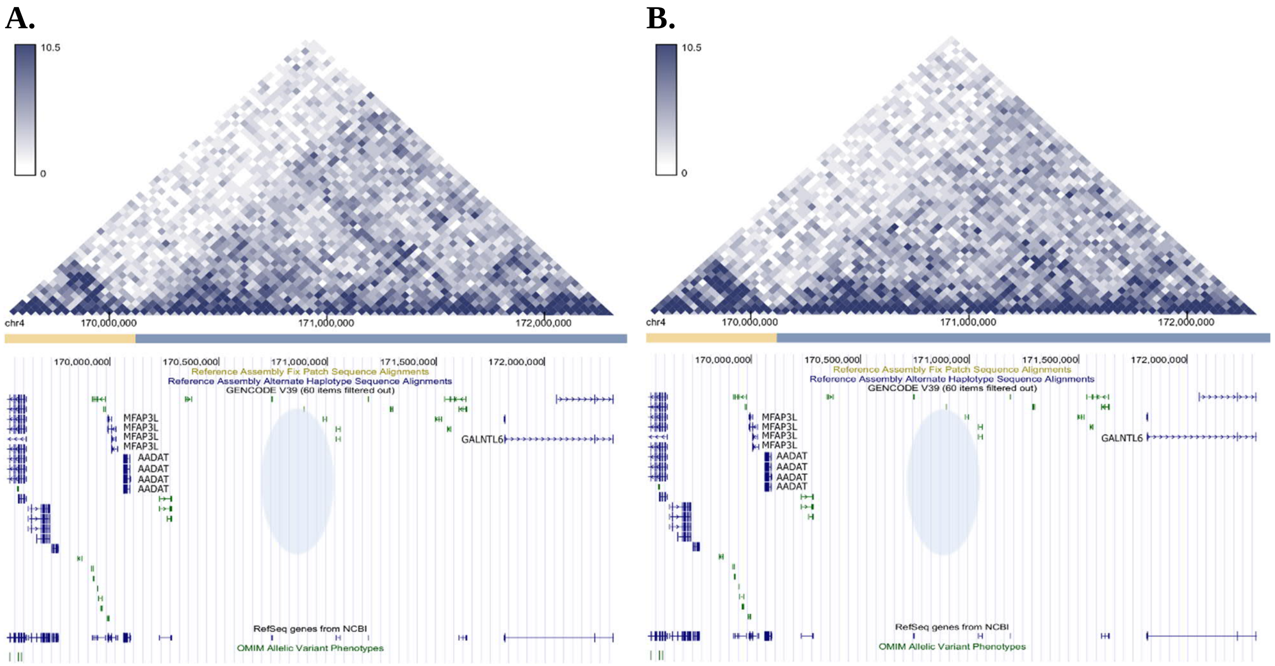

#### Figure S15: Enhancer variant PCHi-C links

**A.** *BDNF* in hippocampus, and **B.** An intronic sliding-window region, on *LINC01500*, links to *DACT1*.

**A.**

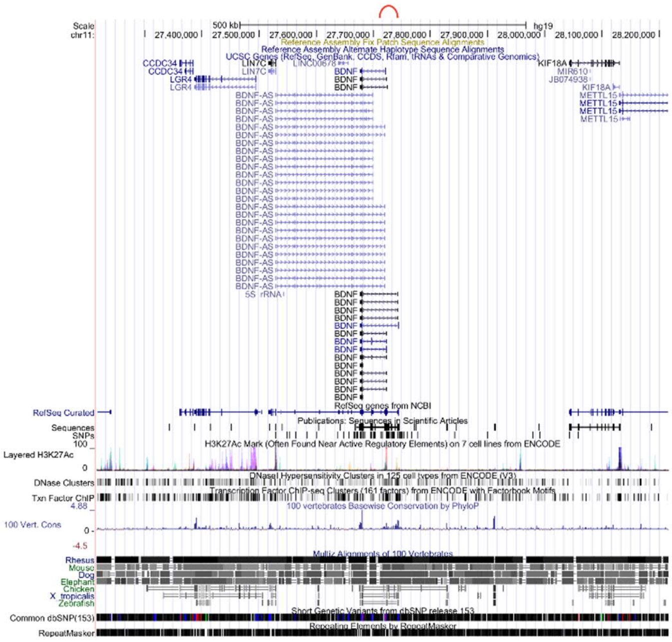

**B.**

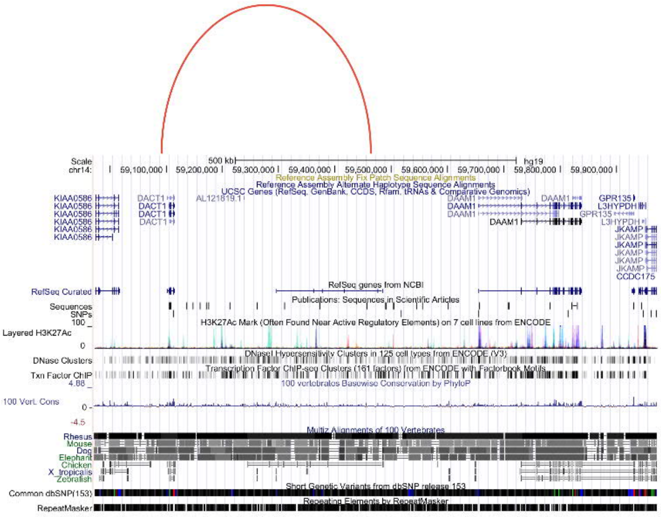

### Figure S16: Stroke enhancer association Manhattan plot

Variants weighted with PHRED scale minor allele frequencies. **A.** MAF≤5%, and **B.** MAF≤1%.

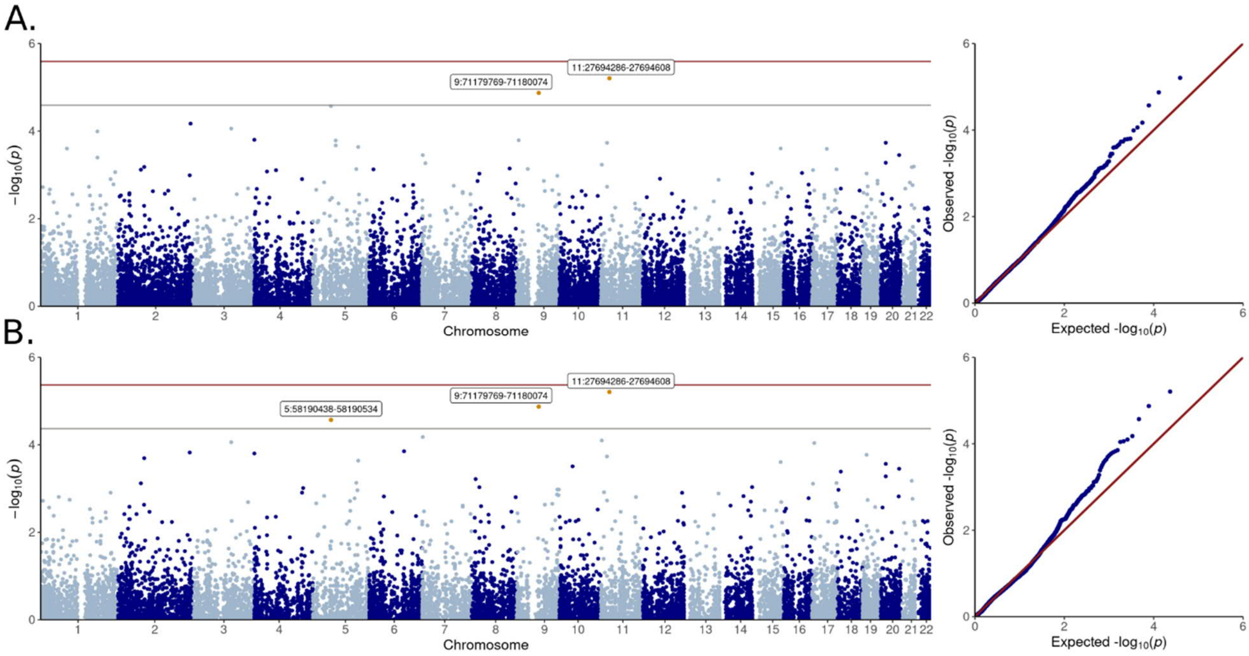

#### Figure S17: Stroke promoter association Manhattan plot

Variants are weighted with PHRED scale minor allele frequencies. **A.** MAF≤5%, and **B.** MAF≤1%.

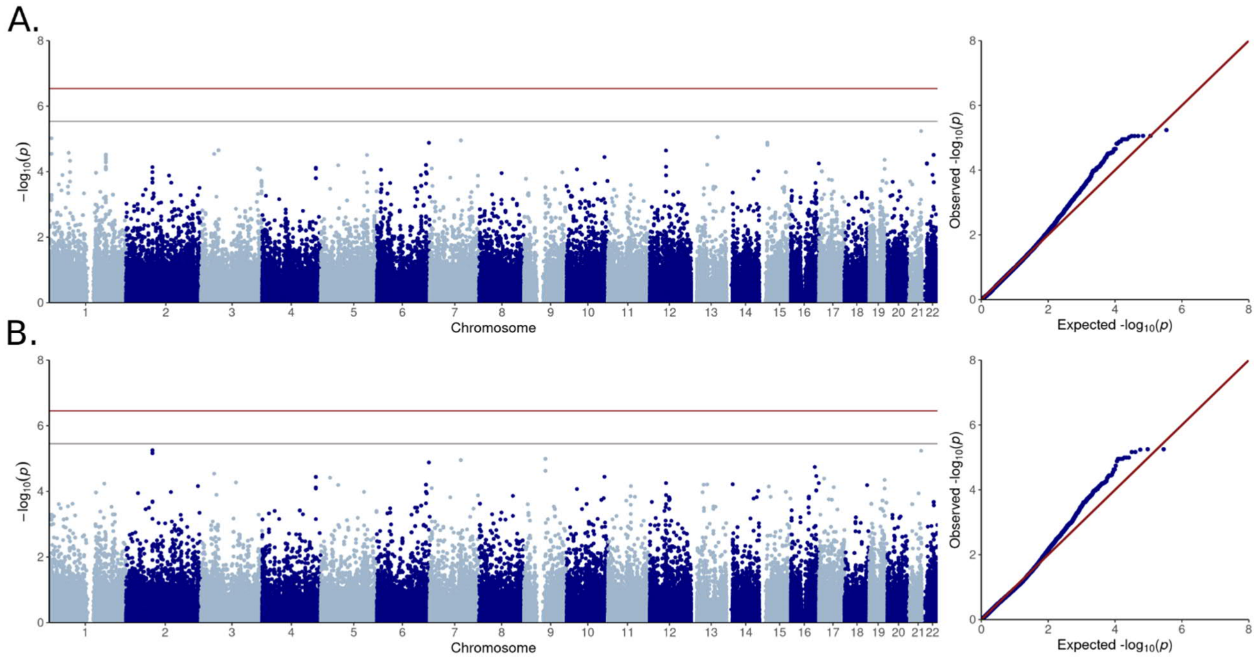

Figure S18: *TRPM2-AS* regional plot and experimental data

**A.** Regional plot of *TRPM2* and *TRPM2-AS* extended region; the discovered promoter is highlighted (red). **B.** *TRPM2-AS* expression in HELA, HEK-293 and HUVEC cell lines, **C.** Relative expression of *TRPM2-AS* and *TRPM2* transcripts in HELA cells (hypoxanthine phosphoribosyltransferase 1 used as reference transcript to normalize quantitative RT-PCR), **D.** Firefly/Renilla luciferase assay of promoter activity. Empty vector (mean=1.0, 12 technical repeats) as transfection control for baseline luciferase activity was compared to *TRPM2-AS* control promoter, i.e., major allele in all identified *TRPM2-AS* variants (mean=56.5, 11 technical repeats, *p*-value=0.00022); which was further compared to a *TRPM2-AS* promoter with rs753589764 minor allele (mean=72.6, 11 technical repeats, *p*-value=0.27): All cloned before firefly reporter gene to evaluate potential transcriptional promoter activity. Statistical significance was assessed with Student's t-test and error bars represent standard error.

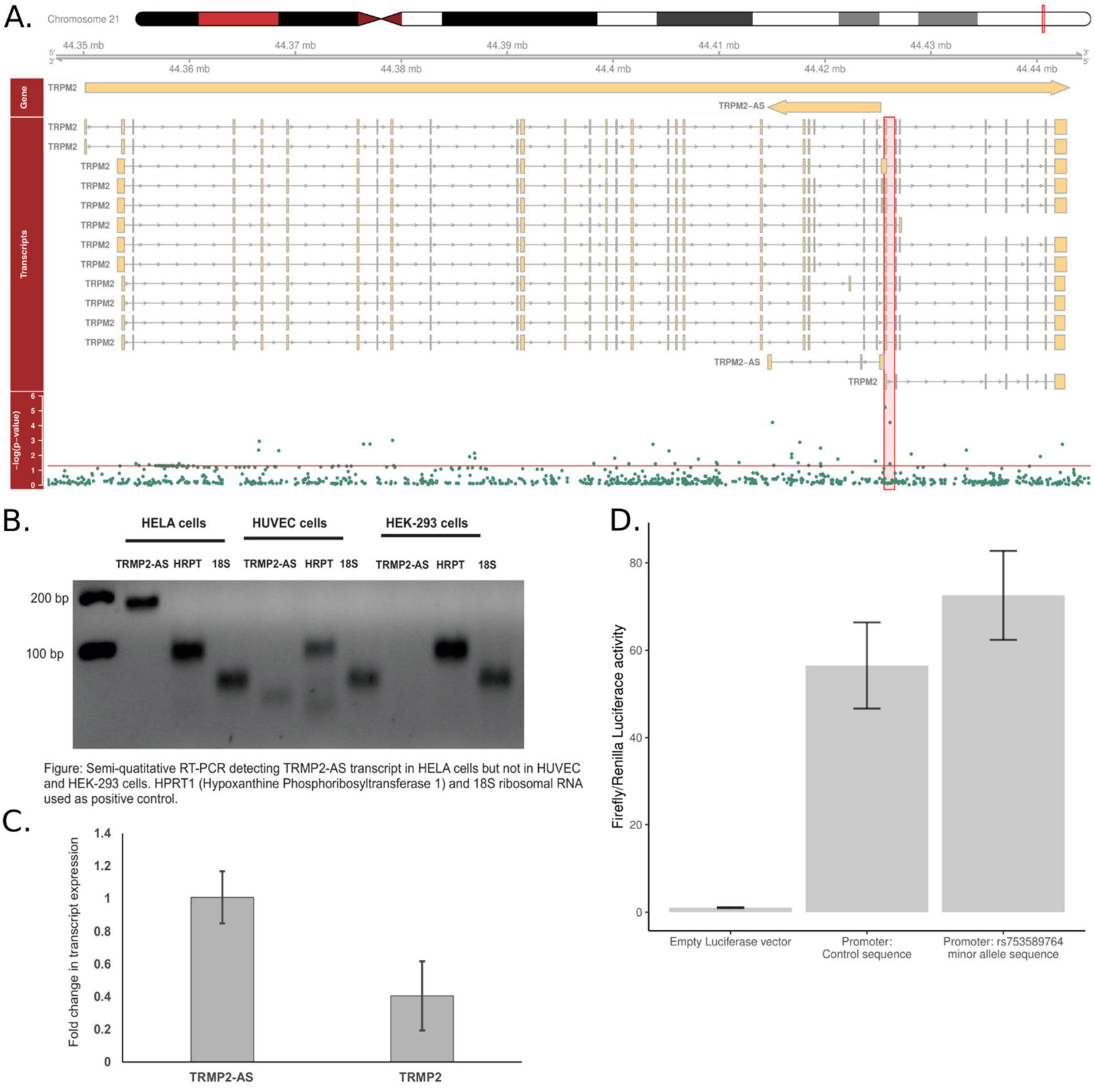

#### Figure S19: Sequencing data processing pipeline (WES and WGS)

**1.** Genomes were sequenced with Illumina HiSeqX and Illumina HiSeq2000 next-generation sequencing, respectively. **2.** Genomes were trimmed with Trimmomatic 0.36 software[2], and read quality was assessed with FastQC software[32]. Next, we aligned reads with GATK's (4.0.1.1) Burrows-Wheeler Aligner (BWA), sorted and marked duplicate reads with Picard's SortSam and MarkDuplicates tools. We recalibrated bases by chromosome with GATK's VQSR and ApplyVQSR tools. **3.** Variants were called by sample with GATK's HaplotypeCaller using ERC mode, then WES and WGS were jointcalled into combined variant call format files with GATK's CombineGVCFs and GenotypeGVCF. 490 exomes passed quality control within WES: Two samples were excluded due to excess heterozygosity (>3 standard deviations from sample mean), one due to relatedness, one due to discordance with FinnDiane GWAS, and one due to being also whole genome sequenced. 583 samples passed QC within WGS: Five samples were excluded due to excess heterozygosity, eight due to failed percentage of mapped deduplicated reads (<91%), and three due to discordance with FinnDiane GWAS. We further performed quality control on individual variants.

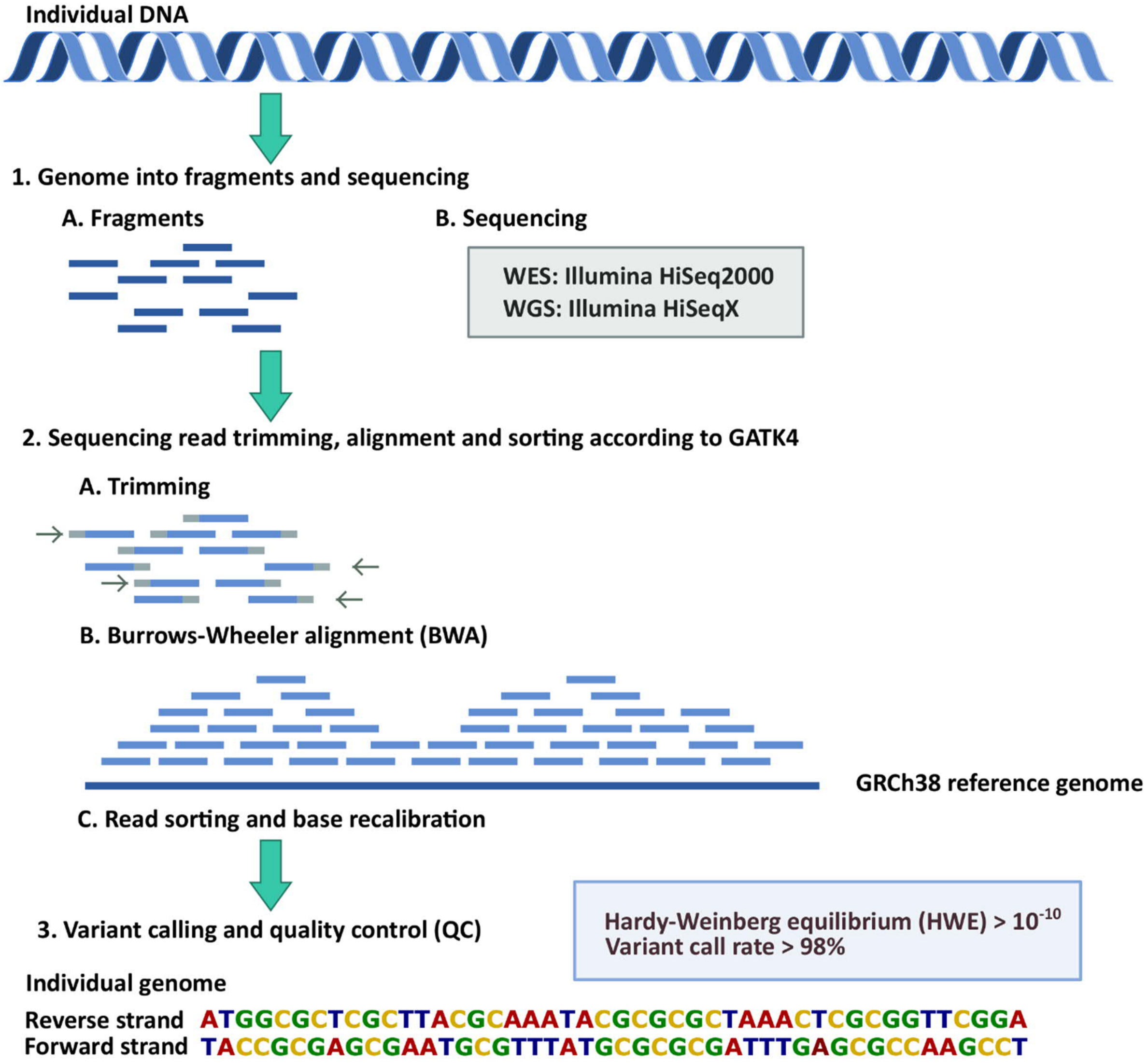

#### Figure S20: GWAS replication data processing pipeline (FinnDiane)

The data had been previously processed to GRCh37 reference genome, importantly, variants had been called with zCall software[33] and low genotyping quality variants excluded. We have now shifted the genotyping positions from GRCh37 to GRCh38 with Picard's LiftoverVCF tool and merged the genotyping batches. Variants with high missingness (>2%), low HWE *p*-value ($<10^{-6}$), or minor allele count <3 were removed. Three individuals were excluded due to ambiguous gender, and six due to high genotype missingness rate (>5%) or heterozygosity (4 SDs from sample mean). Next, the chip genotyping data was pre-phased with Eagle 2.3.5 software[34], imputed to SISu v3 reference panel with Beagle 4.1 software[35], and annotated with SNPEff version 5 software[4]. FIMM HumGen Sequencing Informatics genotype imputation workflow v3.0 V2 was followed.

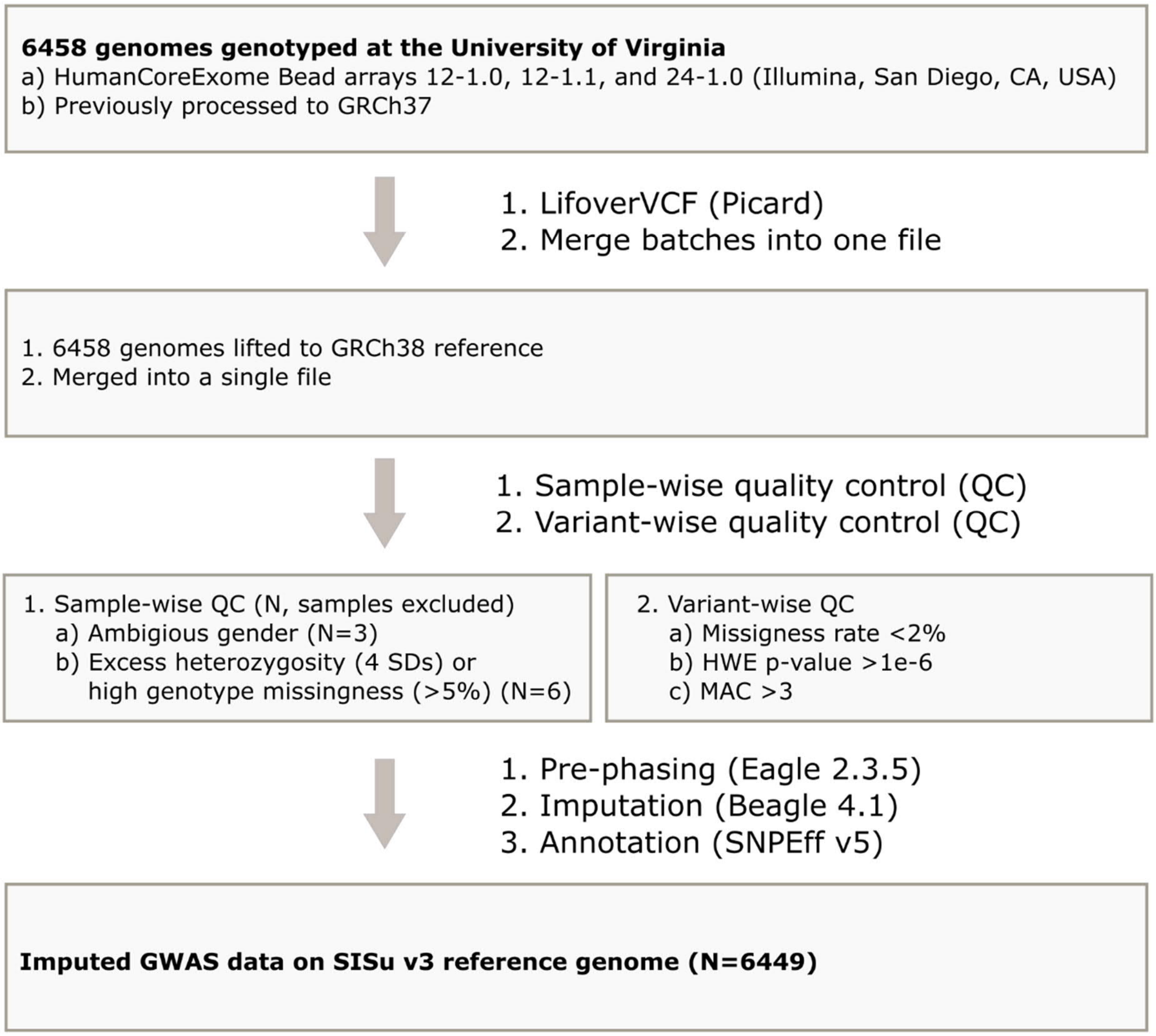

### Supplemental Tables

#### Table S1: Stroke subtype sequencing data clinical characteristics

**A.** Ischemic stroke, **B**. Hemorrhagic stroke. Weighted mean HbA1c is calculated until the stroke event or the end of follow-up. DKD = End-stage renal disease, macro- or microalbuminuria. Mean (SD), *Median (IQR). Student's t-test, Wilcoxon signed rank test or Fisher's exact test.

**A.**

| | WES | | | WGS | | |
|---|---|---|---|---|---|---|
| | **Cases** | **Controls** | ***p*-value** | **Cases** | **Controls** | ***p*-value** |
| N | 49 | 406 | | 64 | 459 | |
| CVD death <2017 (yes/no, yes-%) | 22/27 (45%) | 66/340 (16%) | $1.20 \times 10^{-5}$ | 31/33 (48%) | 83/376 (18%) | $3.93 \times 10^{-7}$ |
| Sex (male/female, %-males) | 26/23 (53%) | 182/224 (45%) | 0.29 | 45/19 (70%) | 236/223 (51%) | 0.0048 |
| Age | 49.40 (12.46) | 60.69 (11.13) | $1.12 \times 10^{-7}$ | 51.49 (10.12) | 58.70 (9.61) | $7.63 \times 10^{-7}$ |
| T1D Duration | 33.63 (10.31) | 47.49 (9.84) | $1.45 \times 10^{-12}$ | 36.63 (9.15) | 46.00 (8.14) | $2.59 \times 10^{-11}$ |
| Calendar year of diabetes onset* | 1971 (11) | 1968 (11) | 0.21 | 1967.5 (12) | 1969 (9) | 0.41 |
| T1D onset age | 15.77 (7.26) | 13.21 (7.27) | 0.023 | 14.86 (8.79) | 12.69 (7.75) | 0.065 |
| Weighted mean HbA1c* | 9.1 (2.11) | 8.51 (1.40) | 0.015 | 8.86 (1.74) | 8.34 (1.57) | 0.0062 |
| HbA1c count* | 23 (31) | 29 (26) | 0.039 | 19 (28.75) | 29 (30) | 0.068 |
| DKD (yes/no, yes-%) | 35/14 (71%) | 201/205 (50%) | 0.0039 | 53/11 (83%) | 206/253 (45%) | $6.76 \times 10^{-9}$ |

**B.**

| | WES | | | WGS | | |
|---|---|---|---|---|---|---|
| | **Cases** | **Controls** | ***p*-value** | **Cases** | **Controls** | ***p*-value** |
| N | 22 | 406 | | 26 | 459 | |
| CVD death <2017 (yes/no, yes-%) | 12/10 (55%) | 66/340 (16%) | $8.46 \times 10^{-5}$ | 16/10 (62%) | 83/376 (18%) | $2.75 \times 10^{-6}$ |
| Sex (male/female, %-males) | 13/9 (59%) | 182/224 (45%) | 0.27 | 19/7 (73%) | 236/223 (51%) | 0.042 |
| Age | 45.98 (10.97) | 60.69 (11.13) | $2.82 \times 10^{-6}$ | 49.42 (11.34) | 58.7 (9.61) | 0.00035 |
| T1D Duration | 32.96 (11.30) | 47.49 (9.84) | $5.24 \times 10^{-6}$ | 36.72 (10.81) | 46.00 (8.14) | 0.00020 |
| Calendar year of T1D onset* | 1968.5 (11.25) | 1968 (11) | 0.90 | 1968 (12.5) | 1969 (9) | 0.85 |
| T1D onset age | 13.02 (5.02) | 13.21 (7.27) | 0.87 | 12.7 (8.37) | 12.69 (7.75) | 1.00 |
| Weighted mean HbA1c* | 8.99 (0.45) | 8.51 (1.40) | 0.050 | 8.92 (1.91) | 8.34 (1.57) | 0.15 |
| HbA1c count* | 13.5 (12.75) | 29 (26) | 0.010 | 22 (25.75) | 29 (30) | 0.092 |
| DKD (yes/no, yes-%) | 15/7 (68%) | 201/205 (50%) | 0.12 | 20/6 (77%) | 206/253 (45%) | 0.0019 |

#### Table S2: Stroke ICD codes in Finnish registry data

For patients in sequencing cohort without data verified by neurologists ($N_{WGS}$=27, $N_{WES}$=2), we considered only the registry data as follows: Stroke was defined as *Stroke, severe* (I60, I61, I62, I63, I64, 430, 431, 432, 433, 434), and we excluded *Stroke, mild* (I65, I66, I67, I68, I69, 435, 436, 437, 438, TIA G45) from controls in order to ensure a clean phenotype (WGS N=5, WES N=0). In T1D specific replication data (GWAS and genotyping), registry-based stroke events were partly verified by neurologists, and whenever the verified data were unavailable, we defined stroke as *Stroke, severe* (I60, I61, I62, I63, I64, 430, 431, 432, 433, 434). Similarly, *Stroke, mild* (I65, I66, I67, I68, I69, 435, 436, 437, 438, TIA G45) were excluded from controls. Controls were followed until death or the end of 2017.

| **Stroke, severe** | | **Stroke mild** | |
|---|---|---|---|
| **Explanation** | **ICD code** | **Explanation** | **ICD code** |
| Subarachnoid hemorrhage | I60 | Occlusion and stenosis of precerebral arteries not resulting in cerebral infarction | I65 |
| Intracerebral hemorrhage | I61 | Occlusion and stenosis of cerebral arteries not resulting in cerebral infarction | I66 |
| Other nontraumatic intracranial haemorrhage | I62 | Other cerebrovascular diseases | I67 |
| Cerebral infarction | I63 | Cerebrovascular disorders in diseases classified elsewhere | I68 |
| Stroke, not specified as hemorrhage or infarction | I64 | Sequellae of cerebrovascular disease | I69 |
| Subarachnoid hemorrhage | 430 | Transient cerebral ischemia | 435 |
| Intracerebral hemorrhage | 431 | Acute but ill-defined cerebrovascular disease | 436 |
| Other and unspecified intracranial hemorrhage | 432 | Other and ill-defined cerebrovascular disease | 437 |
| Occlusion and stenosis of precerebral arteries | 433 | Late effects of cerebrovascular disease | 438 |
| Occlusion of cerebral arteries | 434 | Transient cerebral ischemic attacks and related syndromes | TIA G45 |

Table S3: GWAS replication clinical characteristics (FinnDiane)

Out of the stroke cases, 206 were verified by trained neurologists and 161 were based on registry data. Weighted mean HbA1c is calculated until the stroke event or the end of follow-up. DKD = End-stage renal disease, macro- or microalbuminuria. Mean (SD), *Median (IQR). Student's t-test, Wilcoxon signed rank test or Fisher's exact test.

| | Stroke | | |
|---|---|---|---|
| | **Cases** | **Controls** | ***p*-value** |
| N | 367 | 3578 | |
| Hemorrhagic/Ischemic | 40/164 | | |
| CVD death <2017 (yes/no, yes-%) | 160/207 (44%) | 232/3346 (6.5%) | $2.70 \times 10^{-74}$ |
| Sex (male/female, %-males) | 225/142 (61%) | 1819/1759 (51%) | 0.00015 |
| Age | 51.02 (11.00) | 52.83 (10.28) | 0.0028 |
| T1D Duration | 35.14 (11.20) | 36.55 (9.93) | 0.021 |
| Calendar year of T1D onset* | 1968 (15) | 1981 (15) | $8.53 \times 10^{-60}$ |
| T1D onset age* | 14.21 (14.88) | 14.48 (13.22) | 0.23 |
| Weighted mean HbA1c | 9.10 (1.33) | 8.33 (1.14) | $2.94 \times 10^{-19}$ |
| HbA1c count* | 17 (27) | 22 (27) | 0.0024 |
| DKD (yes/no, yes-%) | 296/56 (84%) | 1305/2108 (38%) | $7.24 \times 10^{-64}$ |

### Table S4: GWAS stroke subtype replication clinical characteristics

Weighted mean HbA1c is calculated until the stroke event or the end of follow-up. DKD = End-stage renal disease, macro- or microalbuminuria. Mean (SD), *Median (IQR). Student's t-test, Wilcoxon signed rank test or Fisher's exact test.

| | Hemorrhagic stroke | | | Ischemic stroke | | |
|---|---|---|---|---|---|---|
| | **Cases** | **Controls** | **p-value** | **Cases** | **Controls** | **p-value** |
| | 40 | 3578 | | 164 | 3578 | |
| CVD death <2017 (yes/no, yes-%) | 20/20 (50%) | 232/3346 (6.5%) | $1.36\times10^{-13}$ | 72/92 (44%) | 232/3346 (6.5%) | $7.18\times10^{-38}$ |
| Sex (male/female, %-males) | 27/13 (68%) | 1819/1759 (51%) | 0.039 | 108/56 (66%) | 1819/1759 (51%) | 0.00016 |
| Age | 49.96 (7.31) | 52.83 (10.28) | 0.019 | 52.23 (10.85) | 52.83 (10.28) | 0.49 |
| T1D Duration | 35.84 (9.37) | 36.55 (9.93) | 0.64 | 35.09 (10.53) | 36.55 (9.93) | 0.083 |
| Calendar year of T1D onset* | 1965.5 (13.25) | 1981 (15) | $1.00\times10^{-10}$ | 1968 (13.25) | 1981 (15) | $4.47\times10^{-32}$ |
| T1D onset age* | 11.33 (11.52) | 14.48 (13.22) | 0.072 | 15.42 (15.63) | 14.48 (13.22) | 0.404 |
| Weighted mean HbA1c | 8.8 (1.43) | 8.33 (1.14) | 0.062 | 9.04 (1.18) | 8.33 (1.14) | $5.73\times10^{-10}$ |
| HbA1c count* | 8 (13.5) | 22 (27) | 0.00046 | 16 (27.25) | 22 (27) | 0.0040 |
| DKD (yes/no, yes-%) | 35/4 (90%) | 1305/2108 (38%) | $4.04\times10^{-11}$ | 131/25 (84%) | 1305/2108 (38%) | $1.88\times10^{-30}$ |

#### Table S5: FinnDiane genotyping clinical characteristics

Out of the stroke cases, 201 were verified by trained neurologists and 102 were based on registry data. Weighted mean HbA1c is calculated until the stroke event or the end of follow-up. DKD = End-stage renal disease, macro- or microalbuminuria. Mean (SD), *Median (IQR). Student's t-test, Wilcoxon signed rank test or Fisher's exact test.

| | Stroke | | |
|---|---|---|---|
| | **Cases** | **Controls** | ***p*-value** |
| N | 303 | 2960 | |
| Hemorrhagic/Ischemic | 39/160 | | |
| CVD death <2017 (yes/no, yes-%) | 131/172 (43%) | 191/2769 (6.5%) | $6.45\times10^{-61}$ |
| Sex (male/female, %-males) | 185/118 (61%) | 1508/1452 (51%) | 0.00088 |
| Age | 50.91 (11.07) | 52.92 (10.42) | 0.0028 |
| T1D Duration | 35.19 (11.26) | 36.60 (10.10) | 0.037 |
| Calendar year of T1D onset* | 1968 (14) | 1981 (15) | $2.35\times10^{-52}$ |
| T1D onset age* | 13.76 (15.50) | 14.39 (13.20) | 0.14 |
| Weighted mean HbA1c | 9.03 (1.25) | 8.35 (1.16) | $8.06\times10^{-15}$ |
| HbA1c count* | 16 (27) | 23 (25) | 0.00030 |
| DKD (yes/no, yes-%) | 247/43 (85%) | 1123/1735 (39%) | $3.77\times10^{-53}$ |

Table S6: Variant type classification for SKAT-O

| | **Protein altering variants (PAV)** | **Protein truncating variants (PTV)** |
|---|---|---|
| **Variant type** | 5 prime UTR premature start codon gain variant | start lost |
| | 5 prime UTR truncation & exon loss variant | stop gained |
| | bidirectional gene fusion | stop lost |
| | gene fusion | bidirectional gene fusion |
| | conservative inframe deletion | gene fusion |
| | conservative inframe insertion | frameshift variant |
| | disruptive inframe deletion | exon loss variant |
| | disruptive inframe insertion | splice acceptor variant |
| | start lost | splice donor variant |
| | stop gained | |
| | stop lost | |
| | exon loss variant | |
| | frameshift variant | |
| | missense variant | |
| | splice acceptor variant | |
| | splice donor variant | |
| | structural interaction variant | |

#### Table S7: CADD functional annotations within annotation classes

Listed are CADD annotation scores, which we used in calculation of annotation principal components (aPC)[15] for STAAR-O sliding-window analyses.

| **Functional score** | **CADD annotations included** |
|---|---|
| **a-PC-Epigenetics** | GC, CpG, EncodeH3K4me1-max, EncodeH3K4me2-max, EncodeH3K4me3-max, EncodeH3K9ac-max, EncodeH3K9me3-max, EncodeH3K27ac-max, EncodeH3K27me3-max, EncodeH3K36me3-max, EncodeH3K79me2-max, EncodeH4K20me1-max, EncodeH2AFZ-max, RemapOverlapTF, RemapOverlapCL, minDistTSS, minDistTSE |
| **aPC-TF** | RemapOverlapTF, RemapOverlapCL |
| **aPC-Conservation** | GerpN, GerpS, priPhCons, mamPhCons, verPhCons, priPhyloP, mamPhyloP, verPhyloP |
| **aPC-Protein-function** | SIFTval, PolyPhenVal, Grantham |
| **aPC-microRNA** | targetScan, mirSVR.Score, mirSVR.E |
| **aPC-Mutation-density** | Freq100bp, Rare100bp, Sngl100bp, Freq1000bp, Rare1000bp, Sngl1000bp, Freq10000bp, Rare10000bp, Sngl10000bp |
| **aPC-TES-TSS-proximity** | minDistTSS, minDistTSE |

#### Table S8: Genotyped variants in FinnDiane

Variants were genotyped for 3,600 individuals (including the positive control), although replication by genotyping entailed only individuals within FinnDiane GWAS data – thus, the kinship matrix – and those with available stroke phenotype and fulfilled control criteria (i.e., age >35 years, diabetes duration >20 years, and no mild strokes in registry data, if data verified neurologists not available) (N=3,263). One mutation carrier was selected from sequencing data to be the positive control, and the patient was later excluded from the replication analyses.

| Variant | REF | ALT | rsnumber | Sequenced | ALT AC[†] |
|---|---|---|---|---|---|
| 1:183648589* | GT | G | . | OK | 0 |
| 1:183653304* | T | C | rs1361824345 | OK | 0 |
| 15:70679607 | C | A | rs185763236 | OK | ≤3 |
| 17:17819659 | G | A | rs114001633 | OK | 16 |
| 19:12897009* | G | A | rs761135089 | OK | 0 |
| 19:51716967 | C | T | rs1396240967 | OK | 0 |
| 19:7766121* | A | G | rs144783051 | OK | 10 |
| 19:7767549 | C | T | rs752738017 | OK | ≤3 |
| 3:3845194 | C | T | rs142381203 | OK | ≤3[‡] |
| 3:3845554 | C | T | rs141825989 | OK | ≤3 |
| 3:3845966 | C | T | rs747455683 | OK | 0 |
| 19:7766659 | G | A | rs140767813 | OK | ≤3 |

*Added for runs 4-10.

†Alternative allele count in all genotyped individuals (excluding the positive control, $N_{total}$=3,599).

‡Analysed in R with linear regression due to no alternative allele carriers after kinship matrix adjustment criteria (N=3,263)

#### Table S9: FinnGen replication ICD codes (GWAS)

Stroke, including SAH, is the main general population replication phenotype. We performed replication also with additional FinnGen stroke phenotypes, most importantly; ischemic stroke (*Ischemic stroke, excluding all hemorrhages*) and hemorrhages (*Nontraumatic intracranial hemorrhages*), but also (*Stroke, excluding SAH*).

| **Phenotype** | **Name** | **ICD codes** |
|---|---|---|
| Nontraumatic intracranial hemorrhage | I9_INTRACRA | I60-61, 430-431 |
| Ischemic stroke, excluding all hemorrhages | I9_STR_EXH | I63-64, 433-434, 436 |
| Stroke, excluding SAH | I9_STR | I61, I63-64, 431, 433-434, 436 |
| Stroke, including SAH | I9_STR_SAH | I60-61, I63-64, 430-431, 433-434, 436 |

Table S24: Physicians and nurses at health care centers participating in the collection of FinnDiane patients

| FinnDiane Study Centers | Physicians and nurses |
|---|---|
| **Anjalankoski Health Centre** | S. Koivula, T. Uggeldahl |
| **Central Finland Central Hospital, Jyväskylä** | T. Forslund, A. Halonen, A. Koistinen, P. Koskiaho, M. Laukkanen, J. Saltevo, M. Tiihonen |
| **Central Hospital of Åland Islands, Mariehamn** | M. Forsen, H. Granlund, A-C. Jonsson, B. Nyroos |
| **Central Hospital of Kanta-Häme, Hämeenlinna** | P. Kinnunen, A. Orvola, T. Salonen, A. Vähänen |
| **Central Hospital of Länsi-Pohja, Kemi** | H. Laukkanen, P. Nyländen, A. Sademies |
| **Central Ostrabothnian Hospital District, Kokkola** | S. Anderson, B. Asplund, U. Byskata, P. Liedes, M. Kuusela, T. Virkkala |
| **City of Espoo Health Centre** | |
| **Espoonlahti** | A. Nikkola, E. Ritola |
| **Tapiola** | M. Niska, H. Saarinen |
| **Samaria** | E. Oukko-Ruponen, T. Virtanen |
| **Viherlaakso** | A. Lyytinen |
| **City of Helsinki Health Centre** | |
| **Puistola** | H. Kari, T. Simonen |
| **Suutarila** | A. Kaprio, J. Kärkkäinen, B. Rantaeskola |
| **Töölö** | P. Kääriäinen, J. Haaga, A-L. Pietiläinen |
| **City of Hyvinkää Health Centre** | S. Klemetti, T. Nyandoto, E. Rontu, S. Satuli-Autere |
| **City of Vantaa Health Centre** | |
| **Korso** | R. Toivonen, H. Virtanen |
| **Länsimäki** | R. Ahonen, M. Ivaska-Suomela, A. Jauhiainen |
| **Martinlaakso** | M. Laine, T. Pellonpää, R. Puranen |
| **Myyrmäki** | A. Airas, J. Laakso, K. Rautavaara |
| **Rekola** | M. Erola, E. Jatkola |
| **Tikkurila** | R. Lönnblad, A. Malm, J. Mäkelä, E. Rautamo |
| **Heinola Health Centre** | P. Hentunen, J. Lagerstam |
| **Helsinki University Central Hospital, Department of Medicine, Division of Nephrology** | A. Ahola, J. Fagerudd, M. Feodoroff, D. Gordin, O. Heikkilä, K Hietala, L. Kyllönen, J. Kytö, S. Lindh, K. Pettersson-Fernholm, M. Rosengård-Bärlund, M. Rönnback, A. Sandelin, A-R Salonen, L. Salovaara, L. Thorn, J. Tuomikangas, T. Vesisenaho, J. Wadén |
| **Herttoniemi Hospital, Helsinki** | V. Sipilä |
| **Hospital of Lounais-Häme, Forssa** | T. Kalliomäki, J. Koskelainen, R. Nikkanen, N. Savolainen, H. Sulonen, E. Valtonen |
| **Iisalmi Hospital** | E. Toivanen |
| **Jokilaakso Hospital, Jämsä** | A. Parta, I. Pirttiniemi |
| **Jorvi Hospital, Helsinki University Central Hospital** | S. Aranko, S. Ervasti, R. Kauppinen-Mäkelin, A. Kuusisto, T. Leppälä, K. Nikkilä, L. Pekkonen |
| **Jyväskylä Health Centre, Kyllö** | K. Nuorva, M. Tiihonen |
| **Kainuu Central Hospital, Kajaani** | S. Jokelainen, P. Kemppainen, A-M. Mankinen, M. Sankari |
| **Kerava Health Centre** | H. Stuckey, P. Suominen |
| **Kirkkonummi Health Centre** | A. Lappalainen, M. Liimatainen, J. Santaholma |
| **Kivelä Hospital, Helsinki** | A. Aimolahti, E. Huovinen |
| **Koskela Hospital, Helsinki** | V. Ilkka, M. Lehtimäki |
| **Kotka Heath Centre** | E. Pälikkö-Kontinen, A. Vanhanen |

| | |
|---|---|
| **Kouvola Health Centre** | E. Koskinen, T. Siitonen |
| **Kuopio University Hospital** | E. Huttunen, R. Ikäheimo, P. Karhapää, P. Kekäläinen, M. Laakso, T. Lakka, E. Lampainen, L. Moilanen, L. Niskanen, U. Tuovinen, I. Vauhkonen, E. Voutilainen |
| **Kuusamo Health Centre** | T. Kääriäinen, E. Isopoussu |
| **Kuusankoski Hospital** | E. Kilkki, I. Koskinen, L. Riihelä |
| **Laakso Hospital, Helsinki** | T. Meriläinen, P. Poukka, R. Savolainen, N. Uhlenius |
| **Lahti City Hospital** | A. Mäkelä, M. Tanner |
| **Lapland Central Hospital, Rovaniemi** | L. Hyvärinen, S. Severinkangas, T. Tulokas |
| **Lappeenranta Health Centre** | P. Linkola, I. Pulli |
| **Lohja Hospital** | T. Granlund, M. Saari, T. Salonen |
| **Loimaa Health Centre** | A. Mäkelä, P. Eloranta |
| **Länsi-Uusimaa Hospital, Tammisaari** | I-M. Jousmaa, J. Rinne |
| **Malmi Hospital, Helsinki** | H. Lanki, S. Moilanen, M. Tilly-Kiesi |
| **Mikkeli Central Hospital** | A. Gynther, R. Manninen, P. Nironen, M. Salminen, T. Vänttinen |
| **Mänttä Regional Hospital** | I. Pirttiniemi, A-M. Hänninen |
| **North Karelian Hospital, Joensuu** | U-M. Henttula, P. Kekäläinen, M. Pietarinen, A. Rissanen, M. Voutilainen |
| **Nurmijärvi Health Centre** | A. Burgos, K. Urtamo |
| **Oulankangas Hospital, Oulainen** | E. Jokelainen, P-L. Jylkkä, E. Kaarlela, J. Vuolaspuro |
| **Oulu Health Centre** | L. Hiltunen, R. Häkkinen, S. Keinänen-Kiukaanniemi |
| **Oulu University Hospital** | R. Ikäheimo |
| **Päijät-Häme Central Hospital** | H. Haapamäki, A. Helanterä, S. Hämäläinen, V. Ilvesmäki, H. Miettinen |
| **Palokka Health Centre** | P. Sopanen, L. Welling |
| **Pieksämäki Hospital** | V. Javtsenko, M. Tamminen |
| **Pietarsaari Hospital** | M-L. Holmbäck, B. Isomaa, L. Sarelin |
| **Pori City Hospital** | P. Ahonen, P. Merensalo, K. Sävelä |
| **Porvoo Hospital** | M. Kallio, B. Rask, S. Rämö |
| **Raahe Hospital** | A. Holma, M. Honkala, A. Tuomivaara, R. Vainionpää |
| **Rauma Hospital** | K. Laine, K. Saarinen, T. Salminen |
| **Riihimäki Hospital** | P. Aalto, E. Immonen, L. Juurinen |
| **Salo Hospital** | A. Alanko, J. Lapinleimu, P. Rautio, M. Virtanen |
| **Satakunta Central Hospital, Pori** | M. Asola, M. Juhola, P. Kunelius, M-L. Lahdenmäki, P. Pääkkönen, M. Rautavirta |
| **Savonlinna Central Hospital** | E. Korpi-Hyövälti, T. Latvala, E. Leijala |
| **South Karelia Central Hospital, Lappeenranta** | T. Ensala, E. Hussi, R. Härkönen, U. Nyholm, J. Toivanen |
| **Tampere Health Centre** | A. Vaden, P. Alarotu, E. Kujansuu, H. Kirkkopelto-Jokinen, M. Helin, S. Gummerus, L. Calonius, T. Niskanen, T. Kaitala, T. Vatanen |
| **Tampere University Hospital** | I. Ala-Houhala, T. Kuningas, P. Lampinen, M. Määttä, H. Oksala, T. Oksanen, K. Salonen, H. Tauriainen, S. Tulokas |
| **Tiirismaa Health Centre, Hollola** | T. Kivelä, L, Petlin, L. Savolainen |
| **Turku Health Centre** | I. Hämäläinen, H. Virtamo, M. Vähätalo |
| **Turku University Central Hospital** | K. Breitholz, R. Eskola, K. Metsärinne, U. Pietilä, P. Saarinen, R. Tuominen, S. Äyräpää |
| **Vaajakoski Health Centre** | K. Mäkinen, P. Sopanen |
| **Valkeakoski Regional Hospital** | S. Ojanen, E. Valtonen, H. Ylönen, M. Rautiainen, T. Immonen |
| **Vammala Regional Hospital** | I. Isomäki, R. Kroneld, M. Tapiolinna-Mäkelä |
| **Vaasa Central Hospital** | S. Bergkulla, U. Hautamäki, V-A. Myllyniemi, I. Rusk |

### STROBE checklist

| Item | STROBE guideline | Extension for Genetic Association Studies | Page No |
|---|---|---|---|
| **Title and Abstract** | (a) Indicate the study's design with a commonly used term in the title or the abstract | | Title page, 1 |
| | (b) Provide in the abstract an informative and balanced summary of what was done and what was found | | 1 |
| **Introduction** | | | |
| **Background rationale** | Explain the scientific background and rationale for the investigation being reported | | 3-5 |
| **Objectives** | State specific objectives, including any pre-specified hypotheses | *State if the study is the first report of a genetic association, a replication effort, or both* | 4-5 |
| **Methods** | | | |
| **Study design** | Present key elements of study design early in the paper | | 6-7, Fig. 1 |
| **Setting** | Describe the setting, locations and relevant dates, including periods of recruitment, exposure, follow-up, and data collection | | 6, Sup. Material pages 1-2 |
| **Participants** | (a) *Cohort study:* give the eligibility criteria, and the sources and methods of selection of participants. Describe methods of follow-up | *Give information on the criteria and methods for selection of subsets of participants from a larger study, when relevant* | 6, Sup. Material pages 1-2, Table S2, Table S8 |
| | *Case–control study*: give the eligibility criteria, and the sources and methods of case ascertainment and control selection. Give the rationale for the choice of cases and controls | | |
| | *Cross-sectional study:* give the eligibility criteria, and the sources and methods of selection of participants | | |
| | (b) *Cohort study:* for matched studies, give matching criteria and number of exposed and unexposed | | |
| | *Case–control study*: for matched studies, give matching criteria and the number of controls per case | | |
| **Variables** | (a) Clearly define all outcomes, exposures, predictors, potential confounders, and effect modifiers. Give diagnostic criteria, if applicable | *(b) Clearly define genetic exposures (genetic variants) using a widely-used nomenclature system. Identify variables likely to be associated with population stratification (confounding by ethnic origin)* | 6, Sup. Material pages 1-2, Table 1, Figures S1-S4, Tables S1, S3-S5 |
| **Data sources/ measurement** | (a) For each variable of interest, give sources of data and details of methods of assessment (measurement). Describe comparability of assessment methods if there is more than one group | *(b) Describe laboratory methods, including source and storage of DNA, genotyping methods and platforms (including the allele calling algorithm used, and its version), error rates and call rates. State the laboratory/center where genotyping was done. Describe comparability of laboratory methods if there is more than one group. Specify whether genotypes were assigned using all of the data from the study simultaneously or in smaller batches* | Sup. Material pages 1-4, Figures S18-S20 |
| **Bias** | (a) Describe any efforts to address potential sources of bias | *(b) For quantitative outcome variables, specify if any investigation of potential bias resulting from pharmacotherapy was undertaken. If relevant, describe the nature and magnitude of the potential bias, and explain what approach was used to deal with this* | 7, Sup. Material pages 2-4 |
| **Study size** | Explain how the study size was arrived at | | 6, Sup. Material pages 1-2, Figures S19-S20, Table S8 |
| **Quantitative variables** | Explain how quantitative variables were handled in the analyses. If applicable, describe which groupings were chosen, and why | *If applicable, describe how effects of treatment were dealt with* | 6-7, Sup. Material pages 1-2, Table S2 |
| **Statistical methods** | (a) Describe all statistical methods, including those used to control for confounding | *State software version used and options (or settings) chosen* | 7-9, Sup. Material pages 2-4 |
| | (b) Describe any methods used to examine subgroups and interactions | | |
| | (c) Explain how missing data were addressed | | |
| | *Cohort study*: if applicable, explain how loss to follow-up was addressed | | |
| | *Case–control study*: if applicable, explain how matching of cases and controls was addressed | | |
| | *Cross-sectional study*: if applicable, describe analytical methods taking account of sampling strategy | | |
| | (e) Describe any sensitivity analyses | | |
| | | *(f) State whether Hardy–Weinberg equilibrium was considered and, if so, how* | Sup. Material page 1, Table 2, Figures S19-S20 |
| | | *(g) Describe any methods used for inferring genotypes or haplotypes* | Sup. Material pages 1-2, Figures S19-S20 |

| | | | |
|---|---|---|---|
| | | *(h) Describe any methods used to assess or address population stratification* | 7, 9, Sup. Material pages 2-4 |
| | | *(i) Describe any methods used to address multiple comparisons or to control risk of false-positive findings* | Sup. Material pages 2-4, Figure 2 |
| | | *(j) Describe any methods used to address and correct for relatedness among subjects* | 8-9, Sup. Material pages 2-3 |
| **Results** | | | |
| **Participants** | (a) Report the numbers of individuals at each stage of the study—e.g., numbers potentially eligible, examined for eligibility, confirmed eligible, included in the study, completing follow-up, and analyzed | *Report numbers of individuals in whom genotyping was attempted and numbers of individuals in whom genotyping was successful* | 6, Sup. Material pages 1-2, Table S2, Table S8, Figures S19-S20 |
| | (b) Give reasons for non-participation at each stage | | |
| | (c) Consider use of a flow diagram | | |
| **Descriptive data** | (a) Give characteristics of study participants (e.g., demographic, clinical, social) and information on exposures and potential confounders | *Consider giving information by genotype* | Table 1, Table S1, Tables S3-S5, Figures S1-S4 |
| | (b) Indicate the number of participants with missing data for each variable of interest | | |
| | (c) *Cohort study*: summarize follow-up time, e.g., average and total amount | | |
| **Outcome data** | *Cohort study*: report numbers of outcome events or summary measures over time | *Report outcomes (phenotypes) for each genotype category over time* | Table 1, Table S1, Tables S3-S5, Figures S1-S4 |
| | *Case–control study*: report numbers in each exposure category, or summary measures of exposure | *Report numbers in each genotype category* | |
| | *Cross-sectional study*: report numbers of outcome events or summary measures | *Report outcomes (phenotypes) for each genotype category* | |
| **Main results** | (a) Give unadjusted estimates and, if applicable, confounder-adjusted estimates and their precision (e.g., 95% confidence intervals). Make clear which confounders were adjusted for and why they were included | | 10-16, Tables 2-3, Figure 3, Figure S18, Tables S10-S23 |
| | (b) Report category boundaries when continuous variables were categorized | | |
| | (c) If relevant, consider translating estimates of relative risk into absolute risk for a meaningful time period | | |
| | | *(d) Report results of any adjustments for multiple comparisons* | |
| **Other analyses** | (a) Report other analyses done—e.g., analyses of subgroups and interactions, and sensitivity analyses | | 10-16 Figures 2 and 4 |
| | | *(b) If numerous genetic exposures (genetic variants) were examined, summarize results from all analyses undertaken* | |
| | | *(c) If detailed results are available elsewhere, state how they can be accessed* | 20, Sup. Material page 4 |
| **Discussion** | | | |
| **Key results** | Summarize key results with reference to study objectives | | 17-19 |
| **Limitations** | Discuss limitations of the study, taking into account sources of potential bias or imprecision. Discuss both direction and magnitude of any potential bias | | 19 |
| **Interpretation** | Give a cautious overall interpretation of results considering objectives, limitations, multiplicity of analyses, results from similar studies, and other relevant evidence | | 17-20 |
| **Generalizability** | Discuss the generalizability (external validity) of the study results | | 20 |
| **Other information** | | | |
| **Funding** | Give the source of funding and the role of the funders for the present study and, if applicable, for the original study on which the present article is based | | 20-23 |

### References

1. Sandholm N, Van Zuydam N, Ahlqvist E, Juliusdottir T, Deshmukh HA, Rayner NW, Di Camillo B, Forsblom C, Fadista J, Ziemek D, et al. The Genetic Landscape of Renal Complications in Type 1 Diabetes. *J. Am. Soc. Nephrol.* 2017;28:557–574.

2. Bolger AM, Lohse M, Usadel B. Trimmomatic: a flexible trimmer for Illumina sequence data. *Bioinformatics*. 2014;30:2114–2120.

3. Van der Auwera G, O'Connor B. Genomics in the Cloud: Using Docker, GATK, and WDL in Terra. 1st Edition. O-Reilly Media; 2020.

4. Cingolani P, Platts A, Wang LL, Coon M, Nguyen T, Wang L, Land SJ, Lu X, Ruden DM. A program for annotating and predicting the effects of single nucleotide polymorphisms, SnpEff: SNPs in the genome of Drosophila melanogaster strain w1118; iso-2; iso-3. *Fly (Austin)*. 2012;6:80–92.

5. Salem RM, Todd JN, Sandholm N, Cole JB, Chen W-M, Andrews D, Pezzolesi MG, McKeigue PM, Hiraki LT, Qiu C, et al. Genome-Wide Association Study of Diabetic Kidney Disease Highlights Biology Involved in Glomerular Basement Membrane Collagen. *J. Am. Soc. Nephrol.* 2019;30:2000.

6. Zhan X, Hu Y, Li B, Abecasis GR, Liu DJ. RVTESTS: an efficient and comprehensive tool for rare variant association analysis using sequence data. *Bioinformatics*. 2016;32:1423–1426.

7. Zhou X, Stephens M. Genome-wide efficient mixed-model analysis for association studies. *Nat. Genet.* 2012;44:821–824.

8. Willer CJ, Li Y, Abecasis GR. METAL: fast and efficient meta-analysis of genomewide association scans. *Bioinformatics*. 2010;26:2190–2191.

9. Chang CC, Chow CC, Tellier LC, Vattikuti S, Purcell SM, Lee JJ. Second-generation PLINK: rising to the challenge of larger and richer datasets. *Gigascience*. 2015;4:s13742-015.

10. Purcell S, Chang C. PLINK v2.00a3LM [Internet]. Available from: www.cog-genomics.org/plink/2.0/

11. Lee S, Emond MJ, Bamshad MJ, Barnes KC, Rieder MJ, Nickerson DA, Team ELP, Christiani DC, Wurfel MM, Lin X. Optimal unified approach for rare-variant association testing with application to small-sample case-control whole-exome sequencing studies. *Am. J. Hum. Genet.* 2012;91:224–237.

12. Lee S, Teslovich TM, Boehnke M, Lin X. General framework for meta-analysis of rare variants in sequencing association studies. *Am. J. Hum. Genet.* 2013;93:42–53.

13. Grami N, Chong M, Lali R, Mohammadi-Shemirani P, Henshall DE, Rannikmäe K, Paré G. Global assessment of Mendelian stroke genetic prevalence in 101 635 individuals from 7 ethnic groups. *Stroke*. 2020;51:1290–1293.

14. Li X, Li Z, Zhou H, Gaynor SM, Liu Y, Chen H, Sun R, Dey R, Arnett DK, Aslibekyan S, et al. Dynamic incorporation of multiple in silico functional annotations empowers rare variant association analysis of large whole-genome sequencing studies at scale. *Nat. Genet.* 2020;52:969–983.

15. Rentzsch P, Witten D, Cooper GM, Shendure J, Kircher M. CADD: predicting the deleteriousness of variants throughout the human genome. *Nucleic Acids Res.* 2019;47:D886–D894.

16. Kircher M, Witten DM, Jain P, O'roak BJ, Cooper GM, Shendure J. A general framework for estimating the relative pathogenicity of human genetic variants. *Nat. Genet.* 2014;46:310–315.

17. Andersson R, Gebhard C, Miguel-Escalada I, Hoof I, Bornholdt J, Boyd M, Chen Y, Zhao X, Schmidl C, Suzuki T, et al. An atlas of active enhancers across human cell types and tissues. *Nature*. 2014;507:455–461.

18. The FANTOM Consortium and the RIKEN PMI and CLST (DGT). A promoter-level mammalian expression atlas. *Nature*. 2014;507:462–470.

19. Abugessaisa I, Noguchi S, Hasegawa A, Harshbarger J, Kondo A, Lizio M, Severin J, Carninci P, Kawaji H, Kasukawa T. FANTOM5 CAGE profiles of human and mouse reprocessed for GRCh38 and GRCm38 genome assemblies. *Sci. Data*. 2017;4:1–10.

20. Pruim RJ, Welch RP, Sanna S, Teslovich TM, Chines PS, Gliedt TP, Boehnke M, Abecasis GR, Willer CJ. LocusZoom: regional visualization of genome-wide association scan results. *Bioinformatics*. 2010;26:2336–2337.

21. Hahne F, Ivanek R. Visualizing genomic data using Gviz and bioconductor. In: Statistical genomics. Springer; 2016. p. 335–351.

22. Võsa U, Claringbould A, Westra H-J, Bonder MJ, Deelen P, Zeng B, Kirsten H, Saha A, Kreuzhuber R, Yazar S, et al. Large-scale cis- and trans-eQTL analyses identify thousands of genetic loci and polygenic scores that regulate blood gene expression. *Nat. Genet.* 2021;53:1300–1310.

23. Boyle AP, Hong EL, Hariharan M, Cheng Y, Schaub MA, Kasowski M, Karczewski KJ, Park J, Hitz BC, Weng S. Annotation of functional variation in personal genomes using RegulomeDB. *Genome Res*. 2012;22:1790–1797.

24. Wang Y, Song F, Zhang B, Zhang L, Xu J, Kuang D, Li D, Choudhary MNK, Li Y, Hu M, et al. The 3D Genome Browser: a web-based browser for visualizing 3D genome organization and long-range chromatin interactions. *Genome Biol*. 2018;19:151.

25. Adzhubei IA, Schmidt S, Peshkin L, Ramensky VE, Gerasimova A, Bork P, Kondrashov AS, Sunyaev SR. A method and server for predicting damaging missense mutations. *Nat. Methods*. 2010;7:248–249.

26. Kumar P, Henikoff S, Ng PC. Predicting the effects of coding non-synonymous variants on protein function using the SIFT algorithm. *Nat. Protoc*. 2009;4:1073–1081.

27. McLaren W, Gil L, Hunt SE, Riat HS, Ritchie GRS, Thormann A, Flicek P, Cunningham F. The Ensembl Variant Effect Predictor. *Genome Biol*. 2016;17:122.

28. Moore CM, Jacobson SA, Fingerlin TE. Power and Sample Size Calculations for Genetic Association Studies in the Presence of Genetic Model Misspecification. *Hum. Hered*. 2019;84:256–271.

29. Chen H, Huffman JE, Brody JA, Wang C, Lee S, Li Z, Gogarten SM, Sofer T, Bielak LF, Bis JC, et al. Efficient Variant Set Mixed Model Association Tests for Continuous and Binary Traits in Large-Scale Whole-Genome Sequencing Studies. *Am. J. Hum. Genet.* 2019;104:260–274.

30. Jurgens SJ, Choi SH, Morrill VN, Chaffin M, Pirruccello JP, Halford JL, Weng L-C, Nauffal V, Roselli C, Hall AW, et al. Analysis of rare genetic variation underlying cardiometabolic diseases and traits among 200,000 individuals in the UK Biobank. *Nat. Genet.* 2022;54:240–250.

31. Backman JD, Li AH, Marcketta A, Sun D, Mbatchou J, Kessler MD, Benner C, Liu D, Locke AE, Balasubramanian S, et al. Exome sequencing and analysis of 454,787 UK Biobank participants. *Nature*. 2021;599:628–634.

32. Andrews S. FastQC: a quality control tool for high throughput sequence data. 2010;Available from: http://www.bioinformatics.babraham.ac.uk/projects/fastqc

33. Goldstein JI, Crenshaw A, Carey J, Grant GB, Maguire J, Fromer M, O'Dushlaine C, Moran JL, Chambert K, Stevens C. zCall: a rare variant caller for array-based genotyping: genetics and population analysis. *Bioinformatics.* 2012;28:2543–2545.

34. Loh P-R, Danecek P, Palamara PF, Fuchsberger C, Reshef YA, Finucane HK, Schoenherr S, Forer L, McCarthy S, Abecasis GR. Reference-based phasing using the Haplotype Reference Consortium panel. *Nat. Genet.* 2016;48:1443–1448.

35. Browning BL, Browning SR. Genotype imputation with millions of reference samples. *Am. J. Hum. Genet.* 2016;98:116–126.
