## Supplementary Tables 10-23 for "Whole-genome sequencing identifies variants in *ANK1*, *LRRN1*, *HAS1,* and other genes and regulatory regions for stroke in type 1 diabetes"

| | |
|---|---|
| **Supplementary Table 10** | S10: Single variant stroke subtype associations |
| **Supplementary Table 11** | S11: Stroke subtype SKAT-O analysis variant number and CMAC |
| **Supplementary Table 12** | S12: Variants in discovered gene aggregate test (SKAT-O) findings with minimal adjustment |
| **Supplementary Table 13** | S13: Variants in discovered gene aggregate test (SKAT-O) findings with additional DKD adjustment |
| **Supplementary Table 14** | S14: SKAT-O replication in T1D with minimal adjustment |
| **Supplementary Table 15** | S15: SKAT-O replication in T1D with additional DKD adjustment |
| **Supplementary Table 16** | S16: SKAT-O replication in the general population (UK Biobank, Jurgens et al. 2022) |
| **Supplementary Table 17** | S17: SKAT-O replication in the general population (UK Biobank, Backman et al. 2022) |
| **Supplementary Table 18** | S18: Discovered genomic windows with replication in T1D (STAAR-O) |
| **Supplementary Table 19** | S19: Variants in the discovered genomic windows (STAAR-O) with replication in T1D and general population |
| **Supplementary Table 20** | S20: Discovered enhancers with replication in T1D (STAAR-O) |
| **Supplementary Table 21** | S21: Variants in the discovered enhancers (STAAR-O) with replication in T1D and general population |
| **Supplementary Table 22** | S22: Discovered promoters with replication in T1D (STAAR-O) |
| **Supplementary Table 23** | S23: Variants in the discovered promoters (STAAR-O) with replication in T1D and general population |

**Supplementary Table 10.** Single variant stroke subtype associations with replication

| | | | | | | | | | | | | | | Replication GWAS (T1D specific) | | Replication GWAS (General population) | |
|---|---|---|---|---|---|---|---|---|---|---|---|---|---|---|---|---|---|
| | | | | Discovery data (WGS: T1D specific) | | | | | | | | | | | | | |
| | **Variant** | **Position** | **Genetic model (†Adjusted for DKD)** | **Potential consequence** | **Data** | **REF/ALT** | **MAF** | **N (Hom.)** | **OR** | **Lower CI** | **Upper CI** | $I^2$ | ***p*-value** | **OR** | ***p*-value** | **OR** | **p-value** |
| **Ischemic stroke** | rs376936219 | 4:170784011 | Additive | *AADAT, MFAP3L, GALNTL6* | WGS | G/T | 4,6 % | - | 5,24 | 2,70 | 10,18 | - | 1,04E-06 | 0,87 | 0,46 | 1,03 | 0,36 |
| | rs4435704 | 4:170787127 | Additive | *AADAT, MFAP3L, GALNTL6* | WGS | T/C | 3,7 % | - | 7,02 | 3,38 | 14,60 | - | 1,76E-07 | - | - | - | - |
| | rs4401420 | 4:170787143 | Additive | *AADAT, MFAP3L, GALNTL6* | WGS | C/T | 4,1 % | - | 5,71 | 2,88 | 11,33 | - | 5,98E-07 | - | - | - | - |
| | rs1728177 | 17:43605161 | Recessive | *DHX8, MEOX1, ETV4* | WGS | G/A | 25,0 % | 36 | 7,49 | 3,50 | 16,04 | - | 2,19E-07 | 1,27 | 0,30 | 1,00 | 0,77 |
| | rs76073237 | 1:160870594 | Recessive† | *ITLN1* , *CD244* | WGS | GTA/G | 36,0 % | 54 | 5,20 | 2,50 | 10,83 | - | 1,08E-05 | - | - | - | - |
| | rs114001633 | 17:17819659 | Additive† | *SREBF1* (p.Pro227Leu) | ++ | G/A | 0,2 % | - | 129716 | 902 | 18656404 | 87 % | 3,41E-06 | 1,82 | 0,54 | 1,07 | 0,67 |
| | rs2224123 | 14:24313752 | Recessive | *LTB4R* | ++ | A/T | 13,8 % | 20 | 18,45 | 4,13 | 82,46 | 87 % | 1,36E-04 | 1,06 | 0,90 | 1,04 | 0,067 |
| **Hemorrhagic stroke** | rs376936219 | 4:170784011 | Additive | *AADAT, MFAP3L, GALNTL6* | WGS | G/T | 4,6 % | - | 2,69 | 0,99 | 7,34 | - | 5,30E-02 | 0,93 | 0,83 | 1,00 | 0,98 |
| | rs4435704 | 4:170787127 | Additive | *AADAT, MFAP3L, GALNTL6* | WGS | T/C | 3,7 % | - | 2,22 | 0,61 | 8,00 | - | 2,25E-01 | - | - | - | - |
| | rs4401420 | 4:170787143 | Additive | *AADAT, MFAP3L, GALNTL6* | WGS | C/T | 4,1 % | - | 2,31 | 0,80 | 6,64 | - | 1,22E-01 | - | - | - | - |
| | rs1728177 | 17:43605161 | Recessive | *DHX8, MEOX1, ETV4* | WGS | G/A | 24,3 % | 27 | 6,46 | 2,43 | 17,17 | - | 1,82E-04 | 0,50 | 0,09 | 0,98 | 0,41 |
| | rs76073237 | 1:160870594 | Recessive† | *ITLN1* , *CD244* | WGS | GTA/G | 35,0 % | 45 | 5,26 | 2,09 | 13,27 | - | 4,29E-04 | - | - | - | - |
| | rs114001633* | 17:17819659 | Additive† | *SREBF1* (p.Pro227Leu) | ++ | G/A | 0,2 % | - | 41632847 | 105163 | 16482021208 | 96 % | 8,96E-09 | 42,70 | 0,023 | 0,74 | 0,32 |
| | rs2224123 | 14:24313752 | Recessive | *LTB4R* | ++ | A/T | 13,5 % | 18 | 267,66 | 27,79 | 2577,76 | 0 % | 1,32E-06 | 0,52 | 0,41 | 1,04 | 0,31 |

All variants were in Hardy-Weinberg equilibrium (HWE p-value>0.05).
*Note that rs114001633 did not pass MAC threshold, but results are still presented (MAC=3).

**Supplementary Table 11.** Variant number (N) and cumulative minor allele count (CMAC) for stroke subtype SKAT-O analyses: **A.** Minimal model, **B.** Additionally DKD adjusted model

**A.**

| PAV 1% | Ischemic stroke | | Hemorrhagic stroke | |
|---|---|---|---|---|
| Gene | N | CMAC | N | CMAC |
| *LRRN1* | 5 | 12 | 7 | 17 |
| *FOXO1* | 2 | 5 | - | - |
| *CLEC4M* | 6 | 8 | 9 | 11 |
| *TARBP2* | 2 | 5 | - | - |
| *RRP9* | 6 | 10 | 4 | 5 |
| *GCDH* | - | - | - | - |
| *SLC18A2* | 3 | 12 | 3 | 8 |

| PAV 5% | Ischemic stroke | | Hemorrhagic stroke | |
|---|---|---|---|---|
| Gene | N | CMAC | N | CMAC |
| *ANK1* | 23 | 225 | 22 | 198 |
| *UTS2* | 2 | 26 | 2 | 18 |
| *APOBEC4* | 8 | 170 | 7 | 123 |

| PTV 1% | Ischemic stroke | | Hemorrhagic stroke | |
|---|---|---|---|---|
| Gene | N | CMAC | N | CMAC |
| *HAS1* | 4 | 7 | - | - |

| PTV 5% | Ischemic stroke | | Hemorrhagic stroke | |
|---|---|---|---|---|
| Gene | N | CMAC | N | CMAC |
| *ARPC5* | 2 | 31 | 2 | 26 |

**B.**

| PAV 1% | Ischemic stroke | | Hemorrhagic stroke | |
|---|---|---|---|---|
| Gene | N | CMAC | N | CMAC |
| *LRRN1* | 5 | 12 | 7 | 17 |
| *CLEC4M* | 6 | 8 | 9 | 11 |
| *GCDH* | - | - | - | - |
| *SLC18A2* | 3 | 12 | 3 | 8 |
| *MAP3K12* | 6 | 17 | 5 | 14 |
| *TARBP2* | 2 | 5 | - | - |
| *MTRNR2L7* | 2 | 8 | 2 | 6 |

| PAV 5% | Ischemic stroke | | Hemorrhagic stroke | |
|---|---|---|---|---|
| Gene | N | CMAC | N | CMAC |
| *ANK1* | 23 | 225 | 22 | 198 |
| *APOBEC4* | 7 | 139 | 7 | 123 |
| *UTS2* | 2 | 24 | 2 | 18 |

| PTV 1% | Ischemic stroke | | Hemorrhagic stroke | |
|---|---|---|---|---|
| Gene | N | CMAC | N | CMAC |
| *HAS1* | 4 | 7 | - | - |
| *UACA* | - | - | - | - |
| *CHEK2* | 2 | 13 | 2 | 10 |

| PTV 5% | Ischemic stroke | | Hemorrhagic stroke | |
|---|---|---|---|---|
| Gene | N | CMAC | N | CMAC |
| *ARPC5* | 2 | 31 | 2 | 26 |

Note that variant number may differ between the stroke phenotypes (stroke, ischemic stroke and hemmorhagic stroke) due to MAF thresholds and differences in variant carriers

**Supplementary Table 12.** Variants in discovered stroke-risk SKAT-O genes with minimal adjustment; and replication in T1D (FinnDiane) and general populaiton (FinnGen); using score test

*Note: Stroke subtype analyses may have included additional variants (i.e. passed MAF threshold in smaller data)*

| | | | | | | | | | | | | FinnDiane discovery cohort WES+WGS; analysed with score test and, when possible, fixed-effects meta-analysis (T1D specific) | | | | | | | | | | | | | | FinnDiane GWAS/genotyping replication using score test (T1D specific) | | | | | | | | | | | | | | | | | | FinnGen GWAS replication (general population) | | | | | | | | | | | | | | | | | | | |
|---|---|---|---|---|---|---|---|---|---|---|---|---|---|---|---|---|---|---|---|---|---|---|---|---|---|---|---|---|---|---|---|---|---|---|---|---|---|---|---|---|---|---|---|---|---|---|---|---|---|---|---|---|---|---|---|---|---|---|---|---|---|---|---|
| | | | | | | | metaSKAT (stroke) | | | | | Stroke | | | | Ischemic stroke | | | | | Hemorrhagic stroke | | | | | Stroke | | | | | | Ischemic stroke | | | | | | Hemorrhagic stroke | | | | | | Stroke | | | | | Stroke, without SAH | | | | | Ischemic stroke | | | | | Hemorrhagic stroke | | | | |
| MODEL | SetID | Position | SNPID | Major | Minor | Data | Score | MAF | MissingRate | MAC | N | Beta | SE | *p*-value | Het $I^2$ | MAF | Beta | SE | *p*-value | Het $I^2$ | MAF | Beta | SE | *p*-value | Het $I^2$ | HWE | MAC | MAF | Beta | SE | *p*-value | HWE | MAC | MAF | Beta | SE | *p*-value | HWE | MAC | MAF | Beta | SE | *p*-value | MAC | MAF | Beta | SE | *p*-value | MAC | MAF | Beta | SE | *p*-value | MAC | MAF | Beta | SE | *p*-value | MAC | MAF | Beta | SE | *p*-value |
| PTV 5% | *ARPC5* | 1:183623459 | rs16861256 | A | G | WGS | 9,090 | 3 % | 0,00 % | 35 | 571 | 1,81 | 0,45 | 5,02E-05 | 0 | 3 % | 1,74 | 0,61 | 4,37E-03 | 0 | 2 % | 1,69 | 0,97 | 8,10E-02 | 0 | 0,48 | 187 | 2,3 % | -0,046 | 0,21 | 8,28E-01 | 0,47 | 178 | 2,3 % | -0,083 | 0,27 | 7,57E-01 | 0,47 | 176 | 2,4 % | 0,799 | 0,47 | 8,85E-02 | 13867 | 2,74 % | -0,055 | 0,04 | 1,47E-01 | 13863 | 2,74 % | -0,055 | 0,04 | 1,61E-01 | 13855 | 2,74 % | -0,063 | 0,04 | 1,23E-01 | 13413 | 2,74 % | -0,078 | 0,08 | 3,04E-01 |
| | | 1:183630530 | . | CTTCATTA | C | WGS | -0,942 | 0 % | 0,00 % | 4 | 571 | -1,40 | 1,22 | 2,52E-01 | 0 | 0 % | -1,29 | 1,40 | 3,58E-01 | 0 | 0 % | -1,13 | 1,97 | 5,65E-01 | 0 | | | | | | | | | | | | | | | | | | | | | | | | | | | | | | | | | | | | | | |
| PAV 5% | *APOBEC4* | 1:183647731 | rs139524898 | A | G | WES | -0,140 | 0 % | 0,42 % | ≤3 | 480 | -1,19 | 1,99 | 5,50E-01 | 0 | 0 % | -1,14 | 2,27 | 6,17E-01 | 0 | 0 % | -1,05 | 3,76 | 7,80E-01 | 0 | | | | | | | | | | | | | | | | | | | | | | | | | | | | | | | | | | | | | | |
| | | | | | G | WGS | -0,161 | 0 % | 0,00 % | ≤3 | 571 | | | | | | | | | | | | | | | | | | | | | | | | | | | | | | | | | | | | | | | | | | | | | | | | | | | | |
| | | 1:183647749 | rs10911390 | C | T | WGS | 9,090 | 3 % | 0,00 % | 35 | 571 | 1,43 | 0,37 | 1,18E-04 | 57,3 | 2 % | 1,23 | 0,50 | 1,27E-02 | 50,2 | 2 % | 1,59 | 0,74 | 3,09E-02 | 0 | 0,50 | 191 | 2,4 % | -0,062 | 0,21 | 7,64E-01 | 0,48 | 182 | 2,4 % | -0,097 | 0,27 | 7,17E-01 | 0,49 | 180 | 2,5 % | 0,766 | 0,46 | 9,83E-02 | 13921 | 2,75 % | -0,054 | 0,04 | 1,59E-01 | 13917 | 2,75 % | -0,056 | 0,04 | 1,54E-01 | 13909 | 2,75 % | -0,064 | 0,04 | 1,19E-01 | 13464 | 2,75 % | -0,074 | 0,08 | 3,30E-01 |
| | | | | | | WES | 1,267 | 2 % | 0,42 % | 19 | 480 | | | | | | | | | | | | | | | | | | | | | | | | | | | | | | | | | | | | | | | | | | | | | | | | | | |
| | | 1:183647959 | rs10911391 | G | A | WES | 1,267 | 2 % | 0,42 % | 19 | 480 | 1,43 | 0,37 | 1,18E-04 | 57,3 | 2 % | 1,23 | 0,50 | 1,27E-02 | 50,2 | 2 % | 1,59 | 0,74 | 3,09E-02 | 0 | 0,50 | 191 | 2,4 % | -0,062 | 0,21 | 7,64E-01 | 0,48 | 182 | 2,4 % | -0,097 | 0,27 | 7,17E-01 | 0,49 | 180 | 2,5 % | 0,766 | 0,46 | 9,83E-02 | 13928 | 2,75 % | -0,054 | 0,04 | 1,59E-01 | 13924 | 2,75 % | -0,056 | 0,04 | 1,54E-01 | 13916 | 2,75 % | -0,064 | 0,04 | 1,20E-01 | 13471 | 2,75 % | -0,075 | 0,08 | 3,22E-01 |
| | | | | | | WGS | 9,090 | 3 % | 0,00 % | 35 | 571 | | | | | | | | | | | | | | | | | | | | | | | | | | | | | | | | | | | | | | | | | | | | | | | | | | |
| | | 1:183648558 | rs16861394 | G | A | WGS | 9,090 | 3 % | 0,00 % | 35 | 571 | 1,43 | 0,37 | 1,18E-04 | 57,3 | 2 % | 1,23 | 0,50 | 1,27E-02 | 50,2 | 2 % | 1,59 | 0,74 | 3,09E-02 | 0 | 0,50 | 191 | 2,4 % | -0,062 | 0,21 | 7,64E-01 | 0,48 | 182 | 2,4 % | -0,097 | 0,27 | 7,17E-01 | 0,49 | 180 | 2,5 % | 0,766 | 0,46 | 9,83E-02 | 13929 | 2,75 % | -0,053 | 0,04 | 1,60E-01 | 13925 | 2,75 % | -0,056 | 0,04 | 1,55E-01 | 13917 | 2,75 % | -0,064 | 0,04 | 1,22E-01 | 13472 | 2,75 % | -0,075 | 0,08 | 3,21E-01 |
| | | | | | | WES | 1,267 | 2 % | 0,42 % | 19 | 480 | | | | | | | | | | | | | | | | | | | | | | | | | | | | | | | | | | | | | | | | | | | | | | | | | | |
| | | 1:183648588 | rs776661029 | T | G | WES | -0,190 | 0 % | 0,42 % | ≤3 | 480 | 1,26 | 1,58 | 4,26E-01 | 34,8 | 0 % | 2,48 | 1,91 | 1,93E-01 | 65,5 | 0 % | -1,08 | 3,44 | 7,54E-01 | 0 | 1 | 10 | 0,1 % | 0,877 | 0,99 | 3,77E-01 | 1 | 8 | 0,1 % | -0,801 | 1,34 | 5,50E-01 | 1 | 9 | 0,1 % | 1,302 | 2,30 | 5,72E-01 | 576 | 0,11 % | 0,222 | 0,20 | 2,60E-01 | 577 | 0,11 % | 0,210 | 0,20 | 3,05E-01 | 577 | 0,11 % | 0,329 | 0,21 | 1,24E-01 | 555 | 0,11 % | 0,046 | 0,39 | 9,05E-01 |
| | | | | | | WGS | 0,694 | 0 % | 0,00 % | ≤3 | 571 | | | | | | | | | | | | | | | | | | | | | | | | | | | | | | | | | | | | | | | | | | | | | | | | | | |
| | | 1:183648589 | . | GT | G | WES | 0,842 | 0 % | 0,42 % | ≤3 | 480 | 6,40 | 2,76 | 2,02E-02 | 0 | | | | | | 0 % | 25,24 | 5,13 | 8,56E-07 | 0 | | | | | | | | | | | | | | | | | | | | | | | | | | | | | | | | | | | | | | |
| | | 1:183648660 | rs182486585 | G | C | WGS | -0,116 | 0 % | 0,00 % | ≤3 | 571 | -1,14 | 3,14 | 7,16E-01 | 0 | 0 % | -1,09 | 3,79 | 7,73E-01 | 0 | 0 % | -1,03 | 6,72 | 8,78E-01 | 0 | | | | | | | | | | | | | | | | | | | | | | | | | | | | | | | | | | | | | | |
| | | 1:183653304 | . | T | C | WES | 0,870 | 0 % | 0,42 % | ≤3 | 480 | 7,79 | 2,99 | 9,21E-03 | 0 | 0 % | 12,49 | 3,69 | 7,04E-04 | 0 | | | | | | | | | | | | | | | | | | | | | | | | | | | | | | | | | | | | | | | | | | | |
| PAV 5% | *UTS2* | 1:7849677 | rs2890565 | C | T | WES | -0,848 | 1 % | 0,00 % | 13 | 480 | 1,66 | 0,57 | 3,53E-03 | 93,1 | 1 % | 2,19 | 0,69 | 1,59E-03 | 92,9 | 1 % | 0,44 | 1,23 | 7,22E-01 | 82,6 | 0,0019 | 120 | 1,5 % | 0,155 | 0,25 | 5,38E-01 | 0,0013 | 112 | 1,5 % | 0,153 | 0,33 | 6,40E-01 | 0,00082 | 105 | 1,4 % | -0,732 | 0,58 | 2,09E-01 | 7082 | 1,40 % | 0,053 | 0,05 | 3,31E-01 | 7081 | 1,40 % | 0,063 | 0,06 | 2,61E-01 | 7078 | 1,40 % | 0,066 | 0,06 | 2,60E-01 | 6850 | 1,40 % | -0,091 | 0,11 | 4,06E-01 |
| | | | | | | WGS | 5,986 | 1 % | 0,00 % | 12 | 571 | | | | | | | | | | | | | | | | | | | | | | | | | | | | | | | | | | | | | | | | | | | | | | | | | | |
| | | 1:7850908 | rs201977733 | C | T | WGS | -0,120 | 0 % | 0,00 % | ≤3 | 571 | -1,14 | 3,08 | 7,12E-01 | 0 | 0 % | -1,08 | 3,93 | 7,83E-01 | 0 | 0 % | -1,04 | 5,89 | 8,60E-01 | 0 | | | | | | | | | | | | | | | | | | | | | | | | | | | | | | | | | | | | | | |
| PAV 1% & PAV 5% | *SLC18A2* | 10:117253425 | rs184073241 | C | T | WGS | -0,096 | 0 % | 0,00 % | ≤3 | 571 | -1,11 | 3,41 | 7,44E-01 | 0 | 0 % | -1,06 | 4,50 | 8,14E-01 | 0 | 0 % | -1,03 | 6,44 | 8,73E-01 | 0 | | | | | | | | | | | | | | | | | | | | | | | | | | | | | | | | | | | | | | |
| | | 10:117257799 | rs199750100 | T | A | WGS | 0,245 | 0 % | 0,00 % | 5 | 571 | 3,02 | 0,84 | 3,32E-04 | 86,3 | 1 % | 4,15 | 1,03 | 5,78E-05 | 84,4 | 0 % | 4,12 | 2,28 | 7,00E-02 | 83,3 | 1 | 34 | 0,5 % | -0,538 | 0,51 | 2,94E-01 | 1 | 34 | 0,5 % | -0,255 | 0,64 | 6,92E-01 | 1 | 33 | 0,5 % | -0,484 | 1,13 | 6,68E-01 | 2062 | 0,41 % | 0,067 | 0,10 | 5,06E-01 | 2061 | 0,41 % | 0,114 | 0,10 | 2,74E-01 | 2059 | 0,41 % | 0,117 | 0,11 | 2,85E-01 | 1991 | 0,41 % | -0,074 | 0,20 | 7,11E-01 |
| | | | | | | WES | 4,017 | 1 % | 0,00 % | 6 | 480 | | | | | | | | | | | | | | | | | | | | | | | | | | | | | | | | | | | | | | | | | | | | | | | | | | |
| | | 10:117267034 | rs367902584 | G | A | WGS | -0,133 | 0 % | 0,00 % | ≤3 | 571 | -1,16 | 2,96 | 6,94E-01 | 0 | 0 % | -1,13 | 3,34 | 7,36E-01 | 0 | 0 % | -1,04 | 6,12 | 8,65E-01 | 0 | | | | | | | | | | | | | | | | | | | | | | | | | | | | | | | | | | | | | | |
| PAV 1% & PAV 5% | *TARBP2* | 12:53503093 | rs146234229 | G | C | WGS | -0,203 | 0 % | 0,00 % | ≤3 | 571 | -1,25 | 1,79 | 4,84E-01 | 0 | 0 % | -1,15 | 2,17 | 5,94E-01 | 0 | 0 % | -1,07 | 3,04 | 7,25E-01 | 0 | 1 | 8 | 0,1 % | 3,316 | 1,09 | 2,20E-03 | 1 | 6 | 0,1 % | 2,804 | 1,59 | 7,83E-02 | 1 | 6 | 0,1 % | 10,403 | 2,80 | 2,05E-04 | 462 | 0,09 % | 0,094 | 0,23 | 6,81E-01 | 462 | 0,09 % | 0,159 | 0,24 | 5,01E-01 | 461 | 0,09 % | 0,202 | 0,25 | 4,18E-01 | 439 | 0,09 % | 0,086 | 0,45 | 8,49E-01 |
| | | | | | | WES | -0,187 | 0 % | 0,00 % | ≤3 | 480 | | | | | | | | | | | | | | | | | | | | | | | | | | | | | | | | | | | | | | | | | | | | | | | | | |
| | | 12:53504426 | rs149488319 | T | C | WGS | 0,759 | 0 % | 0,00 % | ≤3 | 571 | 6,19 | 1,36 | 5,55E-06 | 11,9 | 0 % | 10,96 | 2,01 | 4,77E-08 | 0 | | | | | | 1 | 13 | 0,2 % | -0,251 | 0,79 | 7,50E-01 | 1 | 12 | 0,2 % | -0,963 | 1,03 | 3,50E-01 | 1 | 13 | 0,2 % | 3,808 | 1,71 | 2,63E-02 | 792 | 0,16 % | 0,190 | 0,17 | 2,56E-01 | 791 | 0,16 % | 0,134 | 0,17 | 4,36E-01 | 792 | 0,16 % | 0,233 | 0,18 | 1,95E-01 | 754 | 0,15 % | 0,094 | 0,34 | 7,83E-01 |
| | | | | | | WES | 2,576 | 0 % | 0,00 % | ≤3 | 480 | | | | | | | | | | | | | | | | | | | | | | | | | | | | | | | | | | | | | | | | | | | | | | | | | |
| PAV 1% & PAV 5% | *FOXO1* | 13:40560079 | rs767400829 | G | A | WES | -0,192 | 0 % | 0,42 % | ≤3 | 480 | 1,75 | 1,74 | 3,16E-01 | 61 | 0 % | 3,15 | 2,08 | 1,30E-01 | 79,9 | 0 % | -1,06 | 4,96 | 8,30E-01 | 0 | | | | | | | | | | | | | | | | | | | | | | | | | | | | | | | | | | | | | | |
| | | | | | | WGS | 0,767 | 0 % | 0,00 % | ≤3 | 571 | | | | | | | | | | | | | | | | | | | | | | | | | | | | | | | | | | | | | | | | | | | | | | | | | | |
| | | 13:40666178 | rs773597669 | G | T | WGS | 4,085 | 0 % | 0,00 % | 5 | 571 | 4,39 | 1,04 | 2,29E-05 | 0 | 0 % | 6,17 | 1,52 | 4,78E-05 | 0 | 0 % | 13,35 | 3,80 | 4,44E-04 | 0 | 1 | 39 | 0,5 % | 0,070 | 0,46 | 8,80E-01 | 1 | 36 | 0,5 % | -0,300 | 0,61 | 6,20E-01 | 1 | 37 | 0,5 % | 2,596 | 1,03 | 1,20E-02 | 2821 | 0,56 % | 0,024 | 0,09 | 7,79E-01 | 2820 | 0,56 % | 0,027 | 0,09 | 7,68E-01 | 2819 | 0,56 % | 0,050 | 0,09 | 5,97E-01 | 2716 | 0,56 % | 0,015 | 0,17 | 9,32E-01 |
| PAV 1% & PAV 5% | *GCDH* | 19:12891359 | . | G | T | WGS | 0,791 | 0 % | 0,00 % | ≤3 | 571 | 4,83 | 2,47 | 5,06E-02 | 0 | | | | | | 0 % | 19,54 | 4,54 | 1,67E-05 | 0 | | | | | | | | | | | | | | | | | | | | | | | | | | | | | | | | | | | | | | |
| | | 19:12896058 | rs149120354 | T | C | WES | 0,843 | 0 % | 0,42 % | ≤3 | 480 | 6,44 | 2,76 | 1,98E-02 | 0 | 0 % | 8,87 | 3,16 | 5,06E-03 | 0 | | | | | | 1 | ≤3 | 0,0 % | 2,206 | 1,70 | 1,94E-01 | 1 | ≤3 | 0,0 % | -0,833 | 2,52 | 7,41E-01 | 1 | ≤3 | 0,0 % | -0,819 | 4,35 | 8,51E-01 | 76 | 0,02 % | -0,751 | 0,54 | 1,64E-01 | 76 | 0,02 % | -0,702 | 0,56 | 2,09E-01 | 76 | 0,02 % | -0,624 | 0,59 | 2,86E-01 | 78 | 0,02 % | -0,080 | 1,03 | 9,38E-01 |
| | | 19:12896341 | rs201076343 | G | T | WES | -0,139 | 0 % | 0,42 % | ≤3 | 480 | -1,18 | 2,92 | 6,86E-01 | 0 | 0 % | -1,11 | 3,68 | 7,63E-01 | 0 | 0 % | -1,10 | 3,92 | 7,79E-01 | 0 | | | | | | | | | | | | | | | | | | | | | | | | | | | | | | | | | | | | | | |
| | | 19:12896401 | rs751742575 | C | T | WES | 0,905 | 0 % | 0,42 % | ≤3 | 480 | 10,68 | 3,44 | 1,87E-03 | 0 | 0 % | 18,50 | 4,42 | 2,89E-05 | 0 | | | | | | | | | | | | | | | | | | | | | | | | | | | | | | | | | | | | | | | | | | | |
| | | 19:12897009 | rs761135089 | G | A | WGS | 0,641 | 0 % | 0,00 % | ≤3 | 571 | 2,83 | 2,10 | 1,78E-01 | 0 | | | | | | | | | | | | | | | | | | | | | | | | | | | | | | | | | | | | | | | | | | | | | | | | |
| | | 19:12897824 | rs121434369 | C | T | WES | 0,890 | 0 % | 0,42 % | ≤3 | 480 | 9,21 | 3,22 | 4,20E-03 | 0 | 0 % | 14,22 | 3,91 | 2,79E-04 | 0 | | | | | | 1 | 6 | 0,1 % | 0,863 | 1,16 | 4,57E-01 | 1 | 6 | 0,1 % | 2,281 | 1,45 | 1,17E-01 | 1 | 5 | 0,1 % | -0,574 | 2,75 | 8,35E-01 | 289 | 0,06 % | -0,020 | 0,26 | 9,41E-01 | 289 | 0,06 % | 0,006 | 0,27 | 9,83E-01 | 289 | 0,06 % | 0,084 | 0,29 | 7,68E-01 | 273 | 0,06 % | -0,609 | 0,54 | 2,59E-01 |
| PTV 1% & PTV 5% | *HAS1* | 19:51713664 | . | G | T | WES | -0,267 | 0 % | 0,42 % | ≤3 | 480 | -1,18 | 2,10 | 5,74E-01 | 0 | 0 % | -1,13 | 2,43 | 6,41E-01 | 0 | 0 % | -1,08 | 3,37 | 7,48E-01 | 0 | | | | | | | | | | | | | | | | | | | | | | | | | | | | | | | | | | | | | | |
| | | 19:51713853 | . | G | T | WES | -0,113 | 0 % | 0,63 % | ≤3 | 480 | -1,14 | 3,18 | 7,19E-01 | 0 | 0 % | -1,10 | 3,78 | 7,71E-01 | 0 | 0 % | -1,05 | 5,47 | 8,47E-01 | 0 | | | | | | | | | | | | | | | | | | | | | | | | | | | | | | | | | | | | | | |
| | | 19:51716967 | . | C | T | WES | 0,851 | 0 % | 0,42 % | ≤3 | 480 | 6,80 | 2,83 | 1,61E-02 | 0 | 0 % | 11,07 | 3,49 | 1,52E-03 | 0 | | | | | | | | | | | | | | | | | | | | | | | | | | | | | | | | | | | | | | | | | | | |
| | | 19:51723924 | rs58597876 | C | CACAC | WGS | 2,476 | 0 % | 0,00 % | ≤3 | 571 | 5,91 | 1,55 | 1,30E-04 | 0 | 0 % | 8,89 | 1,83 | 1,19E-06 | 0 | | | | | | | | | | | | | | | | | | | | | | | | | | | | | | | | | | | | | | | | | | | |
| PAV 1% & PAV 5% | *CLEC4M* | 19:7765676 | rs200077446 | G | A | WGS | -0,126 | 0 % | 0,00 % | ≤3 | 571 | -1,15 | 3,03 | 7,03E-01 | 0 | 0 % | -1,11 | 3,48 | 7,50E-01 | 0 | 0 % | -1,03 | 6,30 | 8,70E-01 | 0 | 1 | 9 | 0,1 % | 0,145 | 0,95 | 8,78E-01 | 1 | 9 | 0,1 % | 1,159 | 1,19 | 3,30E-01 | 1 | 8 | 0,1 % | -0,668 | 2,18 | 7,59E-01 | 572 | 0,11 % | -0,071 | 0,18 | 7,00E-01 | 572 | 0,11 % | -0,213 | 0,19 | 2,58E-01 | 571 | 0,11 % | -0,254 | 0,20 | 1,97E-01 | 551 | 0,11 % | 0,577 | 0,37 | 1,18E-01 |
| | | 19:7765776 | rs113080783 | A | G | WES | -0,299 | 0 % | 1,04 % | ≤3 | 480 | -1,19 | 2,00 | 5,51E-01 | 0 | 0 % | -1,12 | 2,32 | 6,27E-01 | 0 | 0 % | -1,07 | 3,53 | 7,61E-01 | 0 | | | | | | | | | | | | | | | | | | | | | | | | | | | | | | | | | | | | | | |
| | | 19:7766121 | rs144783051 | A | G | WES | 0,868 | 0 % | 0,42 % | ≤3 | 480 | 7,65 | 2,97 | 9,96E-03 | 0 | | | | | | 0 % | 27,02 | 5,30 | 3,41E-07 | 0 | 1 | 10 | 0,2 % | -0,136 | 0,87 | 8,75E-01 | 1 | 10 | 0,2 % | 0,566 | 1,04 | 5,88E-01 | 1 | 9 | 0,2 % | -0,806 | 1,75 | 6,45E-01 | *Genotyped | | | | | | | | | | | | | | | | | | | |
| | | 19:7766155 | rs746560445 | G | GT | WGS | 0,773 | 0 % | 0,00 % | ≤3 | 571 | 4,43 | 2,39 | 6,44E-02 | 0 | 0 % | 6,60 | 2,79 | 1,80E-02 | 0 | | | | | | | | | | | | | | | | | | | | | | | | | | | | | | | | | | | | | | | | | | | |
| | | 19:7766659 | rs140767813 | G | A | WES | 0,898 | 0 % | 0,42 % | ≤3 | 480 | 10,00 | 3,34 | 2,73E-03 | 0 | | | | | | 0 % | 17,84 | 4,35 | 4,13E-05 | 0 | 0,00046 | ≤3 | 0,0 % | -0,321 | 1,27 | 8,00E-01 | 0,00049 | ≤3 | 0,0 % | -0,309 | 1,51 | 8,38E-01 | 0,00050 | ≤3 | 0,0 % | -0,277 | 2,56 | 9,14E-01 | *Genotyped | | | | | | | | | | | | | | | | | | | |
| | | 19:7766667 | rs377334555 | C | T | WES | 0,863 | 0 % | 0,42 % | ≤3 | 480 | 7,46 | 2,94 | 1,11E-02 | 0 | | | | | | 0 % | 19,79 | 4,57 | 1,48E-05 | 0 | | | | | | | | | | | | | | | | | | | | | | | | | | | | | | | | | | | | | | |
| | | 19:7767549 | rs752738017 | C | T | WES | 1,782 | 0 % | 0,42 % | ≤3 | 480 | 9,34 | 2,29 | 4,49E-05 | 0 | 0 % | 18,50 | 4,42 | 2,89E-05 | 0 | 0 % | 27,83 | 5,37 | 2,23E-07 | 0 | 1 | ≤3 | 0,0 % | -0,396 | 2,84 | 8,89E-01 | 1 | ≤3 | 0,0 % | -0,271 | 3,38 | 9,36E-01 | 1 | ≤3 | 0,0 % | -0,187 | 5,73 | 9,74E-01 | *Genotyped | | | | | | | | | | | | | | | | | | | |
| | | 19:7768929 | rs184828145 | T | C | WGS | 0,711 | 0 % | 0,00 % | ≤3 | 571 | 3,54 | 2,23 | 1,13E-01 | 0 | | | | | | 0 % | 12,18 | 3,65 | 8,41E-04 | 0 | | | | | | | | | | | | | | | | | | | | | | | | | | | | | | | | | | | | | | |
| | | 19:7768947 | rs201740080 | T | C | WES | -0,126 | 0 % | 0,42 % | ≤3 | 480 | -1,24 | 1,90 | 5,13E-01 | 0 | 0 % | -1,16 | 2,19 | 5,96E-01 | 0 | 0 % | -1,06 | 3,50 | 7,62E-01 | 0 | | | | | | | | | | | | | | | | | | | | | | | | | | | | | | | | | | | | | | |
| | | | | | | WGS | -0,219 | 0 % | 0,00 % | ≤3 | 571 | | | | | | | | | | | | | | | | | | | | | | | | | | | | | | | | | | | | | | | | | | | | | | | | | | |
| | | 19:7768973 | rs753343180 | C | G | WGS | -0,177 | 0 % | 0,00 % | ≤3 | 571 | -1,24 | 2,65 | 6,40E-01 | 0 | 0 % | -1,11 | 3,73 | 7,67E-01 | 0 | 0 % | -1,07 | 4,74 | 8,21E-01 | 0 | 1 | 6 | 0,1 % | 0,345 | 1,16 | 7,66E-01 | 1 | 6 | 0,1 % | 1,772 | 1,46 | 2,23E-01 | 1 | 5 | 0,1 % | -0,999 | 2,75 | 7,17E-01 | 642 | 0,13 % | -0,046 | 0,18 | 8,00E-01 | 642 | 0,13 % | -0,094 | 0,19 | 6,14E-01 | 642 | 0,13 % | -0,010 | 0,19 | 9,59E-01 | 620 | 0,13 % | -0,307 | 0,35 | 3,83E-01 |
| PAV 1% | *LRRN1* | 3:3844558 | rs755350940 | C | T | WGS | -0,209 | 0 % | 0,00 % | ≤3 | 571 | 2,59 | 1,93 | 1,80E-01 | 87,7 | 0 % | 4,66 | 2,40 | 5,17E-02 | 92 | 0 % | -1,03 | 4,77 | 8,28E-01 | 0 | 1 | 21 | 0,3 % | -0,016 | 0,62 | 9,80E-01 | 1 | 21 | 0,3 % | 0,456 | 0,78 | 5,59E-01 | 1 | 20 | 0,3 % | -0,313 | 1,38 | 8,21E-01 | 1483 | 0,29 % | -0,022 | 0,12 | 8,50E-01 | 1483 | 0,29 % | -0,033 | 0,12 | 7,90E-01 | 1482 | 0,29 % | -0,053 | 0,13 | 6,80E-01 | 1437 | 0,29 % | 0,078 | 0,23 | 7,38E-01 |
| | | | | | | WES | 0,906 | 0 % | 0,00 % | ≤3 | 480 | | | | | | | | | | | | | | | | | | | | | | | | | | | | | | | | | | | | | | | | | | | | | | | | | | |
| | | 3:3845194 | rs142381203 | C | T | WGS | 0,796 | 0 % | 0,00 % | ≤3 | 571 | 4,96 | 2,50 | 4,68E-02 | 0 | | | | | | 0 % | 16,41 | 4,18 | 8,69E-05 | 0 | - | ≤3 | 0,0 % | -0,176 | 0,28 | 5,30E-01 | - | ≤3 | 0,0 % | -0,103 | 0,21 | 6,24E-01 | - | ≤3 | 0,0 % | -0,027 | 0,11 | 8,01E-01 | **Genotyped: Linear regression without relatedness adjustment | | | | | | | | | | | | | | | | | | | |
| | | 3:3845210 | rs147007969 | C | T | WES | 2,311 | 0 % | 0,00 % | 4 | 480 | 3,43 | 1,08 | 1,53E-03 | 0 | 0 % | 4,67 | 1,39 | 8,25E-04 | 0 | 0 % | 4,39 | 2,34 | 6,06E-02 | 78 | 1 | 26 | 0,3 % | -0,329 | 0,56 | 5,56E-01 | 1 | 26 | 0,3 % | 0,097 | 0,70 | 8,90E-01 | 1 | 25 | 0,3 % | -0,441 | 1,24 | 7,21E-01 | 1431 | 0,28 % | -0,066 | 0,12 | 5,67E-01 | 1431 | 0,28 % | -0,049 | 0,12 | 6,82E-01 | 1431 | 0,28 % | 0,006 | 0,12 | 9,64E-01 | 1380 | 0,28 % | -0,363 | 0,23 | 1,11E-01 |
| | | | | | | WGS | 0,614 | 0 % | 0,00 % | ≤3 | 571 | | | | | | | | | | | | | | | | | | | | | | | | | | | | | | | | | | | | | | | | | | | | | | | | | | |
| | | 3:3845554 | rs141825989 | C | T | WES | 0,825 | 0 % | 0,00 % | ≤3 | 480 | 5,88 | 2,67 | 2,76E-02 | 0 | 0 % | 7,33 | 2,92 | 1,21E-02 | 0 | | | | | | 1 | ≤3 | 0,0 % | -1,123 | 2,83 | 6,92E-01 | 1 | ≤3 | 0,0 % | -0,939 | 3,37 | 7,81E-01 | 1 | ≤3 | 0,0 % | -1,072 | 5,71 | 8,51E-01 | *Genotyped | | | | | | | | | | | | | | | | | | | |
| | | 3:3845810 | rs770349026 | G | A | WES | 0,751 | 0 % | 0,00 % | ≤3 | 480 | 3,50 | 2,16 | 1,05E-01 | 0 | 0 % | 5,57 | 2,59 | 3,12E-02 | 0 | 0 % | -1,08 | 4,16 | 7,94E-01 | 0 | | | | | | | | | | | | | | | | | | | | | | | | | | | | | | | | | | | | | | |
| | | 3:3845966 | rs747455683 | C | T | WGS | 0,739 | 0 % | 0,00 % | ≤3 | 571 | 3,90 | 2,30 | 8,96E-02 | 0 | | | | | | 0 % | 11,44 | 3,55 | 1,25E-03 | 0 | | | | | | | | | | | | | | | | | | | 281 | 0,06 % | -0,289 | 0,31 | 3,51E-01 | 282 | 0,06 % | -0,450 | 0,32 | 1,59E-01 | 282 | 0,06 % | -0,336 | 0,33 | 3,14E-01 | 274 | 0,06 % | 0,003 | 0,61 | 9,96E-01 |
| | | 3:3846082 | rs543878366 | G | A | WES | -0,158 | 0 % | 0,00 % | ≤3 | 480 | -1,20 | 2,75 | 6,63E-01 | 0 | 0 % | -1,13 | 3,22 | 7,26E-01 | 0 | 0 % | -1,06 | 4,88 | 8,29E-01 | 0 | | | | | | | | | | | | | | | | | | | | | | | | | | | | | | | | | | | | | | |
| PAV 1% | *RRP9* | 3:51934709 | rs142340392 | G | A | WGS | 1,287 | 0 % | 0,00 % | 4 | 571 | 2,25 | 1,32 | 8,85E-02 | 0 | 0 % | 3,55 | 1,54 | 2,09E-02 | 0 | 0 % | -1,05 | 3,87 | 7,86E-01 | 0 | 1 | 15 | 0,2 % | -0,219 | 0,73 | 7,66E-01 | 1 | 14 | 0,2 % | -0,736 | 0,95 | 4,40E-01 | 1 | 14 | 0,2 % | -0,693 | 1,65 | 6,74E-01 | 891 | 0,18 % | -0,308 | 0,15 | 3,45E-02 | 890 | 0,18 % | -0,348 | 0,15 | 2,07E-02 | 890 | 0,18 % | -0,298 | 0,16 | 5,76E-02 | 863 | 0,18 % | -0,398 | 0,29 | 1,75E-01 |
| | | 3:51934759 | rs200319255 | C | A | WGS | 0,906 | 0 % | 0,00 % | ≤3 | 571 | 10,75 | 3,44 | 1,80E-03 | 0 | 0 % | 18,32 | 4,40 | 3,17E-05 | 0 | | | | | | | | | | | | | | | | | | | | | | | | | | | | | | | | | | | | | | | | | | | |
| | | 3:51935206 | rs758330430 | G | A | WGS | 0,682 | 0 % | 0,00 % | ≤3 | 571 | 4,37 | 1,69 | 9,98E-03 | 0 | | | | | | 0 % | 9,04 | 3,19 | 4,63E-03 | 0 | 1 | 6 | 0,1 % | 0,800 | 1,30 | 5,40E-01 | 1 | 5 | 0,1 % | -1,299 | 1,84 | 4,81E-01 | 1 | 5 | 0,1 % | -0,954 | 3,19 | 7,65E-01 | 319 | 0,06 % | 0,510 | 0,27 | 5,59E-02 | 319 | 0,06 % | 0,560 | 0,28 | 4,18E-02 | 319 | 0,06 % | 0,518 | 0,29 | 7,33E-02 | 302 | 0,06 % | 0,938 | 0,55 | 8,53E-02 |
| | | | | | | WES | 0,839 | 0 % | 0,00 % | ≤3 | 480 | | | | | | | | | | | | | | | | | | | | | | | | | | | | | | | | | | | | | | | | | | | | | | | | | | |
| | | 3:51935641 | rs140029162 | G | A | WES | 0,830 | 0 % | 0,00 % | ≤3 | 480 | 5,91 | 2,67 | 2,68E-02 | 0 | 0 % | 8,36 | 3,08 | 6,71E-03 | 0 | | | | | | 1 | 8 | 0,1 % | -1,095 | 1,00 | 2,75E-01 | 1 | 8 | 0,1 % | -0,968 | 1,26 | 4,42E-01 | 1 | 8 | 0,1 % | -0,889 | 2,18 | 6,83E-01 | 272 | 0,05 % | 0,127 | 0,28 | 6,55E-01 | 271 | 0,05 % | 0,060 | 0,30 | 8,40E-01 | 271 | 0,05 % | -0,094 | 0,31 | 7,61E-01 | 261 | 0,05 % | 0,590 | 0,57 | 2,96E-01 |
| | | 3:51937282 | rs145537768 | G | A | WGS | 0,773 | 0 % | 0,00 % | ≤3 | 571 | 4,43 | 2,39 | 6,44E-02 | 0 | 0 % | 6,60 | 2,79 | 1,80E-02 | 0 | | | | | | | | | | | | | | | | | | | | | | | | | | | | | | | | | | | | | | | | | | | |
| | | 3:51941864 | . | A | G | WES | -0,182 | 0 % | 0,00 % | ≤3 | 480 | -1,23 | 2,60 | 6,35E-01 | 0 | 0 % | -1,17 | 2,96 | 6,93E-01 | 0 | 0 % | -1,09 | 3,94 | 7,83E-01 | 0 | | | | | | | | | | | | | | | | | | | | | | | | | | | | | | | | | | | | | | |
| | | 3:51941879 | rs199803038 | G | A | WGS | 0,682 | 0 % | 0,00 % | ≤3 | 571 | 1,81 | 1,79 | 3,10E-01 | 20,6 | 0 % | 2,70 | 1,99 | 1,75E-01 | 34,2 | 0 % | -1,05 | 5,53 | 8,50E-01 | 0 | | | | | | | | | | | | | | | | | | | | | | | | | | | | | | | | | | | | | | |
| | | | | | | WES | -0,113 | 0 % | 0,00 % | ≤3 | 480 | | | | | | | | | | | | | | | | | | | | | | | | | | | | | | | | | | | | | | | | | | | | | | | | | | |
| PAV 5% | *ANK1* | 8:41668453 | rs750580242 | A | T | WGS | 0,682 | 0 % | 0,00 % | ≤3 | 571 | 3,18 | 2,16 | 1,41E-01 | 0 | | | | | | 0 % | 9,04 | 3,19 | 4,63E-03 | 0 | | | | | | | | | | | | | | | | | | | | | | | | | | | | | | | | | | | | | | |
| | | 8:41672752 | rs751965455 | C | T | WES | -0,181 | 0 % | 0,00 % | ≤3 | 480 | -1,24 | 2,62 | 6,36E-01 | 0 | 0 % | -1,19 | 2,87 | 6,78E-01 | 0 | 0 % | -1,05 | 5,24 | 8,41E-01 | 0 | | | | | | | | | | | | | | | | | | | | | | | | | | | | | | | | | | | | | | |
| | | 8:41672844 | rs146416859 | G | A | WES | -0,179 | 0 % | 0,00 % | ≤3 | 480 | 1,82 | 1,76 | 3,01E-01 | 59,2 | 0 % | 3,33 | 2,12 | 1,16E-01 | 76,2 | 0 % | -1,06 | 4,97 | 8,31E-01 | 0 | 1 | 15 | 0,2 % | 0,505 | 0,73 | 4,91E-01 | 1 | 14 | 0,2 % | 0,624 | 0,95 | 5,13E-01 | 1 | 14 | 0,2 % | 3,774 | 1,65 | 2,20E-02 | 716 | 0,14 % | 0,014 | 0,17 | 9,36E-01 | 716 | 0,14 % | 0,016 | 0,17 | 9,27E-01 | 716 | 0,14 % | -0,020 | 0,18 | 9,13E-01 | 691 | 0,14 % | 0,048 | 0,33 | 8,84E-01 |
| | | | | | | WGS | 0,767 | 0 % | 0,00 % | ≤3 | 571 | 1,82 | 1,76 | 3,01E-01 | 59,2 | | | | | | | | | | | | | | | | | | | | | | | | | | | | | | | | | | | | | | | | | | | | | | |
| | | 8:41686157 | rs34664882 | G | A | WES | -1,280 | 2 % | 0,00 % | 15 | 480 | 1,15 | 0,52 | 2,75E-02 | 91,8 | 1 % | 1,17 | 0,63 | 6,44E-02 | 88 | 1 % | 1,07 | 1,04 | 3,03E-01 | 85,3 | 0,36 | 141 | 1,8 % | 0,114 | 0,24 | 6,33E-01 | 0,32 | 132 | 1,7 % | 0,055 | 0,31 | 8,59E-01 | 0,29 | 125 | 1,7 % | -0,758 | 0,55 | 1,70E-01 | 8250 | 1,63 % | -0,068 | 0,05 | 1,73E-01 | 8251 | 1,63 % | -0,053 | 0,05 | 2,99E-01 | 8245 | 1,63 % | -0,063 | 0,05 | 2,43E-01 | 8011 | 1,64 % | -0,113 | 0,10 | 2,55E-01 |
| | | | | | | WGS | 5,510 | 1 % | 0,00 % | 13 | 571 | | | | | | | | | | | | | | | | | | | | | | | | | | | | | | | | | | | | | | | | | | | | | | | | | | |
| | | 8:41688189 | rs199760447 | C | T | WES | 0,882 | 0 % | 0,00 % | ≤3 | 480 | 8,56 | 3,12 | 5,99E-03 | 0 | 0 % | 12,16 | 3,64 | 8,41E-04 | 0 | | | | | | 1 | 6 | 0,1 % | -1,349 | 1,16 | 2,44E-01 | 1 | 6 | 0,1 % | -1,215 | 1,46 | 4,04E-01 | 1 | 6 | 0,1 % | -1,140 | 2,52 | 6,50E-01 | 257 | 0,05 % | 0,110 | 0,28 | 6,95E-01 | 257 | 0,05 % | 0,129 | 0,29 | 6,57E-01 | 257 | 0,05 % | 0,170 | 0,30 | 5,75E-01 | 243 | 0,05 % | -0,358 | 0,59 | 5,47E-01 |
| | | 8:41688538 | rs201439151 | A | G | WGS | -0,450 | 0 % | 0,00 % | ≤3 | 571 | -1,29 | 1,44 | 3,71E-01 | 0 | 0 % | -1,16 | 1,78 | 5,16E-01 | 0 | 0 % | -1,09 | 2,39 | 6,47E-01 | 0 | 1 | 11 | 0,2 % | -0,321 | 0,86 | 7,08E-01 | 1 | 11 | 0,2 % | 0,500 | 1,08 | 6,42E-01 | 1 | 10 | 0,1 % | -0,976 | 1,95 | 6,16E-01 | 917 | 0,18 % | -0,277 | 0,15 | 6,22E-02 | 916 | 0,18 % | -0,274 | 0,15 | 7,46E-02 | 915 | 0,18 % | -0,248 | 0,16 | 1,23E-01 | 898 | 0,18 % | -0,361 | 0,29 | 2,19E-01 |
| | | | | | | WES | -0,172 | 0 % | 0,00 % | ≤3 | 480 | | | | | | | | | | | | | | | | | | | | | | | | | | | | | | | | | | | | | | | | | | | | | | | | | |
| | | 8:41690485 | rs10093583 | T | C | WGS | 1,191 | 0 % | 0,00 % | 4 | 571 | 0,96 | 1,06 | 3,63E-01 | 46,1 | 0 % | 0,75 | 1,39 | 5,86E-01 | 11 | 0 % | -1,10 | 2,00 | 5,84E-01 | 0 | 1 | 41 | 0,5 % | -0,228 | 0,45 | 6,08E-01 | 1 | 39 | 0,5 % | -0,316 | 0,57 | 5,82E-01 | 1 | 38 | 0,5 % | -0,636 | 1,00 | 5,26E-01 | 2757 | 0,54 % | 0,031 | 0,09 | 7,20E-01 | 2756 | 0,54 % | 0,056 | 0,09 | 5,29E-01 | 2753 | 0,54 % | 0,042 | 0,09 | 6,55E-01 | 2675 | 0,55 % | -0,054 | 0,17 | 7,51E-01 |
| | | | | | | WES | -0,332 | 0 % | 0,00 % | ≤3 | 480 | | | | | | | | | | | | | | | | | | | | | | | | | | | | | | | | | | | | | | | | | | | | | | | | | |
| | | 8:41690553 | rs779805849 | A | T | WGS | 0,491 | 0 % | 0,00 % | ≤3 | 571 | 1,25 | 1,59 | 4,34E-01 | 0 | 0 % | 2,62 | 1,95 | 1,79E-01 | 0 | 0 % | -1,04 | 4,34 | 8,11E-01 | 0 | 1 | 5 | 0,1 % | 3,186 | 1,34 | 1,71E-02 | 1 | 4 | 0,1 % | 3,763 | 1,89 | 4,60E-02 | 1 | 4 | 0,1 % | 15,483 | 3,28 | 2,43E-06 | 684 | 0,14 % | -0,165 | 0,17 | 3,44E-01 | 684 | 0,14 % | -0,242 | 0,18 | 1,80E-01 | 684 | 0,14 % | -0,138 | 0,19 | 4,65E-01 | 666 | 0,14 % | -0,003 | 0,34 | 9,92E-01 |
| | | 8:41692743 | rs148275567 | G | A | WGS | 0,118 | 0 % | 0,00 % | ≤3 | 571 | 2,14 | 0,93 | 2,21E-02 | 79,1 | 0 % | 3,31 | 1,07 | 1,95E-03 | 85,1 | 0 % | -1,11 | 2,39 | 6,42E-01 | 0 | 1 | 11 | 0,1 % | -0,020 | 0,86 | 9,82E-01 | 1 | 11 | 0,1 % | 0,804 | 1,08 | 4,54E-01 | 1 | 10 | 0,1 % | -0,716 | 1,95 | 7,13E-01 | 1056 | 0,21 % | -0,165 | 0,14 | 2,28E-01 | 1056 | 0,21 % | -0,128 | 0,14 | 3,65E-01 | 1055 | 0,21 % | -0,089 | 0,15 | 5,45E-01 | 1028 | 0,21 % | -0,231 | 0,27 | 3,96E-01 |
| | | | | | | WES | 2,331 | 0 % | 0,00 % | 4 | 480 | | | | | | | | | | | | | | | | | | | | | | | | | | | | | | | | | | | | | | | | | | | | | | | | | |
| | | 8:41694031 | rs117516263 | G | A | WES | 0,964 | 1 % | 0,00 % | 12 | 480 | -0,14 | 0,47 | 7,67E-01 | 25,5 | 2 % | -0,48 | 0,60 | 4,29E-01 | 0 | 2 % | 0,34 | 0,82 | 6,77E-01 | 69,4 | 0,55 | 114 | 1,4 % | 0,330 | 0,27 | 2,21E-01 | 0,51 | 105 | 1,4 % | 0,240 | 0,35 | 4,96E-01 | 0,50 | 102 | 1,4 % | 1,109 | 0,62 | 7,34E-02 | 6980 | 1,38 % | -0,086 | 0,05 | 1,11E-01 | 6977 | 1,38 % | -0,102 | 0,06 | 6,76E-02 | 6971 | 1,38 % | -0,105 | 0,06 | 7,31E-02 | 6785 | 1,39 % | -0,001 | 0,11 | 9,93E-01 |
| | | | | | | WGS | -1,600 | 2 % | 0,00 % | 19 | 571 | | | | | | | | | | | | | | | | | | | | | | | | | | | | | | | | | | | | | | | | | | | | | | | | | |
| | | 8:41695273 | rs202226361 | C | T | WGS | -0,150 | 0 % | 0,00 % | 5 | 571 | -0,18 | 1,09 | 8,70E-01 | 0 | 0 % | -1,17 | 1,58 | 4,56E-01 | 0 | 0 % | -1,11 | 2,09 | 5,96E-01 | 0 | 1 | 21 | 0,3 % | 1,193 | 0,62 | 5,46E-02 | 1 | 18 | 0,2 % | 0,459 | 0,84 | 5,86E-01 | 1 | 17 | 0,2 % | -0,392 | 1,50 | 7,94E-01 | 1510 | 0,30 % | 0,005 | 0,12 | 9,65E-01 | 1510 | 0,30 % | 0,028 | 0,12 | 8,18E-01 | 1510 | 0,30 % | 0,057 | 0,13 | 6,54E-01 | 1466 | 0,30 % | -0,280 | 0,23 | 2,20E-01 |
| | | 8:41696381 | rs767717861 | G | A | WES | 0,892 | 0 % | 0,00 % | ≤3 | 480 | 9,37 | 3,24 | 3,84E-03 | 0 | 0 % | 13,35 | 3,80 | 4,45E-04 | 0 | | | | | | | | | | | | | | | | | | | | | | | | | | | | | | | | | | | | | | | | | | | |
| | | 8:41696385 | . | G | T | WES | -0,447 | 0 % | 0,00 % | ≤3 | 480 | -1,21 | 1,64 | 4,63E-01 | 0 | 0 % | -1,15 | 1,89 | 5,43E-01 | 0 | 0 % | -1,06 | 3,09 | 7,33E-01 | 0 | | | | | | | | | | | | | | | | | | | | | | | | | | | | | | | | | | | | | | |
| | | 8:41696393 | . | G | T | WES | 0,618 | 0 % | 0,00 % | ≤3 | 480 | 1,91 | 1,76 | 2,78E-01 | 0 | 0 % | 3,16 | 2,07 | 1,27E-01 | 0 | 0 % | -1,05 | 3,68 | 7,75E-01 | 0 | | | | | | | | | | | | | | | | | | | | | | | | | | | | | | | | | | | | | | |
| | | 8:41708920 | rs2304877 | C | T | WES | -4,882 | 4 % | 0,00 % | 41 | 480 | 0,30 | 0,32 | 3,47E-01 | 93,8 | 4 % | 0,29 | 0,40 | 4,63E-01 | 93,5 | 4 % | -0,01 | 0,61 | 9,85E-01 | 31,9 | 0,68 | 317 | 4,0 % | -0,004 | 0,16 | 9,82E-01 | 0,67 | 300 | 4,0 % | -0,083 | 0,21 | 6,93E-01 | 0,80 | 294 | 4,1 % | 0,413 | 0,37 | 2,62E-01 | 20065 | 3,96 % | 0,004 | 0,03 | 9,13E-01 | 20060 | 3,96 % | 0,007 | 0,03 | 8,25E-01 | 20049 | 3,96 % | -0,001 | 0,03 | 9,82E-01 | 19374 | 3,96 % | 0,020 | 0,06 | 7,53E-01 |
| | | | | | | WGS | 7,816 | 3 % | 0,00 % | 32 | 571 | | | | | | | | | | | | | | | | | | | | | | | | | | | | | | | | | | | | | | | | | | | | | | | | | | |
| | | 8:41714174 | rs61753679 | G | T | WES | -1,245 | 2 % | 0,00 % | 22 | 480 | -0,09 | 0,42 | 8,38E-01 | 0 | 2 % | -0,13 | 0,52 | 8,06E-01 | 0 | 2 % | 0,06 | 0,77 | 9,38E-01 | 59,1 | 0,22 | 120 | 1,6 % | 0,188 | 0,26 | 4,74E-01 | 0,56 | 112 | 1,5 % | 0,081 | 0,34 | 8,13E-01 | 0,20 | 111 | 1,6 % | 1,564 | 0,59 | 8,09E-03 | 8343 | 1,65 % | -0,013 | 0,05 | 7,92E-01 | 8338 | 1,65 % | -0,008 | 0,05 | 8,72E-01 | 8337 | 1,65 % | -0,016 | 0,05 | 7,66E-01 | 8059 | 1,65 % | -0,053 | 0,10 | 5,89E-01 |
| | | | | | | WGS | 0,756 | 2 % | 0,00 % | 20 | 571 | | | | | | | | | | | | | | | | | | | | | | | | | | | | | | | | | | | | | | | | | | | | | | | | | | |
| | | 8:41715748 | rs34387324 | G | A | WGS | -0,111 | 0 % | 0,00 % | ≤3 | 571 | -1,20 | 1,63 | 4,61E-01 | 0 | 0 % | -1,14 | 1,91 | 5,53E-01 | 0 | 0 % | -1,07 | 2,67 | 6,87E-01 | 0 | 1 | 10 | 0,1 % | 0,094 | 0,97 | 9,23E-01 | 1 | 10 | 0,1 % | 1,140 | 1,22 | 3,49E-01 | 1 | 9 | 0,1 % | -0,711 | 2,23 | 7,50E-01 | 1206 | 0,24 % | 0,174 | 0,13 | 1,77E-01 | 1205 | 0,24 % | 0,154 | 0,13 | 2,48E-01 | 1203 | 0,24 % | 0,240 | 0,14 | 8,57E-02 | 1139 | 0,23 % | -0,042 | 0,27 | 8,73E-01 |
| | | | | | | WES | -0,341 | 0 % | 0,00 % | ≤3 | 480 | | | | | | | | | | | | | | | | | | | | | | | | | | | | | | | | | | | | | | | | | | | | | | | | | | |
| | | 8:41715770 | rs142690258 | T | C | WES | 0,089 | 1 % | 0,00 % | 5 | 480 | 1,45 | 0,74 | 5,11E-02 | 52,5 | 1 % | 1,82 | 1,00 | 6,89E-02 | 36,6 | 0 % | 3,01 | 1,43 | 3,56E-02 | 86,2 | 1 | 21 | 0,3 % | -0,375 | 0,62 | 5,46E-01 | 1 | 21 | 0,3 % | 0,112 | 0,78 | 8,86E-01 | 1 | 20 | 0,3 % | -0,533 | 1,38 | 7,00E-01 | 2327 | 0,46 % | -0,034 | 0,09 | 7,15E-01 | 2327 | 0,46 % | -0,039 | 0,10 | 6,87E-01 | 2326 | 0,46 % | -0,060 | 0,10 | 5,56E-01 | 2252 | 0,46 % | 0,297 | 0,19 | 1,18E-01 |
| | | | | | | WGS | 2,538 | 1 % | 0,00 % | 7 | 571 | | | | | | | | | | | | | | | | | | | | | | | | | | | | | | | | | | | | | | | | | | | | | | | | | | |
| | | 8:41716970 | rs140085544 | C | T | WGS | 0,901 | 0 % | 0,00 % | ≤3 | 571 | 4,97 | 1,75 | 4,59E-03 | 69,9 | 0 % | 4,65 | 2,41 | 5,34E-02 | 0 | 0 % | 12,33 | 3,66 | 7,51E-04 | 96,6 | 1 | 17 | 0,2 % | 0,886 | 0,69 | 1,99E-01 | 1 | 15 | 0,2 % | 0,459 | 0,92 | 6,18E-01 | 1 | 14 | 0,2 % | -0,488 | 1,65 | 7,67E-01 | 1101 | 0,22 % | -0,071 | 0,14 | 5,99E-01 | 1101 | 0,22 % | -0,042 | 0,14 | 7,66E-01 | 1101 | 0,22 % | -0,007 | 0,15 | 9,63E-01 | 1056 | 0,22 % | -0,392 | 0,27 | 1,45E-01 |
| | | | | | | WES | 0,716 | 0 % | 0,00 % | ≤3 | 480 | | | | | | | | | | | | | | | | | | | | | | | | | | | | | | | | | | | | | | | | | | | | | | | | | | |
| | | 8:41717030 | rs769619019 | T | A | WES | -0,167 | 0 % | 0,00 % | ≤3 | 480 | -1,21 | 2,69 | 6,53E-01 | 0 | 0 % | -1,14 | 3,10 | 7,12E-01 | 0 | 0 % | -1,05 | 4,98 | 8,33E-01 | 0 | | | | | | | | | | | | | | | | | | | | | | | | | | | | | | | | | | | | | | |
| | | 8:41717626 | . | G | A | WGS | -0,138 | 0 % | 0,00 % | ≤3 | 571 | -1,17 | 2,92 | 6,88E-01 | 0 | 0 % | -1,14 | 3,22 | 7,23E-01 | 0 | 0 % | -1,04 | 6,16 | 8,66E-01 | 0 | | | | | | | | | | | | | | | | | | | | | | | | | | | | | | | | | | | | | | |
| | | 8:41717683 | . | T | C | WGS | -0,135 | 0 % | 0,00 % | ≤3 | 571 | -1,16 | 2,94 | 6,93E-01 | 0 | 0 % | -1,10 | 3,60 | 7,61E-01 | 0 | 0 % | -1,04 | 5,49 | 8,50E-01 | 0 | | | | | | | | | | | | | | | | | | | | | | | | | | | | | | | | | | | | | | |
| | | 8:41718164 | . | T | C | WGS | -0,228 | 0 % | 0,00 % | ≤3 | 571 | -1,30 | 2,39 | 5,86E-01 | 0 | 0 % | -1,18 | 2,84 | 6,78E-01 | 0 | 0 % | -1,07 | 4,18 | 7,97E-01 | 0 | | | | | | | | | | | | | | | | | | | | | | | | | | | | | | | | | | | | | | |
| | | 8:41724513 | rs61735313 | G | T | WGS | 2,589 | 1 % | 0,00 % | 11 | 571 | 1,41 | 0,68 | 3,79E-02 | 0 | 1 % | 2,13 | 0,83 | 9,72E-03 | 69,3 | 1 % | -1,11 | 1,55 | 4,73E-01 | 0 | 1 | 77 | 1,0 % | -0,203 | 0,33 | 5,33E-01 | 1 | 73 | 1,0 % | -0,341 | 0,42 | 4,18E-01 | 1 | 72 | 1,0 % | 0,118 | 0,73 | 8,73E-01 | 3998 | 0,79 % | -0,013 | 0,07 | 8,51E-01 | 3998 | 0,79 % | -0,035 | 0,07 | 6,36E-01 | 3998 | 0,79 % | -0,049 | 0,08 | 5,19E-01 | 3847 | 0,79 % | 0,006 | 0,14 | 9,67E-01 |
| | | | | | | WES | 0,460 | 0 % | 0,00 % | 4 | 480 | | | | | | | | | | | | | | | | | | | | | | | | | | | | | | | | | | | | | | | | | | | | | | | | | | |

**Supplementary Table 13.** Variants in discovered stroke-risk SKAT-O genes with additional DKD adjustment; and replication in T1D (FinnDiane) and general populaiton (FinnGen)

*Note: Stroke subtype analyses may have included additional variants (i.e. passed MAF threshold in smaller data)*

FinnDiane discovery cohort WES+WGS; analysed with score test and, when possible, fixed-effects meta-analysis (T1D specific)

| MODEL | SetID | Position | SNPID | Major | Minor | metaSKAT (stroke) Data | Score | MAF | MissingRate | MAC | N | Stroke Beta | SE | *p*-value | Het $I^2$ | Ischemic stroke MAF | Beta | SE | *p*-value | Het $I^2$ | Hemorrhagic stroke MAF | Beta | SE | *p*-value | Het $I^2$ |
|---|---|---|---|---|---|---|---|---|---|---|---|---|---|---|---|---|---|---|---|---|---|---|---|---|---|
| PTV 5% | *ARPC5* | 1:183623459 | rs16861256 | A | G | WGS | 8,984 | 3 % | 0,00 % | 35 | 571 | 1,88 | 0,46 | 4,02E-05 | 0 | 3 % | 1,79 | 0,62 | 4,25E-03 | 0 | 2 % | 1,91 | 1,01 | 5,79E-02 | 0 |
| | | 1:183630530 | . | CTTCATT. | C | WGS | -0,789 | 0 % | 0,00 % | 4 | 571 | -1,48 | 1,37 | 2,80E-01 | 0 | 0 % | -1,35 | 1,64 | 4,10E-01 | 0 | 0 % | -1,18 | 2,10 | 5,74E-01 | 0 |
| PAV 5% | *APOBEC4* | 1:183647731 | rs139524898 | A | G | WGS | -0,104 | 0 % | 0,00 % | ≤3 | 571 | -1,18 | 2,07 | 5,68E-01 | 0 | 0 % | -1,13 | 2,47 | 6,48E-01 | 0 | 0 % | -1,05 | 3,83 | 7,83E-01 | 0 |
| | | | | | | WES | -0,173 | 0 % | 0,42 % | ≤3 | 480 | | | | | | | | | | | | | | |
| | | 1:183647749 | rs10911390 | C | T | WGS | 8,984 | 3 % | 0,00 % | 35 | 571 | 1,57 | 0,39 | 4,71E-05 | 36,8 | 2 % | 1,35 | 0,51 | 8,67E-03 | 34,3 | 2 % | 1,90 | 0,78 | 1,48E-02 | 0 |
| | | | | | | WES | 1,581 | 2 % | 0,42 % | 19 | 480 | | | | | | | | | | | | | | |
| | | 1:183647959 | rs10911391 | G | A | WGS | 8,984 | 3 % | 0,00 % | 35 | 571 | 1,57 | 0,39 | 4,71E-05 | 36,8 | 2 % | 1,35 | 0,51 | 8,67E-03 | 34,3 | 2 % | 1,90 | 0,78 | 1,48E-02 | 0 |
| | | | | | | WES | 1,581 | 2 % | 0,42 % | 19 | 480 | | | | | | | | | | | | | | |
| | | 1:183648558 | rs16861394 | G | A | WES | 1,581 | 2 % | 0,42 % | 19 | 480 | 1,57 | 0,39 | 4,71E-05 | 36,8 | 2 % | 1,35 | 0,51 | 8,67E-03 | 34,3 | 2 % | 1,90 | 0,78 | 1,48E-02 | 0 |
| | | | | | | WGS | 8,984 | 3 % | 0,00 % | 35 | 571 | | | | | | | | | | | | | | |
| | | 1:183648588 | rs776661029 | T | G | WGS | 0,732 | 0 % | 0,00 % | ≤3 | 571 | 1,45 | 1,63 | 3,75E-01 | 45,4 | 0 % | 2,73 | 1,97 | 1,67E-01 | 70,5 | 0 % | -1,08 | 3,10 | 7,28E-01 | 0 |
| | | | | | | WES | -0,188 | 0 % | 0,42 % | ≤3 | 480 | | | | | | | | | | | | | | |
| | | 1:183648589 | . | GT | G | WES | 0,815 | 0 % | 0,42 % | ≤3 | 480 | 5,46 | 2,59 | 3,49E-02 | 0 | | | | | | 0 % | 20,73 | 4,67 | 8,97E-06 | 0 |
| | | 1:183648660 | rs182486585 | G | C | WGS | -0,064 | 0 % | 0,00 % | ≤3 | 571 | -1,07 | 4,11 | 7,94E-01 | 0 | 0 % | -1,04 | 5,58 | 8,52E-01 | 0 | 0 % | -1,02 | 8,97 | 9,10E-01 | 0 |
| | | 1:183653304 | . | T | C | WES | 0,902 | 0 % | 0,42 % | ≤3 | 480 | 10,38 | 3,39 | 2,21E-03 | 0 | 0 % | 16,50 | 4,19 | 8,34E-05 | 0 | | | | | |
| PTV 1% & PTV 5% | *HAS1* | 19:51713664 | . | G | T | WES | -0,304 | 0 % | 0,42 % | ≤3 | 480 | -1,21 | 2,00 | 5,44E-01 | 0 | 0 % | -1,15 | 2,30 | 6,16E-01 | 0 | 0 % | -1,09 | 3,15 | 7,28E-01 | 0 |
| | | 19:51713853 | . | G | T | WES | -0,159 | 0 % | 0,63 % | ≤3 | 480 | -1,21 | 2,76 | 6,61E-01 | 0 | 0 % | -1,14 | 3,24 | 7,24E-01 | 0 | 0 % | -1,08 | 4,50 | 8,11E-01 | 0 |
| | | 19:51716967 | rs1396240967 | C | T | WES | 0,877 | 0 % | 0,42 % | ≤3 | 480 | 8,28 | 3,07 | 7,02E-03 | 0 | 0 % | 13,52 | 3,82 | 4,06E-04 | 0 | | | | | |
| | | 19:51723924 | rs58597876 | C | CACACA | WGS | 2,551 | 0 % | 0,00 % | ≤3 | 571 | 7,05 | 1,66 | 2,23E-05 | 0 | 0 % | 11,03 | 2,02 | 4,71E-08 | 0 | | | | | |
| PAV 5% | *UTS2* | 1:7849677 | rs2890565 | C | T | WES | -0,958 | 1 % | 0,00 % | 13 | 480 | 1,56 | 0,57 | 5,91E-03 | 93,1 | 1 % | 2,06 | 0,69 | 2,63E-03 | 93,1 | 1 % | 0,19 | 1,14 | 8,68E-01 | 67,8 |
| | | | | | | WGS | 5,830 | 1 % | 0,00 % | 12 | 571 | | | | | | | | | | | | | | |
| | | 1:7850908 | rs201977733 | C | T | WGS | -0,068 | 0 % | 0,00 % | ≤3 | 571 | -1,08 | 3,98 | 7,87E-01 | 0 | 0 % | -1,04 | 5,81 | 8,59E-01 | 0 | 0 % | -1,02 | 7,71 | 8,95E-01 | 0 |
| PAV 1% & PAV 5% | *SLC18A2* | 10:117253425 | rs184073241 | C | T | WGS | -0,188 | 0 % | 0,00 % | ≤3 | 571 | -1,25 | 2,58 | 6,28E-01 | 0 | 0 % | -1,16 | 3,09 | 7,08E-01 | 0 | 0 % | -1,07 | 4,54 | 8,13E-01 | 0 |
| | | 10:117257799 | rs199750100 | T | A | WGS | 0,070 | 0 % | 0,00 % | 5 | 571 | 3,01 | 0,85 | 3,90E-04 | 90,8 | 1 % | 3,93 | 1,02 | 1,12E-04 | 91,5 | 0 % | 3,34 | 2,10 | 1,12E-01 | 79,1 |
| | | | | | | WES | 4,112 | 1 % | 0,00 % | 6 | 480 | | | | | | | | | | | | | | |
| | | 10:117267034 | rs367902584 | G | A | WGS | -0,086 | 0 % | 0,00 % | ≤3 | 571 | -1,10 | 3,58 | 7,58E-01 | 0 | 0 % | -1,06 | 4,54 | 8,15E-01 | 0 | 0 % | -1,03 | 7,57 | 8,92E-01 | 0 |
| PAV 1% & PAV 5% | *MTRNR2L7* | 10:37602452 | rs567342729 | T | C | WGS | 0,795 | 0 % | 0,00 % | ≤3 | 571 | 4,41 | 2,36 | 6,11E-02 | 0 | 0 % | -1,08 | 4,12 | 7,93E-01 | 0 | 0 % | 23,97 | 5,00 | 1,64E-06 | 0 |
| | | 10:37602666 | rs12241774 | G | T | WGS | 3,695 | 1 % | 0,00 % | 8 | 571 | 3,81 | 1,01 | 1,77E-04 | 0 | 1 % | 5,72 | 1,32 | 1,49E-05 | 0 | 0 % | 7,75 | 2,97 | 9,04E-03 | 0 |
| PAV 1% & PAV 5% | *MAP3K12* | 12:53481189 | rs377738660 | G | T | WES | 0,910 | 0 % | 0,00 % | ≤3 | 480 | 11,32 | 3,53 | 1,32E-03 | 0 | | | | | | 0 % | 20,71 | 4,67 | 9,11E-06 | 0 |
| | | 12:53481235 | rs115635603 | C | T | WGS | -0,816 | 0 % | 0,00 % | ≤3 | 571 | 0,32 | 0,86 | 7,08E-01 | 66,9 | 0 % | -1,04 | 1,11 | 3,49E-01 | 0 | 1 % | 2,61 | 1,34 | 5,17E-02 | 83,7 |
| | | | | | | WES | 1,251 | 1 % | 0,00 % | 7 | 480 | | | | | | | | | | | | | | |
| | | 12:53483170 | . | C | CT | WGS | -0,067 | 0 % | 0,00 % | ≤3 | 571 | -1,08 | 4,03 | 7,89E-01 | 0 | 0 % | -1,04 | 5,66 | 8,54E-01 | 0 | 0 % | -1,02 | 7,95 | 8,98E-01 | 0 |
| | | 12:53486534 | rs141065572 | G | T | WGS | 0,690 | 0 % | 0,00 % | ≤3 | 571 | 6,07 | 1,36 | 7,98E-06 | 64,4 | 0 % | 11,68 | 2,07 | 1,64E-08 | 0 | | | | | |
| | | | | | | WES | 2,597 | 0 % | 0,00 % | ≤3 | 480 | | | | | | | | | | | | | | |
| | | 12:53486535 | rs199977017 | T | C | WES | 2,597 | 0 % | 0,00 % | ≤3 | 480 | 6,07 | 1,36 | 7,98E-06 | 64,4 | 0 % | 11,68 | 2,07 | 1,64E-08 | 0 | | | | | |
| | | | | | | WGS | 0,690 | 0 % | 0,00 % | ≤3 | 571 | | | | | | | | | | | | | | |
| | | 12:53486960 | . | C | A | WES | -0,203 | 0 % | 0,00 % | ≤3 | 480 | -1,27 | 2,50 | 6,13E-01 | 0 | 0 % | -1,18 | 2,87 | 6,81E-01 | 0 | 0 % | -1,10 | 3,72 | 7,68E-01 | 0 |
| | | 12:53499441 | . | G | A | WES | -0,197 | 0 % | 0,00 % | ≤3 | 480 | -1,26 | 2,52 | 6,18E-01 | 0 | 0 % | -1,19 | 2,81 | 6,72E-01 | 0 | 0 % | -1,06 | 4,89 | 8,29E-01 | 0 |
| PAV 1% & PAV 5% | *TARBP2* | 12:53503093 | rs146234229 | G | C | WGS | -0,249 | 0 % | 0,00 % | ≤3 | 571 | -1,32 | 1,68 | 4,33E-01 | 0 | 0 % | -1,19 | 1,98 | 5,46E-01 | 0 | 0 % | -1,09 | 2,76 | 6,93E-01 | 0 |
| | | | | | | WES | -0,217 | 0 % | 0,00 % | ≤3 | 480 | | | | | | | | | | | | | | |
| | | 12:53504426 | rs149488319 | T | C | WGS | 0,690 | 0 % | 0,00 % | ≤3 | 571 | 6,07 | 1,36 | 7,98E-06 | 64,4 | 0 % | 11,68 | 2,07 | 1,64E-08 | 0 | | | | | |
| | | | | | | WES | 2,597 | 0 % | 0,00 % | ≤3 | 480 | | | | | | | | | | | | | | |
| PTV 1% & PTV 5% | *UACA* | 15:70667338 | rs781623644 | G | A | WES | 1,653 | 0 % | 0,42 % | ≤3 | 480 | 5,04 | 1,44 | 4,49E-04 | 0 | 0 % | 8,05 | 2,14 | 1,67E-04 | 53,7 | 0 % | -1,06 | 3,67 | 7,74E-01 | 0 |
| | | | | | | WGS | 0,790 | 0 % | 0,00 % | ≤3 | 571 | | | | | | | | | | | | | | |
| | | 15:70679607 | rs185763236 | C | A | WGS | 0,814 | 0 % | 0,00 % | ≤3 | 571 | 5,46 | 2,59 | 3,51E-02 | 0 | | | | | | | | | | |
| PAV 1% & PAV 5% | *GCDH* | 19:12891359 | . | G | T | WGS | 0,728 | 0 % | 0,00 % | ≤3 | 571 | 3,71 | 2,26 | 1,01E-01 | 0 | | | | | | 0 % | 14,50 | 3,95 | 2,40E-04 | 0 |
| | | 19:12896058 | rs149120354 | T | C | WES | 0,919 | 0 % | 0,42 % | ≤3 | 480 | 12,59 | 3,70 | 6,69E-04 | 0 | 0 % | 17,25 | 4,28 | 5,60E-05 | 0 | | | | | |
| | | 19:12896341 | rs201076343 | G | T | WES | -0,130 | 0 % | 0,42 % | ≤3 | 480 | -1,17 | 3,00 | 6,96E-01 | 0 | 0 % | -1,10 | 3,78 | 7,70E-01 | 0 | 0 % | -1,10 | 4,08 | 7,88E-01 | 0 |
| | | 19:12896401 | rs751742575 | C | T | WES | 0,906 | 0 % | 0,42 % | ≤3 | 480 | 10,75 | 3,45 | 1,81E-03 | 0 | 0 % | 18,47 | 4,42 | 2,94E-05 | 0 | | | | | |
| | | 19:12897009 | rs761135089 | G | A | WGS | 0,824 | 0 % | 0,00 % | ≤3 | 571 | 5,79 | 2,65 | 2,90E-02 | 0 | | | | | | | | | | |
| | | 19:12897824 | rs121434369 | C | T | WES | 0,809 | 0 % | 0,42 % | ≤3 | 480 | 5,36 | 2,57 | 3,72E-02 | 0 | 0 % | 8,09 | 3,04 | 7,87E-03 | 0 | | | | | |
| PAV 1% & PAV 5% | *CLEC4M* | 19:7765676 | rs200077446 | G | A | WGS | -0,078 | 0 % | 0,00 % | ≤3 | 571 | -1,09 | 3,73 | 7,70E-01 | 0 | 0 % | -1,05 | 4,84 | 8,28E-01 | 0 | 0 % | -1,02 | 7,94 | 8,98E-01 | 0 |
| | | 19:7765776 | rs113080783 | A | G | WES | -0,284 | 0 % | 1,00 % | ≤3 | 480 | -1,21 | 2,06 | 5,58E-01 | 0 | 0 % | -1,14 | 2,37 | 6,31E-01 | 0 | 0 % | -1,07 | 3,70 | 7,72E-01 | 0 |
| | | 19:7766121 | rs144783051 | A | G | WES | 0,838 | 0 % | 0,42 % | ≤3 | 480 | 6,23 | 2,73 | 2,23E-02 | 0 | | | | | | 0 % | 21,09 | 4,71 | 7,44E-06 | 0 |
| | | 19:7766155 | rs746560445 | G | GT | WGS | 0,695 | 0 % | 0,00 % | ≤3 | 571 | 3,31 | 2,18 | 1,29E-01 | 0 | 0 % | 4,58 | 2,42 | 5,91E-02 | 0 | | | | | |
| | | 19:7766659 | rs140767813 | G | A | WES | 0,910 | 0 % | 0,42 % | ≤3 | 480 | 11,32 | 3,53 | 1,34E-03 | 0 | | | | | | 0 % | 20,71 | 4,67 | 9,16E-06 | 0 |
| | | 19:7766667 | rs377334555 | C | T | WES | 0,807 | 0 % | 0,42 % | ≤3 | 480 | 5,31 | 2,57 | 3,85E-02 | 0 | | | | | | 0 % | 13,15 | 3,78 | 5,01E-04 | 0 |
| | | 19:7767549 | rs752738017 | C | T | WES | 1,832 | 0 % | 0,42 % | ≤3 | 480 | 12,15 | 2,58 | 2,38E-06 | 0 | 0 % | 18,47 | 4,42 | 2,94E-05 | 0 | 0 % | 50,78 | 7,20 | 1,74E-12 | 0 |
| | | 19:7768929 | rs184828145 | T | C | WGS | 0,846 | 0 % | 0,00 % | ≤3 | 571 | 6,63 | 2,80 | 1,79E-02 | 0 | | | | | | 0 % | 25,30 | 5,13 | 8,32E-07 | 0 |
| | | 19:7768947 | rs201740080 | T | C | WES | -0,144 | 0 % | 0,42 % | ≤3 | 480 | -1,16 | 2,20 | 5,99E-01 | 0 | 0 % | -1,11 | 2,71 | 6,82E-01 | 0 | 0 % | -1,05 | 4,16 | 8,01E-01 | 0 |
| | | | | | | WGS | -0,095 | 0 % | 0,00 % | ≤3 | 571 | | | | | | | | | | | | | | |
| | | 19:7768973 | rs753343180 | C | G | WGS | -0,191 | 0 % | 0,00 % | ≤3 | 571 | -1,26 | 2,57 | 6,24E-01 | 0 | 0 % | -1,13 | 3,47 | 7,45E-01 | 0 | 0 % | -1,08 | 4,53 | 8,11E-01 | 0 |
| PTV 1% & PTV 5% | *CHEK2* | 22:28687961 | rs587782684 | CG | C | WES | -0,200 | 0 % | 0,42 % | ≤3 | 480 | -1,26 | 2,51 | 6,15E-01 | 0 | 0 % | -1,18 | 2,89 | 6,84E-01 | 0 | 0 % | -1,11 | 3,64 | 7,61E-01 | 0 |
| | | 22:28695868 | rs555607708 | AG | A | WES | 1,683 | 1 % | 0,42 % | 7 | 480 | 2,77 | 0,72 | 1,10E-04 | 58,1 | 1 % | 3,62 | 0,98 | 2,34E-04 | 61,8 | 0 % | 3,75 | 1,56 | 1,62E-02 | 92,2 |
| | | | | | | WGS | 3,725 | 1 % | 0,00 % | 8 | 571 | | | | | | | | | | | | | | |
| PAV 1% | *LRRN1* | 3:3844558 | rs755350940 | C | T | WES | 0,933 | 0 % | 0,00 % | ≤3 | 480 | 1,49 | 1,66 | 3,68E-01 | 92,7 | 0 % | 2,43 | 1,91 | 2,03E-01 | 95,4 | 0 % | -1,07 | 3,43 | 7,54E-01 | 0 |
| | | | | | | WGS | -0,390 | 0 % | 0,00 % | ≤3 | 571 | | | | | | | | | | | | | | |
| | | 3:3845194 | rs142381203 | C | T | WGS | 0,760 | 0 % | 0,00 % | ≤3 | 571 | 4,21 | 2,35 | 7,38E-02 | 0 | | | | | | 0 % | 13,50 | 3,82 | 4,10E-04 | 0 |
| | | 3:3845210 | rs147007969 | C | T | WGS | 0,816 | 0 % | 0,00 % | ≤3 | 571 | 3,51 | 1,10 | 1,35E-03 | 0 | 0 % | 5,43 | 1,49 | 2,57E-04 | 80,9 | 0 % | 5,28 | 2,52 | 3,62E-02 | 45,4 |
| | | | | | | WES | 2,109 | 0 % | 0,00 % | 4 | 480 | | | | | | | | | | | | | | |
| | | 3:3845554 | rs141825989 | C | T | WES | 0,831 | 0 % | 0,00 % | ≤3 | 480 | 6,08 | 2,71 | 2,45E-02 | 0 | 0 % | 7,43 | 2,94 | 1,14E-02 | 0 | | | | | |
| | | 3:3845810 | rs770349026 | G | A | WES | 0,740 | 0 % | 0,00 % | ≤3 | 480 | 3,34 | 2,12 | 1,16E-01 | 0 | 0 % | 5,31 | 2,54 | 3,63E-02 | 0 | 0 % | -1,09 | 4,03 | 7,87E-01 | 0 |
| | | 3:3845966 | rs747455683 | C | T | WGS | 0,869 | 0 % | 0,00 % | ≤3 | 571 | 7,76 | 2,99 | 9,39E-03 | 0 | | | | | | 0 % | 24,63 | 5,07 | 1,18E-06 | 0 |
| | | 3:3846082 | rs543878366 | G | A | WES | -0,102 | 0 % | 0,00 % | ≤3 | 480 | -1,12 | 3,32 | 7,35E-01 | 0 | 0 % | -1,09 | 3,93 | 7,82E-01 | 0 | 0 % | -1,04 | 6,29 | 8,69E-01 | 0 |
| PAV 5% | *ANK1* | 8:41668453 | rs750580242 | A | T | WGS | 0,593 | 0 % | 0,00 % | ≤3 | 571 | 2,49 | 2,05 | 2,24E-01 | 0 | | | | | | 0 % | 6,55 | 2,79 | 1,88E-02 | 0 |
| | | 8:41672752 | rs751965455 | C | T | WES | -0,168 | 0 % | 0,00 % | ≤3 | 480 | -1,22 | 2,69 | 6,51E-01 | 0 | 0 % | -1,17 | 2,96 | 6,91E-01 | 0 | 0 % | -1,05 | 5,48 | 8,48E-01 | 0 |
| | | 8:41672844 | rs146416859 | G | A | WES | -0,119 | 0 % | 0,00 % | ≤3 | 480 | 1,83 | 1,78 | 3,05E-01 | 27,3 | 0 % | 3,21 | 2,10 | 1,26E-01 | 53,1 | 0 % | -1,04 | 6,28 | 8,69E-01 | 0 |
| | | | | | | WGS | 0,695 | 0 % | 0,00 % | ≤3 | 571 | | | | | | | | | | | | | | |
| | | 8:41686157 | rs34664882 | G | A | WES | -1,139 | 2 % | 0,00 % | 15 | 480 | 1,02 | 0,52 | 4,87E-02 | 88,7 | 1 % | 1,01 | 0,62 | 1,04E-01 | 81,4 | 1 % | 0,97 | 1,02 | 3,43E-01 | 80,3 |
| | | | | | | WGS | 4,954 | 1 % | 0,00 % | 13 | 571 | | | | | | | | | | | | | | |
| | | 8:41688189 | rs199760447 | C | T | WES | 0,849 | 0 % | 0,00 % | ≤3 | 480 | 6,67 | 2,80 | 1,73E-02 | 0 | 0 % | 9,40 | 3,25 | 3,77E-03 | 0 | | | | | |
| | | 8:41688538 | rs201439151 | A | G | WGS | -0,408 | 0 % | 0,00 % | ≤3 | 571 | -1,39 | 1,45 | 3,38E-01 | 0 | 0 % | -1,24 | 1,74 | 4,78E-01 | 0 | 0 % | -1,12 | 2,29 | 6,25E-01 | 0 |
| | | | | | | WES | -0,253 | 0 % | 0,00 % | ≤3 | 480 | | | | | | | | | | | | | | |
| | | 8:41690485 | rs10093583 | T | C | WGS | 1,233 | 0 % | 0,00 % | 4 | 571 | 1,02 | 1,09 | 3,47E-01 | 56,3 | 0 % | 0,67 | 1,37 | 6,24E-01 | 16,9 | 0 % | -1,12 | 2,03 | 5,83E-01 | 0 |
| | | | | | | WES | -0,367 | 0 % | 0,00 % | ≤3 | 480 | | | | | | | | | | | | | | |
| | | 8:41690553 | rs779805849 | A | T | WGS | 0,511 | 0 % | 0,00 % | ≤3 | 571 | 1,50 | 1,71 | 3,82E-01 | 0 | 0 % | 2,86 | 2,04 | 1,62E-01 | 0 | 0 % | -1,03 | 5,72 | 8,58E-01 | 0 |
| | | 8:41692743 | rs148275567 | G | A | WGS | 0,061 | 0 % | 0,00 % | ≤3 | 571 | 2,19 | 0,95 | 2,08E-02 | 82,1 | 0 % | 3,45 | 1,09 | 1,47E-03 | 86,9 | 0 % | -1,09 | 2,52 | 6,64E-01 | 0 |
| | | | | | | WES | 2,378 | 0 % | 0,00 % | 4 | 480 | | | | | | | | | | | | | | |
| | | 8:41694031 | rs117516263 | G | A | WES | 1,077 | 1 % | 0,00 % | 12 | 480 | -0,18 | 0,48 | 7,12E-01 | 44,9 | 2 % | -0,53 | 0,61 | 3,88E-01 | 0 | 2 % | 0,31 | 0,83 | 7,11E-01 | 73,6 |
| | | | | | | WGS | -1,850 | 2 % | 0,00 % | 19 | 571 | | | | | | | | | | | | | | |
| | | 8:41695273 | rs202226361 | C | T | WGS | -0,017 | 0 % | 0,00 % | 5 | 571 | -0,02 | 1,14 | 9,85E-01 | 0 | 0 % | -1,20 | 1,54 | 4,36E-01 | 0 | 0 % | -1,11 | 2,05 | 5,88E-01 | 0 |
| | | 8:41696381 | rs767717861 | G | A | WES | 0,863 | 0 % | 0,00 % | ≤3 | 480 | 7,40 | 2,93 | 1,15E-02 | 0 | 0 % | 10,47 | 3,40 | 2,11E-03 | 0 | | | | | |
| | | 8:41696385 | . | G | T | WES | -0,366 | 0 % | 0,00 % | ≤3 | 480 | -1,19 | 1,81 | 5,09E-01 | 0 | 0 % | -1,15 | 2,06 | 5,78E-01 | 0 | 0 % | -1,05 | 3,50 | 7,64E-01 | 0 |
| | | 8:41696393 | . | G | T | WES | 0,764 | 0 % | 0,00 % | ≤3 | 480 | 3,59 | 2,17 | 9,75E-02 | 0 | 0 % | 5,57 | 2,58 | 3,07E-02 | 0 | 0 % | -1,03 | 4,99 | 8,36E-01 | 0 |
| | | 8:41708920 | rs2304877 | C | T | WES | -4,818 | 4 % | 0,00 % | 41 | 480 | 0,39 | 0,33 | 2,40E-01 | 94,7 | 4 % | 0,37 | 0,41 | 3,73E-01 | 94,5 | 4 % | 0,0001 | 0,61 | 1,00E+00 | 29 |
| | | | | | | WGS | 8,349 | 3 % | 0,00 % | 32 | 571 | | | | | | | | | | | | | | |
| | | 8:41714174 | rs61753679 | G | T | WES | -1,373 | 2 % | 0,00 % | 22 | 480 | -0,16 | 0,42 | 7,09E-01 | 0 | 2 % | -0,19 | 0,52 | 7,22E-01 | 0 | 2 % | 0,02 | 0,76 | 9,80E-01 | 56,4 |
| | | | | | | WGS | 0,487 | 2 % | 0,00 % | 20 | 571 | | | | | | | | | | | | | | |
| | | 8:41715748 | rs34387324 | G | A | WGS | -0,065 | 0 % | 0,00 % | ≤3 | 571 | -1,32 | 1,57 | 3,98E-01 | 0 | 0 % | -1,22 | 1,82 | 5,03E-01 | 0 | 0 % | -1,13 | 2,34 | 6,29E-01 | 0 |
| | | | | | | WES | -0,475 | 0 % | 0,00 % | ≤3 | 480 | | | | | | | | | | | | | | |
| | | 8:41715770 | rs142690258 | T | C | WES | -0,138 | 1 % | 0,00 % | 5 | 480 | 1,31 | 0,74 | 7,57E-02 | 69,1 | 1 % | 1,82 | 1,01 | 7,11E-02 | 75 | 0 % | 2,12 | 1,28 | 9,93E-02 | 85,5 |
| | | | | | | WGS | 2,544 | 1 % | 0,00 % | 7 | 571 | | | | | | | | | | | | | | |
| | | 8:41716970 | rs140085544 | C | T | WGS | 0,822 | 0 % | 0,00 % | ≤3 | 571 | 3,90 | 1,60 | 1,49E-02 | 0 | 0 % | 4,32 | 2,34 | 6,49E-02 | 0 | 0 % | 8,92 | 3,16 | 4,78E-03 | 92,3 |
| | | | | | | WES | 0,700 | 0 % | 0,00 % | ≤3 | 480 | | | | | | | | | | | | | | |
| | | 8:41717030 | rs769619019 | T | A | WES | -0,227 | 0 % | 0,00 % | ≤3 | 480 | -1,31 | 2,40 | 5,85E-01 | 0 | 0 % | -1,21 | 2,72 | 6,57E-01 | 0 | 0 % | -1,08 | 4,15 | 7,94E-01 | 0 |
| | | 8:41717626 | . | G | A | WGS | -0,091 | 0 % | 0,00 % | ≤3 | 571 | -1,11 | 3,50 | 7,51E-01 | 0 | 0 % | -1,07 | 4,31 | 8,03E-01 | 0 | 0 % | -1,03 | 7,59 | 8,92E-01 | 0 |
| | | 8:41717683 | . | T | C | WGS | -0,082 | 0 % | 0,00 % | ≤3 | 571 | -1,09 | 3,66 | 7,65E-01 | 0 | 0 % | -1,05 | 5,15 | 8,39E-01 | 0 | 0 % | -1,03 | 6,97 | 8,83E-01 | 0 |
| | | 8:41718164 | . | T | C | WGS | -0,099 | 0 % | 0,00 % | ≤3 | 571 | -1,12 | 3,36 | 7,39E-01 | 0 | 0 % | -1,05 | 4,89 | 8,30E-01 | 0 | 0 % | -1,03 | 6,47 | 8,73E-01 | 0 |
| | | 8:41724513 | rs61735313 | G | T | WGS | 2,475 | 1 % | 0,00 % | 11 | 571 | 1,26 | 0,67 | 5,90E-02 | 0 | 1 % | 2,02 | 0,82 | 1,35E-02 | 65,3 | 1 % | -1,08 | 1,64 | 5,08E-01 | 0 |
| | | | | | | WES | 0,356 | 0 % | 0,00 % | 4 | 480 | | | | | | | | | | | | | | |

FinnDiane GWAS/genotyping replication using score test (T1D specific)

| Position | Stroke HWE | MAC | MAF | Beta | SE | *p*-value | Ischemic stroke HWE | MAC | MAF | Beta | SE | *p*-value | Hemorrhagic stroke HWE | MAC | MAF | Beta | SE | *p*-value |
|---|---|---|---|---|---|---|---|---|---|---|---|---|---|---|---|---|---|---|
| 1:183623459 | 0,49 | 183 | 2,4 % | -0,006 | 0,21 | 9,75E-01 | 0,47 | 174 | 2,4 % | -0,041 | 0,27 | 8,81E-01 | 0,47 | 172 | 2,5 % | 0,826 | 0,47 | 7,82E-02 |
| 1:183647749 | 0,50 | 187 | 2,5 % | -0,025 | 0,21 | 9,03E-01 | 0,48 | 178 | 2,5 % | -0,056 | 0,27 | 8,34E-01 | 0,49 | 176 | 2,5 % | 0,790 | 0,46 | 8,81E-02 |
| 1:183647959 | 0,50 | 187 | 2,5 % | -0,025 | 0,21 | 9,03E-01 | 0,48 | 178 | 2,5 % | -0,056 | 0,27 | 8,34E-01 | 0,49 | 176 | 2,5 % | 0,790 | 0,46 | 8,81E-02 |
| 1:183648558 | 0,50 | 187 | 2,5 % | -0,025 | 0,21 | 9,03E-01 | 0,48 | 178 | 2,5 % | -0,056 | 0,27 | 8,34E-01 | 0,49 | 176 | 2,5 % | 0,790 | 0,46 | 8,81E-02 |
| 1:183648588 | 1 | 9 | 0,1 % | 1,221 | 1,04 | 2,42E-01 | 1 | 7 | 0,1 % | -0,591 | 1,44 | 6,80E-01 | 1 | 8 | 0,1 % | 1,756 | 2,46 | 4,75E-01 |
| 1:7849677 | 0,049 | 112 | 1,5 % | 0,276 | 0,26 | 2,91E-01 | 0,038 | 104 | 1,4 % | 0,272 | 0,34 | 4,26E-01 | 0,029 | 97 | 1,4 % | -0,738 | 0,61 | 2,25E-01 |
| 10:117257799 | 1 | 34 | 0,5 % | -0,538 | 0,51 | 2,87E-01 | 1 | 34 | 0,5 % | -0,260 | 0,64 | 6,85E-01 | 1 | 33 | 0,5 % | -0,502 | 1,11 | 6,52E-01 |
| 10:37602666 | 1 | 38 | 0,5 % | -0,069 | 0,46 | 8,80E-01 | 1 | 37 | 0,5 % | 0,424 | 0,59 | 4,70E-01 | 1 | 34 | 0,5 % | -0,827 | 1,05 | 4,30E-01 |
| 12:53481189 | 1 | ≤3 | 0,0 % | -1,640 | 1,61 | 3,09E-01 | 1 | ≤3 | 0,0 % | -1,465 | 2,04 | 4,74E-01 | 1 | ≤3 | 0,0 % | -1,416 | 3,51 | 6,87E-01 |
| 12:53481235 | 1 | 28 | 0,4 % | -0,139 | 0,54 | 7,98E-01 | 1 | 28 | 0,4 % | 0,473 | 0,69 | 4,91E-01 | 1 | 26 | 0,4 % | -0,403 | 1,21 | 7,40E-01 |
| 12:53486534 | 1 | 12 | 0,2 % | 0,150 | 0,81 | 8,53E-01 | 1 | 11 | 0,1 % | -0,668 | 1,07 | 5,32E-01 | 1 | 12 | 0,2 % | 4,365 | 1,76 | 1,33E-02 |
| 12:53486535 | 1 | 12 | 0,2 % | 0,150 | 0,81 | 8,53E-01 | 1 | 11 | 0,1 % | -0,668 | 1,07 | 5,32E-01 | 1 | 12 | 0,2 % | 4,365 | 1,76 | 1,33E-02 |
| 12:53503093 | 1 | 7 | 0,1 % | 3,816 | 1,16 | 1,00E-03 | 1 | 5 | 0,0 % | 3,645 | 1,77 | 3,94E-02 | 1 | 5 | 0,1 % | 12,963 | 3,11 | 3,07E-05 |
| 12:53504426 | 1 | 12 | 0,2 % | 0,150 | 0,81 | 8,53E-01 | 1 | 11 | 0,1 % | -0,668 | 1,07 | 5,32E-01 | 1 | 12 | 0,2 % | 4,365 | 1,76 | 1,33E-02 |
| 15:70667338 | 1 | 7 | 0,1 % | -0,665 | 1,19 | 5,77E-01 | 1 | 6 | 0,1 % | -0,553 | 1,51 | 7,15E-01 | 1 | 7 | 0,1 % | -0,200 | 2,61 | 9,39E-01 |
| 15:70679607 | 1 | ≤3 | 0,0 % | 8,315 | 2,80 | 2,99E-03 | 1 | ≤3 | 0,0 % | 14,061 | 3,36 | 2,93E-05 | *Genotyped | | | | | |
| 19:12896058 | 1 | ≤3 | 0,0 % | 2,298 | 1,67 | 1,69E-01 | 1 | ≤3 | 0,0 % | -0,916 | 2,50 | 7,14E-01 | 1 | ≤3 | 0,0 % | -0,897 | 4,30 | 8,35E-01 |
| 19:12897824 | 1 | 6 | 0,1 % | 0,746 | 1,14 | 5,13E-01 | 1 | 6 | 0,1 % | 2,192 | 1,45 | 1,29E-01 | 1 | 5 | 0,1 % | -0,586 | 2,72 | 8,30E-01 |
| 19:7765676 | 1 | 9 | 0,1 % | 0,130 | 0,93 | 8,89E-01 | 1 | 9 | 0,1 % | 1,146 | 1,18 | 3,32E-01 | 1 | 8 | 0,1 % | -0,638 | 2,15 | 7,67E-01 |
| 19:7766121 | 1 | 10 | 0,2 % | -0,046 | 0,85 | 9,57E-01 | 1 | 10 | 0,3 % | 0,652 | 1,04 | 5,31E-01 | 1 | 9 | 0,2 % | -0,694 | 1,73 | 6,89E-01 |
| 19:7766659 | 0,00048 | ≤3 | 0,0 % | -0,136 | 1,25 | 9,14E-01 | 0,00050 | ≤3 | 0,0 % | -0,154 | 1,51 | 9,19E-01 | 0,00052 | ≤3 | 0,1 % | -0,145 | 2,54 | 9,54E-01 |
| 19:7767549 | 1 | ≤3 | 0,0 % | -0,118 | 2,81 | 9,67E-01 | 1 | ≤3 | 0,0 % | -0,024 | 3,37 | 9,94E-01 | 1 | ≤3 | 0,0 % | 0,040 | 5,69 | 9,94E-01 |
| 19:7768973 | 1 | 5 | 0,1 % | 0,865 | 1,25 | 4,89E-01 | 1 | 5 | 0,1 % | 2,530 | 1,58 | 1,10E-01 | 1 | 4 | 0,1 % | -0,686 | 3,04 | 8,22E-01 |
| 22:28695868 | 0,032 | 67 | 0,9 % | -0,944 | 0,34 | 5,30E-03 | 0,036 | 66 | 1,0 % | -0,717 | 0,43 | 9,52E-02 | 0,036 | 66 | 1,0 % | -0,812 | 0,75 | 2,77E-01 |
| 3:3844558 | 1 | 20 | 0,3 % | 0,053 | 0,63 | 9,32E-01 | 1 | 20 | 0,3 % | 0,547 | 0,79 | 4,91E-01 | 1 | 19 | 0,3 % | -0,296 | 1,40 | 8,33E-01 |
| 3:3845194 | - | ≤3 | 0,0 % | -0,204 | 0,28 | 4,59E-01 | - | ≤3 | 0,0 % | -0,121 | 0,21 | 5,65E-01 | - | ≤3 | 0,0 % | -0,032 | 0,11 | 7,63E-01 |
| 3:3845210 | 1 | 25 | 0,3 % | -0,288 | 0,56 | 6,08E-01 | 1 | 25 | 0,3 % | 0,136 | 0,71 | 8,48E-01 | 1 | 24 | 0,3 % | -0,428 | 1,25 | 7,31E-01 |
| 3:3845554 | 1 | ≤3 | 0,0 % | -0,657 | 2,80 | 8,15E-01 | 1 | ≤3 | 0,0 % | -0,513 | 3,36 | 8,79E-01 | 1 | ≤3 | 0,0 % | -0,687 | 5,68 | 9,04E-01 |
| 8:41672844 | 1 | 15 | 0,2 % | 0,500 | 0,72 | 4,89E-01 | 1 | 14 | 0,2 % | 0,640 | 0,95 | 4,99E-01 | 1 | 14 | 0,2 % | 3,679 | 1,63 | 2,40E-02 |
| 8:41686157 | 0,34 | 135 | 1,7 % | 0,149 | 0,24 | 5,37E-01 | 0,30 | 126 | 1,7 % | 0,108 | 0,32 | 7,33E-01 | 0,27 | 119 | 1,7 % | -0,732 | 0,56 | 1,90E-01 |
| 8:41688189 | 1 | 6 | 0,1 % | -1,317 | 1,14 | 2,48E-01 | 1 | 6 | 0,1 % | -1,209 | 1,45 | 4,03E-01 | 1 | 6 | 0,1 % | -1,121 | 2,49 | 6,52E-01 |
| 8:41688538 | 1 | 11 | 0,2 % | -0,367 | 0,84 | 6,63E-01 | 1 | 11 | 0,2 % | 0,450 | 1,07 | 6,74E-01 | 1 | 10 | 0,2 % | -1,000 | 1,93 | 6,04E-01 |
| 8:41690485 | 1 | 36 | 0,5 % | -0,140 | 0,47 | 7,65E-01 | 1 | 34 | 0,5 % | -0,258 | 0,61 | 6,72E-01 | 1 | 33 | 0,5 % | -0,642 | 1,06 | 5,46E-01 |
| 8:41690553 | 1 | 5 | 0,1 % | 3,033 | 1,32 | 2,11E-02 | 1 | 4 | 0,1 % | 3,737 | 1,87 | 4,62E-02 | 1 | 4 | 0,1 % | 15,089 | 3,25 | 3,37E-06 |
| 8:41692743 | 1 | 10 | 0,1 % | -0,170 | 0,88 | 8,48E-01 | 1 | 10 | 0,1 % | 0,749 | 1,12 | 5,04E-01 | 1 | 9 | 0,1 % | -0,935 | 2,03 | 6,45E-01 |
| 8:41694031 | 0,54 | 110 | 1,4 % | 0,220 | 0,27 | 4,15E-01 | 0,51 | 102 | 1,4 % | 0,214 | 0,36 | 5,47E-01 | 0,50 | 99 | 1,4 % | 1,065 | 0,62 | 8,65E-02 |
| 8:41695273 | 1 | 19 | 0,2 % | 0,213 | 0,64 | 7,41E-01 | 1 | 17 | 0,2 % | -0,647 | 0,86 | 4,52E-01 | 1 | 17 | 0,2 % | -0,551 | 1,48 | 7,10E-01 |
| 8:41708920 | 0,83 | 298 | 4,0 % | -0,023 | 0,17 | 8,92E-01 | 0,82 | 283 | 4,0 % | -0,045 | 0,22 | 8,33E-01 | 1 | 276 | 4,0 % | 0,234 | 0,38 | 5,33E-01 |
| 8:41714174 | 0,20 | 115 | 1,6 % | 0,129 | 0,26 | 6,24E-01 | 0,55 | 108 | 1,6 % | 0,091 | 0,35 | 7,93E-01 | 0,19 | 107 | 1,6 % | 1,567 | 0,59 | 8,37E-03 |
| 8:41715748 | 1 | 9 | 0,1 % | 0,263 | 1,01 | 7,96E-01 | 1 | 9 | 0,1 % | 1,418 | 1,29 | 2,70E-01 | 1 | 8 | 0,1 % | -0,653 | 2,37 | 7,83E-01 |
| 8:41715770 | 1 | 21 | 0,3 % | -0,607 | 0,61 | 3,21E-01 | 1 | 21 | 0,3 % | -0,080 | 0,78 | 9,18E-01 | 1 | 20 | 0,3 % | -0,762 | 1,37 | 5,77E-01 |
| 8:41716970 | 1 | 16 | 0,2 % | 0,904 | 0,70 | 1,97E-01 | 1 | 14 | 0,2 % | 0,515 | 0,95 | 5,87E-01 | 1 | 13 | 0,2 % | -0,535 | 1,69 | 7,52E-01 |
| 8:41724513 | 1 | 74 | 1,0 % | -0,183 | 0,33 | 5,77E-01 | 1 | 70 | 1,0 % | -0,315 | 0,43 | 4,61E-01 | 1 | 69 | 1,0 % | 0,149 | 0,74 | 8,41E-01 |

FinnGen GWAS replication (general population, no diabetic kidney disease adjustment)

| Position | Stroke MAC | MAF | Beta | SE | *p*-value | Stroke, without SAH MAC | MAF | Beta | SE | *p*-value | Ischemic stroke MAC | MAF | Beta | SE | *p*-value | Hemorrhagic stroke MAC | MAF | Beta | SE | *p*-value |
|---|---|---|---|---|---|---|---|---|---|---|---|---|---|---|---|---|---|---|---|---|
| 1:183623459 | 13867 | 2,74 % | -0,055 | 0,04 | 1,47E-01 | 13863 | 2,74 % | -0,055 | 0,04 | 1,61E-01 | 13855 | 2,74 % | -0,063 | 0,04 | 1,23E-01 | 13413 | 2,74 % | -0,078 | 0,08 | 3,04E-01 |
| 1:183647749 | 13921 | 2,75 % | -0,054 | 0,04 | 1,59E-01 | 13917 | 2,75 % | -0,056 | 0,04 | 1,54E-01 | 13909 | 2,75 % | -0,064 | 0,04 | 1,19E-01 | 13464 | 2,75 % | -0,074 | 0,08 | 3,30E-01 |
| 1:183647959 | 13928 | 2,75 % | -0,054 | 0,04 | 1,59E-01 | 13924 | 2,75 % | -0,056 | 0,04 | 1,54E-01 | 13916 | 2,75 % | -0,064 | 0,04 | 1,20E-01 | 13471 | 2,75 % | -0,075 | 0,08 | 3,22E-01 |
| 1:183648558 | 13929 | 2,75 % | -0,053 | 0,04 | 1,60E-01 | 13925 | 2,75 % | -0,056 | 0,04 | 1,55E-01 | 13917 | 2,75 % | -0,064 | 0,04 | 1,22E-01 | 13472 | 2,75 % | -0,075 | 0,08 | 3,21E-01 |
| 1:183648588 | 576 | 0,11 % | 0,222 | 0,20 | 2,60E-01 | 577 | 0,11 % | 0,210 | 0,20 | 3,05E-01 | 577 | 0,11 % | 0,329 | 0,21 | 1,24E-01 | 555 | 0,11 % | 0,046 | 0,39 | 9,05E-01 |
| 1:7849677 | 7082 | 1,40 % | 0,053 | 0,05 | 3,31E-01 | 7081 | 1,40 % | 0,063 | 0,06 | 2,61E-01 | 7078 | 1,40 % | 0,066 | 0,06 | 2,60E-01 | 6850 | 1,40 % | -0,091 | 0,11 | 4,06E-01 |
| 10:117257799 | 2062 | 0,41 % | 0,067 | 0,10 | 5,06E-01 | 2061 | 0,41 % | 0,114 | 0,10 | 2,74E-01 | 2059 | 0,41 % | 0,117 | 0,11 | 2,85E-01 | 1991 | 0,41 % | -0,074 | 0,20 | 7,11E-01 |
| 10:37602666 | 2480 | 0,49 % | -0,026 | 0,09 | 7,69E-01 | 2480 | 0,49 % | -0,036 | 0,09 | 6,94E-01 | 2479 | 0,49 % | -0,003 | 0,10 | 9,71E-01 | 2398 | 0,49 % | 0,023 | 0,18 | 8,97E-01 |
| 12:53481189 | 221 | 0,04 % | 0,247 | 0,28 | 3,81E-01 | 221 | 0,04 % | 0,349 | 0,29 | 2,29E-01 | 221 | 0,04 % | 0,496 | 0,30 | 1,00E-01 | 201 | 0,04 % | -0,683 | 0,62 | 2,70E-01 |
| 12:53481235 | 1392 | 0,28 % | -0,074 | 0,12 | 5,36E-01 | 1391 | 0,28 % | -0,064 | 0,12 | 6,01E-01 | 1388 | 0,27 % | -0,043 | 0,13 | 7,36E-01 | 1341 | 0,27 % | -0,126 | 0,24 | 6,01E-01 |
| 12:53486534 | 716 | 0,14 % | 0,158 | 0,17 | 3,63E-01 | 716 | 0,14 % | 0,092 | 0,18 | 6,10E-01 | 715 | 0,14 % | 0,221 | 0,19 | 2,38E-01 | 685 | 0,14 % | 0,017 | 0,36 | 9,61E-01 |
| 12:53486535 | 752 | 0,15 % | 0,173 | 0,17 | 3,08E-01 | 751 | 0,15 % | 0,114 | 0,18 | 5,15E-01 | 751 | 0,15 % | 0,215 | 0,18 | 2,42E-01 | 718 | 0,15 % | 0,086 | 0,35 | 8,04E-01 |
| 12:53503093 | 462 | 0,09 % | 0,094 | 0,23 | 6,81E-01 | 462 | 0,09 % | 0,159 | 0,24 | 5,01E-01 | 461 | 0,09 % | 0,202 | 0,25 | 4,18E-01 | 439 | 0,09 % | 0,086 | 0,45 | 8,49E-01 |
| 12:53504426 | 792 | 0,16 % | 0,190 | 0,17 | 2,56E-01 | 791 | 0,16 % | 0,134 | 0,17 | 4,36E-01 | 792 | 0,16 % | 0,233 | 0,18 | 1,95E-01 | 754 | 0,15 % | 0,094 | 0,34 | 7,83E-01 |
| 15:70667338 | 301 | 0,06 % | 0,094 | 0,27 | 7,27E-01 | 302 | 0,06 % | 0,124 | 0,28 | 6,57E-01 | 302 | 0,06 % | 0,191 | 0,29 | 5,12E-01 | 296 | 0,06 % | 0,024 | 0,53 | 9,64E-01 |
| 19:12896058 | 76 | 0,02 % | -0,751 | 0,54 | 1,64E-01 | 76 | 0,02 % | -0,702 | 0,56 | 2,09E-01 | 76 | 0,02 % | -0,624 | 0,59 | 2,86E-01 | 78 | 0,02 % | -0,080 | 1,03 | 9,38E-01 |
| 19:12897824 | 289 | 0,06 % | -0,020 | 0,26 | 9,41E-01 | 289 | 0,06 % | 0,006 | 0,27 | 9,83E-01 | 289 | 0,06 % | 0,084 | 0,29 | 7,68E-01 | 273 | 0,06 % | -0,609 | 0,54 | 2,59E-01 |
| 19:7765676 | 572 | 0,11 % | -0,071 | 0,18 | 7,00E-01 | 572 | 0,11 % | -0,213 | 0,19 | 2,58E-01 | 571 | 0,11 % | -0,254 | 0,20 | 1,97E-01 | 551 | 0,11 % | 0,577 | 0,37 | 1,18E-01 |
| 19:7766121 | *Genotyped | | | | | | | | | | | | | | | | | | | |
| 19:7766659 | *Genotyped | | | | | | | | | | | | | | | | | | | |
| 19:7767549 | *Genotyped | | | | | | | | | | | | | | | | | | | |
| 19:7768973 | 642 | 0,13 % | -0,046 | 0,18 | 8,00E-01 | 642 | 0,13 % | -0,094 | 0,19 | 6,14E-01 | 642 | 0,13 % | -0,010 | 0,19 | 9,59E-01 | 620 | 0,13 % | -0,307 | 0,35 | 3,83E-01 |
| 22:28695868 | 3185 | 0,63 % | 0,060 | 0,08 | 4,62E-01 | 3184 | 0,63 % | 0,073 | 0,08 | 3,83E-01 | 3181 | 0,63 % | 0,092 | 0,09 | 2,96E-01 | 3064 | 0,63 % | -0,045 | 0,16 | 7,86E-01 |
| 3:3844558 | 1483 | 0,29 % | -0,022 | 0,12 | 8,50E-01 | 1483 | 0,29 % | -0,033 | 0,12 | 7,90E-01 | 1482 | 0,29 % | -0,053 | 0,13 | 6,80E-01 | 1437 | 0,29 % | 0,078 | 0,23 | 7,38E-01 |
| 3:3845194 | **Genotyped: Linear regression without relatedness adjustment | | | | | | | | | | | | | | | | | | | |
| 3:3845210 | 1431 | 0,28 % | -0,066 | 0,12 | 5,67E-01 | 1431 | 0,28 % | -0,049 | 0,12 | 6,82E-01 | 1431 | 0,28 % | 0,006 | 0,12 | 9,64E-01 | 1380 | 0,28 % | -0,363 | 0,23 | 1,11E-01 |
| 3:3845554 | *Genotyped | | | | | | | | | | | | | | | | | | | |
| 3:3845966 | 281 | 0,06 % | -0,289 | 0,31 | 3,51E-01 | 282 | 0,06 % | -0,450 | 0,32 | 1,59E-01 | 282 | 0,06 % | -0,336 | 0,33 | 3,14E-01 | 274 | 0,06 % | 0,003 | 0,61 | 9,96E-01 |
| 8:41672844 | 716 | 0,14 % | 0,014 | 0,17 | 9,36E-01 | 716 | 0,14 % | 0,016 | 0,17 | 9,27E-01 | 716 | 0,14 % | -0,020 | 0,18 | 9,13E-01 | 691 | 0,14 % | 0,048 | 0,33 | 8,84E-01 |
| 8:41686157 | 8250 | 1,63 % | -0,068 | 0,05 | 1,73E-01 | 8251 | 1,63 % | -0,053 | 0,05 | 2,99E-01 | 8245 | 1,63 % | -0,063 | 0,05 | 2,43E-01 | 8011 | 1,64 % | -0,113 | 0,10 | 2,55E-01 |
| 8:41688189 | 257 | 0,05 % | 0,110 | 0,28 | 6,95E-01 | 257 | 0,05 % | 0,129 | 0,29 | 6,57E-01 | 257 | 0,05 % | 0,170 | 0,30 | 5,75E-01 | 243 | 0,05 % | -0,358 | 0,59 | 5,47E-01 |
| 8:41688538 | 917 | 0,18 % | -0,277 | 0,15 | 6,22E-02 | 916 | 0,18 % | -0,274 | 0,15 | 7,46E-02 | 915 | 0,18 % | -0,248 | 0,16 | 1,23E-01 | 898 | 0,18 % | -0,361 | 0,29 | 2,19E-01 |
| 8:41690485 | 2757 | 0,54 % | 0,031 | 0,09 | 7,20E-01 | 2756 | 0,54 % | 0,056 | 0,09 | 5,29E-01 | 2753 | 0,54 % | 0,042 | 0,09 | 6,55E-01 | 2675 | 0,55 % | -0,054 | 0,17 | 7,51E-01 |
| 8:41690553 | 684 | 0,14 % | -0,165 | 0,17 | 3,44E-01 | 684 | 0,14 % | -0,242 | 0,18 | 1,80E-01 | 684 | 0,14 % | -0,138 | 0,19 | 4,65E-01 | 666 | 0,14 % | -0,003 | 0,34 | 9,92E-01 |
| 8:41692743 | 1056 | 0,21 % | -0,165 | 0,14 | 2,28E-01 | 1056 | 0,21 % | -0,128 | 0,14 | 3,65E-01 | 1055 | 0,21 % | -0,089 | 0,15 | 5,45E-01 | 1028 | 0,21 % | -0,231 | 0,27 | 3,96E-01 |
| 8:41694031 | 6980 | 1,38 % | -0,086 | 0,05 | 1,11E-01 | 6977 | 1,38 % | -0,102 | 0,06 | 6,76E-02 | 6971 | 1,38 % | -0,105 | 0,06 | 7,31E-02 | 6785 | 1,39 % | -0,001 | 0,11 | 9,93E-01 |
| 8:41695273 | 1510 | 0,30 % | 0,005 | 0,12 | 9,65E-01 | 1510 | 0,30 % | 0,028 | 0,12 | 8,18E-01 | 1510 | 0,30 % | 0,057 | 0,13 | 6,54E-01 | 1466 | 0,30 % | -0,280 | 0,23 | 2,20E-01 |
| 8:41708920 | 20065 | 3,96 % | 0,004 | 0,03 | 9,13E-01 | 20060 | 3,96 % | 0,007 | 0,03 | 8,25E-01 | 20049 | 3,96 % | -0,001 | 0,03 | 9,82E-01 | 19374 | 3,96 % | 0,020 | 0,06 | 7,53E-01 |
| 8:41714174 | 8343 | 1,65 % | -0,013 | 0,05 | 7,92E-01 | 8338 | 1,65 % | -0,008 | 0,05 | 8,72E-01 | 8337 | 1,65 % | -0,016 | 0,05 | 7,66E-01 | 8059 | 1,65 % | -0,053 | 0,10 | 5,89E-01 |
| 8:41715748 | 1206 | 0,24 % | 0,174 | 0,13 | 1,77E-01 | 1205 | 0,24 % | 0,154 | 0,13 | 2,48E-01 | 1203 | 0,24 % | 0,240 | 0,14 | 8,57E-02 | 1139 | 0,23 % | -0,042 | 0,27 | 8,73E-01 |
| 8:41715770 | 2327 | 0,46 % | -0,034 | 0,09 | 7,15E-01 | 2327 | 0,46 % | -0,039 | 0,10 | 6,87E-01 | 2326 | 0,46 % | -0,060 | 0,10 | 5,56E-01 | 2252 | 0,46 % | 0,297 | 0,19 | 1,18E-01 |
| 8:41716970 | 1101 | 0,22 % | -0,071 | 0,14 | 5,99E-01 | 1101 | 0,22 % | -0,042 | 0,14 | 7,66E-01 | 1101 | 0,22 % | -0,007 | 0,15 | 9,63E-01 | 1056 | 0,22 % | -0,392 | 0,27 | 1,45E-01 |
| 8:41724513 | 3998 | 0,79 % | -0,013 | 0,07 | 8,51E-01 | 3998 | 0,79 % | -0,035 | 0,07 | 6,36E-01 | 3998 | 0,79 % | -0,049 | 0,08 | 5,19E-01 | 3847 | 0,79 % | 0,006 | 0,14 | 9,67E-01 |

**Supplementary Table 14.** SKAT-O gene-aggregate replication in T1D with minimal adjustment with GWAS and genotyping in FinnDiane

| | Gene | Phenotype | Variant N | Missingness rate (min) | Missingness rate (max) | MAF (min) | MAF (max) | Burden Score | *p*-value Burden | *p*-value SKAT | *p*-value SKAT-O | *p*-value SMMAT-E* |
|---|---|---|---|---|---|---|---|---|---|---|---|---|
| **PAV 1%** | *LRRN1* | Stroke | 4 | 0 % | 18 % | 0,02 % | 0,96 % | -101,65 | 0,138 | 0,519 | 0,215 | 0,388 |
| | | Ischemic | 3 | 0 % | 18 % | 0,02 % | 0,35 % | -2,65 | 0,935 | 0,998 | 1,000 | 0,996 |
| | | Hemorrhagic | 4 | 0 % | 18 % | 0,02 % | 0,98 % | -27,76 | 0,246 | 0,651 | 0,367 | 0,591 |
| | *FOXO1* | Stroke | 1 | 0 % | 0 % | 0,49 % | 0,49 % | 9,51 | 0,809 | 0,809 | 0,809 | 0,809 |
| | | Ischemic | 1 | 0 % | 0 % | 0,48 % | 0,48 % | -10,92 | 0,676 | 0,676 | 0,676 | 0,676 |
| | | Hemorrhagic | 1 | 0 % | 0 % | 0,51 % | 0,51 % | 34,23 | 0,017 | 0,017 | 0,017 | 0,017 |
| | *CLEC4M* | Stroke | 6 | 0 % | 43 % | 0,02 % | 0,22 % | 4,23 | 0,926 | 0,989 | 1,000 | 0,994 |
| | | Ischemic | 6 | 0 % | 43 % | 0,02 % | 0,23 % | 28,08 | 0,374 | 0,489 | 0,549 | 0,431 |
| | | Hemorrhagic | 6 | 0 % | 43 % | 0,02 % | 0,22 % | 12,88 | 0,422 | 0,128 | 0,212 | 0,227 |
| | *TARBP2* | Stroke | 2 | 0 % | 0 % | 0,10 % | 0,17 % | 48,45 | 0,120 | 0,054 | 0,079 | 0,013 |
| | | Ischemic | 2 | 0 % | 0 % | 0,08 % | 0,16 % | 5,07 | 0,807 | 0,302 | 0,411 | 0,251 |
| | | Hemorrhagic | 2 | 0 % | 0 % | 0,08 % | 0,18 % | 41,00 | 1,67E-04 | 2,35E-03 | 2,59E-04 | 1,08E-04 |
| | *RRP9* | Stroke | 3 | 0 % | 0 % | 0,08 % | 0,19 % | -14,77 | 0,685 | 0,662 | 0,810 | 0,731 |
| | | Ischemic | 3 | 0 % | 0 % | 0,07 % | 0,19 % | -29,72 | 0,241 | 0,598 | 0,350 | 0,583 |
| | | Hemorrhagic | 3 | 0 % | 0 % | 0,07 % | 0,19 % | -7,24 | 0,575 | 0,918 | 0,725 | 0,893 |
| | *GCDH* | Stroke | 3 | 0 % | 0 % | 0,04 % | 0,12 % | 30,39 | 0,305 | 0,707 | 0,439 | 0,499 |
| | | Ischemic | 3 | 0 % | 0 % | 0,03 % | 0,12 % | 29,34 | 0,164 | 0,289 | 0,248 | 0,290 |
| | | Hemorrhagic | 3 | 0 % | 0 % | 0,03 % | 0,11 % | -3,79 | 0,689 | 0,974 | 0,833 | 0,946 |
| | *SLC18A2* | Stroke | 2 | 0 % | 0 % | 0,01 % | 0,43 % | -53,56 | 0,149 | 0,160 | 0,166 | 0,424 |
| | | Ischemic | 2 | 0 % | 0 % | 0,01 % | 0,45 % | -16,87 | 0,511 | 0,538 | 0,586 | 0,833 |
| | | Hemorrhagic | 2 | 0 % | 0 % | 0,01 % | 0,46 % | -7,92 | 0,550 | 0,567 | 0,626 | 0,876 |
| **PAV 5%** | *UTS2* | Stroke | 1 | 0 % | 0 % | 1,52 % | 1,52 % | 66,48 | 0,235 | 0,235 | 0,235 | 0,235 |
| | | Ischemic | 1 | 0 % | 0 % | 1,50 % | 1,50 % | 35,80 | 0,354 | 0,354 | 0,354 | 0,354 |
| | | Hemorrhagic | 1 | 0 % | 0 % | 1,45 % | 1,45 % | -19,99 | 0,292 | 0,292 | 0,292 | 0,292 |
| | *APOBEC4* | Stroke | 4 | 0 % | 0 % | 0,13 % | 2,42 % | -90,90 | 0,583 | 0,558 | 0,625 | 0,700 |
| | | Ischemic | 4 | 0 % | 0 % | 0,11 % | 2,43 % | -82,68 | 0,465 | 0,533 | 0,521 | 0,581 |
| | | Hemorrhagic | 4 | 0 % | 0 % | 0,12 % | 2,49 % | 87,15 | 0,127 | 0,141 | 0,137 | 0,357 |
| | *ANK1* | Stroke | 15 | 0 % | 0 % | 0,06 % | 4,02 % | 106,50 | 0,437 | 0,876 | 0,642 | 0,736 |
| | | Ischemic | 15 | 0 % | 0 % | 0,05 % | 4,01 % | 52,00 | 0,580 | 0,969 | 0,786 | 0,874 |
| | | Hemorrhagic | 15 | 0 % | 0 % | 0,05 % | 4,06 % | 0,50 | 0,991 | 0,429 | 0,633 | 0,697 |
| | *SLC18A2* | Stroke | 2 | 0 % | 0 % | 0,01 % | 0,43 % | -53,56 | 0,149 | 0,160 | 0,166 | 0,424 |
| | | Ischemic | 2 | 0 % | 0 % | 0,01 % | 0,45 % | -16,87 | 0,511 | 0,538 | 0,586 | 0,833 |
| | | Hemorrhagic | 2 | 0 % | 0 % | 0,01 % | 0,46 % | -7,92 | 0,550 | 0,567 | 0,626 | 0,876 |
| | *TARBP2* | Stroke | 2 | 0 % | 0 % | 0,10 % | 0,17 % | 48,45 | 0,120 | 0,054 | 0,079 | 0,013 |
| | | Ischemic | 2 | 0 % | 0 % | 0,08 % | 0,16 % | 5,07 | 0,807 | 0,302 | 0,411 | 0,251 |
| | | Hemorrhagic | 2 | 0 % | 0 % | 0,08 % | 0,18 % | 41,00 | 1,67E-04 | 2,35E-04 | 2,59E-04 | 1,08E-04 |
| | *FOXO1* | Stroke | 1 | 0 % | 0 % | 0,49 % | 0,49 % | 9,51 | 0,809 | 0,809 | 0,809 | 0,809 |
| | | Ischemic | 1 | 0 % | 0 % | 0,48 % | 0,48 % | -10,92 | 0,676 | 0,676 | 0,676 | 0,676 |
| | | Hemorrhagic | 1 | 0 % | 0 % | 0,51 % | 0,51 % | 34,23 | 0,017 | 0,017 | 0,017 | 0,017 |
| | *CLEC4M* | Stroke | 6 | 0 % | 43 % | 0,02 % | 0,22 % | 4,23 | 0,926 | 0,989 | 1,000 | 0,994 |
| | | Ischemic | 6 | 0 % | 43 % | 0,02 % | 0,23 % | 28,08 | 0,374 | 0,489 | 0,549 | 0,431 |
| | | Hemorrhagic | 6 | 0 % | 43 % | 0,02 % | 0,22 % | 12,88 | 0,422 | 0,128 | 0,212 | 0,227 |
| | *GCDH* | Stroke | 3 | 0 % | 0 % | 0,04 % | 0,12 % | 30,39 | 0,305 | 0,707 | 0,439 | 0,499 |
| | | Ischemic | 3 | 0 % | 0 % | 0,03 % | 0,12 % | 29,34 | 0,164 | 0,289 | 0,248 | 0,290 |
| | | Hemorrhagic | 3 | 0 % | 0 % | 0,03 % | 0,11 % | -3,79 | 0,689 | 0,974 | 0,833 | 0,946 |
| **PTV 5%** | *ARPC5* | Stroke | 1 | 0 % | 0 % | 2,38 % | 2,38 % | -30,00 | 0,585 | 0,585 | 0,585 | 0,585 |
| | | Ischemic | 1 | 0 % | 0 % | 2,38 % | 2,38 % | -22,34 | 0,551 | 0,551 | 0,551 | 0,551 |
| | | Hemorrhagic | 1 | 0 % | 0 % | 2,44 % | 2,44 % | 29,01 | 0,125 | 0,125 | 0,125 | 0,125 |

*SMATT-E = variant-set mixed model association hybrid test

**SKAT-O gene-aggregate replication with low-frequency variants in T1D with GWAS and genotyping in FinnDiane:** MAF≤5% models for discovery cohort MAF≤1% genes, if low-frequency variants (MAF 1-5%) available for replication cohort

| | | | | | | | | | | | | |
|---|---|---|---|---|---|---|---|---|---|---|---|---|
| **PAV 5%** | *LRRN1* | Stroke | 6 | 0 % | 18 % | 0,02 % | 4,43 % | -62,45 | 0,499 | 0,492 | 0,680 | 0,506 |
| | | Ischemic | 6 | 0 % | 18 % | 0,02 % | 4,48 % | 104,24 | 0,104 | 0,081 | 0,123 | 0,075 |
| | | Hemorrhagic | 6 | 0 % | 18 % | 0,02 % | 4,30 % | -54,54 | 0,090 | 0,675 | 0,153 | 0,306 |
| | *RRP9* | Stroke | 4 | 0 % | 0 % | 0,08 % | 1,28 % | -14,05 | 0,824 | 0,916 | 1,000 | 0,916 |
| | | Ischemic | 4 | 0 % | 0 % | 0,07 % | 1,30 % | -14,20 | 0,746 | 0,780 | 0,861 | 0,659 |
| | | Hemorrhagic | 4 | 0 % | 0 % | 0,07 % | 1,30 % | 11,55 | 0,598 | 0,450 | 0,607 | 0,644 |

*SMATT-E = variant-set mixed model association hybrid test

**Supplementary Table 15.** SKAT-O gene-aggregate replication in T1D with additional DKD adjustment with GWAS and genotyping in FinnDiane

| | Gene | Phenotype | Variant N | Missingness rate (min) | Missingness rate (max) | MAF (min) | MAF (max) | Burden Score | *p*-value Burden | *p*-value SKAT | *p*-value SKAT-O | *p*-value SMMAT-E* |
|---|---|---|---|---|---|---|---|---|---|---|---|---|
| **PAV 1%** | *LRRN1* | Stroke | 4 | 0 % | 17 % | 0,02 % | 0,94 % | -55,00 | 0,354 | 0,820 | 0,503 | 0,661 |
| | | Ischemic | 4 | 0 % | 17 % | 0,02 % | 0,98 % | 30,83 | 0,467 | 0,770 | 0,632 | 0,789 |
| | | Hemorrhagic | 4 | 0 % | 17 % | 0,02 % | 0,96 % | -21,94 | 0,298 | 0,750 | 0,431 | 0,658 |
| | *CLEC4M* | Stroke | 6 | 0 % | 43 % | 0,02 % | 0,23 % | 9,60 | 0,818 | 0,972 | 1,000 | 0,966 |
| | | Ischemic | 6 | 0 % | 43 % | 0,02 % | 0,25 % | 31,56 | 0,283 | 0,323 | 0,428 | 0,208 |
| | | Hemorrhagic | 6 | 0 % | 43 % | 0,02 % | 0,23 % | 14,65 | 0,297 | 0,117 | 0,187 | 0,254 |
| | *GCDH* | Stroke | 3 | 0 % | 0 % | 0,04 % | 0,12 % | 26,50 | 0,374 | 0,689 | 0,516 | 0,447 |
| | | Ischemic | 3 | 0 % | 0 % | 0,03 % | 0,13 % | 25,78 | 0,250 | 0,428 | 0,362 | 0,429 |
| | | Hemorrhagic | 3 | 0 % | 0 % | 0,03 % | 0,12 % | -3,98 | 0,680 | 0,967 | 0,821 | 0,942 |
| | *SLC18A2* | Stroke | 2 | 0 % | 0 % | 0,01 % | 0,45 % | -53,43 | 0,122 | 0,125 | 0,129 | 0,377 |
| | | Ischemic | 2 | 0 % | 0 % | 0,01 % | 0,48 % | -17,69 | 0,480 | 0,485 | 0,521 | 0,824 |
| | | Hemorrhagic | 2 | 0 % | 0 % | 0,01 % | 0,48 % | -9,71 | 0,493 | 0,493 | 0,520 | 0,839 |
| | *MAP3K12* | Stroke | 4 | 0 % | 0 % | 0,04 % | 0,37 % | -29,25 | 0,588 | 0,642 | 0,736 | 0,650 |
| | | Ischemic | 4 | 0 % | 0 % | 0,04 % | 0,39 % | -18,79 | 0,630 | 0,881 | 0,775 | 0,806 |
| | | Hemorrhagic | 4 | 0 % | 0 % | 0,04 % | 0,38 % | 33,97 | 0,095 | 0,033 | 0,054 | 0,014 |
| | *TARBP2* | Stroke | 2 | 0 % | 0 % | 0,09 % | 0,16 % | 52,64 | 0,063 | 0,057 | 0,078 | 0,051 |
| | | Ischemic | 2 | 0 % | 0 % | 0,07 % | 0,15 % | 7,73 | 0,680 | 0,350 | 0,467 | 0,326 |
| | | Hemorrhagic | 2 | 0 % | 0 % | 0,07 % | 0,17 % | 42,17 | 1,05E-05 | 9,95E-05 | 1,66E-05 | 6,90E-05 |
| | *MTRNR2L7* | Stroke | 1 | 0 % | 0 % | 0,50 % | 0,50 % | -12,98 | 0,754 | 0,754 | 0,754 | 0,754 |
| | | Ischemic | 1 | 0 % | 0 % | 0,52 % | 0,52 % | 16,41 | 0,583 | 0,583 | 0,583 | 0,583 |
| | | Hemorrhagic | 1 | 0 % | 0 % | 0,49 % | 0,49 % | -11,14 | 0,466 | 0,466 | 0,466 | 0,466 |
| **PAV 5%** | *ANK1* | Stroke | 15 | 0 % | 0 % | 0,07 % | 3,96 % | 44,63 | 0,736 | 0,940 | 0,906 | 0,940 |
| | | Ischemic | 15 | 0 % | 0 % | 0,06 % | 3,97 % | 43,51 | 0,634 | 0,957 | 0,834 | 0,902 |
| | | Hemorrhagic | 15 | 0 % | 0 % | 0,06 % | 4,00 % | -5,56 | 0,903 | 0,573 | 0,782 | 0,807 |
| | *APOBEC4* | Stroke | 4 | 0 % | 0 % | 0,12 % | 2,48 % | -8,72 | 0,953 | 0,814 | 1,000 | 0,568 |
| | | Ischemic | 4 | 0 % | 0 % | 0,10 % | 2,49 % | -38,04 | 0,707 | 0,768 | 0,769 | 0,797 |
| | | Hemorrhagic | 4 | 0 % | 0 % | 0,11 % | 2,55 % | 96,82 | 0,058 | 0,069 | 0,061 | 0,136 |
| | *UTS2* | Stroke | 1 | 0 % | 0 % | 1,49 % | 1,49 % | 76,94 | 0,146 | 0,146 | 0,146 | 0,146 |
| | | Ischemic | 1 | 0 % | 0 % | 1,46 % | 1,46 % | 41,79 | 0,262 | 0,262 | 0,262 | 0,262 |
| | | Hemorrhagic | 1 | 0 % | 0 % | 1,41 % | 1,41 % | -18,22 | 0,323 | 0,323 | 0,323 | 0,323 |
| | *MTRNR2L7* | Stroke | 1 | 0 % | 0 % | 0,50 % | 0,50 % | -12,98 | 0,754 | 0,754 | 0,754 | 0,754 |
| | | Ischemic | 1 | 0 % | 0 % | 0,52 % | 0,52 % | 16,41 | 0,583 | 0,583 | 0,583 | 0,583 |
| | | Hemorrhagic | 1 | 0 % | 0 % | 0,49 % | 0,49 % | -11,14 | 0,466 | 0,466 | 0,466 | 0,466 |
| | *SLC18A2* | Stroke | 2 | 0 % | 0 % | 0,01 % | 0,45 % | -53,43 | 0,122 | 0,125 | 0,129 | 0,377 |
| | | Ischemic | 2 | 0 % | 0 % | 0,01 % | 0,48 % | -17,69 | 0,480 | 0,485 | 0,521 | 0,824 |
| | | Hemorrhagic | 2 | 0 % | 0 % | 0,01 % | 0,48 % | -9,71 | 0,493 | 0,493 | 0,520 | 0,839 |
| | *MAP3K12* | Stroke | 5 | 0 % | 0 % | 0,04 % | 2,05 % | -101,48 | 0,164 | 0,244 | 0,260 | 0,334 |
| | | Ischemic | 5 | 0 % | 0 % | 0,04 % | 2,11 % | -33,04 | 0,526 | 0,926 | 0,701 | 0,839 |
| | | Hemorrhagic | 5 | 0 % | 0 % | 0,04 % | 2,11 % | 19,73 | 0,459 | 0,118 | 0,192 | 0,079 |
| | *TARBP2* | Stroke | 2 | 0 % | 0 % | 0,09 % | 0,16 % | 52,64 | 0,063 | 0,057 | 0,078 | 0,051 |
| | | Ischemic | 2 | 0 % | 0 % | 0,07 % | 0,15 % | 7,73 | 0,680 | 0,350 | 0,467 | 0,326 |
| | | Hemorrhagic | 2 | 0 % | 0 % | 0,07 % | 0,17 % | 42,17 | 1,05E-05 | 9,95E-05 | 1,66E-05 | 6,90E-05 |
| | *CLEC4M* | Stroke | 6 | 0 % | 43 % | 0,02 % | 0,23 % | 9,60 | 0,818 | 0,972 | 1,000 | 0,966 |
| | | Ischemic | 6 | 0 % | 43 % | 0,02 % | 0,25 % | 31,56 | 0,283 | 0,323 | 0,428 | 0,208 |
| | | Hemorrhagic | 6 | 0 % | 43 % | 0,02 % | 0,23 % | 14,65 | 0,297 | 0,117 | 0,187 | 0,254 |
| | *GCDH* | Stroke | 3 | 0 % | 0 % | 0,04 % | 0,12 % | 26,50 | 0,374 | 0,689 | 0,516 | 0,447 |
| | | Ischemic | 3 | 0 % | 0 % | 0,03 % | 0,13 % | 25,78 | 0,250 | 0,428 | 0,362 | 0,429 |
| | | Hemorrhagic | 3 | 0 % | 0 % | 0,03 % | 0,12 % | -3,98 | 0,680 | 0,967 | 0,821 | 0,942 |
| **PTV 1%** | *UACA* | Stroke | 3 | 0 % | 17 % | 0,02 % | 0,09 % | -4,24 | 0,856 | 0,285 | 0,416 | 0,387 |
| | | Ischemic | 3 | 0 % | 16 % | 0,02 % | 0,09 % | 10,13 | 0,560 | 0,150 | 0,231 | 0,117 |
| | | Hemorrhagic | 2 | 0 % | 0 % | 0,06 % | 0,09 % | -2,95 | 0,721 | 0,917 | 0,829 | 0,953 |
| | *CHEK2* | Stroke | 3 | 0 % | 0 % | 0,09 % | 0,88 % | -82,88 | 0,170 | 0,030 | 0,046 | 0,038 |
| | | Ischemic | 3 | 0 % | 0 % | 0,09 % | 0,93 % | -38,66 | 0,374 | 0,331 | 0,453 | 0,372 |
| | | Hemorrhagic | 3 | 0 % | 0 % | 0,09 % | 0,95 % | -21,00 | 0,310 | 0,584 | 0,428 | 0,673 |
| **PTV 5%** | *ARPC5* | Stroke | 1 | 0 % | 0 % | 2,44 % | 2,44 % | -4,52 | 0,926 | 0,926 | 0,926 | 0,926 |
| | | Ischemic | 1 | 0 % | 0 % | 2,44 % | 2,44 % | -8,55 | 0,798 | 0,798 | 0,798 | 0,798 |
| | | Hemorrhagic | 1 | 0 % | 0 % | 2,50 % | 2,50 % | 32,03 | 0,057 | 0,057 | 0,057 | 0,057 |
| | *UACA* | Stroke | 3 | 0 % | 17 % | 0,02 % | 0,09 % | -4,24 | 0,856 | 0,285 | 0,416 | 0,387 |
| | | Ischemic | 3 | 0 % | 16 % | 0,02 % | 0,09 % | 10,13 | 0,560 | 0,150 | 0,231 | 0,117 |
| | | Hemorrhagic | 2 | 0 % | 0 % | 0,06 % | 0,09 % | -2,95 | 0,721 | 0,917 | 0,829 | 0,953 |
| | *CHEK2* | Stroke | 3 | 0 % | 0 % | 0,09 % | 0,88 % | -82,88 | 0,170 | 0,030 | 0,046 | 0,038 |
| | | Ischemic | 3 | 0 % | 0 % | 0,09 % | 0,93 % | -38,66 | 0,374 | 0,331 | 0,453 | 0,372 |
| | | Hemorrhagic | 3 | 0 % | 0 % | 0,09 % | 0,95 % | -21,00 | 0,310 | 0,584 | 0,428 | 0,673 |

*SMATT-E = variant-set mixed model association hybrid test

**SKAT-O gene-aggregate replication with low-frequency variants in T1D with GWAS and genotyping in FinnDiane**: MAF≤5% models for discovery cohort MAF≤1% genes, if low-frequency variants (MAF 1-5%) available for replication cohort

| | | | | | | | | | | | | |
|---|---|---|---|---|---|---|---|---|---|---|---|---|
| **PAV 5%** | *LRRN1* | Stroke | 6 | 0 % | 17 % | 0,02 % | 4,41 % | -3,51 | 0,966 | 0,546 | 0,734 | 0,750 |
| | | Ischemic | 6 | 0 % | 17 % | 0,02 % | 4,46 % | 127,38 | 0,030 | 0,037 | 0,039 | 0,028 |
| | | Hemorrhagic | 6 | 0 % | 17 % | 0,02 % | 4,28 % | -45,32 | 0,122 | 0,765 | 0,204 | 0,379 |
| | *MAP3K12* | Stroke | 5 | 0 % | 0 % | 0,04 % | 2,05 % | -101,48 | 0,164 | 0,244 | 0,260 | 0,334 |
| | | Ischemic | 5 | 0 % | 0 % | 0,04 % | 2,11 % | -33,04 | 0,526 | 0,926 | 0,701 | 0,839 |
| | | Hemorrhagic | 5 | 0 % | 0 % | 0,04 % | 2,11 % | 19,74 | 0,459 | 0,118 | 0,192 | 0,079 |

*SMATT-E = variant-set mixed model association hybrid test

**Supplementary Table 16.** Jurgens et al. UK Biobank gene burden replication (N=200,337)

| Phenotype | Gene | Variants N | Carriers N | Score | Score statistic | p-value |
|---|---|---|---|---|---|---|
| Ischemic stroke | *HAS1* | 20 | 388 | 1,549 | 0,742 | 0,458 |
| | *UACA* | 77 | 295 | -1,202 | -0,678 | 0,498 |
| | *CHEK2* | 37 | 288 | 2,825 | 1,601 | 0,109 |
| Stroke | *HAS1* | 20 | 388 | 7,624 | 2,535 | 0,012 |
| | *UACA* | 77 | 295 | 0,063 | 0,024 | 0,981 |
| | *CHEK2* | 37 | 288 | 3,289 | 1,293 | 0,196 |

Gene burden replication in the general population UK Biobank study with ultra-rare loss-of-function variants (MAF≤0.1%)

Logistic mixed-effects model with saddle-point approximation adjusted for age, sex, and significant ancestral principal components, with a sparse kinship matrix as a random effect

We attempted to replicate genes identified with protein truncating variants (PTV): *ARPC5* , *UACA* , *CHEK2* and *HAS1*

**Supplementary Table 17:** Backman et al. UK Biobank gene burden replication (N=454,787)

| Model | Gene | Model | Trait | OR | CI Lower | CI Upper | p-value |
|---|---|---|---|---|---|---|---|
| **Stroke - PAV** | *UTS2* | 0.001% | Stroke | 1,570 | 0,291 | 8,475 | 0,600 |
| | *UTS2* | 0.01% | Stroke | 0,921 | 0,306 | 2,769 | 0,884 |
| | *UTS2* | 0.1% | Stroke | 1,598 | 0,646 | 3,956 | 0,311 |
| | *UTS2* | 1 % | Stroke | 1,598 | 0,646 | 3,956 | 0,311 |
| | *UTS2* | singleton | Stroke | 10,228 | 1,051 | 99,548 | 0,045 |
| **Stroke - PAV** | *APOBEC4* | 0.001% | Stroke | 0,360 | 0,049 | 2,628 | 0,313 |
| | *APOBEC4* | 0.01% | Stroke | 1,004 | 0,319 | 3,163 | 0,995 |
| | *APOBEC4* | 0.1% | Stroke | 0,917 | 0,422 | 1,993 | 0,827 |
| | *APOBEC4* | 1 % | Stroke | 0,917 | 0,422 | 1,993 | 0,827 |
| | *APOBEC4* | singleton | Stroke | 0,364 | 0,002 | 55,661 | 0,693 |
| **Stroke - PAV** | *LRRN1* | 0.001% | Stroke | 2,449 | 1,116 | 5,375 | 0,026 |
| | *LRRN1* | 0.01% | Stroke | 1,571 | 0,739 | 3,342 | 0,240 |
| | *LRRN1* | 0.1% | Stroke | 1,298 | 0,886 | 1,903 | 0,181 |
| | *LRRN1* | 1 % | Stroke | 1,298 | 0,886 | 1,903 | 0,181 |
| | *LRRN1* | singleton | Stroke | 3,732 | 1,009 | 13,806 | 0,049 |
| **Stroke - PAV** | *RRP9* | 0.001% | Stroke | 2,092 | 0,833 | 5,257 | 0,116 |
| | *RRP9* | 0.01% | Stroke | 1,388 | 0,736 | 2,616 | 0,311 |
| | *RRP9* | 0.1% | Stroke | 1,388 | 0,736 | 2,616 | 0,311 |
| | *RRP9* | 1 % | Stroke | 1,025 | 0,810 | 1,297 | 0,837 |
| | *RRP9* | singleton | Stroke | 6,084 | 0,591 | 62,648 | 0,129 |
| **Stroke - PAV** | *ANK1* | 0.001% | Stroke | 1,127 | 0,655 | 1,940 | 0,666 |
| | *ANK1* | 0.01% | Stroke | 1,046 | 0,761 | 1,438 | 0,782 |
| | *ANK1* | 0.1% | Stroke | 0,893 | 0,719 | 1,109 | 0,305 |
| | *ANK1* | 1 % | Stroke | 0,919 | 0,770 | 1,097 | 0,348 |
| | *ANK1* | singleton | Stroke | 1,483 | 0,521 | 4,220 | 0,461 |
| **Stroke - PAV** | *SLC18A2* | 0.001% | Stroke | 0,482 | 0,165 | 1,408 | 0,182 |
| | *SLC18A2* | 0.01% | Stroke | 0,919 | 0,469 | 1,804 | 0,807 |
| | *SLC18A2* | 0.1% | Stroke | 0,736 | 0,445 | 1,219 | 0,234 |
| | *SLC18A2* | 1 % | Stroke | 0,736 | 0,445 | 1,219 | 0,234 |
| | *SLC18A2* | singleton | Stroke | 0,358 | 0,043 | 2,952 | 0,340 |
| **Stroke - PAV** | *MAP3K12* | 0.001% | Stroke | 1,269 | 0,473 | 3,404 | 0,637 |
| | *MAP3K12* | 0.01% | Stroke | 1,854 | 0,995 | 3,455 | 0,052 |
| | *MAP3K12* | 0.1% | Stroke | 1,659 | 1,029 | 2,674 | 0,038 |
| | *MAP3K12* | 1 % | Stroke | 1,659 | 1,029 | 2,674 | 0,038 |
| | *MAP3K12* | singleton | Stroke | 4,726 | 1,437 | 15,541 | 0,011 |
| **Stroke - PAV** | *TARBP2* | 0.001% | Stroke | 2,664 | 0,770 | 9,220 | 0,122 |
| | *TARBP2* | 0.01% | Stroke | 1,761 | 0,772 | 4,020 | 0,179 |
| | *TARBP2* | 0.1% | Stroke | 1,287 | 0,678 | 2,443 | 0,440 |
| | *TARBP2* | 1 % | Stroke | 1,287 | 0,678 | 2,443 | 0,440 |
| | *TARBP2* | singleton | Stroke | 0,358 | 0,035 | 3,676 | 0,387 |
| **Stroke - PAV** | *FOXO1* | 0.001% | Stroke | 4,606 | 0,962 | 22,051 | 0,056 |
| | *FOXO1* | 0.01% | Stroke | 2,127 | 0,985 | 4,592 | 0,055 |
| | *FOXO1* | 0.1% | Stroke | 2,127 | 0,985 | 4,592 | 0,055 |
| | *FOXO1* | 1 % | Stroke | 2,127 | 0,985 | 4,592 | 0,055 |
| | *FOXO1* | singleton | Stroke | 5,366 | 1,366 | 21,084 | 0,016 |
| **Stroke - PAV** | *CLEC4M* | 0.001% | Stroke | 3,493 | 0,432 | 28,260 | 0,241 |
| | *CLEC4M* | 0.01% | Stroke | 0,896 | 0,238 | 3,377 | 0,871 |
| | *CLEC4M* | 0.1% | Stroke | 0,774 | 0,328 | 1,825 | 0,558 |
| | *CLEC4M* | 1 % | Stroke | 1,006 | 0,806 | 1,255 | 0,958 |
| | *CLEC4M* | singleton | Stroke | 17,024 | 0,362 | 801,076 | 0,149 |
| **Stroke - PAV** | *GCDH* | 0.001% | Stroke | 2,044 | 0,818 | 5,109 | 0,126 |
| | *GCDH* | 0.01% | Stroke | 1,542 | 0,919 | 2,589 | 0,101 |
| | *GCDH* | 0.1% | Stroke | 1,451 | 0,985 | 2,138 | 0,059 |
| | *GCDH* | 1 % | Stroke | 1,451 | 0,985 | 2,138 | 0,059 |
| | *GCDH* | singleton | Stroke | 1,778 | 0,307 | 10,283 | 0,520 |
| **Ischemic - PAV** | *UTS2* | 0.001% | Ischemic | 0,760 | 0,138 | 4,198 | 0,753 |
| | *UTS2* | 0.01% | Ischemic | 0,664 | 0,221 | 1,997 | 0,466 |
| | *UTS2* | 0.1% | Ischemic | 0,811 | 0,331 | 1,983 | 0,646 |
| | *UTS2* | 1 % | Ischemic | 0,811 | 0,331 | 1,983 | 0,646 |
| | *UTS2* | singleton | Ischemic | 28,785 | 0,468 | 1772,120 | 0,110 |
| **Ischemic - PAV** | *APOBEC4* | 0.001% | Ischemic | 0,354 | 0,053 | 2,378 | 0,285 |
| | *APOBEC4* | 0.01% | Ischemic | 0,916 | 0,304 | 2,755 | 0,876 |
| | *APOBEC4* | 0.1% | Ischemic | 1,290 | 0,614 | 2,711 | 0,501 |
| | *APOBEC4* | 1 % | Ischemic | 1,290 | 0,614 | 2,711 | 0,501 |
| | *APOBEC4* | singleton | Ischemic | 0,362 | 0,003 | 49,128 | 0,685 |
| **Ischemic - PAV** | *LRRN1* | 0.001% | Ischemic | 1,990 | 0,696 | 5,691 | 0,199 |
| | *LRRN1* | 0.01% | Ischemic | 1,762 | 0,833 | 3,725 | 0,138 |
| | *LRRN1* | 0.1% | Ischemic | 1,377 | 0,940 | 2,016 | 0,100 |
| | *LRRN1* | 1 % | Ischemic | 1,377 | 0,940 | 2,016 | 0,100 |
| | *LRRN1* | singleton | Ischemic | 2,398 | 0,353 | 16,304 | 0,371 |
| **Ischemic - PAV** | *RRP9* | 0.001% | Ischemic | 1,260 | 0,511 | 3,105 | 0,616 |
| | *RRP9* | 0.01% | Ischemic | 0,805 | 0,429 | 1,509 | 0,498 |
| | *RRP9* | 0.1% | Ischemic | 0,805 | 0,429 | 1,509 | 0,498 |
| | *RRP9* | 1 % | Ischemic | 0,895 | 0,709 | 1,131 | 0,354 |
| | *RRP9* | singleton | Ischemic | 1,599 | 0,145 | 17,622 | 0,702 |

| | | | | | | | |
|---|---|---|---|---|---|---|---|
| **Ischemic - PAV** | *ANK1* | 0.001% | Ischemic | 1,154 | 0,665 | 2,002 | 0,610 |
| | *ANK1* | 0.01% | Ischemic | 1,042 | 0,760 | 1,428 | 0,799 |
| | *ANK1* | 0.1% | Ischemic | 0,950 | 0,767 | 1,176 | 0,637 |
| | *ANK1* | 1 % | Ischemic | 1,024 | 0,861 | 1,219 | 0,788 |
| | *ANK1* | singleton | Ischemic | 1,357 | 0,490 | 3,757 | 0,557 |
| **Ischemic - PAV** | *SLC18A2* | 0.001% | Ischemic | 0,621 | 0,219 | 1,759 | 0,370 |
| | *SLC18A2* | 0.01% | Ischemic | 1,119 | 0,575 | 2,178 | 0,741 |
| | *SLC18A2* | 0.1% | Ischemic | 0,866 | 0,527 | 1,423 | 0,571 |
| | *SLC18A2* | 1 % | Ischemic | 0,866 | 0,527 | 1,423 | 0,571 |
| | *SLC18A2* | singleton | Ischemic | 1,167 | 0,138 | 9,873 | 0,887 |
| **Ischemic - PAV** | *MAP3K12* | 0.001% | Ischemic | 2,150 | 0,987 | 4,684 | 0,054 |
| | *MAP3K12* | 0.01% | Ischemic | 2,270 | 1,269 | 4,063 | 0,006 |
| | *MAP3K12* | 0.1% | Ischemic | 1,558 | 0,900 | 2,696 | 0,113 |
| | *MAP3K12* | 1 % | Ischemic | 1,558 | 0,900 | 2,696 | 0,113 |
| | *MAP3K12* | singleton | Ischemic | 3,942 | 1,051 | 14,788 | 0,042 |
| **Ischemic - PAV** | *TARBP2* | 0.001% | Ischemic | 1,712 | 0,501 | 5,855 | 0,391 |
| | *TARBP2* | 0.01% | Ischemic | 1,563 | 0,705 | 3,467 | 0,272 |
| | *TARBP2* | 0.1% | Ischemic | 0,962 | 0,517 | 1,788 | 0,902 |
| | *TARBP2* | 1 % | Ischemic | 0,962 | 0,517 | 1,788 | 0,902 |
| | *TARBP2* | singleton | Ischemic | 0,356 | 0,033 | 3,869 | 0,396 |
| **Ischemic - PAV** | *FOXO1* | 0.001% | Ischemic | 1,544 | 0,288 | 8,269 | 0,612 |
| | *FOXO1* | 0.01% | Ischemic | 1,338 | 0,487 | 3,673 | 0,572 |
| | *FOXO1* | 0.1% | Ischemic | 1,338 | 0,487 | 3,673 | 0,572 |
| | *FOXO1* | 1 % | Ischemic | 1,338 | 0,487 | 3,673 | 0,572 |
| | *FOXO1* | singleton | Ischemic | 3,160 | 0,176 | 56,752 | 0,435 |
| **Ischemic - PAV** | *CLEC4M* | 0.001% | Ischemic | 4,469 | 0,492 | 40,574 | 0,183 |
| | *CLEC4M* | 0.01% | Ischemic | 0,887 | 0,236 | 3,341 | 0,859 |
| | *CLEC4M* | 0.1% | Ischemic | 0,667 | 0,273 | 1,628 | 0,373 |
| | *CLEC4M* | 1 % | Ischemic | 1,037 | 0,834 | 1,289 | 0,745 |
| | *CLEC4M* | singleton | Ischemic | 30,255 | 0,486 | 1884,860 | 0,106 |
| **Ischemic - PAV** | *GCDH* | 0.001% | Ischemic | 1,601 | 0,644 | 3,977 | 0,311 |
| | *GCDH* | 0.01% | Ischemic | 0,796 | 0,478 | 1,326 | 0,380 |
| | *GCDH* | 0.1% | Ischemic | 1,149 | 0,784 | 1,684 | 0,477 |
| | *GCDH* | 1 % | Ischemic | 1,149 | 0,784 | 1,684 | 0,477 |
| | *GCDH* | singleton | Ischemic | 1,627 | 0,293 | 9,049 | 0,578 |
| **PTV - Stroke** | *ARPC5* | 0.001% | Stroke | 0,356 | 0,006 | 20,694 | 0,618 |
| | *ARPC5* | 0.01% | Stroke | 1,860 | 0,314 | 11,021 | 0,494 |
| | *ARPC5* | 0.1% | Stroke | 1,860 | 0,314 | 11,021 | 0,494 |
| | *ARPC5* | 1 % | Stroke | 1,860 | 0,314 | 11,021 | 0,494 |
| **PTV - Stroke** | *UACA* | 0.001% | Stroke | 2,539 | 1,091 | 5,905 | 0,031 |
| | *UACA* | 0.01% | Stroke | 1,865 | 1,046 | 3,326 | 0,035 |
| | *UACA* | 0.1% | Stroke | 1,683 | 0,876 | 3,232 | 0,118 |
| | *UACA* | 1 % | Stroke | 1,683 | 0,876 | 3,232 | 0,118 |
| | *UACA* | singleton | Stroke | 3,809 | 1,035 | 14,020 | 0,044 |
| **PTV - Stroke** | *HAS1* | 0.001% | Stroke | 3,807 | 0,843 | 17,194 | 0,082 |
| | *HAS1* | 0.01% | Stroke | 3,430 | 0,918 | 12,815 | 0,067 |
| | *HAS1* | 0.1% | Stroke | 1,551 | 1,032 | 2,333 | 0,035 |
| | *HAS1* | 1 % | Stroke | 1,551 | 1,032 | 2,333 | 0,035 |
| | *HAS1* | singleton | Stroke | 0,358 | 0,008 | 15,516 | 0,593 |
| **PTV - Stroke** | *CHEK2* | 0.001% | Stroke | 0,823 | 0,232 | 2,921 | 0,764 |
| | *CHEK2* | 0.01% | Stroke | 0,804 | 0,365 | 1,771 | 0,588 |
| | *CHEK2* | 0.1% | Stroke | 0,907 | 0,499 | 1,649 | 0,749 |
| | *CHEK2* | 1 % | Stroke | 1,098 | 0,797 | 1,513 | 0,568 |
| | *CHEK2* | singleton | Stroke | 3,607 | 0,438 | 29,704 | 0,233 |
| **PTV - Ischemic** | *ARPC5* | 0.001% | Ischemic | 0,357 | 0,006 | 22,473 | 0,626 |
| | *ARPC5* | 0.01% | Ischemic | 0,997 | 0,137 | 7,262 | 0,997 |
| | *ARPC5* | 0.1% | Ischemic | 0,997 | 0,137 | 7,262 | 0,997 |
| | *ARPC5* | 1 % | Ischemic | 0,997 | 0,137 | 7,262 | 0,997 |
| **PTV - Ischemic** | *UACA* | 0.001% | Ischemic | 0,514 | 0,156 | 1,697 | 0,275 |
| | *UACA* | 0.01% | Ischemic | 1,001 | 0,493 | 2,030 | 0,998 |
| | *UACA* | 0.1% | Ischemic | 0,944 | 0,494 | 1,805 | 0,862 |
| | *UACA* | 1 % | Ischemic | 0,944 | 0,494 | 1,805 | 0,862 |
| | *UACA* | singleton | Ischemic | 0,357 | 0,042 | 3,010 | 0,344 |
| **PTV - Ischemic** | *HAS1* | 0.001% | Ischemic | 0,683 | 0,140 | 3,316 | 0,636 |
| | *HAS1* | 0.01% | Ischemic | 0,918 | 0,238 | 3,534 | 0,901 |
| | *HAS1* | 0.1% | Ischemic | 1,011 | 0,630 | 1,623 | 0,964 |
| | *HAS1* | 1 % | Ischemic | 1,011 | 0,630 | 1,623 | 0,964 |
| | *HAS1* | singleton | Ischemic | 0,352 | 0,010 | 12,257 | 0,564 |
| **PTV - Ischemic** | *CHEK2* | 0.001% | Ischemic | 1,035 | 0,319 | 3,355 | 0,954 |
| | *CHEK2* | 0.01% | Ischemic | 1,092 | 0,523 | 2,283 | 0,814 |
| | *CHEK2* | 0.1% | Ischemic | 1,267 | 0,715 | 2,246 | 0,418 |
| | *CHEK2* | 0,01 | Ischemic | 1,088 | 0,798 | 1,484 | 0,595 |
| | *CHEK2* | singleton | Ischemic | 0,922 | 0,135 | 6,296 | 0,934 |

**Gene burden replication in the general population UK Biobank study with putative LoF variants (M1) and deleterious missense variants (M3).**

1. Genes which we discovered with PAVs (replicated with M3)
2. Genes which we discovered with PTVs (replicated with M1)

**Supplementary Table 18:** Significant ($p<5\times10^{-8}$) and suggestively significant ($p<5\times10^{-7}$) genomic windows (STAAR-O) with replication in T1D

| | | | | | Discovery Stroke (WGS: T1D specific) | | | Discovery Ischemic stroke (WGS: T1D specific) | | | Discovery Hemorrhagic stroke (WGS: T1D specific) | | | Replication GWAS Stroke (T1D specific) | | | Replication GWAS Ischemic stroke (T1D specific) | | | Replication GWAS Hemorrhagic stroke (T1D specific) | | |
|---|---|---|---|---|---|---|---|---|---|---|---|---|---|---|---|---|---|---|---|---|---|---|
| **MODEL** | **CHR** | **START** | **END** | **Closest gene** | **N** | **CMAC** | ***p*-value** | **N** | **CMAC** | ***p*-value** | **N** | **CMAC** | ***p*-value** | **N** | **CMAC** | ***p*-value** | **N** | **CMAC** | ***p*-value** | **N** | **CMAC** | ***p*-value** |
| **MAF<5%** | 13 | 85912001 | 85916000 | *MOB1AP1, SLITRK6* | 21 | 142 | 1,79E-07 | 17 | 110 | 1,80E-03 | 16 | 106 | 4,81E-09 | 12 | 737 | 0,188 | 12 | 686 | 0,670 | 12 | 665 | 0,710 |
| | 4 | 170752001 | 170756000 | *LINC02382* | 24 | 328 | 8,88E-08 | 19 | 272 | 2,64E-07 | 20 | 232 | 1,00E-03 | 14 | 1726 | 0,453 | 14 | 1639 | 0,813 | 14 | 1594 | 0,829 |
| | 4 | 170754001 | 170758000 | *LINC02382* | 19 | 239 | 8,55E-08 | 15 | 203 | 1,62E-07 | 15 | 156 | 1,75E-01 | 10 | 778 | 0,703 | 10 | 742 | 0,974 | 10 | 719 | 0,744 |
| | 4 | 170756001 | 170760000 | *LINC02382* | 20 | 152 | 7,30E-09 | 15 | 128 | 2,53E-09 | 12 | 95 | 1,39E-01 | 11 | 589 | 0,945 | 11 | 557 | 0,956 | 11 | 540 | 0,529 |
| | 4 | 170758001 | 170762000 | *LINC02382* | 30 | 302 | 9,42E-09 | 23 | 254 | 2,11E-08 | 20 | 195 | 5,50E-02 | 18 | 1725 | 0,594 | 18 | 1651 | 0,916 | 18 | 1601 | 0,845 |
| | 4 | 170760001 | 170764000 | *LINC02382* | 33 | 353 | 2,50E-07 | 29 | 303 | 3,06E-07 | 27 | 238 | 1,55E-01 | 22 | 2117 | 0,235 | 22 | 2038 | 0,760 | 22 | 1971 | 0,859 |
| | 4 | 170766001 | 170770000 | *LINC02382* | 21 | 284 | 2,61E-07 | 21 | 248 | 8,49E-07 | 18 | 226 | 2,30E-02 | 14 | 1581 | 0,940 | 14 | 1527 | 0,180 | 14 | 1457 | 0,863 |
| | 4 | 170768001 | 170772000 | *LINC02382* | 20 | 264 | 3,08E-08 | 19 | 226 | 5,43E-08 | 18 | 179 | 3,90E-02 | 11 | 718 | 0,979 | 11 | 690 | 0,644 | 11 | 660 | 0,431 |
| | 4 | 170770001 | 170774000 | *LINC02382* | 22 | 265 | 1,66E-07 | 21 | 227 | 4,36E-07 | 21 | 194 | 8,00E-03 | 11 | 700 | 0,972 | 11 | 672 | 0,457 | 11 | 642 | 0,484 |
| | 4 | 170776001 | 170780000 | *LINC02382* | 30 | 497 | 5,40E-08 | 28 | 426 | 1,64E-07 | 28 | 354 | 2,80E-02 | 17 | 1813 | 0,315 | 17 | 1756 | 0,542 | 17 | 1684 | 0,722 |
| | 4 | 170778001 | 170782000 | *LINC02382* | 26 | 547 | 7,44E-08 | 24 | 469 | 1,79E-07 | 24 | 382 | 3,70E-02 | 13 | 2195 | 0,701 | 13 | 2118 | 0,748 | 13 | 2036 | 0,938 |
| | 4 | 170780001 | 170784000 | *LINC02382* | 29 | 520 | 7,20E-08 | 28 | 446 | 1,82E-07 | 26 | 357 | 3,40E-02 | 16 | 2424 | 0,587 | 16 | 2322 | 0,744 | 16 | 2240 | 0,908 |
| | 4 | 170782001 | 170786000 | *LINC02382* | 58 | 934 | 3,40E-08 | 54 | 797 | 1,29E-07 | 52 | 662 | 2,00E-03 | 22 | 2382 | 0,040 | 22 | 2281 | 0,244 | 22 | 2187 | 0,791 |
| | 4 | 170784001 | 170788000 | *LINC02382* | 65 | 1190 | 1,10E-08 | 61 | 1008 | 1,24E-08 | 62 | 886 | 3,00E-03 | 22 | 2934 | 0,043 | 22 | 2822 | 0,176 | 22 | 2707 | 0,808 |
| | 4 | 170786001 | 170790000 | *LINC02382* | 43 | 931 | 2,88E-09 | 41 | 783 | 9,77E-09 | 42 | 677 | 5,00E-03 | 20 | 3177 | 0,831 | 20 | 3052 | 0,441 | 20 | 2945 | 0,765 |
| | 4 | 170788001 | 170792000 | *LINC02382* | 34 | 667 | 1,90E-09 | 31 | 564 | 3,95E-08 | 31 | 448 | 8,00E-03 | 22 | 2717 | 0,916 | 22 | 2608 | 0,656 | 22 | 2521 | 0,852 |
| | 4 | 170790001 | 170794000 | *LINC02382* | 33 | 538 | 8,78E-08 | 31 | 460 | 1,67E-07 | 30 | 361 | 5,00E-02 | 18 | 2564 | 0,847 | 18 | 2464 | 0,799 | 18 | 2379 | 0,982 |
| | 4 | 170792001 | 170796000 | *LINC02382* | 31 | 445 | 1,33E-07 | 29 | 384 | 1,69E-07 | 28 | 303 | 6,90E-02 | 17 | 2228 | 0,876 | 17 | 2136 | 0,743 | 17 | 2059 | 0,927 |
| | 4 | 170794001 | 170798000 | *LINC02382* | 25 | 424 | 2,47E-08 | 22 | 353 | 9,47E-08 | 23 | 278 | 2,00E-02 | 14 | 986 | 0,881 | 14 | 939 | 0,673 | 14 | 906 | 0,956 |
| | 4 | 170796001 | 170800000 | *LINC02382* | 23 | 415 | 2,24E-08 | 20 | 343 | 1,12E-07 | 22 | 276 | 8,00E-03 | 12 | 871 | 0,896 | 12 | 828 | 0,838 | 12 | 797 | 0,671 |
| | 4 | 170798001 | 170802000 | *LINC02382* | 24 | 646 | 2,29E-08 | 23 | 541 | 1,06E-07 | 24 | 433 | 9,00E-03 | 13 | 1699 | 0,907 | 13 | 1633 | 0,352 | 13 | 1565 | 0,877 |
| | 4 | 170800001 | 170804000 | *LINC02382* | 25 | 562 | 3,40E-08 | 24 | 470 | 1,46E-07 | 24 | 377 | 1,60E-02 | 12 | 1952 | 0,753 | 12 | 1879 | 0,519 | 12 | 1805 | 0,909 |
| | 4 | 170806001 | 170810000 | *LINC02382* | 29 | 538 | 3,33E-07 | 28 | 461 | 2,63E-07 | 28 | 383 | 1,60E-01 | 17 | 2394 | 0,962 | 17 | 2295 | 0,917 | 17 | 2210 | 0,937 |
| | 4 | 170808001 | 170812000 | *LINC02382* | 36 | 675 | 7,28E-08 | 34 | 575 | 1,28E-07 | 34 | 465 | 8,60E-02 | 15 | 2380 | 0,929 | 15 | 2279 | 0,819 | 15 | 2201 | 0,980 |
| | 4 | 170812001 | 170816000 | *LINC02382* | 21 | 423 | 2,06E-07 | 19 | 361 | 2,29E-07 | 19 | 299 | 1,20E-01 | 13 | 1664 | 0,840 | 13 | 1601 | 0,698 | 13 | 1537 | 0,853 |
| | 4 | 170814001 | 170818000 | *LINC02382* | 25 | 469 | 2,09E-07 | 23 | 404 | 1,76E-07 | 23 | 314 | 2,08E-01 | 17 | 2196 | 0,577 | 17 | 2108 | 0,853 | 17 | 2040 | 0,903 |
| | 4 | 170816001 | 170820000 | *LINC02382* | 36 | 505 | 2,13E-08 | 35 | 429 | 6,57E-08 | 31 | 337 | 8,20E-02 | 25 | 3055 | 0,853 | 25 | 2914 | 0,564 | 25 | 2836 | 0,873 |
| | 4 | 170818001 | 170822000 | *LINC02382* | 39 | 559 | 1,04E-08 | 38 | 473 | 7,14E-08 | 34 | 380 | 4,60E-02 | 25 | 3218 | 0,891 | 25 | 3069 | 0,518 | 25 | 2982 | 0,939 |
| | 4 | 170822001 | 170826000 | *LINC02382* | 27 | 369 | 1,12E-07 | 27 | 316 | 2,37E-07 | 26 | 258 | 1,03E-01 | 20 | 1853 | 0,951 | 20 | 1771 | 0,482 | 20 | 1701 | 0,637 |
| | 4 | 170824001 | 170828000 | *LINC02382* | 29 | 564 | 7,49E-08 | 29 | 486 | 7,28E-08 | 27 | 361 | 2,91E-01 | 22 | 3282 | 0,778 | 22 | 3141 | 0,818 | 22 | 3040 | 0,864 |
| | 4 | 170826001 | 170830000 | *LINC02382* | 23 | 550 | 8,02E-08 | 23 | 468 | 8,48E-08 | 21 | 338 | 1,99E-01 | 14 | 2740 | 0,766 | 14 | 2624 | 0,583 | 14 | 2548 | 0,921 |
| | 4 | 170828001 | 170832000 | *LINC02382* | 24 | 398 | 6,91E-08 | 24 | 331 | 1,62E-07 | 23 | 267 | 2,50E-02 | 9 | 932 | 0,929 | 9 | 887 | 0,274 | 9 | 858 | 0,930 |
| | 4 | 170830001 | 170834000 | *LINC02382* | 27 | 365 | 1,35E-07 | 26 | 303 | 3,31E-07 | 27 | 254 | 7,80E-03 | 15 | 1542 | 0,943 | 15 | 1471 | 0,335 | 15 | 1424 | 0,751 |
| | 4 | 170844001 | 170848000 | *LINC02382* | 18 | 198 | 3,44E-07 | 18 | 163 | 3,59E-07 | 17 | 131 | 4,70E-02 | 14 | 1367 | 0,892 | 14 | 1304 | 0,471 | 14 | 1265 | 0,868 |
| | 4 | 170846001 | 170850000 | *LINC02382* | 15 | 282 | 3,04E-08 | 15 | 244 | 1,03E-08 | 14 | 189 | 1,60E-01 | 11 | 1582 | 0,644 | 11 | 1514 | 0,723 | 11 | 1468 | 0,913 |
| | 4 | 170854001 | 170858000 | *LINC02382* | 19 | 189 | 4,85E-07 | 18 | 259 | 8,75E-08 | 17 | 202 | 5,30E-02 | 15 | 1950 | 0,985 | 15 | 1861 | 0,768 | 15 | 1796 | 0,699 |
| | 4 | 170862001 | 170866000 | *LINC02382* | 21 | 155 | 4,96E-07 | 20 | 130 | 2,55E-07 | 20 | 104 | 8,90E-02 | 10 | 1000 | 0,794 | 10 | 945 | 0,702 | 10 | 912 | 0,904 |
| | 4 | 170864001 | 170868000 | *LINC02382* | 18 | 209 | 4,14E-07 | 16 | 178 | 2,20E-07 | 16 | 144 | 1,27E-01 | 13 | 1408 | 0,924 | 13 | 1335 | 0,858 | 13 | 1288 | 0,766 |
| | 4 | 170874001 | 170878000 | *LINC02382* | 28 | 295 | 1,72E-07 | 26 | 253 | 2,68E-07 | 26 | 207 | 6,70E-02 | 13 | 1825 | 0,979 | 13 | 1732 | 0,913 | 13 | 1675 | 0,076 |
| | 4 | 170878001 | 170882000 | *LINC02382* | 18 | 321 | 2,32E-07 | 16 | 265 | 7,63E-07 | 15 | 205 | 8,84E-01 | 14 | 2150 | 0,837 | 14 | 2026 | 0,570 | 14 | 1980 | 0,561 |
| | 4 | 170880001 | 170884000 | *LINC02382* | 19 | 265 | 4,55E-07 | 20 | 255 | 2,26E-06 | 20 | 205 | 7,70E-02 | 18 | 2168 | 0,943 | 18 | 2050 | 0,323 | 18 | 2002 | 0,851 |
| | 4 | 171010001 | 171014000 | *LINC02382* | 27 | 277 | 3,67E-07 | 27 | 243 | 2,60E-07 | 26 | 200 | 1,87E-01 | 16 | 1194 | 0,879 | 16 | 1138 | 0,855 | 15 | 744 | 0,738 |
| | 4 | 171032001 | 171036000 | *LINC02382* | 19 | 223 | 3,50E-07 | 19 | 185 | 1,10E-06 | 18 | 161 | 5,00E-03 | 12 | 1344 | 0,996 | 12 | 1263 | 0,119 | 12 | 1244 | 0,777 |
| | 4 | 171050001 | 171054000 | *LINC02382* | 28 | 221 | 4,71E-07 | 26 | 191 | 8,34E-08 | 20 | 151 | 4,93E-01 | 14 | 1573 | 0,885 | 14 | 1499 | 0,512 | 12 | 732 | 0,302 |
| | 4 | 171070001 | 171074000 | *LINC02382* | 45 | 542 | 3,71E-07 | 45 | 447 | 1,75E-07 | 42 | 325 | 2,80E-02 | 39 | 3763 | 0,941 | 39 | 3554 | 0,143 | 36 | 2388 | 0,516 |
| | 4 | 171078001 | 171082000 | *LINC02382* | 25 | 415 | 2,06E-07 | 25 | 349 | 2,19E-07 | 23 | 269 | 7,30E-02 | 12 | 643 | 0,957 | 12 | 596 | 0,111 | 12 | 593 | 0,624 |
| | 5 | 149692001 | 149696000 | *ARHGEF37, LOC105378224* | 20 | 103 | 3,95E-07 | 19 | 93 | 1,78E-08 | 17 | 74 | 7,17E-01 | 12 | 813 | 0,089 | 12 | 760 | 0,484 | - | - | - |
| **MAF<1%** | 12 | 40950001 | 40954000 | *CNTN1* | 13 | 36 | 6,51E-08 | 11 | 26 | 2,20E-04 | 12 | 24 | 2,10E-08 | 2 | 18 | 0,326 | 2 | 17 | 0,239 | 2 | 17 | 0,578 |
| | 14 | 59004001 | 59008000 | *LINC01500* | 13 | 19 | 2,53E-07 | 13 | 17 | 6,66E-06 | 12 | 14 | 3,56E-01 | 4 | 56 | 0,015 | 4 | 50 | 0,088 | 4 | 46 | 0,027 |
| | 2 | 85328001 | 85332000 | *TGOLN2* | 14 | 31 | 2,65E-07 | 11 | 23 | 1,10E-04 | 10 | 18 | 2,10E-03 | 7 | 215 | 0,679 | 7 | 202 | 0,122 | 7 | 205 | 0,037 |
| | 3 | 139636001 | 139640000 | *NMNAT3* | 7 | 40 | 4,75E-07 | 7 | 36 | 9,41E-10 | 7 | 22 | 8,54E-01 | 4 | 127 | 0,705 | 4 | 120 | 0,747 | 4 | 115 | 0,162 |
| | 3 | 2384001 | 2388000 | *CNTN4* | 20 | 51 | 2,58E-07 | 19 | 42 | 3,73E-06 | 18 | 33 | 2,45E-05 | 7 | 129 | 0,777 | 7 | 124 | 0,813 | 7 | 120 | 0,565 |
| | 5 | 31732001 | 31736000 | *PDZD2* | 15 | 45 | 4,70E-07 | 15 | 38 | 1,96E-07 | 15 | 28 | 6,35E-01 | 6 | 199 | 0,880 | 6 | 193 | 0,512 | 6 | 182 | 0,462 |
| | 5 | 58516001 | 58520000 | *GAPT, LINC02108* | 5 | 11 | 3,30E-07 | 4 | 5 | 8,55E-03 | 5 | 15 | 8,81E-05 | 2 | 71 | 0,122 | 2 | 63 | 0,528 | 2 | 60 | 0,766 |

**Supplementary Table 19:** Variants in the discovered genomic windows (STAAR-O) with replication in T1D (FinnDiane) and general population (FinnGen)

*Note: Stroke subtype analyses may have included additional variants (i.e. passed MAF threshold in smaller data)*

| MODEL | CLOSEST GENE | WINDOW | CHR | POS | rsnumber | RegulomeD | GTEx (m | eQTLGen | PCHi-C (hi | REF | ALT | CADD | TF | TSS/TES | EPIGEN | CONS | PROT | microRN | MUTAT | Stroke MAF | Stroke EFF |
|---|---|---|---|---|---|---|---|---|---|---|---|---|---|---|---|---|---|---|---|---|---|
| | | | | | | | | | | | | PHRED SCALE SCORE | | | | | | | | Discovery cohort FinnDiane WGS using score test (T1D specific) | |
| MAF<1% | *CNTN1* | 12:40950001-40954000 | 12 | 40950375 | rs114294967 | 0.18412 (7) | - | - | - | C | A | 3,12 | 1,34 | 2,69 | 1,87 | 3,66 | 3,00 | 2,97 | 7,16 | 0 % | -1,374 |
| | | | 12 | 40950918 | - | - | - | - | - | C | CAT | 0,49 | 1,34 | 2,70 | 0,73 | 1,61 | 3,00 | 2,97 | 11,12 | 0 % | -1,126 |
| | | | 12 | 40950957 | rs201239947 | 0.34847 (6) | - | - | - | C | CAT | 0,95 | 1,34 | 2,70 | 0,65 | 4,06 | 3,00 | 2,97 | 11,37 | 1 % | 3,870 |
| | | | 12 | 40951144 | rs1235859151 | 0.17448 (6) | - | - | - | T | C | 16,42 | 1,34 | 2,70 | 3,94 | 10,79 | 3,00 | 2,97 | 10,79 | 0 % | 2,127 |
| | | | 12 | 40951472 | rs1417797054 | 0.99 (5) | - | - | - | T | G | 5,05 | 1,34 | 2,71 | 2,07 | 5,54 | 3,00 | 2,97 | 9,78 | 0 % | -1,309 |
| | | | 12 | 40951573 | rs145977060 | 0.58955 (5) | - | - | - | G | GT | 1,48 | 1,34 | 2,71 | 2,43 | 5,36 | 3,00 | 2,97 | 9,88 | 0 % | -1,130 |
| | | | 12 | 40951712 | rs747062390 | 0.00667 (6) | - | - | - | AT | A | 3,02 | 1,34 | 2,72 | 0,63 | 12,06 | 3,00 | 2,97 | 12,29 | 1 % | 4,919 |
| | | | 12 | 40952116 | rs117378297 | 0.4986 (6) | - | - | - | A | G | 5,14 | 1,34 | 2,73 | 4,24 | 3,79 | 3,00 | 2,97 | 11,11 | 0 % | -1,126 |
| | | | 12 | 40952223 | rs181026504 | 0.18412 (7) | - | - | - | T | C | 0,29 | 1,34 | 2,73 | 4,09 | 0,33 | 3,00 | 2,97 | 11,18 | 1 % | -0,315 |
| | | | 12 | 40952332 | rs779970390 | 0.18412 (7) | - | - | - | G | C | 6,35 | 1,34 | 2,73 | 1,75 | 9,31 | 3,00 | 2,97 | 11,70 | 0 % | 6,000 |
| | | | 12 | 40953276 | rs577550508 | 0.60906 (4) | - | - | *YAF2* first | G | T | 19,10 | 6,45 | 2,76 | 4,66 | 20,36 | 3,00 | 2,97 | 9,52 | 0 % | 6,000 |
| | | | 12 | 40953677 | rs969389248 | 0.2909 (6) | - | - | *YAF2* first | T | C | 9,62 | 1,34 | 2,77 | 1,45 | 10,07 | 3,00 | 2,97 | 8,82 | 0 % | 6,625 |
| | | | 12 | 40953688 | rs570584679 | 0.42214 (6) | - | - | *YAF2* first | A | G | 0,52 | 1,34 | 2,77 | 1,28 | 0,71 | 3,00 | 2,97 | 8,70 | 0 % | 1,720 |
| MAF<1% | *TGOLN2* | 2:85328001-85332000 | 2 | 85328016 | rs376006909 | 0.70497 (4) | - | - | 27 region: | G | T | 0,79 | 36,22 | 21,87 | 32,47 | 0,07 | 3,00 | 2,97 | 0,67 | 0 % | 3,425 |
| | | | 2 | 85328462 | rs1182214256 | 0.4945 (3a) | - | - | 27 region: | C | T | 10,12 | 25,20 | 17,57 | 31,06 | 6,25 | 3,00 | 2,97 | 1,35 | 0 % | 8,356 |
| | | | 2 | 85328585 | rs542060093 | 1 (2c) | - | - | 27 region: | C | T | 8,59 | 17,88 | 13,93 | 21,88 | 4,55 | 3,00 | 2,97 | 2,64 | 0 % | 5,039 |
| | | | 2 | 85329098 | rs1252548396 | 0.60906 (4) | - | - | 27 region: | A | G | 4,78 | 12,73 | 9,53 | 9,98 | 3,48 | 3,00 | 2,97 | 3,17 | 0 % | 3,173 |
| | | | 2 | 85329702 | rs1188244022 | 0.60906 (4) | - | - | 27 region: | T | A | 11,54 | 12,21 | 10,50 | 9,83 | 12,01 | 3,00 | 2,97 | 3,09 | 0 % | -1,306 |
| | | | 2 | 85330021 | - | - | - | - | - | A | G | 5,27 | 1,34 | 10,30 | 1,95 | 4,99 | 3,00 | 2,97 | 5,59 | 0 % | -1,337 |
| | | | 2 | 85330469 | rs184693178 | 0 (6) | - | - | 27 region: | T | A | 2,96 | 1,34 | 10,16 | 2,46 | 3,53 | 3,00 | 2,97 | 1,72 | 0 % | 1,209 |
| | | | 2 | 85330639 | rs149295883 | 0.191 (5) | - | *RETSAT,A* | 27 region: | C | T | 4,61 | 1,34 | 10,15 | 0,95 | 5,44 | 3,00 | 2,97 | 3,33 | 0 % | 7,296 |
| | | | 2 | 85331227 | rs1366422232 | 0.1525 (3a) | - | - | 27 region: | A | C | 7,72 | 7,64 | 11,03 | 12,76 | 11,27 | 3,00 | 2,97 | 2,95 | 0 % | -1,448 |
| | | | 2 | 85331292 | rs188184516 | 0.60906 (4) | - | - | 27 region: | C | T | 0,26 | 7,64 | 11,16 | 13,00 | 0,03 | 3,00 | 2,97 | 1,22 | 0 % | 9,005 |
| | | | 2 | 85331379 | rs1682917715 | - | - | - | - | C | G | 1,53 | 5,42 | 11,35 | 10,78 | 0,91 | 3,00 | 2,97 | 1,35 | 0 % | 11,612 |
| | | | 2 | 85331645 | - | - | - | - | - | G | C | 4,51 | 9,19 | 12,00 | 8,57 | 5,56 | 3,00 | 2,97 | 2,07 | 0 % | -1,112 |
| | | | 2 | 85331688 | rs143332348 | 0.5497 (3a) | - | - | 27 region: | A | C | 6,94 | 3,99 | 12,12 | 9,57 | 7,32 | 3,00 | 2,97 | 1,94 | 1 % | 1,488 |
| | | | 2 | 85331818 | rs1310271374 | 0.5321 (5) | - | - | 27 region: | A | G | 7,40 | 5,42 | 12,52 | 1,48 | 7,59 | 3,00 | 2,97 | 1,47 | 0 % | 4,587 |
| MAF<1% | *LINC01500* | 14:59004001-59008000 | 14 | 59004302 | - | - | - | - | - | A | G | 2,68 | 11,53 | 3,26 | 5,07 | 1,62 | 3,00 | 2,97 | 2,66 | 0 % | -1,146 |
| | | | 14 | 59004483 | rs976297611 | 0.60906 (4) | - | - | *DACT1* in | C | A | 0,74 | 1,34 | 3,30 | 3,69 | 1,12 | 3,00 | 2,97 | 3,39 | 0 % | 4,723 |
| | | | 14 | 59004745 | rs1884935226 | - | - | - | - | C | T | 2,46 | 1,34 | 3,36 | 0,78 | 4,07 | 3,00 | 2,97 | 2,02 | 0 % | -1,366 |
| | | | 14 | 59004836 | rs1241730246 | 0.37269 (6) | - | - | *DACT1* in | ATCA | A | 1,31 | 1,34 | 3,39 | 0,93 | 5,65 | 3,00 | 2,97 | 2,54 | 0 % | -1,538 |
| | | | 14 | 59005000 | rs569284427 | 0.18412 (7) | - | - | *DACT1* in | A | G | 5,43 | 1,34 | 3,43 | 4,14 | 5,97 | 3,00 | 2,97 | 5,38 | 0 % | -1,337 |
| | | | 14 | 59005063 | rs181980596 | 0.58955 (5) | - | - | *DACT1* in | A | G | 6,89 | 3,99 | 3,45 | 5,65 | 7,42 | 3,00 | 2,97 | 3,36 | 0 % | 7,341 |
| | | | 14 | 59005164 | rs745771590 | 0.58955 (5) | - | - | *DACT1* in | G | A | 0,42 | 1,34 | 3,48 | 4,75 | 1,39 | 3,00 | 2,97 | 1,10 | 0 % | -1,281 |
| | | | 14 | 59005236 | rs574091763 | 0.58955 (5) | - | - | *DACT1* in | A | G | 0,54 | 1,34 | 3,50 | 3,30 | 0,47 | 3,00 | 2,97 | 2,10 | 0 % | -1,103 |
| | | | 14 | 59005248 | rs1346810491 | 0.20355 (5) | - | - | *DACT1* in | T | G | 3,76 | 1,34 | 3,50 | 2,78 | 4,60 | 3,00 | 2,97 | 3,54 | 0 % | -1,232 |
| | | | 14 | 59006333 | rs1176375897 | 0.63479 (5) | - | - | *DACT1* in | A | AAA | 2,82 | 5,42 | 3,89 | 0,42 | 8,75 | 3,00 | 2,97 | 1,15 | 0 % | -1,230 |
| | | | 14 | 59006632 | rs1192857274 | 0.7615 (5) | - | - | *DACT1* in | A | G | 0,68 | 6,45 | 4,03 | 5,03 | 1,02 | 3,00 | 2,97 | 1,57 | 0 % | -1,271 |
| | | | 14 | 59007383 | rs1281241634 | 0.2909 (6) | - | - | *DACT1* in | C | A | 2,98 | 1,34 | 4,48 | 1,74 | 4,02 | 3,00 | 2,97 | 3,75 | 0 % | -1,475 |
| | | | 14 | 59007972 | rs1054862723 | 0.7707 (6) | - | - | *DACT1* in | A | G | 4,90 | 1,34 | 5,01 | 2,09 | 3,75 | 3,00 | 2,97 | 5,30 | 0 % | -1,337 |
| MAF<1% | *CNTN4* | 3:2384001-2388000 | 3 | 2384190 | rs781618452 | 0.81869 (6) | - | - | - | CATT | C | 1,80 | 3,99 | 0,31 | 2,18 | 5,63 | 3,00 | 2,97 | 0,09 | 0 % | 0,975 |
| | | | 3 | 2384207 | rs536421003 | 0.44483 (6) | - | - | - | T | C | 6,82 | 3,99 | 0,31 | 2,08 | 6,28 | 3,00 | 2,97 | 0,14 | 1 % | 0,868 |
| | | | 3 | 2384386 | rs771362280 | 0.13454 (5) | - | - | - | C | G | 1,34 | 6,88 | 0,31 | 5,06 | 1,65 | 3,00 | 2,97 | 0,10 | 0 % | 4,875 |
| | | | 3 | 2384404 | rs545611879 | 0 (5) | - | - | - | C | T | 3,32 | 6,88 | 0,31 | 4,08 | 3,57 | 3,00 | 2,97 | 0,09 | 0 % | -1,195 |
| | | | 3 | 2384477 | rs1384295553 | 0.65638 (5) | - | - | - | G | C | 12,75 | 4,89 | 0,31 | 2,27 | 17,41 | 3,00 | 2,97 | 0,17 | 0 % | -1,153 |
| | | | 3 | 2384890 | rs149716010 | 0.13454 (5) | - | - | *CNTN4* in | T | C | 0,97 | 1,34 | 0,31 | 2,02 | 0,64 | 3,00 | 2,97 | 0,14 | 1 % | 2,299 |
| | | | 3 | 2384937 | rs911340675 | 0.13454 (5) | - | - | *CNTN4* in | G | A | 2,97 | 1,34 | 0,31 | 2,94 | 4,93 | 3,00 | 2,97 | 0,10 | 0 % | -1,317 |
| | | | 3 | 2384958 | rs192643682 | 0.13454 (5) | - | - | *CNTN4* in | G | A | 0,09 | 1,34 | 0,31 | 3,15 | 0,23 | 3,00 | 2,97 | 0,16 | 0 % | 0,999 |
| | | | 3 | 2384993 | - | - | - | - | - | T | C | 0,12 | 1,34 | 0,31 | 3,28 | 0,08 | 3,00 | 2,97 | 0,15 | 0 % | -1,386 |
| | | | 3 | 2385225 | rs1444081748 | 0.18412 (7) | - | - | *CNTN4* in | T | TGC. | 2,55 | 1,34 | 0,31 | 0,56 | 10,06 | 3,00 | 2,97 | 0,18 | 0 % | 5,502 |
| | | | 3 | 2385227 | rs1374949310 | 0.18412 (7) | - | - | *CNTN4* in | T | A | 0,61 | 1,34 | 0,31 | 0,56 | 0,35 | 3,00 | 2,97 | 0,18 | 0 % | 3,774 |
| | | | 3 | 2385228 | rs572060569 | 0.18412 (7) | - | - | *CNTN4* in | A | G | 3,08 | 1,34 | 0,31 | 0,57 | 1,98 | 3,00 | 2,97 | 0,18 | 0 % | 1,610 |
| | | | 3 | 2385871 | rs557034302 | 0.34125 (6) | - | - | *CNTN4* in | G | A | 0,86 | 1,34 | 0,31 | 1,70 | 1,25 | 3,00 | 2,97 | 0,06 | 0 % | -1,503 |
| | | | 3 | 2385952 | rs934528344 | 0 (6) | - | - | *CNTN4* in | C | T | 2,71 | 1,34 | 0,31 | 1,72 | 2,72 | 3,00 | 2,97 | 0,09 | 0 % | -1,504 |
| | | | 3 | 2386125 | rs934755027 | 0.20829 (6) | - | - | *CNTN4* in | A | G | 7,18 | 1,34 | 0,31 | 2,30 | 3,48 | 3,00 | 2,97 | 0,13 | 0 % | 3,122 |
| | | | 3 | 2386718 | rs1466300012 | 0.49716 (6) | - | - | *CNTN4* in | A | G | 6,10 | 1,34 | 0,30 | 1,30 | 4,96 | 3,00 | 2,97 | 0,31 | 0 % | 7,443 |
| | | | 3 | 2387292 | rs770050189 | 0.18412 (7) | - | - | *CNTN4* in | C | G | 0,04 | 1,34 | 0,30 | 0,70 | 0,08 | 3,00 | 2,97 | 0,21 | 0 % | 4,097 |
| | | | 3 | 2387492 | rs757336335 | - | - | - | *CNTN4* in | C | G | 0,07 | 1,34 | 0,30 | 0,71 | 0,17 | 3,00 | 2,97 | 0,15 | 0 % | -1,117 |
| | | | 3 | 2387742 | - | - | - | - | - | A | G | 4,40 | 1,34 | 0,30 | 2,31 | 2,97 | 3,00 | 2,97 | 0,20 | 0 % | -1,132 |
| | | | 3 | 2387800 | rs554832365 | 0.25628 (6) | - | - | *CNTN4* in | G | A | 15,05 | 1,34 | 0,30 | 3,71 | 19,57 | 3,00 | 2,97 | 0,14 | 0 % | 4,407 |
| MAF<1% | *NMNAT3* | 3:139636001-139640000 | 3 | 139636256 | rs192927615 | 0.65422 (6) | - | - | - | T | C | 10,68 | 3,99 | 5,78 | 1,66 | 15,42 | 3,00 | 2,97 | 4,80 | 1 % | 2,510 |
| | | | 3 | 139636448 | rs76008532 | 0.60906 (4) | *NMNAT3* | - | - | A | G | 17,61 | 6,00 | 5,65 | 4,23 | 17,68 | 3,00 | 2,97 | 3,22 | 0 % | 5,602 |
| | | | 3 | 139636640 | rs575465521 | 0.23772 (5) | - | - | - | C | T | 20,26 | 3,99 | 5,54 | 5,31 | 23,15 | 3,00 | 2,97 | 5,02 | 0 % | -1,419 |
| | | | 3 | 139637811 | rs113688313 | 0.13454 (5) | - | - | - | A | G | 5,63 | 1,34 | 5,02 | 2,77 | 5,39 | 3,00 | 2,97 | 6,19 | 1 % | 0,455 |
| | | | 3 | 139637890 | rs1460012412 | 0.13454 (5) | - | - | - | A | C | 4,14 | 1,34 | 4,99 | 2,11 | 2,79 | 3,00 | 2,97 | 6,93 | 0 % | -1,309 |
| | | | 3 | 139638322 | - | - | - | - | - | T | G | 1,21 | 1,34 | 4,85 | 3,54 | 1,21 | 3,00 | 2,97 | 6,38 | 0 % | -1,329 |
| | | | 3 | 139639253 | rs183054474 | 0.24688 (5) | - | - | - | C | T | 8,75 | 3,99 | 4,60 | 3,49 | 6,97 | 3,00 | 2,97 | 2,79 | 1 % | 3,143 |
| MAF<1% | *GAPT, LINC02108* | 5:58516001-58520000 | 5 | 58516862 | rs189910885 | 0.38564 (6) | - | - | - | T | C | 5,35 | 1,34 | 4,72 | 0,95 | 4,95 | 3,00 | 2,97 | 4,92 | 0 % | 6,129 |
| | | | 5 | 58517679 | rs539571055 | 0.58955 (5) | ? | - | - | T | G | 8,11 | 3,99 | 4,90 | 2,33 | 8,47 | 3,00 | 2,97 | 3,43 | 0 % | 4,110 |
| | | | 5 | 58517792 | rs150448509 | 0.58955 (5) | - | - | - | G | A | 0,95 | 1,34 | 4,92 | 3,78 | 0,74 | 3,00 | 2,97 | 2,70 | 0 % | 4,788 |
| | | | 5 | 58518723 | rs554752975 | 0.18412 (7) | - | - | - | T | C | 6,10 | 1,34 | 5,13 | 1,29 | 5,09 | 3,00 | 2,97 | 6,83 | 0 % | -1,140 |
| | | | 5 | 58519917 | rs958186884 | 0.18412 (7) | ? | - | - | G | T | 2,01 | 1,34 | 5,44 | 1,77 | 3,62 | 3,00 | 2,97 | 7,46 | 0 % | 4,082 |
| MAF<1% | *PDZD2* | 5:31732001-31736000 | 5 | 31732083 | rs575726103 | 0.13454 (5) | - | - | *PDZD2* an | C | T | 3,73 | 9,69 | 2,33 | 5,79 | 2,93 | 3,00 | 2,97 | 9,30 | 0 % | -1,475 |
| | | | 5 | 31732315 | rs114748678 | 0.58955 (5) | - | - | *PDZD2* an | C | T | 2,27 | 5,42 | 2,33 | 3,14 | 2,71 | 3,00 | 2,97 | 6,53 | 1 % | 4,599 |
| | | | 5 | 31732616 | rs540364560 | 0.18412 (7) | - | - | *PDZD2* an | C | T | 12,00 | 1,34 | 2,32 | 5,12 | 12,47 | 3,00 | 2,97 | 7,31 | 0 % | -1,210 |
| | | | 5 | 31732890 | rs1047761267 | 0.18412 (7) | - | - | *PDZD2* an | C | T | 0,93 | 1,34 | 2,31 | 1,06 | 0,71 | 3,00 | 2,97 | 9,60 | 0 % | -1,164 |
| | | | 5 | 31733184 | - | - | - | - | - | C | A | 0,22 | 7,29 | 2,30 | 5,12 | 0,47 | 3,00 | 2,97 | 7,14 | 0 % | -1,287 |
| | | | 5 | 31733316 | rs145397958 | 0.13454 (5) | - | *PDZD2* | *PDZD2* an | T | C | 4,59 | 7,29 | 2,30 | 6,21 | 1,50 | 3,00 | 2,97 | 3,94 | 0 % | -1,246 |
| | | | 5 | 31733378 | rs948315861 | 0.13454 (5) | - | - | *PDZD2* an | G | A | 1,90 | 8,88 | 2,29 | 7,42 | 1,90 | 3,00 | 2,97 | 5,48 | 0 % | -1,475 |
| | | | 5 | 31733512 | - | - | - | - | - | T | C | 1,78 | 10,70 | 2,29 | 6,44 | 0,60 | 3,00 | 2,97 | 7,70 | 0 % | -1,154 |
| | | | 5 | 31733779 | rs1749678064 | - | - | - | - | T | C | 1,32 | 10,06 | 2,28 | 5,85 | 0,63 | 3,00 | 2,97 | 4,67 | 0 % | -1,344 |
| | | | 5 | 31734539 | rs145610804 | 0.30584 (6) | - | - | *PDZD2* an | G | A | 1,36 | 1,34 | 2,26 | 0,85 | 1,63 | 3,00 | 2,97 | 3,07 | 0 % | -1,533 |
| | | | 5 | 31734734 | rs143615818 | 0.13454 (5) | - | - | *PDZD2* an | G | A | 1,72 | 3,99 | 2,26 | 3,10 | 2,57 | 3,00 | 2,97 | 3,06 | 0 % | 3,592 |
| | | | 5 | 31734760 | rs115717328 | 0.13454 (5) | - | - | *PDZD2* an | T | C | 4,89 | 6,45 | 2,25 | 4,80 | 4,58 | 3,00 | 2,97 | 3,27 | 1 % | 4,599 |
| | | | 5 | 31734869 | rs761403037 | 0.84691 (3a | - | - | *PDZD2* an | G | A | 9,55 | 6,45 | 2,25 | 4,15 | 7,55 | 3,00 | 2,97 | 2,88 | 0 % | -1,118 |
| | | | 5 | 31735171 | - | - | - | - | - | G | A | 1,14 | 8,63 | 2,24 | 7,97 | 1,43 | 3,00 | 2,97 | 2,04 | 0 % | -1,137 |
| | | | 5 | 31735616 | rs891357795 | 0.18412 (7) | - | - | *PDZD2* an | A | G | 0,50 | 1,34 | 2,23 | 0,67 | 0,08 | 3,00 | 2,97 | 2,65 | 1 % | 0,160 |
| MAF<5% | *MOB1AP1, SLITRK6* | 13:85912001-85916000 | 13 | 85912240 | rs576724884 | 0.42365 (6) | - | - | *MIR4500H* | G | A | 5,40 | 1,34 | 1,59 | 0,15 | 5,68 | 3,00 | 2,97 | 0,06 | 0 % | -1,131 |
| | | | 13 | 85912252 | rs370553970 | 0.67385 (6) | - | - | *MIR4500H* | T | C | 5,87 | 1,34 | 1,59 | 0,16 | 1,60 | 3,00 | 2,97 | 0,07 | 0 % | -1,127 |
| | | | 13 | 85912295 | rs1476733006 | 0.18412 (7) | - | - | *MIR4500H* | G | C | 7,96 | 1,34 | 1,59 | 0,16 | 7,45 | 3,00 | 2,97 | 0,07 | 0 % | 3,508 |
| | | | 13 | 85912423 | rs182183815 | 0.18412 (7) | - | - | *MIR4500H* | C | T | 6,33 | 1,34 | 1,59 | 0,18 | 6,08 | 3,00 | 2,97 | 0,14 | 0 % | -1,150 |
| | | | 13 | 85912476 | rs1037250817 | - | - | - | *MIR4500H* | C | T | 1,84 | 1,34 | 1,59 | 0,17 | 0,53 | 3,00 | 2,97 | 0,17 | 0 % | -1,628 |
| | | | 13 | 85912535 | rs570834227 | 0.18412 (7) | - | - | *MIR4500H* | T | C | 8,10 | 1,34 | 1,59 | 0,15 | 6,97 | 3,00 | 2,97 | 0,11 | 0 % | 2,893 |
| | | | 13 | 85912638 | rs749552742 | 0.49716 (6) | - | - | - | GT | G | 7,13 | 1,34 | 1,60 | 0,16 | 9,21 | 3,00 | 2,97 | 0,15 | 2 % | 2,494 |
| | | | 13 | 85912766 | rs1376936399 | - | - | - | - | C | A | 3,95 | 1,34 | 1,60 | 0,15 | 3,65 | 3,00 | 2,97 | 0,18 | 0 % | 2,957 |
| | | | 13 | 85913481 | rs551699196 | 0.18412 (7) | - | - | - | C | T | 5,98 | 1,34 | 1,62 | 0,15 | 6,18 | 3,00 | 2,97 | 0,20 | 2 % | 2,494 |
| | | | 13 | 85913506 | rs543942416 | 0.18412 (7) | - | - | - | GT | G | 6,93 | 1,34 | 1,62 | 0,15 | 9,38 | 3,00 | 2,97 | 0,17 | 1 % | 0,308 |
| | | | 13 | 85913615 | - | - | - | - | - | A | T | 13,06 | 1,34 | 1,62 | 0,15 | 12,24 | 3,00 | 2,97 | 0,15 | 0 % | 3,845 |
| | | | 13 | 85914083 | rs531175441 | 0.26281 (6) | - | - | - | A | G | 3,00 | 1,34 | 1,64 | 0,00 | 0,42 | 3,00 | 2,97 | 0,27 | 0 % | 2,964 |
| | | | 13 | 85914177 | rs914554122 | - | - | - | - | C | T | 1,66 | 1,34 | 1,64 | 0,15 | 0,32 | 3,00 | 2,97 | 0,23 | 0 % | 2,968 |
| | | | 13 | 85914224 | rs190326090 | - | - | - | - | G | C | 9,17 | 1,34 | 1,64 | 0,16 | 10,82 | 3,00 | 2,97 | 0,15 | 3 % | -0,398 |
| | | | 13 | 85914235 | - | - | - | - | - | T | G | 9,43 | 1,34 | 1,64 | 0,16 | 9,33 | 3,00 | 2,97 | 0,16 | 0 % | 5,150 |
| | | | 13 | 85914242 | rs539363858 | 0.2212 (6) | - | - | - | C | T | 6,62 | 1,34 | 1,64 | 0,16 | 7,51 | 3,00 | 2,97 | 0,17 | 0 % | 4,178 |
| | | | 13 | 85914341 | rs192795728 | 0.23675 (6) | - | - | - | A | G | 5,26 | 1,34 | 1,64 | 0,17 | 3,65 | 3,00 | 2,97 | 0,15 | 1 % | 0,245 |
| | | | 13 | 85914381 | rs1304139912 | 0.18412 (7) | - | - | - | C | G | 5,39 | 1,34 | 1,64 | 0,16 | 6,49 | 3,00 | 2,97 | 0,13 | 0 % | 1,777 |
| | | | 13 | 85915006 | rs535131400 | 0.18653 (6) | - | - | - | A | G | 17,51 | 1,34 | 1,66 | 0,15 | 14,74 | 3,00 | 2,97 | 0,13 | 1 % | 0,736 |
| | | | 13 | 85915137 | rs147527898 | 0.18412 (7) | - | - | - | G | T | 9,49 | 1,34 | 1,67 | 0,15 | 10,88 | 3,00 | 2,97 | 0,18 | 2 % | 2,494 |
| | | | 13 | 85915307 | rs563110572 | 0.18412 (7) | - | - | - | C | T | 2,71 | 1,34 | 1,67 | 0,17 | 3,34 | 3,00 | 2,97 | 0,15 | 0 % | 9,107 |
| MAF<5% | *LINC02382* | 4:170752001-170756000 | 4 | 170752424 | rs529444136 | 0.17931 (6) | - | - | - | G | T | 0,17 | 1,34 | 5,67 | 0,88 | 0,65 | 3,00 | 2,97 | 2,08 | 0 % | -1,128 |
| | | | 4 | 170752505 | - | - | - | - | - | GTAT | G | 4,44 | 1,34 | 5,65 | 0,73 | 10,06 | 3,00 | 2,97 | 1,55 | 1 % | 0,171 |
| | | | 4 | 170752534 | rs12641264 | 0.13144 (6) | - | - | - | T | A | 11,53 | 1,34 | 5,64 | 0,97 | 9,14 | 3,00 | 2,97 | 1,63 | 0 % | 3,693 |
| | | | 4 | 170752821 | rs115070815 | 0.18412 (7) | - | - | - | C | T | 0,35 | 1,34 | 5,56 | 2,54 | 0,18 | 3,00 | 2,97 | 0,73 | 4 % | -0,367 |
| | | | 4 | 170752822 | rs144656210 | 0.18412 (7) | - | - | - | G | A | 2,28 | 1,34 | 5,56 | 2,50 | 5,55 | 3,00 | 2,97 | 0,73 | 1 % | 0,780 |
| | | | 4 | 170752870 | rs28550091 | 0.18412 (7) | - | - | - | C | T | 8,82 | 1,34 | 5,55 | 1,53 | 12,04 | 3,00 | 2,97 | 0,45 | 4 % | 1,828 |
| | | | 4 | 170752975 | rs35342597 | 0.67105 (6) | *LINC024* | *MFAP3L* | - | C | T | 1,76 | 1,34 | 5,52 | 1,02 | 3,60 | 3,00 | 2,97 | 1,31 | 4 % | -0,121 |
| | | | 4 | 170753064 | rs148690806 | - | - | - | - | T | C | 1,72 | 1,34 | 5,49 | 1,19 | 0,96 | 3,00 | 2,97 | 1,33 | 1 % | 3,176 |
| | | | 4 | 170753186 | rs577665150 | 0.18412 (7) | - | - | - | T | G | 3,44 | 1,34 | 5,46 | 2,13 | 5,19 | 3,00 | 2,97 | 1,00 | 0 % | -1,265 |
| | | | 4 | 170753287 | rs72696883 | 0.12326 (5) | *LINC024* | - | - | C | A | 4,18 | 1,34 | 5,43 | 1,27 | 7,54 | 3,00 | 2,97 | 1,64 | 1 % | 3,176 |
| | | | 4 | 170753551 | rs1735171873 | - | - | - | - | T | A | 1,52 | 1,34 | 5,36 | 1,58 | 0,76 | 3,00 | 2,97 | 2,45 | 0 % | 5,440 |
| | | | 4 | 170753927 | rs528985548 | - | - | - | - | G | A | 7,86 | 1,34 | 5,27 | 1,90 | 9,69 | 3,00 | 2,97 | 2,60 | 0 % | -1,337 |
| | | | 4 | 170753952 | rs72696884 | 0.18412 (7) | *LINC024* | - | - | T | C | 3,82 | 1,34 | 5,26 | 2,02 | 4,50 | 3,00 | 2,97 | 1,56 | 1 % | 3,176 |
| | | | 4 | 170753980 | rs72696886 | 0.18412 (7) | *LINC024* | - | - | G | T | 0,56 | 1,34 | 5,25 | 2,68 | 0,90 | 3,00 | 2,97 | 1,18 | 1 % | 3,176 |
| MAF<5% | *LINC02382* | 4:170752001-170756000, 4:170754001-170758000 | 4 | 170754152 | rs111286719 | 0.009 (6) | - | - | - | A | T | 7,37 | 1,34 | 5,21 | 1,08 | 7,47 | 3,00 | 2,97 | 1,53 | 5 % | 1,970 |
| | | | 4 | 170754359 | rs112412452 | 0.60906 (4) | - | - | - | G | A | 1,42 | 4,89 | 5,16 | 1,70 | 1,15 | 3,00 | 2,97 | 0,98 | 4 % | 1,828 |
| | | | 4 | 170754470 | rs1043198722 | - | - | - | - | A | T | 3,47 | 4,89 | 5,13 | 2,92 | 3,51 | 3,00 | 2,97 | 0,68 | 0 % | -1,204 |
| | | | 4 | 170754545 | - | - | - | - | - | A | C | 1,90 | 4,89 | 5,12 | 3,87 | 0,86 | 3,00 | 2,97 | 0,86 | 0 % | 3,845 |
| | | | 4 | 170754570 | rs190108896 | 0.16377 (5) | - | - | - | G | A | 3,06 | 3,99 | 5,11 | 4,74 | 4,41 | 3,00 | 2,97 | 0,97 | 0 % | -1,331 |
| | | | 4 | 170754830 | rs79377369 | 0.13454 (5) | - | - | - | G | A | 4,09 | 1,34 | 5,05 | 2,99 | 6,48 | 3,00 | 2,97 | 1,11 | 1 % | -0,009 |
| | | | 4 | 170754917 | rs112947513 | 0.13454 (5) | - | - | - | A | G | 5,49 | 1,34 | 5,03 | 2,91 | 5,08 | 3,00 | 2,97 | 1,36 | 0 % | 3,693 |
| | | | 4 | 170754921 | rs542829421 | 0.13454 (5) | - | - | - | A | C | 5,52 | 1,34 | 5,03 | 3,17 | 5,31 | 3,00 | 2,97 | 1,29 | 0 % | -1,166 |
| | | | 4 | 170755170 | rs184695069 | 0.18412 (7) | - | - | - | G | A | 1,21 | 1,34 | 4,97 | 0,67 | 1,15 | 3,00 | 2,97 | 2,16 | 1 % | 0,159 |
| | | | 4 | 170755995 | rs1246405789 | - | - | - | - | G | T | 1,83 | 1,34 | 4,79 | 3,95 | 5,38 | 3,00 | 2,97 | 2,06 | 0 % | 3,580 |
| MAF<5% | *LINC02382* | 4:170754001-170758000, 4:170756001-170760000 | 4 | 170756321 | rs755197200 | 0.55602 (6) | - | - | - | T | C | 4,53 | 1,34 | 4,72 | 2,00 | 5,13 | 3,00 | 2,97 | 2,19 | 0 % | -1,464 |
| | | | 4 | 170756517 | rs1403204 | 0.18412 (7) | - | - | - | G | A | 1,32 | 1,34 | 4,68 | 3,96 | 0,81 | 3,00 | 2,97 | 1,08 | 5 % | 1,970 |
| | | | 4 | 170756763 | rs988478193 | 0.60906 (4) | - | - | - | T | C | 5,45 | 5,42 | 4,64 | 1,51 | 4,24 | 3,00 | 2,97 | 1,44 | 0 % | -1,373 |
| | | | 4 | 170756858 | rs1037043516 | 0.60906 (4) | - | - | - | T | C | 10,12 | 7,00 | 4,62 | 3,71 | 11,97 | 3,00 | 2,97 | 0,83 | 0 % | 10,752 |
| | | | 4 | 170756901 | rs143648810 | 0.78994 (3a | - | - | - | G | C | 5,17 | 7,00 | 4,61 | 4,84 | 9,20 | 3,00 | 2,97 | 0,98 | 1 % | 0,616 |
| | | | 4 | 170757099 | rs186858083 | 0.60906 (4) | - | - | - | A | T | 15,70 | 6,45 | 4,57 | 3,80 | 13,85 | 3,00 | 2,97 | 2,64 | 1 % | 0,159 |
| | | | 4 | 170757161 | rs115891080 | 0.60906 (4) | - | - | - | T | C | 11,26 | 3,99 | 4,56 | 3,12 | 11,57 | 3,00 | 2,97 | 1,83 | 0 % | 2,223 |
| | | | 4 | 170757262 | rs186578521 | 0.34847 (6) | - | - | - | C | G | 0,61 | 1,34 | 4,54 | 4,30 | 0,45 | 3,00 | 2,97 | 2,22 | 0 % | -1,243 |
| | | | 4 | 170757781 | rs58057756 | 0.18412 (7) | - | - | - | C | A | 5,95 | 3,99 | 4,44 | 1,31 | 9,60 | 3,00 | 2,97 | 1,38 | 0 % | 3,693 |
| MAF<5% | *LINC02382* | 4:170756001-170760000, 4:170758001-170762000 | 4 | 170758215 | rs60387969 | 0.24039 (6) | - | - | - | T | C | 8,26 | 1,34 | 4,37 | 0,83 | 8,69 | 3,00 | 2,97 | 2,77 | 0 % | 3,693 |
| | | | 4 | 170758303 | rs566837688 | - | - | - | - | A | G | 2,22 | 1,34 | 4,35 | 0,69 | 3,22 | 3,00 | 2,97 | 1,71 | 0 % | 4,828 |
| | | | 4 | 170758398 | rs1735242085 | - | - | - | - | T | A | 3,77 | 1,34 | 4,33 | 0,95 | 4,01 | 3,00 | 2,97 | 3,17 | 0 % | 4,868 |
| | | | 4 | 170758594 | rs778068567 | 0.25628 (6) | - | - | - | A | C | 2,81 | 1,34 | 4,30 | 1,92 | 2,92 | 3,00 | 2,97 | 1,55 | 0 % | 2,360 |
| | | | 4 | 170758855 | rs1434730892 | 0.16346 (6) | - | - | - | TG | T | 3,84 | 1,34 | 4,26 | 2,91 | 8,32 | 3,00 | 2,97 | 1,43 | 0 % | -1,174 |
| | | | 4 | 170758992 | rs184303058 | 0.58955 (5) | - | - | - | T | C | 5,91 | 3,99 | 4,23 | 2,48 | 6,07 | 3,00 | 2,97 | 1,38 | 1 % | 0,225 |
| | | | 4 | 170759024 | rs59810874 | 0.60906 (4) | - | - | - | G | T | 2,16 | 3,99 | 4,23 | 2,49 | 5,19 | 3,00 | 2,97 | 0,85 | 0 % | 3,693 |
| | | | 4 | 170759083 | rs112651432 | 0.74553 (2c | - | - | - | T | C | 7,06 | 10,31 | 4,22 | 3,42 | 7,06 | 3,00 | 2,97 | 0,97 | 3 % | 2,237 |
| | | | 4 | 170759333 | rs1735257798 | - | - | - | - | A | T | 5,33 | 8,63 | 4,18 | 3,26 | 5,26 | 3,00 | 2,97 | 2,39 | 0 % | 4,868 |
| | | | 4 | 170759337 | rs1735257818 | - | - | - | - | G | T | 3,79 | 8,01 | 4,18 | 3,53 | 5,99 | 3,00 | 2,97 | 2,26 | 0 % | 4,868 |
| | | | 4 | 170759869 | rs60300588,rs7 | - | - | - | - | CA | C | 3,48 | 3,99 | 4,09 | 2,75 | 6,36 | 3,00 | 2,97 | 2,21 | 0 % | 3,693 |
| MAF<5% | *LINC02382* | 4:170758001-170762000, 4:170760001-170764000 | 4 | 170760039 | rs564883996 | 0.37351 (6) | - | - | - | G | C | 6,18 | 7,64 | 4,07 | 3,35 | 8,12 | 3,00 | 2,97 | 2,10 | 0 % | -1,117 |
| | | | 4 | 170760371 | rs1482882170 | - | - | - | - | T | A | 5,06 | 1,34 | 4,02 | 1,15 | 4,83 | 3,00 | 2,97 | 1,35 | 0 % | -1,186 |
| | | | 4 | 170760419 | rs28834492 | 0.18412 (7) | - | - | - | T | C | 4,25 | 1,34 | 4,01 | 1,69 | 4,91 | 3,00 | 2,97 | 0,78 | 4 % | 0,389 |
| | | | 4 | 170760429 | rs13129621 | 0.18412 (7) | - | - | - | A | C | 4,45 | 1,34 | 4,01 | 1,82 | 5,72 | 3,00 | 2,97 | 0,79 | 0 % | -1,316 |
| | | | 4 | 170760432 | rs77340396 | 0.18412 (7) | - | - | - | T | C | 4,42 | 1,34 | 4,01 | 1,56 | 5,63 | 3,00 | 2,97 | 0,70 | 4 % | 1,828 |
| | | | 4 | 170760580 | rs188030416 | 0.21214 (6) | - | - | - | A | G | 4,91 | 1,34 | 3,99 | 1,91 | 4,42 | 3,00 | 2,97 | 1,62 | 0 % | -1,331 |
| | | | 4 | 170760629 | rs72981901 | 0.18412 (7) | - | - | - | C | A | 3,56 | 1,34 | 3,98 | 1,31 | 7,02 | 3,00 | 2,97 | 1,39 | 0 % | 3,693 |
| | | | 4 | 170761207 | rs58390360 | 0.13454 (5) | - | - | - | A | C | 8,68 | 1,34 | 3,90 | 3,96 | 8,56 | 3,00 | 2,97 | 1,24 | 0 % | 3,693 |
| | | | 4 | 170761213 | - | - | - | - | - | C | A | 7,14 | 1,34 | 3,90 | 2,74 | 9,21 | 3,00 | 2,97 | 1,03 | 0 % | -1,175 |
| | | | 4 | 170761230 | rs771340747 | 0 (5) | - | - | - | C | T | 5,53 | 1,34 | 3,89 | 2,48 | 6,16 | 3,00 | 2,97 | 1,36 | 0 % | -1,098 |
| | | | 4 | 170761285 | rs116608709 | 0.28786 (6) | - | - | - | C | A | 3,23 | 1,34 | 3,89 | 0,99 | 5,65 | 3,00 | 2,97 | 1,42 | 2 % | 0,544 |
| | | | 4 | 170761294 | - | - | - | - | - | A | T | 1,07 | 1,34 | 3,88 | 0,82 | 0,69 | 3,00 | 2,97 | 1,11 | 0 % | -1,199 |
| | | | 4 | 170761335 | rs72696892 | 0.31452 (6) | *LINC024* | - | - | C | T | 1,10 | 1,34 | 3,88 | 0,55 | 1,31 | 3,00 | 2,97 | 1,10 | 1 % | 3,176 |
| | | | 4 | 170761366 | rs1412944629 | 0.55602 (6) | - | - | - | T | G | 8,32 | 1,34 | 3,87 | 0,52 | 6,63 | 3,00 | 2,97 | 1,45 | 0 % | -1,395 |
| | | | 4 | 170761407 | rs6816393 | 0 (6) | - | - | - | A | G | 9,49 | 1,34 | 3,87 | 0,63 | 7,18 | 3,00 | 2,97 | 1,65 | 4 % | 1,828 |
| | | | 4 | 170761506 | rs1735295056 | - | - | - | - | T | A | 9,21 | 1,34 | 3,85 | 1,87 | 10,56 | 3,00 | 2,97 | 1,83 | 0 % | 4,868 |

| | | | | | | | | | | | | | | | | | | | | | | | | | | | | | | | | | | | | |
|---|---|---|---|---|---|---|---|---|---|---|---|---|---|---|---|---|---|---|---|---|---|---|---|---|---|---|---|---|---|---|---|---|---|---|---|---|
| | | | 4 | 170761700 | rs10016324 | 0.13454 (5) | - | - | - | G | T | 1,40 | 1,34 | 3,83 | 4,37 | 1,46 | 3,00 | 2,97 | 1,07 | 5 % | 1,970 | 0,349 | 1,62E-08 | ≤3 | 52 | 4 % | 2,573 | [illegible] | [illegible] | [illegible] | [illegible] | [illegible] | [illegible] | [illegible] | [illegible] | [illegible] |
| | | | 4 | 170761796 | rs77679939 | 0.52984 (5) | - | - | - | T | C | 1,73 | 1,34 | 3,82 | 3,30 | 1,03 | 3,00 | 2,97 | 1,18 | 0 % | 3,693 | 2,255 | 1,02E-01 | - | ≤3 | - | - | [illegible] | [illegible] | [illegible] | [illegible] | [illegible] | [illegible] | [illegible] | [illegible] | [illegible] |
| | | | 4 | 170761969 | rs1020203326 | 0.2909 (6) | - | - | - | G | A | 1,51 | 3,99 | 3,79 | 6,35 | 1,90 | 3,00 | 2,97 | 1,40 | 0 % | -1,429 | 2,197 | 5,16E-01 | - | ≤3 | 0 % | -1,261 | [illegible] | [illegible] | [illegible] | [illegible] | [illegible] | [illegible] | [illegible] | [illegible] | [illegible] |
| MAF<5% | *LINC02382* | 4:170760001-170764000 | 4 | 170762039 | rs546575781 | 0.32023 (6) | - | - | - | A | T | 2,86 | 5,42 | 3,78 | 3,10 | 2,53 | 3,00 | 2,97 | 1,40 | 0 % | -1,117 | 3,354 | 7,39E-01 | - | ≤3 | 0 % | -1,074 | [illegible] | [illegible] | [illegible] | [illegible] | [illegible] | [illegible] | [illegible] | [illegible] | [illegible] |
| | | | 4 | 170762136 | rs188678575 | 0.18412 (7) | - | - | - | A | G | 2,10 | 5,42 | 3,77 | 3,81 | 1,99 | 3,00 | 2,97 | 0,96 | 0 % | -1,245 | 1,861 | 5,04E-01 | - | ≤3 | 0 % | -1,141 | 2,266 | 6,15E-01 | - | ≤3 | 0 % | -1,066 | 3,301 | 7,47E-01 | ≤3 |
| | | | 4 | 170762194 | rs76539211 | 0.18412 (7) | - | - | - | A | G | 2,87 | 5,42 | 3,76 | 3,25 | 2,66 | 3,00 | 2,97 | 1,54 | 1 % | 0,010 | 0,831 | 9,90E-01 | - | 9 | 1 % | -0,178 | 1,033 | 8,63E-01 | - | 8 | 1 % | -1,108 | 1,761 | 5,29E-01 | 7 |
| | | | 4 | 170762470 | rs568950756 | 0.18412 (7) | - | - | - | T | G | 0,57 | 1,34 | 3,73 | 1,19 | 0,96 | 3,00 | 2,97 | 0,93 | 0 % | -1,315 | 1,507 | 3,83E-01 | - | ≤3 | 0 % | -1,178 | 1,873 | 5,29E-01 | - | ≤3 | 0 % | -1,107 | 2,454 | 6,52E-01 | ≤3 |
| | | | 4 | 170762583 | rs184310675 | 0.18412 (7) | - | *MFAP3L* | - | G | T | 0,22 | 1,34 | 3,71 | 1,05 | 0,76 | 3,00 | 2,97 | 1,05 | 1 % | -0,058 | 0,676 | 9,32E-01 | - | 14 | 1 % | -0,553 | 0,862 | 5,21E-01 | - | 12 | 1 % | 1,475 | 1,148 | 1,99E-01 | 13 |
| | | | 4 | 170762595 | rs58855968 | 0.18412 (7) | - | - | - | T | C | 10,89 | 1,34 | 3,71 | 1,07 | 9,11 | 3,00 | 2,97 | 1,11 | 0 % | 3,693 | 2,255 | 1,02E-01 | - | ≤3 | - | - | - | - | - | - | - | - | - | - | - |
| | | | 4 | 170762698 | rs139565070 | 0.50466 (6) | - | - | - | T | C | 6,48 | 1,34 | 3,70 | 0,54 | 9,35 | 3,00 | 2,97 | 1,50 | 0 % | -1,120 | 3,341 | 7,38E-01 | - | ≤3 | 0 % | -1,061 | 4,539 | 8,15E-01 | - | ≤3 | 0 % | -1,038 | 6,032 | 8,63E-01 | ≤3 |
| | | | 4 | 170762856 | rs1274124147 | 0.2909 (6) | - | - | - | TAA | T | 4,13 | 6,45 | 3,68 | 4,69 | 6,72 | 3,00 | 2,97 | 1,23 | 0 % | 2,666 | 1,971 | 1,76E-01 | - | ≤3 | 0 % | -1,124 | 3,315 | 7,35E-01 | - | ≤3 | 0 % | -1,081 | 4,098 | 7,92E-01 | ≤3 |
| | | | 4 | 170763285 | rs62346544 | - | - | - | - | G | A | 1,05 | 3,99 | 3,62 | 1,91 | 1,79 | 3,00 | 2,97 | 1,03 | 0 % | -1,208 | 1,638 | 4,61E-01 | - | ≤3 | 0 % | -1,124 | 1,992 | 5,73E-01 | - | ≤3 | 0 % | -1,058 | 3,066 | 7,30E-01 | ≤3 |
| | | | 4 | 170763412 | rs185306197 | 0 (6) | - | - | - | T | A | 5,22 | 3,99 | 3,61 | 1,51 | 7,27 | 3,00 | 2,97 | 1,42 | 0 % | -1,215 | 1,951 | 5,34E-01 | - | ≤3 | 0 % | -1,115 | 2,485 | 6,54E-01 | - | ≤3 | 0 % | -1,056 | 3,507 | 7,63E-01 | ≤3 |
| | | | 4 | 170763536 | rs10012355 | 0.18412 (7) | - | - | - | A | T | 4,15 | 1,34 | 3,59 | 2,29 | 5,80 | 3,00 | 2,97 | 0,66 | 4 % | 1,828 | 0,380 | 1,48E-06 | ≤3 | 47 | 4 % | 2,558 | 0,486 | 1,38E-07 | ≤3 | 41 | 3 % | 0,529 | 0,911 | 5,62E-01 | 29 |
| | | | 4 | 170763560 | rs114170797 | 0.34125 (6) | - | - | - | A | T | 6,52 | 1,34 | 3,59 | 1,72 | 8,41 | 3,00 | 2,97 | 0,81 | 1 % | 1,077 | 0,775 | 1,65E-01 | - | 11 | 1 % | 1,670 | 0,979 | 8,81E-02 | - | 10 | 1 % | -1,101 | 1,721 | 5,22E-01 | 7 |
| | | | 4 | 170763581 | rs62346545 | 0.20285 (6) | - | - | - | C | T | 8,93 | 1,34 | 3,59 | 1,18 | 7,88 | 3,00 | 2,97 | 0,85 | 1 % | 0,312 | 0,905 | 7,30E-01 | - | 8 | 1 % | 0,059 | 1,130 | 9,58E-01 | - | 7 | 1 % | -1,077 | 1,951 | 5,81E-01 | 6 |
| | | | 4 | 170763813 | rs1431085476 | - | - | - | - | T | G | 2,47 | 3,99 | 3,56 | 2,27 | 2,49 | 3,00 | 2,97 | 1,47 | 0 % | 5,625 | 2,619 | 3,17E-02 | - | ≤3 | 0 % | 7,332 | 2,917 | 1,20E-02 | - | ≤3 | - | - | - | - | - |
| MAF<5% | *LINC02382* | 4:170766001-170770000 | 4 | 170766316 | rs10021720 | 0.42214 (6) | - | - | - | T | C | 8,06 | 6,10 | 3,29 | 2,50 | 10,56 | 3,00 | 2,97 | 2,33 | 4 % | 0,389 | 0,390 | 3,19E-01 | - | 46 | 4 % | 0,269 | 0,510 | 5,98E-01 | - | 40 | 4 % | 1,542 | 0,738 | 3,67E-02 | 39 |
| | | | 4 | 170766384 | rs562154999 | 0.18412 (7) | - | - | - | C | A | 8,92 | 6,00 | 3,28 | 5,86 | 5,99 | 3,00 | 2,97 | 2,97 | 0 % | 2,621 | 1,965 | 1,82E-01 | - | ≤3 | 0 % | 4,471 | 2,372 | 5,95E-02 | - | ≤3 | 0 % | -1,028 | 6,733 | 8,79E-01 | ≤3 |
| | | | 4 | 170766573 | rs183554664 | 0.18412 (7) | - | - | - | C | A | 0,05 | 7,64 | 3,26 | 3,12 | 0,10 | 3,00 | 2,97 | 1,71 | 0 % | -1,140 | 2,268 | 6,15E-01 | - | ≤3 | 0 % | -1,081 | 2,937 | 7,13E-01 | - | ≤3 | 0 % | -1,038 | 4,579 | 8,21E-01 | ≤3 |
| | | | 4 | 170766604 | rs28698569 | 0.18412 (7) | - | - | - | A | T | 7,91 | 7,00 | 3,26 | 2,67 | 8,35 | 3,00 | 2,97 | 1,17 | 4 % | 0,389 | 0,390 | 3,19E-01 | - | 46 | 4 % | 0,269 | 0,510 | 5,98E-01 | - | 40 | 4 % | 1,542 | 0,738 | 3,67E-02 | 39 |
| | | | 4 | 170766629 | - | - | - | - | - | C | T | 3,47 | 6,88 | 3,26 | 1,76 | 2,58 | 3,00 | 2,97 | 1,30 | 0 % | -1,280 | 2,475 | 6,05E-01 | - | ≤3 | 0 % | -1,127 | 3,324 | 7,35E-01 | - | ≤3 | 0 % | -1,084 | 4,150 | 7,94E-01 | ≤3 |
| | | | 4 | 170766872 | rs116755485 | 0.18412 (7) | - | - | - | A | G | 10,86 | 1,34 | 3,23 | 1,75 | 13,46 | 3,00 | 2,97 | 2,07 | 0 % | 2,223 | 1,862 | 2,33E-01 | - | ≤3 | 0 % | 3,708 | 2,204 | 9,25E-02 | - | ≤3 | 0 % | -1,066 | 4,468 | 8,11E-01 | ≤3 |
| | | | 4 | 170766940 | rs540832527 | 0.18412 (7) | - | - | - | A | G | 2,29 | 1,34 | 3,23 | 2,35 | 1,68 | 3,00 | 2,97 | 1,18 | 1 % | -0,370 | 0,974 | 7,04E-01 | - | 7 | 1 % | -1,190 | 1,292 | 3,57E-01 | - | 6 | 1 % | 1,512 | 1,619 | 3,50E-01 | 7 |
| | | | 4 | 170767096 | rs1297236206 | 0.58955 (5) | - | - | - | T | C | 1,06 | 3,99 | 3,21 | 2,18 | 0,84 | 3,00 | 2,97 | 1,70 | 0 % | 4,331 | 2,375 | 6,82E-02 | - | ≤3 | 0 % | 6,654 | 2,800 | 1,75E-02 | - | ≤3 | - | - | - | - | - |
| | | | 4 | 170767274 | rs188278143 | 0.13454 (5) | - | - | - | C | A | 0,45 | 7,29 | 3,20 | 8,57 | 0,91 | 3,00 | 2,97 | 2,17 | 1 % | -0,818 | 0,735 | 2,66E-01 | - | 12 | 1 % | -0,447 | 0,884 | 6,14E-01 | - | 12 | 1 % | -1,119 | 1,312 | 3,94E-01 | 11 |
| | | | 4 | 170767551 | rs778567298 | 0.18412 (7) | - | - | - | T | C | 3,19 | 1,34 | 3,17 | 2,11 | 1,96 | 3,00 | 2,97 | 1,85 | 0 % | 3,202 | 2,176 | 1,41E-01 | - | ≤3 | 0 % | 4,543 | 2,430 | 6,15E-02 | - | ≤3 | - | - | - | - | - |
| | | | 4 | 170767758 | rs116638566 | 0.18412 (7) | - | - | - | C | T | 3,04 | 1,34 | 3,15 | 0,58 | 4,07 | 3,00 | 2,97 | 1,16 | 1 % | 0,276 | 0,908 | 7,61E-01 | - | 8 | 1 % | -0,059 | 1,096 | 9,57E-01 | - | 7 | 1 % | 1,723 | 1,696 | 3,10E-01 | 7 |
| | | | 4 | 170767819 | rs73866743 | 0.18412 (7) | - | - | - | A | G | 0,21 | 1,34 | 3,15 | 0,64 | 0,03 | 3,00 | 2,97 | 1,59 | 4 % | 0,389 | 0,390 | 3,19E-01 | - | 46 | 4 % | 0,269 | 0,510 | 5,98E-01 | - | 40 | 4 % | 1,542 | 0,738 | 3,67E-02 | 39 |
| MAF<5% | *LINC02382* | 4:170766001-170770000, 4:170768001-170772000 | 4 | 170768175 | rs562764842 | 0.57429 (3a | - | - | - | A | G | 2,32 | 6,45 | 3,11 | 5,83 | 0,82 | 3,00 | 2,97 | 1,43 | 0 % | -1,137 | 3,145 | 7,18E-01 | - | ≤3 | 0 % | -1,089 | 3,794 | 7,74E-01 | - | ≤3 | 0 % | -1,030 | 6,668 | 8,77E-01 | ≤3 |
| | | | 4 | 170768386 | rs13435705 | 0.8051 (6) | - | - | - | C | A | 4,02 | 1,34 | 3,09 | 2,22 | 4,39 | 3,00 | 2,97 | 1,37 | 4 % | 0,389 | 0,390 | 3,19E-01 | - | 46 | 4 % | 0,269 | 0,510 | 5,98E-01 | - | 40 | 4 % | 1,542 | 0,738 | 3,67E-02 | 39 |
| | | | 4 | 170768533 | rs2140134 | 0.18347 (6) | - | - | - | G | A | 13,45 | 1,34 | 3,08 | 2,26 | 9,11 | 3,00 | 2,97 | 2,94 | 5 % | 1,970 | 0,349 | 1,62E-08 | ≤3 | 52 | 4 % | 2,573 | 0,473 | 5,27E-08 | ≤3 | 44 | 3 % | 1,762 | 0,852 | 3,86E-02 | 33 |
| | | | 4 | 170768590 | - | - | - | - | - | T | G | 1,86 | 1,34 | 3,08 | 1,79 | 1,99 | 3,00 | 2,97 | 2,19 | 0 % | -1,275 | 2,455 | 6,03E-01 | - | ≤3 | 0 % | -1,141 | 3,107 | 7,14E-01 | - | ≤3 | 0 % | -1,069 | 4,261 | 8,02E-01 | ≤3 |
| | | | 4 | 170769213 | rs1402294288 | 0.71614 (5) | - | - | - | G | A | 1,02 | 1,34 | 3,02 | 2,34 | 1,72 | 3,00 | 2,97 | 1,72 | 0 % | -1,216 | 2,724 | 6,55E-01 | - | ≤3 | 0 % | -1,093 | 3,892 | 7,79E-01 | - | ≤3 | 0 % | -1,068 | 4,803 | 8,24E-01 | ≤3 |
| | | | 4 | 170769371 | rs535477830 | 0 (5) | - | - | - | TAA | T | 1,51 | 1,34 | 3,01 | 2,99 | 3,36 | 3,00 | 2,97 | 2,48 | 0 % | -1,282 | 1,807 | 4,78E-01 | - | ≤3 | 0 % | -1,178 | 2,072 | 5,70E-01 | - | ≤3 | 0 % | -1,070 | 3,315 | 7,47E-01 | ≤3 |
| | | | 4 | 170769443 | rs1193054142 | - | - | - | - | T | C | 8,63 | 1,34 | 3,00 | 2,98 | 4,41 | 3,00 | 2,97 | 3,16 | 0 % | 3,519 | 2,219 | 1,13E-01 | - | ≤3 | 0 % | 5,429 | 2,581 | 3,55E-02 | - | ≤3 | - | - | - | - | - |
| | | | 4 | 170769507 | rs1735426267 | - | - | - | - | G | T | 0,51 | 1,34 | 3,00 | 2,18 | 1,57 | 3,00 | 2,97 | 3,46 | 0 % | -1,167 | 2,926 | 6,90E-01 | - | ≤3 | 0 % | -1,088 | 3,831 | 7,76E-01 | - | ≤3 | 0 % | -1,050 | 5,181 | 8,39E-01 | ≤3 |
| | | | 4 | 170769815 | rs559562424 | 0.4372 (6) | - | - | - | T | A | 0,09 | 5,42 | 2,97 | 2,45 | 0,31 | 3,00 | 2,97 | 4,71 | 0 % | -1,391 | 2,236 | 5,34E-01 | - | ≤3 | 0 % | -1,229 | 2,593 | 6,36E-01 | - | ≤3 | 0 % | -1,096 | 3,627 | 7,63E-01 | ≤3 |
| MAF<5% | *LINC02382* | 4:170768001-170772000, 4:170770001-170774000 | 4 | 170770374 | rs10004230 | 0.31452 (6) | - | - | - | T | C | 7,54 | 3,99 | 2,93 | 3,20 | 6,18 | 3,00 | 2,97 | 2,04 | 5 % | 1,970 | 0,349 | 1,62E-08 | ≤3 | 52 | 4 % | 2,573 | 0,473 | 5,27E-08 | ≤3 | 44 | 3 % | 1,762 | 0,852 | 3,86E-02 | 33 |
| | | | 4 | 170770422 | rs541864333 | - | - | - | - | A | G | 2,76 | 3,99 | 2,92 | 5,22 | 1,96 | 3,00 | 2,97 | 2,38 | 0 % | -1,462 | 2,178 | 5,02E-01 | - | ≤3 | 0 % | -1,330 | 2,367 | 5,74E-01 | - | ≤3 | 0 % | -1,105 | 3,668 | 7,63E-01 | ≤3 |
| | | | 4 | 170770755 | rs183755902 | - | - | - | - | A | C | 0,59 | 5,42 | 2,90 | 2,16 | 1,37 | 3,00 | 2,97 | 1,00 | 0 % | -1,170 | 2,150 | 5,86E-01 | - | ≤3 | 0 % | -1,138 | 2,422 | 6,39E-01 | - | ≤3 | 0 % | -1,045 | 4,548 | 8,18E-01 | ≤3 |
| | | | 4 | 170770995 | rs72696893 | 0.13454 (5) | *LINC024…* | - | - | T | C | 0,07 | 1,34 | 2,88 | 4,93 | 0,18 | 3,00 | 2,97 | 1,75 | 1 % | 3,176 | 0,950 | 8,28E-04 | - | 7 | 0 % | 2,354 | 1,886 | 2,12E-01 | - | ≤3 | 0 % | 9,278 | 2,276 | 4,58E-05 | 4 |
| | | | 4 | 170771046 | rs140904005 | 0.32576 (5) | - | - | - | T | C | 4,06 | 1,34 | 2,88 | 2,52 | 3,91 | 3,00 | 2,97 | 1,36 | 2 % | 0,048 | 0,528 | 9,28E-01 | - | 25 | 2 % | 0,421 | 0,644 | 5,14E-01 | - | 24 | 2 % | -1,123 | 1,052 | 2,86E-01 | 20 |
| | | | 4 | 170771078 | rs185235422 | 0.3882 (5) | - | - | - | T | C | 5,12 | 1,34 | 2,87 | 0,91 | 4,75 | 3,00 | 2,97 | 1,59 | 0 % | 2,360 | 1,895 | 2,13E-01 | - | ≤3 | 0 % | 3,682 | 2,201 | 9,44E-02 | - | ≤3 | 0 % | -1,059 | 4,839 | 8,27E-01 | ≤3 |
| | | | 4 | 170771148 | rs78567784 | 0.13454 (5) | - | - | - | C | T | 1,93 | 1,34 | 2,87 | 1,11 | 4,15 | 3,00 | 2,97 | 1,30 | 0 % | 3,693 | 2,255 | 1,02E-01 | - | ≤3 | - | - | - | - | - | - | - | - | - | - | - |
| | | | 4 | 170771170 | rs147909608 | 0.14427 (5) | - | - | - | A | G | 5,59 | 1,34 | 2,87 | 1,10 | 3,49 | 3,00 | 2,97 | 0,89 | 0 % | 3,367 | 2,133 | 1,15E-01 | - | ≤3 | 0 % | 5,011 | 2,481 | 4,34E-02 | - | ≤3 | 0 % | -1,048 | 5,578 | 8,51E-01 | ≤3 |
| | | | 4 | 170771408 | rs189249515 | 0.13454 (5) | - | - | - | G | T | 0,65 | 1,34 | 2,85 | 2,52 | 1,42 | 3,00 | 2,97 | 1,89 | 0 % | -1,264 | 1,527 | 4,08E-01 | - | ≤3 | 0 % | -1,167 | 1,832 | 5,24E-01 | - | ≤3 | 0 % | -1,066 | 2,752 | 6,99E-01 | ≤3 |
| | | | 4 | 170771544 | - | - | - | - | - | C | CAC… | 2,00 | 3,99 | 2,84 | 3,78 | 3,00 | 3,00 | 2,97 | 2,11 | 4 % | 1,828 | 0,380 | 1,48E-06 | ≤3 | 47 | 4 % | 2,558 | 0,486 | 1,38E-07 | ≤3 | 41 | 3 % | 0,529 | 0,911 | 5,62E-01 | 29 |
| | | | 4 | 170771590 | rs566113215 | 0 (6) | - | - | - | GT | G | 0,48 | 1,34 | 2,83 | 2,14 | 0,96 | 3,00 | 2,97 | 2,00 | 1 % | -1,319 | 1,030 | 2,00E-01 | - | 6 | 1 % | -1,190 | 1,274 | 3,50E-01 | - | 6 | 1 % | -1,103 | 1,705 | 5,18E-01 | 6 |
| MAF<5% | *LINC02382* | 4:170770001-170774000 | 4 | 170772031 | rs1579113631 | - | - | - | - | A | G | 6,38 | 1,34 | 2,80 | 1,02 | 2,83 | 3,00 | 2,97 | 3,18 | 0 % | -1,179 | 2,832 | 6,77E-01 | - | ≤3 | 0 % | -1,120 | 3,328 | 7,37E-01 | - | ≤3 | 0 % | -1,044 | 5,402 | 8,47E-01 | ≤3 |
| | | | 4 | 170772102 | rs527869746 | 0.42619 (6) | - | - | - | G | GA | 5,35 | 1,34 | 2,80 | 0,57 | 9,05 | 3,00 | 2,97 | 2,97 | 1 % | 3,176 | 0,950 | 8,28E-04 | - | 7 | 0 % | 2,354 | 1,886 | 2,12E-01 | - | ≤3 | 0 % | 9,278 | 2,276 | 4,58E-05 | 4 |

| | | | | | | | | | | | | | | | | | | | | | | | | | | | | | | | | | | | | | | | | | | | | | | | | | | | | | | | | | | | | | | | | | | | | | | | | | |
|---|---|---|---|---|---|---|---|---|---|---|---|---|---|---|---|---|---|---|---|---|---|---|---|---|---|---|---|---|---|---|---|---|---|---|---|---|---|---|---|---|---|---|---|---|---|---|---|---|---|---|---|---|---|---|---|---|---|---|---|---|---|---|---|---|---|---|---|---|---|---|---|---|---|
| | | | 4 | 170787356 | rs540888361 | 0.18412 (7) | - | - | - | C | CCA | 1,74 | 1,34 | 2,94 | 0,49 | 2,31 | 3,00 | 2,97 | 2,11 | 4 % | 1,719 | 0,395 | 1,32E-05 | ≤3 | 38 | 3 % | 2,227 | 0,555 | 6,09E-05 | ≤3 | 32 | 3 % | 1,548 | 0,944 | 1,01E-01 | - | 26 | 4,6 % | -0,091 | 6,370 | 5,60E-01 | 8 | 353 | 4,6 % | -0,128 | 4,949 | 5,25E-01 | 8 | 334 | 4,6 % | 0,012 | 2,814 | 9,72E-01 | 8 | 326 | 4,6 % | 0,035 | 0,031 | 2,59E-01 | 4,6 % | 0,050 | 0,032 | 1,15E-01 | 4,6 % | 0,035 | 0,033 | 2,95E-01 | 4,6 % | -0,006 | 0,061 | 9,27E-01 |
| | | | 4 | 170787474 | rs377000943 | 0.18412 (7) | - | - | - | A | C | 3,42 | 1,34 | 2,95 | 0,33 | 2,31 | 3,00 | 2,97 | 2,58 | 0 % | -1,174 | 2,869 | 6,82E-01 | - | ≤3 | 0 % | -1,120 | 3,334 | 7,37E-01 | - | ≤3 | 0 % | -1,038 | 5,826 | 8,59E-01 | - | ≤3 | - | - | - | - | - | - | - | - | - | - | - | - | - | - | - | - | - | - | - | - | - | - | - | - | - | - | - | - | - | - | - | - | - | - |
| | | | 4 | 170787525 | rs1256975703 | 0.80633 (6) | - | - | - | C | T | 2,59 | 1,34 | 2,96 | 0,07 | 2,31 | 3,00 | 2,97 | 2,89 | 0 % | -1,116 | 3,360 | 7,40E-01 | - | ≤3 | 0 % | -1,058 | 4,598 | 8,18E-01 | - | ≤3 | 0 % | -1,032 | 6,393 | 8,72E-01 | - | ≤3 | - | - | - | - | - | - | - | - | - | - | - | - | - | - | - | - | - | - | - | - | - | - | - | - | - | - | - | - | - | - | - | - | - | - |
| | | | 4 | 170787601 | rs1217502911 | 0.18412 (7) | - | - | - | C | A | 2,40 | 1,34 | 2,96 | 0,22 | 2,31 | 3,00 | 2,97 | 5,29 | 1 % | -0,102 | 1,078 | 9,24E-01 | - | 6 | 1 % | 0,614 | 1,331 | 6,45E-01 | - | 6 | 1 % | -1,074 | 2,130 | 6,14E-01 | - | 5 | - | - | - | - | - | - | - | - | - | - | - | - | - | - | - | - | - | - | - | - | - | - | - | - | - | - | - | - | - | - | - | - | - | - |
| | | | 4 | 170787618 | rs1244139676 | 0.18412 (7) | - | - | - | G | A | 2,31 | 1,34 | 2,96 | 0,20 | 2,31 | 3,00 | 2,97 | 6,49 | 1 % | -1,245 | 0,947 | 1,89E-01 | - | 9 | 1 % | -1,143 | 1,197 | 3,40E-01 | - | 9 | 1 % | -1,090 | 1,681 | 5,17E-01 | - | 9 | - | - | - | - | - | - | - | - | - | - | - | - | - | - | - | - | - | - | - | - | - | - | - | - | - | - | - | - | - | - | - | - | - | - |
| | | | 4 | 170787748 | rs375843326 | 0.55602 (6) | - | - | - | C | T | 5,36 | 1,34 | 2,97 | 0,22 | 2,31 | 3,00 | 2,97 | 10,68 | 2 % | 1,872 | 0,540 | 5,33E-04 | - | 24 | 2 % | 2,322 | 0,712 | 1,10E-03 | - | 20 | 1 % | -1,101 | 1,446 | 4,46E-01 | - | 13 | - | - | - | - | - | - | - | - | - | - | - | - | - | - | - | - | - | - | - | - | - | - | - | - | - | - | - | - | - | - | - | - | - | - |
| | | | 4 | 170787749 | rs186319670 | 0.00083 (6) | - | - | - | G | A | 4,95 | 1,34 | 2,97 | 0,22 | 2,31 | 3,00 | 2,97 | 10,68 | 4 % | 0,389 | 0,390 | 3,19E-01 | - | 46 | 4 % | 0,269 | 0,510 | 5,98E-01 | - | 40 | 4 % | 1,542 | 0,738 | 3,67E-02 | - | 39 | 3,9 % | -0,003 | 5,878 | 9,85E-01 | 5 | 283 | 3,9 % | 0,368 | 4,604 | 9,07E-02 | 5 | 274 | 3,9 % | 0,220 | 2,585 | 5,69E-01 | 4 | 262 | 4,1 % | 0,021 | 0,032 | 5,12E-01 | 4,1 % | 0,016 | 0,033 | 6,38E-01 | 4,1 % | 0,015 | 0,034 | 6,55E-01 | 4,1 % | 0,044 | 0,064 | 4,93E-01 |
| | | | 4 | 170787869 | rs1431098100 | 0.01899 (6) | - | - | - | A | T | 5,83 | 1,34 | 2,98 | 0,14 | 2,31 | 3,00 | 2,97 | 11,27 | 1 % | -0,541 | 0,626 | 3,88E-01 | - | 17 | 2 % | -0,630 | 0,761 | 4,07E-01 | - | 16 | 2 % | -1,104 | 1,164 | 3,43E-01 | - | 15 | - | - | - | - | - | - | - | - | - | - | - | - | - | - | - | - | - | - | - | - | - | - | - | - | - | - | - | - | - | - | - | - | - | - |
| | | | 4 | 170787902 | rs1407561532 | 0.11093 (6) | - | - | - | A | G | 4,85 | 1,34 | 2,99 | 0,21 | 2,31 | 3,00 | 2,97 | 10,40 | 0 % | -1,137 | 3,145 | 7,18E-01 | - | ≤3 | 0 % | -1,089 | 3,794 | 7,74E-01 | - | ≤3 | 0 % | -1,030 | 6,668 | 8,77E-01 | - | ≤3 | - | - | - | - | - | - | - | - | - | - | - | - | - | - | - | - | - | - | - | - | - | - | - | - | - | - | - | - | - | - | - | - | - | - |
| | | | 4 | 170787970 | rs181802136 | 0.18412 (7) | - | - | - | G | A | 2,66 | 1,34 | 2,99 | 0,34 | 2,31 | 3,00 | 2,97 | 6,96 | 4 % | 0,389 | 0,390 | 3,19E-01 | - | 46 | 4 % | 0,269 | 0,510 | 5,98E-01 | - | 40 | 4 % | 1,542 | 0,738 | 3,67E-02 | - | 39 | 4,2 % | -0,051 | 6,139 | 7,53E-01 | 5 | 309 | 4,2 % | 0,305 | 4,814 | 1,42E-01 | 5 | 300 | 4,2 % | 0,153 | 2,707 | 6,79E-01 | 4 | 287 | 4,3 % | 0,030 | 0,031 | 3,26E-01 | 4,3 % | 0,023 | 0,032 | 4,64E-01 | 4,3 % | 0,026 | 0,033 | 4,32E-01 | 4,3 % | 0,045 | 0,062 | 4,69E-01 |
| MAF<5% | *LINC02382* | 4:170786001-170790000, 4:170788001-170792000 | 4 | 170788002 | rs146327279 | 0.34847 (6) | - | - | - | G | C | 2,01 | 1,34 | 2,99 | 0,37 | 2,31 | 3,00 | 2,97 | 8,02 | 5 % | 1,970 | 0,349 | 1,62E-08 | ≤3 | 52 | 4 % | 2,573 | 0,473 | 5,27E-08 | ≤3 | 44 | 3 % | 1,762 | 0,852 | 3,86E-02 | - | 33 | 5,3 % | -0,116 | 7,052 | 4,12E-01 | 11 | 402 | 5,3 % | -0,113 | 5,491 | 5,33E-01 | 11 | 383 | 5,4 % | -0,042 | 3,121 | 8,96E-01 | 11 | 372 | 5,3 % | 0,030 | 0,028 | 2,86E-01 | 5,3 % | 0,049 | 0,029 | 9,47E-02 | 5,3 % | 0,031 | 0,030 | 3,16E-01 | 5,2 % | 0,004 | 0,056 | 9,42E-01 |
| | | | 4 | 170788018 | rs1323230347 | 0.18412 (7) | - | - | - | C | T | 3,64 | 1,34 | 3,00 | 0,37 | 2,31 | 3,00 | 2,97 | 7,98 | 1 % | 0,095 | 0,841 | 9,10E-01 | - | 9 | 1 % | 0,770 | 0,992 | 4,38E-01 | - | 9 | 1 % | -1,114 | 1,601 | 4,87E-01 | - | 7 | - | - | - | - | - | - | - | - | - | - | - | - | - | - | - | - | - | - | - | - | - | - | - | - | - | - | - | - | - | - | - | - | - | - |
| | | | 4 | 170788081 | rs368809151 | 0.18412 (7) | *LINC024* | - | - | A | C | 6,19 | 1,34 | 3,00 | 0,29 | 2,31 | 3,00 | 2,97 | 11,25 | 1 % | 2,812 | 1,020 | 5,82E-03 | - | 6 | 0 % | 2,354 | 1,886 | 2,12E-01 | - | ≤3 | 0 % | 9,278 | 2,276 | 4,58E-05 | - | 4 | 0,7 % | -0,150 | 2,583 | 6,99E-01 | - | 55 | 0,6 % | -0,740 | 1,980 | 1,43E-01 | - | 51 | 0,7 % | -0,632 | 1,143 | 4,70E-01 | - | 51 | 0,7 % | 0,093 | 0,078 | 2,33E-01 | 0,7 % | 0,110 | 0,080 | 1,71E-01 | 0,7 % | 0,072 | 0,084 | 3,89E-01 | 0,7 % | 0,095 | 0,156 | 5,42E-01 |
| | | | 4 | 170788272 | rs1326843031 | 0.15359 (6) | - | - | - | G | C | 3,13 | 1,34 | 3,02 | 0,28 | 2,31 | 3,00 | 2,97 | 6,33 | 0 % | 5,150 | 2,532 | 4,19E-02 | - | ≤3 | - | - | - | - | - | - | - | - | - | - | - | - | - | - | - | - | - | - | - | - | - | - | - | - | - | - | - | - | - | - | - | - | - | - | - | - | - | - | - | - | - | - | - | - | - | - | - | - |
| | | | 4 | 170788361 | rs10013912 | 0.20829 (6) | - | - | - | C | T | 4,50 | 1,34 | 3,02 | 0,27 | 2,31 | 3,00 | 2,97 | 7,12 | 4 % | 1,828 | 0,380 | 1,48E-06 | ≤3 | 47 | 4 % | 2,558 | 0,486 | 1,38E-07 | ≤3 | 41 | 3 % | 0,529 | 0,911 | 5,62E-01 | - | 29 | 4,7 % | -0,111 | 6,565 | 4,66E-01 | 6 | 357 | 4,7 % | -0,020 | 5,120 | 9,19E-01 | 6 | 342 | 4,7 % | 0,049 | 2,905 | 8,86E-01 | 6 | 331 | 4,6 % | 0,020 | 0,030 | 4,98E-01 | 4,6 % | 0,039 | 0,031 | 2,11E-01 | 4,6 % | 0,024 | 0,033 | 4,62E-01 | 4,6 % | -0,009 | 0,060 | 8,75E-01 |
| | | | 4 | 170788367 | rs9990639 | 0.18412 (7) | - | - | - | G | A | 4,53 | 1,34 | 3,03 | 0,27 | 2,31 | 3,00 | 2,97 | 6,38 | 4 % | 0,389 | 0,390 | 3,19E-01 | - | 46 | 4 % | 0,269 | 0,510 | 5,98E-01 | - | 40 | 4 % | 1,542 | 0,738 | 3,67E-02 | - | 39 | 4,2 % | -0,051 | 6,139 | 7,52E-01 | 5 | 309 | 4,2 % | 0,305 | 4,814 | 1,42E-01 | 5 | 300 | 4,2 % | 0,153 | 2,707 | 6,79E-01 | 4 | 287 | 4,3 % | 0,030 | 0,031 | 3,26E-01 | 4,3 % | 0,023 | 0,032 | 4,63E-01 | 4,3 % | 0,026 | 0,033 | 4,32E-01 | 4,3 % | 0,045 | 0,062 | 4,69E-01 |
| | | | 4 | 170788601 | rs1053018603 | 0.58955 (5) | - | - | - | C | G | 4,38 | 1,34 | 3,05 | 0,62 | 2,31 | 3,00 | 2,97 | 1,77 | 0 % | -1,167 | 2,918 | 6,89E-01 | - | ≤3 | 0 % | -1,095 | 3,699 | 7,67E-01 | - | ≤3 | 0 % | -1,043 | 5,560 | 8,51E-01 | - | ≤3 | 0,1 % | -0,161 | 0,865 | 8,89E-01 | - | 6 | 0,1 % | -1,432 | 0,629 | 3,68E-01 | - | 5 | 0,1 % | 8,219 | 0,398 | 1,07E-03 | - | 6 | 0,0 % | -0,398 | 0,322 | 2,16E-01 | 0,0 % | -0,337 | 0,334 | 3,12E-01 | 0,0 % | -0,336 | 0,351 | 3,39E-01 | 0,0 % | -0,717 | 0,617 | 2,45E-01 |
| | | | 4 | 170788635 | rs185230497 | 0.60906 (4) | - | - | - | C | T | 4,24 | 1,34 | 3,05 | 0,61 | 2,31 | 3,00 | 2,97 | 1,55 | 2 % | 0,868 | 0,478 | 6,92E-02 | - | 28 | 2 % | 0,796 | 0,640 | 2,14E-01 | - | 23 | 2 % | 2,239 | 0,920 | 1,50E-02 | - | 22 | 2,8 % | 0,027 | 5,040 | 8,93E-01 | ≤3 | 205 | 2,8 % | 0,387 | 3,944 | 1,27E-01 | ≤3 | 198 | 2,8 % | 0,549 | 2,213 | 2,24E-01 | ≤3 | 190 | 2,9 % | 0,002 | 0,037 | 9,53E-01 | 2,9 % | -0,009 | 0,038 | 8,05E-01 | 2,9 % | -0,013 | 0,040 | 7,38E-01 | 2,9 % | 0,073 | 0,074 | 3,30E-01 |
| | | | 4 | 170788649 | rs181822876 | - | - | - | - | C | T | 3,68 | 1,34 | 3,05 | 0,77 | 2,31 | 3,00 | 2,97 | 1,52 | 0 % | 1,554 | 1,723 | 3,67E-01 | - | ≤3 | 0 % | 2,574 | 1,965 | 1,90E-01 | - | ≤3 | 0 % | -1,044 | 5,470 | 8,49E-01 | - | ≤3 | - | - | - | - | - | - | - | - | - | - | - | - | - | - | - | - | - | - | - | - | - | - | - | - | - | - | - | - | - | - | - | - | - | - |
| | | | 4 | 170788782 | rs150571420 | 0.60906 (4) | - | - | - | G | A | 2,81 | 1,34 | 3,06 | 0,43 | 2,31 | 3,00 | 2,97 | 0,64 | 4 % | 1,777 | 0,385 | 3,88E-06 | ≤3 | 46 | 4 % | 2,558 | 0,486 | 1,38E-07 | ≤3 | 41 | 3 % | -0,225 | 0,948 | 8,13E-01 | - | 28 | 4,7 % | -0,111 | 6,565 | 4,66E-01 | 6 | 357 | 4,7 % | -0,020 | 5,120 | 9,19E-01 | 6 | 342 | 4,7 % | 0,049 | 2,905 | 8,86E-01 | 6 | 331 | 4,6 % | 0,020 | 0,030 | 4,98E-01 | 4,6 % | 0,039 | 0,031 | 2,11E-01 | 4,6 % | 0,024 | 0,033 | 4,62E-01 | 4,6 % | -0,009 | 0,060 | 8,75E-01 |
| | | | 4 | 170788783 | rs1309535225 | 0.60906 (4) | - | - | - | A | G | 4,07 | 1,34 | 3,06 | 0,43 | 2,31 | 3,00 | 2,97 | 0,52 | 0 % | -1,117 | 3,354 | 7,39E-01 | - | ≤3 | 0 % | -1,074 | 4,148 | 7,96E-01 | - | ≤3 | 0 % | -1,029 | 6,833 | 8,80E-01 | - | ≤3 | 0,1 % | 0,657 | 1,220 | 4,23E-01 | - | 12 | 0,1 % | -0,848 | 0,887 | 4,52E-01 | - | 10 | 0,1 % | -0,789 | 0,511 | 6,87E-01 | - | 10 | 0,1 % | -0,031 | 0,224 | 8,92E-01 | 0,1 % | 0,055 | 0,232 | 8,11E-01 | 0,1 % | 0,082 | 0,244 | 7,36E-01 | 0,1 % | -0,643 | 0,448 | 1,51E-01 |
| | | | 4 | 170788790 | rs140483240 | 0.70302 (3a | - | - | - | G | T | 1,96 | 1,34 | 3,06 | 0,42 | 2,31 | 3,00 | 2,97 | 0,46 | 0 % | 3,693 | 2,255 | 1,02E-01 | - | ≤3 | - | - | - | - | - | - | - | - | - | - | - | - | 0,1 % | -0,875 | 1,110 | 3,32E-01 | - | 10 | 0,1 % | -0,766 | 0,883 | 4,99E-01 | - | 10 | 0,1 % | -0,691 | 0,511 | 7,24E-01 | - | 10 | 0,1 % | -0,119 | 0,173 | 4,90E-01 | 0,1 % | -0,075 | 0,178 | 6,72E-01 | 0,1 % | 0,002 | 0,185 | 9,93E-01 | 0,1 % | -0,695 | 0,361 | 5,44E-02 |
| | | | 4 | 170788804 | rs531396986 | 0.1525 (3a) | - | - | - | C | G | 1,85 | 1,34 | 3,06 | 0,39 | 2,31 | 3,00 | 2,97 | 0,37 | 0 % | -1,139 | 3,135 | 7,16E-01 | - | ≤3 | 0 % | -1,091 | 3,788 | 7,73E-01 | - | ≤3 | 0 % | -1,030 | 6,716 | 8,78E-01 | - | ≤3 | - | - | - | - | - | - | - | - | - | - | - | - | - | - | - | - | - | - | - | - | - | - | - | - | - | - | - | - | - | - | - | - | - | - |
| | | | 4 | 170788833 | rs191650202 | 0.58955 (5) | - | - | - | G | A | 2,25 | 1,34 | 3,07 | 0,36 | 2,31 | 3,00 | 2,97 | 0,39 | 0 % | 2,360 | 1,895 | 2,13E-01 | - | ≤3 | 0 % | 3,682 | 2,201 | 9,44E-02 | - | ≤3 | 0 % | -1,059 | 4,839 | 8,27E-01 | - | ≤3 | 0,5 % | -0,386 | 2,113 | 4,15E-01 | - | 36 | 0,5 % | 0,396 | 1,682 | 5,06E-01 | - | 36 | 0,5 % | -0,919 | 0,932 | 3,92E-01 | - | 33 | 0,5 % | -0,177 | 0,087 | 4,35E-02 | 0,5 % | -0,182 | 0,090 | 4,43E-02 | 0,5 % | -0,187 | 0,095 | 4,79E-02 | 0,5 % | -0,147 | 0,172 | 3,92E-01 |
| | | | 4 | 170789043 | rs183483907 | 0.3538 (3a) | - | - | - | G | T | 1,48 | 6,45 | 3,08 | 1,72 | 4,05 | 3,00 | 2,97 | 1,15 | 3 % | 0,866 | 0,497 | 8,15E-02 | - | 29 | 2 % | 0,954 | 0,660 | 1,48E-01 | - | 25 | 2 % | 1,563 | 0,950 | 9,98E-02 | - | 23 | 2,1 % | -0,089 | 4,420 | 6,93E-01 | ≤3 | 158 | 2,0 % | -0,517 | 3,400 | 7,88E-02 | ≤3 | 147 | 2,1 % | 0,155 | 1,961 | 7,61E-01 | ≤3 | 147 | 2,2 % | 0,058 | 0,043 | 1,80E-01 | 2,2 % | 0,067 | 0,045 | 1,30E-01 | 2,2 % | 0,061 | 0,047 | 1,93E-01 | 2,2 % | 0,056 | 0,086 | 5,15E-01 |
| | | | 4 | 170789426 | rs149229838 | 0.03222 (5) | *AADAT (* | - | - | A | G | 5,23 | 3,99 | 3,12 | 1,43 | 3,14 | 3,00 | 2,97 | 2,69 | 1 % | 0,122 | 0,709 | 8,64E-01 | - | 14 | 1 % | -0,437 | 0,910 | 6,31E-01 | - | 12 | 1 % | 0,633 | 1,330 | 6,34E-01 | - | 12 | 0,9 % | -0,381 | 2,913 | 2,67E-01 | ≤3 | 68 | 0,9 % | -0,140 | 2,289 | 7,49E-01 | ≤3 | 66 | 0,9 % | -0,796 | 1,295 | 3,03E-01 | ≤3 | 63 | 0,8 % | 0,013 | 0,071 | 8,51E-01 | 0,8 % | -0,018 | 0,073 | 8,11E-01 | 0,8 % | -0,026 | 0,077 | 7,33E-01 | 0,8 % | 0,206 | 0,141 | 1,43E-01 |
| | | | 4 | 170789747 | rs28380441 | 0 (6) | - | - | - | T | C | 4,41 | 1,34 | 3,15 | 2,43 | 4,47 | 3,00 | 2,97 | 1,30 | 5 % | 1,970 | 0,349 | 1,62E-08 | ≤3 | 52 | 4 % | 2,573 | 0,473 | 5,27E-08 | ≤3 | 44 | 3 % | 1,762 | 0,852 | 3,86E-02 | - | 33 | 5,3 % | -0,116 | 7,052 | 4,13E-01 | 11 | 402 | 5,3 % | -0,113 | 5,490 | 5,34E-01 | 11 | 383 | 5,4 % | -0,042 | 3,121 | 8,96E-01 | 11 | 372 | 5,3 % | 0,030 | 0,028 | 2,86E-01 | 5,3 % | 0,049 | 0,029 | 9,47E-02 | 5,3 % | 0,031 | 0,030 | 3,16E-01 | 5,2 % | 0,004 | 0,056 | 9,43E-01 |
| | | | 4 | 170789973 | rs141868957 | 0.18412 (7) | - | - | - | A | C | 3,58 | 1,34 | 3,17 | 1,74 | 4,80 | 3,00 | 2,97 | 1,47 | 1 % | 0,276 | 0,908 | 7,61E-01 | - | 8 | 1 % | -0,059 | 1,096 | 9,57E-01 | - | 7 | 1 % | 1,723 | 1,696 | 3,10E-01 | - | 7 | 0,6 % | -0,219 | 2,464 | 5,90E-01 | - | 52 | 0,6 % | -0,439 | 1,913 | 4,01E-01 | - | 49 | 0,6 % | -0,432 | 1,095 | 6,36E-01 | - | 48 | 0,7 % | 0,061 | 0,078 | 4,30E-01 | 0,7 % | 0,074 | 0,080 | 3,55E-01 | 0,7 % | 0,012 | 0,084 | 8,90E-01 | 0,7 % | 0,208 | 0,155 | 1,81E-01 |
| [illegible] | | | | | | | | | | | | | | | | | | | | | | | | | | | | | | | | | | | | | | | | | | | | | | | | | | | | | | | | | | | | | | | | | | | | | | | | | |

| | | | | | | | | | | | | | | | | | | | | | | | |
|---|---|---|---|---|---|---|---|---|---|---|---|---|---|---|---|---|---|---|---|---|---|---|---|
| | | | 4 | 170813751 | rs149666742 | 0.2212 (6) | - | - | - | A | AT | 1,46 | 1,34 | 14,65 | 2,07 | 4,23 | 3,00 | 2,97 | 2,74 | 1 % | 3,092 | 1,049 | [illegible] |
| | | | 4 | 170813781 | rs72984170 | 0.58955 (5) | - | - | - | C | A | 1,47 | 3,99 | 14,85 | 3,17 | 1,42 | 3,00 | 2,97 | 2,82 | 1 % | -0,558 | 0,932 | [illegible] |
| | | | 4 | 170813900 | rs142757933 | 0.60906 (4) | - | - | - | A | G | 6,28 | 5,42 | 15,81 | 5,82 | 10,03 | 3,00 | 2,97 | 2,39 | 0 % | -1,160 | 2,966 | [illegible] |
| | | | 4 | 170813935 | rs62344263 | 0.75409 (3a | - | - | - | T | A | 3,58 | 3,99 | 16,14 | 5,89 | 6,53 | 3,00 | 2,97 | 1,98 | 4 % | 0,483 | 0,366 | [illegible] |
| MAF<5% | LINC02382 | 4:170812001-170816000, | 4 | 170814235 | rs80268011 | 0.18412 (7) | - | - | - | C | T | 6,61 | 1,34 | 21,62 | 4,87 | 9,64 | 3,00 | 2,97 | 2,11 | 4 % | 1,828 | 0,380 | [illegible] |
| | | 4:170814001-170818000 | 4 | 170814460 | rs17056210 | 0.13454 (5) | - | - | - | C | T | 3,48 | 1,34 | 25,12 | 7,07 | 3,10 | 3,00 | 2,97 | 2,42 | 1 % | -0,558 | 0,932 | [illegible] |
| | | | 4 | 170814527 | rs1197911746 | 0.13454 (5) | - | - | - | A | G | 2,79 | 1,34 | 28,32 | 4,01 | 3,01 | 3,00 | 2,97 | 1,45 | 0 % | -1,139 | 3,104 | [illegible] |
| | | | 4 | 170814555 | rs79138394 | 0.22339 (6) | - | - | - | T | G | 3,48 | 1,34 | 32,13 | 3,84 | 3,71 | 3,00 | 2,97 | 1,24 | 0 % | -1,208 | 1,638 | [illegible] |
| | | | 4 | 170814578 | rs1331123101 | 0.18412 (7) | - | - | - | C | T | 1,83 | 1,34 | 26,49 | 4,50 | 1,66 | 3,00 | 2,97 | 1,15 | 0 % | 3,693 | 2,255 | [illegible] |
| | | | 4 | 170814600 | rs28534908 | 0.34523 (6) | - | - | - | A | G | 0,83 | 1,34 | 24,43 | 3,19 | 1,03 | 3,00 | 2,97 | 1,73 | 4 % | 0,483 | 0,366 | [illegible] |
| | | | 4 | 170814695 | rs754483226 | 0.2212 (6) | - | - | - | G | T | 1,70 | 1,34 | 20,34 | 2,33 | 2,14 | 3,00 | 2,97 | 2,02 | 0 % | 4,499 | 2,407 | [illegible] |
| | | | 4 | 170814791 | rs17056213 | 0.18412 (7) | - | - | - | T | C | 3,50 | 1,34 | 18,29 | 2,79 | 2,68 | 3,00 | 2,97 | 1,54 | 5 % | 1,921 | 0,352 | [illegible] |
| | | | 4 | 170815003 | rs28646080 | 0.14464 (6) | - | - | - | A | G | 2,36 | 1,34 | 15,69 | 1,89 | 1,60 | 3,00 | 2,97 | 1,23 | 5 % | 1,921 | 0,352 | [illegible] |
| | | | 4 | 170815112 | rs78042725 | 0.18412 (7) | - | - | - | G | A | 6,98 | 1,34 | 14,83 | 5,12 | 9,27 | 3,00 | 2,97 | 0,95 | 1 % | 0,275 | 0,648 | [illegible] |
| | | | 4 | 170815217 | rs189064370 | 0.13454 (5) | - | - | - | G | A | 4,02 | 1,34 | 14,16 | 3,80 | 4,33 | 3,00 | 2,97 | 1,58 | 1 % | -0,841 | 0,709 | [illegible] |
| | | | 4 | 170815542 | rs754114601 | 0.18412 (7) | - | - | - | A | T | 1,36 | 1,34 | 12,65 | 2,36 | 0,30 | 3,00 | 2,97 | 1,29 | 0 % | 1,330 | 1,621 | [illegible] |
| | | | 4 | 170815545 | rs150108258 | 0.18412 (7) | - | - | - | C | T | 2,71 | 1,34 | 12,64 | 2,33 | 3,36 | 3,00 | 2,97 | 1,38 | 0 % | -1,329 | 1,194 | [illegible] |
| | | | 4 | 170815696 | rs190860986 | 0.24955 (6) | - | - | - | G | T | 1,64 | 1,34 | 12,15 | 1,04 | 1,09 | 3,00 | 2,97 | 1,24 | 5 % | 1,921 | 0,352 | [illegible] |
| MAF<5% | LINC02382 | 4:170814001-170818000, | 4 | 170816037 | rs72984174 | 0.43933 (5) | - | - | - | G | A | 5,12 | 5,42 | 11,30 | 3,82 | 3,57 | 3,00 | 2,97 | 2,01 | 0 % | 1,274 | 1,628 | [illegible] |
| | | 4:170816001-170820000 | 4 | 170816112 | rs72698830 | 1 (3a) | LINC024. | - | - | C | T | 2,60 | 5,42 | 11,14 | 4,95 | 0,56 | 3,00 | 2,97 | 2,18 | 1 % | 3,092 | 1,049 | [illegible] |
| | | | 4 | 170816478 | rs114343841 | 0.13454 (5) | - | - | - | G | T | 4,95 | 1,34 | 10,47 | 4,38 | 4,42 | 3,00 | 2,97 | 2,93 | 3 % | -0,598 | 0,412 | [illegible] |
| | | | 4 | 170816511 | rs114994390 | 0.1033 (5) | - | - | - | C | T | 3,13 | 1,34 | 10,41 | 3,78 | 0,88 | 3,00 | 2,97 | 2,36 | 0 % | -1,195 | 2,903 | [illegible] |
| | | | 4 | 170816582 | rs113810432 | 0.13454 (5) | - | - | - | CAG | C | 9,43 | 1,34 | 10,30 | 2,70 | 10,59 | 3,00 | 2,97 | 2,11 | 4 % | 1,828 | 0,380 | [illegible] |
| | | | 4 | 170816779 | rs996473946 | 0.18412 (7) | - | - | - | G | A | 5,83 | 1,34 | 10,00 | 0,93 | 4,75 | 3,00 | 2,97 | 6,58 | 0 % | 2,505 | 1,930 | [illegible] |
| | | | 4 | 170817189 | - | - | - | - | - | T | C | 3,78 | 1,34 | 9,46 | 3,81 | 0,80 | 3,00 | 2,97 | 6,00 | 0 % | -1,158 | 2,962 | [illegible] |
| | | | 4 | 170817700 | rs28586273 | 0.13454 (5) | - | - | - | T | A | 1,85 | 1,34 | 8,89 | 3,08 | 0,99 | 3,00 | 2,97 | 1,44 | 4 % | 1,828 | 0,380 | [illegible] |
| | | | 4 | 170817766 | rs578122364 | 0.27598 (6) | - | - | - | C | T | 0,11 | 1,34 | 8,83 | 1,68 | 0,06 | 3,00 | 2,97 | 1,56 | 0 % | -1,316 | 2,383 | [illegible] |
| | | | 4 | 170817861 | rs72984178 | 0.72948 (6) | - | - | - | A | C | 1,56 | 1,34 | 8,73 | 1,25 | 1,78 | 3,00 | 2,97 | 0,89 | 0 % | 1,274 | 1,628 | [illegible] |
| | | | 4 | 170817911 | rs186744836 | 0.18412 (7) | - | - | - | A | T | 10,25 | 1,34 | 8,69 | 1,51 | 8,98 | 3,00 | 2,97 | 0,97 | 0 % | -1,160 | 1,844 | [illegible] |
| MAF<5% | LINC02382 | 4:170816001-170820000, | 4 | 170818179 | rs1368006 | 0.4855 (6) | - | - | - | C | T | 3,46 | 1,34 | 8,44 | 1,84 | 4,86 | 3,00 | 2,97 | 1,03 | 3 % | 2,237 | 0,442 | [illegible] |
| | | 4:170818001-170822000 | 4 | 170818186 | rs72698832 | 0.18412 (7) | LINC024. | - | - | T | C | 3,05 | 1,34 | 8,44 | 2,00 | 2,62 | 3,00 | 2,97 | 1,00 | 1 % | 3,092 | 1,049 | [illegible] |
| | | | 4 | 170818302 | rs1372369542 | 0.35235 (6) | - | - | - | A | ACG | 1,57 | 1,34 | 8,34 | 1,10 | 3,42 | 3,00 | 2,97 | 0,99 | 1 % | 0,510 | 0,966 | [illegible] |
| | | | 4 | 170818303 | rs1474390910 | 0.17046 (6) | - | - | - | T | TCC( | 3,09 | 1,34 | 8,33 | 1,10 | 6,07 | 3,00 | 2,97 | 0,90 | 0 % | 4,342 | 2,377 | [illegible] |
| | | | 4 | 170818326 | rs188816203 | 0 (6) | - | - | - | C | T | 2,34 | 1,34 | 8,32 | 1,16 | 1,91 | 3,00 | 2,97 | 0,75 | 0 % | -1,450 | 1,609 | [illegible] |
| | | | 4 | 170818566 | rs72698834 | 0.17046 (6) | LINC024. | - | - | C | A | 5,60 | 1,34 | 8,12 | 1,25 | 9,19 | 3,00 | 2,97 | 1,28 | 1 % | 3,092 | 1,049 | [illegible] |
| | | | 4 | 170818585 | rs58800547 | 0.18412 (7) | - | - | - | A | C | 5,47 | 3,99 | 8,11 | 1,13 | 3,47 | 3,00 | 2,97 | 0,87 | 1 % | -0,558 | 0,932 | [illegible] |
| | | | 4 | 170818781 | rs28634540 | 0.58955 (5) | - | - | - | C | T | 0,09 | 3,99 | 7,96 | 2,03 | 0,06 | 3,00 | 2,97 | 0,82 | 4 % | 1,828 | 0,380 | [illegible] |
| | | | 4 | 170818803 | rs60112988 | 0.57638 (5) | - | - | - | A | G | 9,10 | 3,99 | 7,94 | 1,77 | 7,11 | 3,00 | 2,97 | 1,00 | 1 % | -0,558 | 0,932 | [illegible] |
| | | | 4 | 170818873 | rs1438476322 | 0.94 (5) | - | - | - | G | A | 0,10 | 7,29 | 7,89 | 2,00 | 0,06 | 3,00 | 2,97 | 1,35 | 0 % | 4,057 | 2,324 | [illegible] |
| | | | 4 | 170819021 | rs189541185 | 0.94867 (3a | - | - | - | T | C | 7,79 | 8,29 | 7,78 | 3,89 | 9,89 | 3,00 | 2,97 | 0,76 | 0 % | 5,150 | 2,532 | [illegible] |
| | | | 4 | 170819066 | rs13104718 | 0.60906 (4) | - | - | - | C | T | 9,92 | 7,29 | 7,75 | 2,15 | 9,55 | 3,00 | 2,97 | 0,88 | 3 % | 0,866 | 0,497 | [illegible] |
| | | | 4 | 170819320 | rs567783696 | 0.1131 (6) | - | - | - | A | G | 3,61 | 1,34 | 7,59 | 1,95 | 1,64 | 3,00 | 2,97 | 1,32 | 0 % | -1,201 | 2,081 | [illegible] |
| | | | 4 | 170819351 | - | - | - | - | - | T | C | 0,20 | 1,34 | 7,57 | 0,95 | 0,14 | 3,00 | 2,97 | 1,37 | 0 % | -1,331 | 2,346 | [illegible] |
| | | | 4 | 170819523 | rs75902432 | 0.18412 (7) | - | - | - | A | G | 3,39 | 1,34 | 7,46 | 2,51 | 2,50 | 3,00 | 2,97 | 0,78 | 1 % | 0,318 | 0,753 | [illegible] |
| | | | 4 | 170819569 | rs78706251 | 0.18412 (7) | - | - | - | C | T | 1,61 | 1,34 | 7,43 | 1,33 | 1,58 | 3,00 | 2,97 | 1,08 | 4 % | 1,828 | 0,380 | [illegible] |
| | | | 4 | 170819625 | rs72698835 | 0.18412 (7) | LINC024. | - | - | G | C | 0,25 | 1,34 | 7,40 | 2,17 | 0,50 | 3,00 | 2,97 | 1,14 | 1 % | 3,092 | 1,049 | [illegible] |
| | | | 4 | 170819694 | rs72698836 | 0.13454 (5) | - | - | - | C | T | 2,80 | 1,34 | 7,36 | 1,94 | 3,89 | 3,00 | 2,97 | 1,02 | 5 % | 1,921 | 0,352 | [illegible] |
| | | | 4 | 170819751 | rs543559469 | 0.69 (5) | - | - | - | C | G | 0,18 | 1,34 | 7,32 | 2,50 | 0,04 | 3,00 | 2,97 | 1,09 | 1 % | 0,357 | 0,751 | [illegible] |
| | | | 4 | 170819801 | rs1379992748 | 0.18412 (7) | - | - | - | G | A | 3,20 | 1,34 | 7,29 | 1,17 | 3,21 | 3,00 | 2,97 | 0,88 | 0 % | -1,156 | 3,030 | [illegible] |
| | | | 4 | 170819824 | rs554274784 | 0.18412 (7) | - | - | - | C | T | 1,09 | 1,34 | 7,28 | 0,73 | 0,43 | 3,00 | 2,97 | 0,90 | 0 % | -1,554 | 2,142 | [illegible] |
| | | | 4 | 170819852 | rs76264493 | 0 (6) | - | - | - | C | CA | 2,78 | 1,34 | 7,27 | 0,64 | 3,98 | 3,00 | 2,97 | 0,89 | 1 % | 1,969 | 0,906 | [illegible] |
| | | | 4 | 170819870 | - | - | - | - | - | C | T | 4,70 | 1,34 | 7,26 | 0,56 | 2,59 | 3,00 | 2,97 | 0,91 | 0 % | 9,433 | 3,249 | [illegible] |
| | | | 4 | 170819884 | rs28623002 | 0.18412 (7) | - | - | - | G | A | 3,29 | 1,34 | 7,25 | 0,46 | 6,02 | 3,00 | 2,97 | 0,90 | 4 % | 0,483 | 0,366 | [illegible] |
| | | | 4 | 170819932 | rs191087508 | 0.18412 (7) | - | - | - | G | A | 1,92 | 1,34 | 7,22 | 0,56 | 1,71 | 3,00 | 2,97 | 1,28 | 0 % | 6,637 | 2,798 | [illegible] |
| MAF<5% | LINC02382 | 4:170818001-170822000 | 4 | 170820134 | rs183398487 | 0.18412 (7) | - | - | - | T | A | 2,85 | 1,34 | 7,11 | 1,70 | 2,61 | 3,00 | 2,97 | 2,21 | 0 % | 1,238 | 1,625 | [illegible] |
| | | | 4 | 170820189 | rs10031519 | 0.58046 (6) | - | - | - | T | G | 6,39 | 1,34 | 7,08 | 2,81 | 5,59 | 3,00 | 2,97 | 2,15 | 4 % | 1,828 | 0,380 | [illegible] |
| | | | 4 | 170820243 | rs139608776 | 0.18412 (7) | - | - | - | CT | C | 4,42 | 1,34 | 7,05 | 2,31 | 7,53 | 3,00 | 2,97 | 1,97 | 4 % | 0,483 | 0,366 | [illegible] |
| | | | 4 | 170820313 | rs186527689 | 0.18412 (7) | - | - | - | T | C | 2,26 | 1,34 | 7,02 | 1,56 | 2,79 | 3,00 | 2,97 | 1,74 | 0 % | -1,108 | 3,496 | [illegible] |
| | | | 4 | 170820456 | rs569906656 | - | - | - | - | C | T | 1,23 | 1,34 | 6,94 | 3,76 | 1,14 | 3,00 | 2,97 | 2,74 | 0 % | 2,360 | 1,907 | [illegible] |
| | | | 4 | 170820477 | rs57587583 | 0.11188 (6) | - | - | - | G | GA | 0,05 | 1,34 | 6,93 | 3,47 | 0,29 | 3,00 | 2,97 | 2,07 | 1 % | -0,648 | 0,864 | [illegible] |
| | | | 4 | 170820506 | rs954061318 | 0.18412 (7) | - | - | - | C | T | 3,57 | 1,34 | 6,92 | 3,37 | 3,99 | 3,00 | 2,97 | 2,43 | 0 % | -1,216 | 2,724 | [illegible] |
| | | | 4 | 170820760 | rs938420468 | 0.34847 (6) | - | - | - | T | C | 2,89 | 1,34 | 6,79 | 1,94 | 0,92 | 3,00 | 2,97 | 1,83 | 0 % | -1,114 | 3,380 | [illegible] |
| | | | 4 | 170820766 | rs28688612 | 0.17046 (6) | - | - | - | C | T | 13,78 | 1,34 | 6,79 | 1,93 | 11,70 | 3,00 | 2,97 | 1,82 | 4 % | 1,828 | 0,380 | [illegible] |
| | | | 4 | 170820783 | rs12647730 | 0.18412 (7) | - | - | - | C | T | 6,96 | 1,34 | 6,78 | 2,00 | 3,17 | 3,00 | 2,97 | 1,91 | 1 % | -0,558 | 0,932 | [illegible] |
| | | | 4 | 170820786 | rs1263196790 | 0.18412 (7) | - | - | - | T | C | 10,98 | 1,34 | 6,78 | 2,50 | 8,64 | 3,00 | 2,97 | 2,04 | 0 % | -1,373 | 2,260 | [illegible] |
| | | | 4 | 170821013 | rs35436492 | 0.8051 (6) | - | - | - | T | C | 0,12 | 1,34 | 6,68 | 1,22 | 0,41 | 3,00 | 2,97 | 2,40 | 3 % | 0,866 | 0,497 | [illegible] |
| | | | 4 | 170821792 | rs1384876597 | 0.18412 (7) | - | - | - | G | T | 0,25 | 1,34 | 6,35 | 3,40 | 0,59 | 3,00 | 2,97 | 2,37 | 0 % | -1,332 | 2,323 | [illegible] |
| | | | 4 | 170821885 | rs72698837 | 0.18412 (7) | LINC024. | - | - | C | T | 1,99 | 3,99 | 6,31 | 3,76 | 2,88 | 3,00 | 2,97 | 3,73 | 1 % | 3,092 | 1,049 | [illegible] |
| MAF<5% | LINC02382 | 4:170822001-170826000 | 4 | 170822209 | rs1374449197 | 0.13454 (5) | - | - | - | T | C | 0,65 | 1,34 | 6,19 | 2,15 | 0,97 | 3,00 | 2,97 | 3,15 | 0 % | -1,313 | 2,381 | [illegible] |
| | | | 4 | 170822214 | rs552212403 | 0.13454 (5) | - | - | - | G | A | 0,23 | 1,34 | 6,18 | 2,17 | 0,77 | 3,00 | 2,97 | 3,15 | 0 % | 0,696 | 1,413 | [illegible] |
| | | | 4 | 170822614 | rs28419134 | 0.8051 (6) | - | - | - | T | C | 4,86 | 1,34 | 6,04 | 0,88 | 5,25 | 3,00 | 2,97 | 3,17 | 4 % | 0,483 | 0,366 | [illegible] |
| | | | 4 | 170822753 | rs10026794 | 0.18412 (7) | - | - | - | C | T | 7,02 | 1,34 | 5,99 | 0,78 | 10,70 | 3,00 | 2,97 | 2,69 | 4 % | 1,828 | 0,380 | [illegible] |
| | | | 4 | 170822816 | rs894132328 | 0.18412 (7) | - | - | - | T | C | 1,46 | 1,34 | 5,97 | 1,29 | 1,25 | 3,00 | 2,97 | 3,40 | 0 % | -1,216 | 2,724 | [illegible] |
| | | | 4 | 170822857 | rs188174580 | 0.18412 (7) | - | - | - | C | T | 7,27 | 1,34 | 5,95 | 1,08 | 9,90 | 3,00 | 2,97 | 3,02 | 0 % | -1,264 | 2,488 | [illegible] |
| | | | 4 | 170822953 | rs148125420 | 0.18412 (7) | - | - | - | C | G | 0,29 | 1,34 | 5,92 | 2,09 | 0,76 | 3,00 | 2,97 | 3,29 | 0 % | -1,285 | 2,441 | [illegible] |
| | | | 4 | 170823014 | rs200795565 | 0.34523 (6) | LINC024. | - | - | A | AT | 6,37 | 1,34 | 5,90 | 2,47 | 8,80 | 3,00 | 2,97 | 2,90 | 1 % | 3,092 | 1,049 | [illegible] |
| | | | 4 | 170823501 | rs9996374 | 0.18412 (7) | - | - | - | A | T | 0,67 | 1,34 | 5,75 | 2,60 | 0,37 | 3,00 | 2,97 | 2,90 | 4 % | 1,828 | 0,380 | [illegible] |
| | | | 4 | 170823577 | rs550398230 | 0.18412 (7) | - | - | - | C | A | 1,67 | 1,34 | 5,72 | 3,39 | 2,98 | 3,00 | 2,97 | 2,24 | 0 % | -1,275 | 1,757 | [illegible] |
| | | | 4 | 170823690 | rs938288306 | 0.11318 (6) | - | - | - | G | A | 0,15 | 1,34 | 5,69 | 0,69 | 0,55 | 3,00 | 2,97 | 3,24 | 0 % | -1,442 | 2,223 | [illegible] |
| | | | 4 | 170823913 | rs57208717 | 1 (3a) | - | - | - | T | A | 6,05 | 1,34 | 5,62 | 4,87 | 7,10 | 3,00 | 2,97 | 2,12 | 1 % | -0,558 | 0,932 | [illegible] |
| MAF<5% | LINC02382 | 4:170822001-170826000, | 4 | 170824028 | rs74682859 | 0.85988 (3a | - | - | - | C | A | 0,80 | 1,34 | 5,59 | 4,35 | 0,86 | 3,00 | 2,97 | 1,40 | 0 % | -1,285 | 2,441 | [illegible] |
| | | 4:170824001-170828000 | 4 | 170824033 | rs62344264 | 0.75204 (3a | - | - | - | C | A | 0,72 | 1,34 | 5,59 | 3,92 | 0,64 | 3,00 | 2,97 | 1,70 | 1 % | 0,312 | 0,905 | [illegible] |
| | | | 4 | 170824102 | rs28729811 | 0.60906 (4) | - | - | - | T | G | 1,70 | 1,34 | 5,57 | 2,40 | 1,05 | 3,00 | 2,97 | 1,67 | 4 % | 0,483 | 0,366 | [illegible] |
| | | | 4 | 170824139 | rs145563971 | 0.58955 (5) | - | - | - | C | A | 0,70 | 1,34 | 5,56 | 2,03 | 1,09 | 3,00 | 2,97 | 2,02 | 2 % | 0,179 | 0,508 | [illegible] |
| | | | 4 | 170824199 | rs148861690 | 0.98086 (3a | - | - | - | T | G | 1,78 | 1,34 | 5,54 | 2,24 | 0,27 | 3,00 | 2,97 | 3,63 | 0 % | 0,305 | 1,243 | [illegible] |
| | | | 4 | 170824300 | rs182491187 | 0.13454 (5) | - | - | - | A | T | 11,04 | 1,34 | 5,52 | 1,47 | 15,28 | 3,00 | 2,97 | 1,70 | 1 % | 3,898 | 1,020 | [illegible] |
| | | | 4 | 170824418 | - | - | - | - | - | T | C | 1,72 | 1,34 | 5,48 | 1,56 | 2,64 | 3,00 | 2,97 | 2,77 | 0 % | 4,477 | 2,404 | [illegible] |
| | | | 4 | 170824718 | rs72984186 | 0.22365 (6) | - | - | - | A | G | 1,84 | 1,34 | 5,41 | 2,68 | 1,42 | 3,00 | 2,97 | 3,76 | 0 % | 1,274 | 1,628 | [illegible] |
| | | | 4 | 170824793 | rs7693814 | 0 (6) | LINC024. | - | - | A | G | 4,94 | 1,34 | 5,39 | 2,14 | 5,53 | 3,00 | 2,97 | 3,83 | 1 % | 3,092 | 1,049 | [illegible] |
| | | | 4 | 170824832 | rs530401118 | 0.67385 (6) | LINC024. | - | - | T | TTAT | 2,58 | 1,34 | 5,38 | 1,60 | 7,93 | 3,00 | 2,97 | 4,35 | 1 % | 3,092 | 1,049 | [illegible] |
| | | | 4 | 170825051 | rs17056219 | 0.13454 (5) | - | - | - | A | G | 0,51 | 1,34 | 5,32 | 2,30 | 0,63 | 3,00 | 2,97 | 5,39 | 5 % | 1,921 | 0,352 | [illegible] |
| | | | 4 | 170825145 | rs530322997 | 0 (5) | - | - | - | T | A | 0,10 | 1,34 | 5,30 | 1,68 | 0,02 | 3,00 | 2,97 | 4,34 | 0 % | -1,442 | 2,223 | [illegible] |
| | | | 4 | 170825182 | rs76086095 | 0.13454 (5) | - | - | - | T | A | 3,13 | 3,99 | 5,29 | 1,90 | 3,46 | 3,00 | 2,97 | 3,68 | 1 % | -0,301 | 0,613 | [illegible] |
| | | | 4 | 170825490 | rs1458838998 | 0.18412 (7) | - | - | - | G | T | 1,83 | 1,34 | 5,21 | 2,96 | 2,78 | 3,00 | 2,97 | 4,92 | 0 % | -1,314 | 2,378 | [illegible] |
| | | | 4 | 170825635 | rs532250991 | 0.50466 (6) | - | - | - | T | C | 6,58 | 1,34 | 5,17 | 0,79 | 8,20 | 3,00 | 2,97 | 3,90 | 1 % | 0,031 | 0,820 | [illegible] |
| MAF<5% | LINC02382 | 4:170824001-170828000, | 4 | 170826105 | rs10005720 | 0.58283 (6) | - | - | - | T | C | 6,97 | 1,34 | 5,06 | 1,83 | 5,48 | 3,00 | 2,97 | 2,94 | 4 % | 1,828 | 0,380 | [illegible] |
| | | 4:170826001-170830000 | 4 | 170826286 | - | - | - | - | - | A | C | 5,03 | 1,34 | 5,02 | 4,30 | 4,04 | 3,00 | 2,97 | 2,66 | 0 % | -1,129 | 3,246 | [illegible] |
| | | | 4 | 170826289 | rs34247689 | 0.41664 (3a | - | - | - | C | G | 0,29 | 1,34 | 5,02 | 4,28 | 0,54 | 3,00 | 2,97 | 3,32 | 0 % | 1,274 | 1,628 | [illegible] |
| | | | 4 | 170826639 | rs539760175 | 0.60906 (4) | - | - | - | G | T | 0,31 | 1,34 | 4,94 | 6,33 | 0,64 | 3,00 | 2,97 | 2,49 | 0 % | -1,340 | 1,644 | [illegible] |
| | | | 4 | 170826685 | rs115325823 | 0.611 (5) | - | - | - | T | C | 4,48 | 3,99 | 4,93 | 5,66 | 2,71 | 3,00 | 2,97 | 2,42 | 0 % | -1,329 | 1,699 | [illegible] |
| | | | 4 | 170826749 | rs10016425 | 0.58955 (5) | - | - | - | G | A | 0,29 | 3,99 | 4,92 | 4,28 | 0,79 | 3,00 | 2,97 | 1,57 | 4 % | 1,828 | 0,380 | [illegible] |
| | | | 4 | 170826774 | - | - | - | - | - | T | C | 8,27 | 3,99 | 4,91 | 2,98 | 8,12 | 3,00 | 2,97 | 1,27 | 0 % | 3,235 | 2,171 | [illegible] |
| | | | 4 | 170826932 | rs77317701 | 0.58955 (5) | - | - | - | G | T | 0,28 | 1,34 | 4,88 | 1,74 | 0,72 | 3,00 | 2,97 | 2,00 | 4 % | 1,828 | 0,380 | [illegible] |
| | | | 4 | 170827136 | rs6839990 | 0.18412 (7) | - | - | - | G | C | 2,47 | 1,34 | 4,83 | 6,52 | 4,23 | 3,00 | 2,97 | 3,55 | 5 % | 1,921 | 0,352 | [illegible] |
| | | | 4 | 170827217 | rs142466235 | 0.44531 (6) | - | - | - | TC | T | 4,69 | 1,34 | 4,81 | 1,82 | 7,39 | 3,00 | 2,97 | 3,45 | 0 % | 1,274 | 1,628 | [illegible] |
| | | | 4 | 170827275 | rs77114548 | 0.34955 (6) | - | - | - | C | A | 0,29 | 3,99 | 4,80 | 3,52 | 0,54 | 3,00 | 2,97 | 3,37 | 4 % | 1,828 | 0,380 | [illegible] |
| | | | 4 | 170827723 | rs79549014 | 0.191 (5) | - | - | - | C | A | 2,24 | 9,87 | 4,71 | 10,99 | 2,50 | 3,00 | 2,97 | 4,11 | 1 % | -0,547 | 0,898 | [illegible] |
| | | | 4 | 170827952 | rs184188275 | 0.18412 (7) | - | - | - | A | T | 3,77 | 8,01 | 4,66 | 9,98 | 3,84 | 3,00 | 2,97 | 2,91 | 4 % | 1,828 | 0,380 | [illegible] |
| | | | 4 | 170827953 | rs188244305 | 0.18412 (7) | - | - | - | T | G | 2,86 | 8,01 | 4,66 | 10,00 | 1,85 | 3,00 | 2,97 | 2,91 | 4 % | 1,828 | 0,380 | [illegible] |
| MAF<5% | LINC02382 | 4:170826001-170830000, | 4 | 170828090 | rs10026172 | 0.60906 (4) | - | - | - | C | T | 1,66 | 11,69 | 4,64 | 4,32 | 1,55 | 3,00 | 2,97 | 3,54 | 4 % | 0,483 | 0,366 | [illegible] |
| | | 4:170828001-170832000 | 4 | 170828357 | rs34095614 | 0.32023 (6) | - | - | - | G | GAT. | 5,30 | 6,00 | 4,58 | 3,01 | 6,09 | 3,00 | 2,97 | 4,89 | 5 % | 1,921 | 0,352 | [illegible] |
| | | | 4 | 170828364 | rs114761463 | 0 (6) | LINC024. | - | - | A | T | 1,41 | 6,00 | 4,58 | 2,97 | 0,73 | 3,00 | 2,97 | 4,61 | 1 % | 3,092 | 1,049 | [illegible] |
| | | | 4 | 170829315 | rs567761490 | 0.18412 (7) | - | - | - | A | G | 7,23 | 1,34 | 4,41 | 1,15 | 3,90 | 3,00 | 2,97 | 3,73 | 0 % | -1,327 | 2,348 | [illegible] |
| | | | 4 | 170829475 | rs1432084 | 0.33118 (6) | - | - | - | A | G | 0,79 | 1,34 | 4,38 | 4,53 | 0,70 | 3,00 | 2,97 | 2,55 | 5 % | 1,921 | 0,352 | [illegible] |
| | | | 4 | 170829510 | rs1273667849 | 0 (6) | - | - | - | C | T | 3,37 | 1,34 | 4,37 | 3,89 | 3,65 | 3,00 | 2,97 | 2,08 | 0 % | 1,054 | 1,578 | [illegible] |
| | | | 4 | 170829704 | rs72698844 | 0.18412 (7) | LINC024. | - | - | T | G | 2,42 | 1,34 | 4,34 | 3,40 | 1,87 | 3,00 | 2,97 | 2,76 | 1 % | 3,092 | 1,049 | [illegible] |
| | | | 4 | 170829847 | rs1218435781 | 0.67105 (6) | - | - | - | T | G | 2,78 | 1,34 | 4,31 | 2,38 | 1,86 | 3,00 | 2,97 | 4,27 | 0 % | 3,519 | 2,219 | [illegible] |
| | | | 4 | 170829956 | rs72698846 | 0.18412 (7) | LINC024. | - | - | T | G | 6,26 | 1,34 | 4,30 | 4,19 | 5,09 | 3,00 | 2,97 | 1,75 | 1 % | 3,092 | 1,049 | [illegible] |
| MAF<5% | LINC02382 | 4:170828001-170832000, | 4 | 170830022 | rs531920548 | 0.01156 (6) | - | - | - | G | C | 8,40 | 3,99 | 4,28 | 4,90 | 6,24 | 3,00 | 2,97 | 1,93 | 0 % | -1,386 | 2,250 | [illegible] |
| | | 4:170830001-170834000 | 4 | 170830125 | rs936226009 | 0.34523 (6) | - | - | - | A | C | 1,44 | 3,99 | 4,27 | 4,23 | 1,48 | 3,00 | 2,97 | 2,61 | 0 % | -1,128 | 3,319 | [illegible] |
| | | | 4 | 170830426 | rs78863675 | 0.66931 (6) | - | - | - | G | A | 1,48 | 1,34 | 4,22 | 0,97 | 2,62 | 3,00 | 2,97 | 3,74 | 1 % | 0,312 | 0,905 | [illegible] |
| | | | 4 | 170830584 | rs28376630 | 0.18412 (7) | - | - | CBR4 in D | T | C | 5,21 | 1,34 | 4,19 | 0,52 | 5,58 | 3,00 | 2,97 | 3,54 | 4 % | 1,828 | 0,380 | [illegible] |
| | | | 4 | 170830672 | rs138868482 | 0.77072 (6) | - | - | CBR4 in D | A | G | 4,07 | 1,34 | 4,18 | 0,22 | 5,65 | 3,00 | 2,97 | 2,81 | 0 % | 1,274 | 1,628 | [illegible] |
| | | | 4 | 170830687 | rs142037372 | 0.16346 (6) | - | - | CBR4 in D | T | C | 2,64 | 1,34 | 4,18 | 0,25 | 2,85 | 3,00 | 2,97 | 2,94 | 4 % | -0,191 | 0,419 | [illegible] |
| | | | 4 | 170831097 | rs190103069 | 0.4986 (6) | - | - | CBR4 in D | C | T | 1,54 | 1,34 | 4,11 | 2,98 | 1,77 | 3,00 | 2,97 | 3,95 | 3 % | 0,908 | 0,433 | [illegible] |
| | | | 4 | 170831158 | - | - | - | - | - | G | C | 3,79 | 1,34 | 4,10 | 4,84 | 9,05 | 3,00 | 2,97 | 5,21 | 0 % | -1,161 | 2,937 | [illegible] |
| | | | 4 | 170831357 | rs72698847 | 0.18412 (7) | - | - | CBR4 in D | C | A | 0,43 | 1,34 | 4,07 | 3,76 | 0,38 | 3,00 | 2,97 | 4,09 | 1 % | 3,092 | 1,049 | [illegible] |
| | | | 4 | 170831465 | rs72698848 | 0.31452 (6) | LINC024. | - | CBR4 in D | T | C | 2,42 | 1,34 | 4,05 | 3,78 | 1,80 | 3,00 | 2,97 | 3,07 | 1 % | 3,092 | 1,049 | [illegible] |
| | | | 4 | 170831657 | - | - | - | - | - | T | C | 7,10 | 1,34 | 4,03 | 3,83 | 5,28 | 3,00 | 2,97 | 3,60 | 0 % | -1,305 | 2,382 | [illegible] |
| | | | 4 | 170831695 | rs553835661 | 0.13454 (5) | - | - | CBR4 in D | A | G | 0,69 | 1,34 | 4,02 | 3,41 | 1,00 | 3,00 | 2,97 | 2,57 | 0 % | -1,347 | 2,299 | [illegible] |
| | | | 4 | 170831812 | rs1425140629 | 0.18412 (7) | - | - | CBR4 in D | G | A | 1,46 | 1,34 | 4,00 | 3,83 | 1,53 | 3,00 | 2,97 | 3,38 | 0 % | -1,296 | 2,415 | [illegible] |
| | | | 4 | 170831815 | rs72698849 | 0.18412 (7) | - | - | CBR4 in D | G | C | 0,50 | 1,34 | 4,00 | 3,47 | 1,10 | 3,00 | 2,97 | 3,60 | 5 % | 1,921 | 0,352 | [illegible] |
| | | | 4 | 170831873 | rs143630387 | 0.13454 (5) | - | - | CBR4 in D | G | A | 11,13 | 1,34 | 3,99 | 1,95 | 13,43 | 3,00 | 2,97 | 2,70 | 1 % | -0,701 | 0,816 | [illegible] |
| MAF<5% | LINC02382 | 4:170830001-170834000 | 4 | 170832243 | rs181294724 | 0.18412 (7) | - | - | CBR4 in D | A | G | 7,29 | 1,34 | 3,94 | 1,57 | 4,57 | 3,00 | 2,97 | 3,19 | 1 % | -0,426 | 0,906 | [illegible] |
| | | | 4 | 170832273 | rs1048215378 | 0.66931 (6) | - | - | CBR4 in D | T | C | 2,86 | 1,34 | 3,93 | 2,04 | 1,51 | 3,00 | 2,97 | 2,68 | 0 % | -1,525 | 2,134 | [illegible] |
| | | | 4 | 170832433 | rs576207599 | 0.5295 (6) | - | - | CBR4 in D | T | C | 7,39 | 1,34 | 3,91 | 0,77 | 3,25 | 3,00 | 2,97 | 4,31 | 0 % | 3,901 | 2,298 | [illegible] |
| | | | 4 | 170832495 | rs72698850 | 0.18412 (7) | - | - | CBR4 in D | G | T | 0,24 | 1,34 | 3,90 | 1,94 | 0,95 | 3,00 | 2,97 | 4,93 | 1 % | 1,059 | 1,080 | [illegible] |
| | | | 4 | 170832597 | rs17056221 | 0.14931 (6) | LINC024. | - | CBR4 in D | G | A | 0,06 | 1,34 | 3,89 | 2,44 | 0,41 | 3,00 | 2,97 | 3,91 | 1 % | 3,092 | 1,049 | [illegible] |
| | | | 4 | 170832716 | rs10520194 | 0.15359 (6) | - | - | CBR4 in D | C | T | 0,05 | 1,34 | 3,87 | 2,01 | 0,43 | 3,00 | 2,97 | 2,44 | 4 % | 1,828 | 0,380 | [illegible] |
| | | | 4 | 170832726 | rs62344265 | 0.2909 (6) | - | - | CBR4 in D | C | T | 2,50 | 1,34 | 3,87 | 1,88 | 3,88 | 3,00 | 2,97 | 2,62 | 4 % | 0,483 | 0,366 | [illegible] |
| | | | 4 | 170833256 | rs72698851 | 0.58955 (5) | LINC024. | - | CBR4 in D | A | G | 4,75 | 9,00 | 3,80 | 2,15 | 2,69 | 3,00 | 2,97 | 5,59 | 1 % | 3,092 | 1,049 | [illegible] |
| | | | 4 | 170833302 | rs141379824 | 0.58955 (5) | - | - | CBR4 in D | G | T | 8,06 | 8,29 | 3,79 | 2,16 | 13,56 | 3,00 | 2,97 | 4,83 | 1 % | 0,648 | 1,011 | [illegible] |
| | | | 4 | 170833486 | rs72698852 | 0.18412 (7) | LINC024. | - | CBR4 in D | T | C | 6,86 | 5,42 | 3,77 | 1,18 | 6,18 | 3,00 | 2,97 | 4,82 | 1 % | 3,092 | 1,049 | [illegible] |
| | | | 4 | 170833541 | rs17056236 | 0.34955 (6) | - | - | CBR4 in D | A | G | 6,75 | 5,42 | 3,76 | 1,22 | 5,35 | 3,00 | 2,97 | 4,78 | 0 % | 1,274 | 1,628 | [illegible] |
| | | | 4 | 170833837 | rs72698854 | 0.18412 (7) | LINC024. | - | CBR4 in D | A | G | 8,48 | 6,45 | 3,72 | 5,31 | 9,11 | 3,00 | 2,97 | 6,38 | 1 % | 3,092 | 1,049 | [illegible] |
| MAF<5% | LINC02382 | 4:170844001-170848000 | 4 | 170844117 | rs145511795,rs: | - | LINC024. | - | - | CATT | C | 8,45 | 1,34 | 2,75 | 2,27 | 12,50 | 3,00 | 2,97 | 7,74 | 1 % | 3,092 | 1,049 | [illegible] |
| | | | 4 | 170844264 | - | - | - | - | - | C | G | 7,31 | 1,34 | 2,74 | 1,40 | 9,24 | 3,00 | 2,97 | 6,77 | 0 % | -1,145 | 3,061 | [illegible] |
| | | | 4 | 170844780 | rs72986119 | 0.18412 (7) | - | - | - | C | T | 1,96 | 1,34 | 2,70 | 2,52 | 0,78 | 3,00 | 2,97 | 3,99 | 0 % | 1,274 | 1,628 | [illegible] |
| | | | 4 | 170844880 | rs548529862 | 0.18412 (7) | - | - | - | T | C | 5,97 | 1,34 | 2,69 | 1,48 | 5,34 | 3,00 | 2,97 | 4,45 | 0 % | -1,278 | 1,874 | [illegible] |
| | | | 4 | 170844897 | rs559076967 | 0.18412 (7) | - | - | - | A | T | 4,78 | 1,34 | 2,69 | 1,13 | 5,99 | 3,00 | 2,97 | 5,14 | 0 % | -1,350 | 2,295 | [illegible] |
| | | | 4 | 170845408 | rs528238359 | 0.18412 (7) | - | - | - | A | C | 2,70 | 1,34 | 2,66 | 1,48 | 2,90 | 3,00 | 2,97 | 6,38 | 0 % | -1,194 | 2,760 | [illegible] |
| | | | 4 | 170845634 | rs182237622 | 0.13454 (5) | - | - | - | C | T | 2,68 | 1,34 | 2,64 | 3,88 | 2,51 | 3,00 | 2,97 | 6,26 | 0 % | -1,173 | 2,872 | [illegible] |
| | | | 4 | 170845888 | rs72698867 | 0.13454 (5) | LINC024. | - | - | A | G | 1,66 | 1,34 | 2,63 | 2,26 | 1,40 | 3,00 | 2,97 | 6,30 | 1 % | 3,092 | 1,049 | [illegible] |
| MAF<5% | LINC02382 | 4:170844001-170848000, | 4 | 170846022 | rs113166986 | 0.53679 (5) | - | - | - | GA | G | 0,80 | 1,34 | 2,62 | 3,13 | 1,15 | 3,00 | 2,97 | 4,87 | 1 % | 3,092 | 1,049 | [illegible] |
| | | 4:170846001-170850000 | 4 | 170846332 | rs141697881 | 0.00545 (5) | LINC024. | - | - | A | G | 3,91 | 1,34 | 2,60 | 3,20 | 4,40 | 3,00 | 2,97 | 3,52 | 1 % | 3,092 | 1,049 | [illegible] |
| | | | 4 | 170846523 | rs988582933 | 0.18412 (7) | - | - | AADAT in | G | A | 1,08 | 1,34 | 2,58 | 4,01 | 1,23 | 3,00 | 2,97 | 3,73 | 0 % | -1,416 | 1,123 | [illegible] |
| | | | 4 | 170846645 | rs1246592130 | 0.18412 (7) | - | - | AADAT in | T | A | 2,96 | 1,34 | 2,58 | 3,59 | 4,19 | 3,00 | 2,97 | 3,18 | 0 % | 0,694 | 1,421 | [illegible] |
| | | | 4 | 170846681 | rs72698872 | 0.18412 (7) | LINC024. | - | AADAT in | A | G | 3,42 | 1,34 | 2,57 | 3,47 | 4,30 | 3,00 | 2,97 | 4,83 | 1 % | 3,092 | 1,049 | [illegible] |
| | | | 4 | 170846766 | rs1368007 | 0.32023 (6) | - | - | AADAT in | A | G | 5,95 | 1,34 | 2,57 | 1,69 | 7,29 | 3,00 | 2,97 | 2,97 | 4 % | 1,828 | 0,380 | [illegible] |
| | | | 4 | 170847006 | rs186822606 | 0.18412 (7) | - | - | AADAT in | T | A | 3,80 | 1,34 | 2,55 | 1,63 | 5,30 | 3,00 | 2,97 | 4,72 | 0 % | -1,340 | 1,644 | [illegible] |
| | | | 4 | 170847420 | rs10010530 | 0.55602 (6) | - | - | AADAT in | C | T | 1,34 | 1,34 | 2,53 | 2,27 | 1,70 | 3,00 | 2,97 | 5,12 | 4 % | 0,483 | 0,366 | [illegible] |
| | | | 4 | 170847448 | rs34920093 | 0.62527 (6) | - | - | AADAT in | TG | T | 1,60 | 1,34 | 2,53 | 2,33 | 3,13 | 3,00 | 2,97 | 4,67 | 4 % | 1,828 | 0,380 | [illegible] |
| | | | 4 | 170847603 | rs1394900993 | 0.13454 (5) | - | - | AADAT in | C | T | 1,96 | 5,42 | 2,52 | 4,10 | 1,55 | 3,00 | 2,97 | 6,21 | 0 % | 4,649 | 2,436 | [illegible] |
| MAF<5% | LINC02382 | 4:170846001-170850000 | 4 | 170848060 | rs1297115632 | 0.13454 (5) | - | - | AADAT in | T | C | 1,00 | 3,99 | 2,49 | 4,18 | 0,82 | 3,00 | 2,97 | 3,92 | 0 % | -1,201 | 2,734 | [illegible] |
| | | | 4 | 170848492 | rs538630385 | 0.18412 (7) | - | - | AADAT in | C | A | 3,82 | 1,34 | 2,47 | 3,40 | 4,35 | 3,00 | 2,97 | 4,95 | 0 % | -1,274 | 2,481 | [illegible] |
| | | | 4 | 170849346 | rs139927327 | 0.40391 (6) | - | - | AADAT in | T | C | 5,83 | 1,34 | 2,42 | 1,76 | 5,75 | 3,00 | 2,97 | 1,75 | 5 % | 0,480 | 0,355 | [illegible] |
| | | | 4 | 170849508 | rs112194785 | 0.18412 (7) | - | - | AADAT in | G | A | 1,43 | 1,34 | 2,41 | 2,74 | 2,87 | 3,00 | 2,97 | 4,47 | 3 % | 2,121 | 0,435 | [illegible] |
| | | | 4 | 170849538 | rs115403732 | 0.18412 (7) | - | - | AADAT in | C | T | 2,63 | 1,34 | 2,41 | 2,68 | 4,00 | 3,00 | 2,97 | 3,82 | 1 % | 1,077 | 0,775 | [illegible] |
| MAF<5% | LINC02382 | 4:170854001-170858000 | 4 | 170854339 | rs185527439 | 0.60906 (4) | - | - | - | A | G | 12,45 | 6,88 | 2,30 | 3,66 | 15,33 | 3,00 | 2,97 | 2,74 | 1 % | -1,236 | 1,173 | [illegible] |
| | | | 4 | 170854405 | rs1228031407 | 0.58955 (5) | - | - | - | TTA | T | 12,31 | 7,64 | 2,30 | 5,37 | 14,04 | 3,00 | 2,97 | 2,66 | 0 % | -1,141 | 3,100 | [illegible] |
| | | | 4 | 170854628 | rs76452905 | 0.58283 (6) | - | - | - | A | G | 2,19 | 5,42 | 2,31 | 2,11 | 1,96 | 3,00 | 2,97 | 1,36 | 3 % | 2,121 | 0,435 | [illegible] |
| | | | 4 | 170854683 | rs72698883 | 0.18412 (7) | - | - | - | T | C | 6,40 | 3,99 | 2,31 | 3,63 | 4,76 | 3,00 | 2,97 | 1,02 | 0 % | -1,417 | 2,247 | [illegible] |
| | | | 4 | 170854736 | rs79561364 | 0.18412 (7) | - | - | - | C | T | 8,61 | 4,89 | 2,32 | 3,06 | 9,10 | 3,00 | 2,97 | 0,75 | 3 % | 2,121 | 0,435 | [illegible] |

| | | | | | | | | | | | | | | | | | | |
|---|---|---|---|---|---|---|---|---|---|---|---|---|---|---|---|---|---|---|
| | | | 4 | 170854952 | rs188994677 | 0.38564 (6) | - | - | - | G | A | 0,11 | 3,99 | 2,33 | 0,72 | 0,52 | 3,00 | [illegible] |
| | | | 4 | 170855163 | rs10000123 | 0.44483 (6) | - | - | - | A | C | 8,86 | 3,99 | 2,34 | 1,61 | 7,61 | 3,00 | [illegible] |
| | | | 4 | 170855495 | rs747354854 | 0.18412 (7) | - | - | - | C | T | 2,41 | 1,34 | 2,36 | 1,01 | 5,52 | 3,00 | [illegible] |
| | | | 4 | 170855546 | rs79547949 | 0.48494 (6) | - | - | - | T | C | 7,02 | 1,34 | 2,36 | 1,50 | 5,86 | 3,00 | [illegible] |
| | | | 4 | 170855780 | rs199878234 | 0.53679 (5) | - | - | - | GA | G | 2,64 | 1,34 | 2,37 | 5,50 | 4,43 | 3,00 | [illegible] |
| | | | 4 | 170855906 | rs566583851 | 0.13454 (5) | - | - | - | T | C | 10,68 | 1,34 | 2,38 | 5,53 | 8,48 | 3,00 | [illegible] |
| | | | 4 | 170856228 | rs901436938 | 0.13454 (5) | - | - | - | A | G | 14,31 | 1,34 | 2,40 | 3,47 | 14,52 | 3,00 | [illegible] |
| | | | 4 | 170856453 | rs146720515 | 0.52984 (5) | *LINC024.* | - | - | C | T | 2,41 | 1,34 | 2,41 | 0,80 | 1,90 | 3,00 | [illegible] |
| | | | 4 | 170856570 | rs200480372 | 0.77072 (6) | - | - | - | C | CA | 4,10 | 1,34 | 2,41 | 0,47 | 7,21 | 3,00 | [illegible] |
| | | | 4 | 170857229 | rs183527895 | 0.18412 (7) | - | - | - | C | T | 4,45 | 1,34 | 2,45 | 1,38 | 3,61 | 3,00 | [illegible] |
| | | | 4 | 170857316 | rs77908121 | 0 (6) | - | - | - | G | A | 0,02 | 1,34 | 2,46 | 1,77 | 0,02 | 3,00 | [illegible] |
| | | | 4 | 170857330 | rs1322137497 | 0.1131 (6) | - | - | - | A | C | 3,45 | 1,34 | 2,46 | 1,54 | 2,98 | 3,00 | [illegible] |
| | | | 4 | 170857752 | rs80233182 | 0.46462 (5) | - | - | - | A | C | 5,13 | 1,34 | 2,48 | 1,56 | 4,48 | 3,00 | [illegible] |
| | | | 4 | 170857830 | rs201406548 | 0.42858 (5) | *LINC024.* | - | - | CAG | C | 3,71 | 1,34 | 2,49 | 4,34 | 6,97 | 3,00 | [illegible] |
| MAF<5% | *LINC02382* | 4:170862001-170866000 | 4 | 170862192 | rs549485552 | 0.18412 (7) | - | - | - | T | A | 3,47 | 1,34 | 2,77 | 2,87 | 3,29 | 3,00 | [illegible] |
| | | | 4 | 170862367 | rs1731569341 | - | - | - | - | T | G | 7,16 | 3,99 | 2,78 | 2,22 | 7,36 | 3,00 | [illegible] |
| | | | 4 | 170862729 | rs114653527 | 0.19425 (6) | - | - | - | T | A | 3,47 | 1,34 | 2,81 | 0,92 | 4,51 | 3,00 | [illegible] |
| | | | 4 | 170862730 | rs188355402 | 0.27598 (6) | - | - | - | A | T | 0,65 | 1,34 | 2,81 | 0,92 | 0,98 | 3,00 | [illegible] |
| | | | 4 | 170862791 | rs1420660689 | 0.45238 (6) | - | - | - | A | G | 6,12 | 1,34 | 2,81 | 1,23 | 5,64 | 3,00 | [illegible] |
| | | | 4 | 170862855 | rs1441677858 | 0.18412 (7) | - | - | - | A | G | 3,44 | 1,34 | 2,82 | 0,80 | 1,39 | 3,00 | [illegible] |
| | | | 4 | 170862969 | rs186455959 | 0.11237 (6) | - | - | - | A | G | 3,70 | 1,34 | 2,83 | 0,62 | 5,48 | 3,00 | [illegible] |
| | | | 4 | 170863314 | rs573036449 | 0.18412 (7) | - | - | - | C | T | 6,48 | 1,34 | 2,85 | 2,04 | 7,23 | 3,00 | [illegible] |
| | | | 4 | 170863391 | rs763664038 | 0 (6) | - | - | - | C | T | 13,29 | 1,34 | 2,86 | 1,54 | 14,38 | 3,00 | [illegible] |
| | | | 4 | 170863505 | rs115132921 | 0.18412 (7) | *LINC024.* | - | - | G | T | 1,29 | 1,34 | 2,87 | 1,08 | 1,92 | 3,00 | [illegible] |
| | | | 4 | 170863612 | rs191288081 | 0 (6) | - | - | - | C | T | 6,64 | 1,34 | 2,88 | 1,87 | 6,50 | 3,00 | [illegible] |
| | | | 4 | 170863798 | rs187726451 | 0.18412 (7) | - | - | - | C | T | 5,94 | 1,34 | 2,89 | 0,72 | 6,68 | 3,00 | [illegible] |
| MAF<5% | *LINC02382* | 4:170862001-170866000, 4:170864001-170868000 | 4 | 170864060 | rs527689082 | 0.43272 (6) | - | - | - | G | GT | 1,28 | 1,34 | 2,91 | 0,85 | 2,54 | 3,00 | [illegible] |
| | | | 4 | 170864180 | rs78739837 | 0.18412 (7) | - | *MFAP3L* | - | C | T | 1,69 | 1,34 | 2,92 | 1,57 | 4,05 | 3,00 | [illegible] |
| | | | 4 | 170864227 | rs1201298272 | 0.43133 (6) | - | - | - | A | G | 1,29 | 1,34 | 2,92 | 1,52 | 1,30 | 3,00 | [illegible] |
| | | | 4 | 170864277 | rs762662749 | 0.18412 (7) | - | - | - | G | A | 5,22 | 1,34 | 2,93 | 1,82 | 4,83 | 3,00 | [illegible] |
| | | | 4 | 170864331 | rs571930754 | 0.18412 (7) | - | - | - | C | T | 2,74 | 1,34 | 2,93 | 1,15 | 4,12 | 3,00 | [illegible] |
| | | | 4 | 170864480 | rs6840051 | 0.35507 (6) | - | - | - | T | A | 5,11 | 1,34 | 2,95 | 1,09 | 4,53 | 3,00 | [illegible] |
| | | | 4 | 170864955 | rs994480 | 0.16346 (6) | - | - | - | G | A | 0,35 | 3,99 | 2,98 | 2,50 | 0,25 | 3,00 | [illegible] |
| | | | 4 | 170865050 | rs72700815 | 0.18412 (7) | *LINC024.* | - | - | A | G | 4,71 | 1,34 | 2,99 | 2,26 | 4,52 | 3,00 | [illegible] |
| | | | 4 | 170865488 | rs184978053 | 0.58955 (5) | | | - | A | T | 2,78 | 5,42 | 3,03 | 1,75 | 1,94 | 3,00 | [illegible] |
| MAF<5% | *LINC02382* | 4:170864001-170868000 | 4 | 170866008 | rs938232046 | 0.13454 (5) | - | - | - | G | A | 1,76 | 6,45 | 3,07 | 7,37 | 1,52 | 3,00 | [illegible] |
| | | | 4 | 170866117 | rs139703247 | 0.13454 (5) | *AADAT* | - | - | TC | T | 1,72 | 5,42 | 3,08 | 7,29 | 2,54 | 3,00 | [illegible] |
| | | | 4 | 170866143 | rs189454604 | 0.49945 (5) | - | - | - | T | C | 0,89 | 5,42 | 3,08 | 8,13 | 0,61 | 3,00 | [illegible] |
| | | | 4 | 170866192 | rs139358235 | 0.02 (6) | - | - | - | C | G | 1,49 | 5,42 | 3,09 | 7,20 | 1,77 | 3,00 | [illegible] |
| | | | 4 | 170866407 | rs989112 | 0.18412 (7) | - | - | - | T | C | 0,70 | 1,34 | 3,11 | 2,55 | 0,40 | 3,00 | [illegible] |
| | | | 4 | 170866617 | rs1432090 | 0.18412 (7) | - | - | - | C | T | 0,31 | 1,34 | 3,13 | 2,25 | 0,51 | 3,00 | [illegible] |
| | | | 4 | 170866691 | rs79337986 | 0.18412 (7) | - | - | - | C | T | 4,48 | 1,34 | 3,13 | 3,92 | 6,59 | 3,00 | [illegible] |
| | | | 4 | 170867465 | rs147790568 | 0.18412 (7) | - | - | - | A | G | 5,94 | 1,34 | 3,20 | 1,52 | 5,36 | 3,00 | [illegible] |
| | | | 4 | 170867542 | rs72700821 | 0.22365 (6) | *LINC024.* | - | - | G | T | 1,68 | 3,99 | 3,21 | 3,44 | 2,87 | 3,00 | [illegible] |
| MAF<5% | *LINC02382* | 4:170874001-170878000 | 4 | 170874007 | rs138700014 | 0.18412 (7) | - | - | - | C | A | 1,53 | 1,34 | 3,98 | 0,95 | 1,73 | 3,00 | [illegible] |
| | | | 4 | 170874060 | rs572738152 | 0.18412 (7) | *LINC024.* | - | - | C | T | 1,02 | 1,34 | 3,99 | 0,79 | 3,19 | 3,00 | [illegible] |
| | | | 4 | 170874113 | rs907907968 | 0.18412 (7) | - | - | - | T | C | 0,76 | 1,34 | 3,99 | 0,56 | 0,38 | 3,00 | [illegible] |
| | | | 4 | 170874154 | rs192725480 | 0.23808 (6) | *LINC024.* | - | - | G | C | 0,80 | 1,34 | 4,00 | 0,14 | 3,41 | 3,00 | [illegible] |
| | | | 4 | 170874200 | rs190624611 | 0.18412 (7) | - | - | - | T | C | 1,99 | 1,34 | 4,01 | 0,16 | 3,20 | 3,00 | [illegible] |
| | | | 4 | 170874299 | rs111680007 | 0.18412 (7) | - | - | - | G | A | 2,10 | 1,34 | 4,02 | 0,24 | 3,24 | 3,00 | [illegible] |
| | | | 4 | 170874470 | rs1440586778 | 0.40391 (6) | - | - | *GALNTL6* | A | AC | 8,13 | 1,34 | 4,05 | 0,21 | 9,36 | 3,00 | [illegible] |
| | | | 4 | 170874471 | rs1181302600 | 0.67105 (6) | - | - | *GALNTL6* | T | A | 9,23 | 1,34 | 4,05 | 0,21 | 9,26 | 3,00 | [illegible] |
| | | | 4 | 170874478 | rs1361865340 | 0.38564 (6) | - | - | *GALNTL6* | T | G | 8,45 | 1,34 | 4,05 | 0,21 | 8,10 | 3,00 | [illegible] |
| | | | 4 | 170874706 | rs552467358 | 0.15367 (6) | - | - | *GALNTL6* | C | T | 3,40 | 1,34 | 4,08 | 0,13 | 0,81 | 3,00 | [illegible] |
| | | | 4 | 170875286 | rs745623208 | 0.13454 (5) | - | - | *GALNTL6* | T | C | 10,88 | 1,34 | 4,17 | 1,27 | 13,05 | 3,00 | [illegible] |
| | | | 4 | 170875627 | rs62344314 | 0.39217 (6) | - | - | *GALNTL6* | T | C | 9,13 | 1,34 | 4,23 | 1,00 | 7,01 | 3,00 | [illegible] |
| | | | 4 | 170875635 | rs145117641 | 0.33117 (6) | - | - | *GALNTL6* | T | C | 9,03 | 1,34 | 4,23 | 1,21 | 6,77 | 3,00 | [illegible] |
| | | | 4 | 170875687 | rs183523078 | 0.58955 (5) | - | - | *GALNTL6* | A | G | 0,12 | 1,34 | 4,24 | 1,01 | 0,37 | 3,00 | [illegible] |
| | | | 4 | 170875977 | rs111989100 | 0.60906 (4) | - | - | *GALNTL6* | C | A | 1,62 | 13,50 | 4,29 | 4,77 | 4,83 | 3,00 | [illegible] |
| | | | 4 | 170876175 | rs79550062 | 0.60906 (4) | - | - | *GALNTL6* | C | T | 3,27 | 13,83 | 4,32 | 6,42 | 5,74 | 3,00 | [illegible] |
| | | | 4 | 170876356 | rs574913099 | 0.47482 (3a | - | - | *GALNTL6* | C | T | 1,88 | 1,34 | 4,35 | 3,21 | 1,78 | 3,00 | [illegible] |
| | | | 4 | 170876382 | rs920290630 | 0.47043 (3a | - | - | *GALNTL6* | A | G | 3,62 | 3,99 | 4,36 | 3,29 | 2,84 | 3,00 | [illegible] |
| | | | 4 | 170876397 | rs192796507 | 0.57256 (3a | - | - | *GALNTL6* | T | G | 1,27 | 3,99 | 4,36 | 3,85 | 0,46 | 3,00 | [illegible] |
| | | | 4 | 170876548 | rs76073793 | 0.3231 (3a) | - | - | *GALNTL6* | T | C | 3,30 | 5,42 | 4,39 | 8,33 | 2,88 | 3,00 | [illegible] |
| | | | 4 | 170876962 | - | - | - | - | - | T | C | 12,34 | 1,34 | 4,46 | 1,29 | 16,17 | 3,00 | [illegible] |
| | | | 4 | 170877027 | rs1231225559 | 0.18412 (7) | - | - | *GALNTL6* | G | T | 1,87 | 1,34 | 4,47 | 1,81 | 3,84 | 3,00 | [illegible] |
| | | | 4 | 170877315 | rs145066146 | 0.18412 (7) | - | - | *GALNTL6* | T | C | 3,22 | 1,34 | 4,53 | 1,02 | 3,20 | 3,00 | [illegible] |
| | | | 4 | 170877317 | rs188746883 | 0.18412 (7) | *LINC024.* | - | *GALNTL6* | T | C | 1,97 | 1,34 | 4,53 | 0,91 | 1,49 | 3,00 | [illegible] |
| | | | 4 | 170877353 | rs548216664 | 0.13144 (6) | - | - | *GALNTL6* | A | C | 0,51 | 1,34 | 4,53 | 0,90 | 0,92 | 3,00 | [illegible] |
| | | | 4 | 170877382 | rs1424870637 | 0.17931 (6) | - | - | *GALNTL6* | G | T | 1,90 | 1,34 | 4,54 | 1,12 | 3,23 | 3,00 | [illegible] |
| | | | 4 | 170877457 | rs775377567 | 0.74742 (6) | - | - | *GALNTL6* | A | G | 6,00 | 1,34 | 4,55 | 1,12 | 7,11 | 3,00 | [illegible] |
| | | | 4 | 170877650 | rs1161712604 | 0.27391 (6) | - | - | *GALNTL6* | T | C | 3,37 | 1,34 | 4,59 | 1,49 | 2,83 | 3,00 | [illegible] |
| MAF<5% | *LINC02382* | 4:170878001-170882000 | 4 | 170878202 | - | - | - | - | - | T | G | 2,93 | 3,99 | 4,70 | 2,87 | 1,81 | 3,00 | [illegible] |
| | | | 4 | 170878264 | rs72700830 | 0.18412 (7) | *LINC024.* | - | - | C | A | 1,20 | 3,99 | 4,71 | 2,17 | 1,76 | 3,00 | [illegible] |
| | | | 4 | 170878383 | rs147899213 | 0 (6) | - | - | - | CAT | C | 0,69 | 3,99 | 4,74 | 0,77 | 1,88 | 3,00 | [illegible] |
| | | | 4 | 170878865 | rs536397968 | 0.18412 (7) | - | - | - | G | A | 2,85 | 1,34 | 4,84 | 1,32 | 4,66 | 3,00 | [illegible] |
| | | | 4 | 170878907 | rs200807377 | 0.335 (6) | - | - | - | A | AT | 5,01 | 1,34 | 4,85 | 1,77 | 6,79 | 3,00 | [illegible] |
| | | | 4 | 170878953 | rs183028368 | 0.0135 (5) | - | - | - | A | G | 5,81 | 1,34 | 4,86 | 1,53 | 4,14 | 3,00 | [illegible] |
| | | | 4 | 170879076 | rs749369894 | 0.18412 (7) | - | - | - | G | A | 1,48 | 1,34 | 4,88 | 3,29 | 1,92 | 3,00 | [illegible] |
| | | | 4 | 170879516 | rs114586839 | 0.20285 (6) | - | - | - | C | G | 2,83 | 1,34 | 4,98 | 4,14 | 4,73 | 3,00 | [illegible] |
| | | | 4 | 170879546 | rs187751552 | 0.17046 (6) | - | - | - | T | C | 3,21 | 1,34 | 4,99 | 2,49 | 3,71 | 3,00 | [illegible] |
| | | | 4 | 170879608 | rs1487452274 | 0.37351 (6) | - | - | - | T | C | 1,89 | 1,34 | 5,00 | 1,72 | 1,86 | 3,00 | [illegible] |
| | | | 4 | 170879622 | - | - | - | - | - | G | C | 0,37 | 1,34 | 5,01 | 2,17 | 1,11 | 3,00 | [illegible] |
| | | | 4 | 170879944 | rs565902012 | 0.55602 (6) | - | - | - | G | A | 0,86 | 1,34 | 5,08 | 1,06 | 1,98 | 3,00 | [illegible] |
| MAF<5% | *LINC02382* | 4:170878001-170882000, 4:170880001-170884000 | 4 | 170880245 | rs72700832 | 0.18412 (7) | - | - | - | C | G | 0,52 | 1,34 | 5,15 | 1,56 | 1,33 | 3,00 | [illegible] |
| | | | 4 | 170880467 | rs72700833 | 0.34847 (6) | *LINC024.* | - | - | G | T | 1,12 | 1,34 | 5,20 | 2,80 | 1,93 | 3,00 | [illegible] |
| | | | 4 | 170881053 | rs72700834 | 0 (6) | *LINC024.* | - | - | G | A | 0,84 | 3,99 | 5,35 | 1,48 | 1,72 | 3,00 | [illegible] |
| | | | 4 | 170881742 | rs111448462 | 0 (5) | - | - | - | G | A | 4,04 | 1,34 | 5,53 | 0,83 | 6,99 | 3,00 | [illegible] |
| | | | 4 | 170881832 | rs111419883 | 0.13454 (5) | - | - | - | G | A | 1,26 | 3,99 | 5,56 | 1,39 | 1,09 | 3,00 | [illegible] |
| | | | 4 | 170881960 | rs111630640,rs | - | - | - | - | TGAA | T | 2,03 | 5,42 | 5,59 | 1,03 | 5,98 | 3,00 | [illegible] |
| MAF<5% | *LINC02382* | 4:170880001-170884000 | 4 | 170882063 | rs113023025 | 0.19425 (6) | *LINC024.* | - | - | C | T | 2,95 | 3,99 | 5,62 | 1,34 | 4,15 | 3,00 | [illegible] |
| | | | 4 | 170882194 | rs111846962 | 0.18412 (7) | *LINC024.* | - | - | A | T | 0,55 | 1,34 | 5,66 | 0,69 | 0,27 | 3,00 | [illegible] |
| | | | 4 | 170882461 | rs762583989 | 0.65422 (6) | - | - | - | G | T | 1,17 | 1,34 | 5,74 | 0,58 | 1,25 | 3,00 | [illegible] |
| | | | 4 | 170882691 | rs112585186 | 0.53625 (6) | - | - | - | T | G | 2,87 | 1,34 | 5,81 | 0,77 | 4,23 | 3,00 | [illegible] |
| | | | 4 | 170882892 | rs113320048 | 0.18412 (7) | *LINC024.* | - | - | A | G | 1,09 | 1,34 | 5,88 | 1,56 | 1,18 | 3,00 | [illegible] |
| | | | 4 | 170882925 | rs112061184 | 0.18412 (7) | - | - | - | T | C | 2,96 | 1,34 | 5,89 | 1,53 | 4,04 | 3,00 | [illegible] |
| | | | 4 | 170883106 | rs561448616 | 0.18412 (7) | - | - | - | T | C | 2,49 | 1,34 | 5,95 | 1,46 | 1,34 | 3,00 | [illegible] |
| | | | 4 | 170883257 | rs111934042 | 0.13454 (5) | *LINC024.* | - | - | C | G | 0,86 | 1,34 | 6,00 | 4,73 | 1,04 | 3,00 | [illegible] |
| | | | 4 | 170883467 | rs77858156 | 0.41041 (6) | - | - | - | A | G | 0,64 | 1,34 | 6,07 | 2,05 | 0,24 | 3,00 | [illegible] |
| | | | 4 | 170883712 | rs199748629 | 0.27792 (6) | - | - | - | G | GA | 3,85 | 1,34 | 6,16 | 1,81 | 7,59 | 3,00 | [illegible] |
| | | | 4 | 170883746 | rs113791122 | 0.14464 (6) | *LINC024.* | - | - | T | C | 0,24 | 1,34 | 6,18 | 0,87 | 0,26 | 3,00 | [illegible] |
| | | | 4 | 170883820 | rs533004652 | 0.7707 (6) | - | - | - | G | A | 0,08 | 3,99 | 6,20 | 2,78 | 0,38 | 3,00 | [illegible] |
| | | | 4 | 170883938 | rs72700839 | 0 (6) | *LINC024.* | - | - | G | A | 4,05 | 3,99 | 6,25 | 2,37 | 8,79 | 3,00 | [illegible] |
| MAF<5% | *LINC02382* | 4:171010001-171014000 | 4 | 171010026 | rs144818511 | 0.16346 (6) | - | - | - | C | CT | 0,34 | 1,34 | 2,23 | 3,75 | 0,58 | 3,00 | [illegible] |
| | | | 4 | 171010114 | rs1361426784 | 0.29006 (6) | - | - | - | G | A | 1,10 | 1,34 | 2,23 | 1,64 | 2,49 | 3,00 | [illegible] |
| | | | 4 | 171010123 | rs189172170 | 0.18412 (7) | - | - | - | C | T | 0,48 | 1,34 | 2,23 | 1,39 | 0,81 | 3,00 | [illegible] |
| | | | 4 | 171010144 | rs192896819 | 0.18412 (7) | - | - | - | A | G | 3,29 | 1,34 | 2,23 | 1,32 | 5,35 | 3,00 | [illegible] |
| | | | 4 | 171010199 | - | - | - | - | - | T | G | 3,82 | 1,34 | 2,24 | 1,74 | 3,96 | 3,00 | [illegible] |
| | | | 4 | 171010542 | rs62346230 | 0 (5) | - | - | - | C | T | 1,42 | 3,99 | 2,25 | 6,84 | 0,94 | 3,00 | [illegible] |
| | | | 4 | 171010607 | rs555388198 | 0.18412 (7) | - | - | - | C | G | 2,67 | 1,34 | 2,26 | 3,07 | 3,99 | 3,00 | [illegible] |
| | | | 4 | 171010705 | rs1201231557 | 0.18412 (7) | - | - | - | G | A | 5,80 | 1,34 | 2,26 | 0,82 | 8,51 | 3,00 | [illegible] |
| | | | 4 | 171010979 | - | - | - | - | - | G | T | 0,52 | 1,34 | 2,27 | 1,82 | 0,36 | 3,00 | [illegible] |
| | | | 4 | 171011089 | - | - | - | - | - | G | C | 7,87 | 1,34 | 2,28 | 2,38 | 9,57 | 3,00 | [illegible] |
| | | | 4 | 171011103 | rs540602121 | 0.18412 (7) | - | - | - | A | G | 10,04 | 1,34 | 2,28 | 1,95 | 9,57 | 3,00 | [illegible] |
| | | | 4 | 171011475 | rs532596999 | 0.35267 (5) | - | - | - | G | GA | 3,98 | 1,34 | 2,30 | 2,49 | 5,96 | 3,00 | [illegible] |
| | | | 4 | 171011987 | rs181690010 | 0 (5) | - | - | - | C | T | 3,72 | 1,34 | 2,33 | 2,64 | 3,79 | 3,00 | [illegible] |
| | | | 4 | 171011994 | rs141793130 | 0.13454 (5) | - | - | - | C | T | 1,47 | 1,34 | 2,33 | 2,66 | 0,89 | 3,00 | [illegible] |
| | | | 4 | 171012207 | rs780713731 | 0 (6) | - | - | - | C | T | 7,16 | 1,34 | 2,34 | 2,71 | 7,35 | 3,00 | [illegible] |
| | | | 4 | 171012269 | rs889526921 | 0.18412 (7) | - | - | - | T | C | 5,44 | 1,34 | 2,34 | 3,60 | 4,02 | 3,00 | [illegible] |
| | | | 4 | 171012748 | rs185844156 | 0 (5) | - | - | - | G | C | 3,50 | 1,34 | 2,37 | 1,63 | 5,75 | 3,00 | [illegible] |
| | | | 4 | 171012759 | rs76621736 | 0.13454 (5) | - | - | - | C | T | 4,96 | 1,34 | 2,37 | 1,55 | 5,75 | 3,00 | [illegible] |
| | | | 4 | 171012958 | rs116262263 | 0.13454 (5) | - | - | - | C | T | 2,73 | 1,34 | 2,38 | 3,98 | 3,79 | 3,00 | [illegible] |
| | | | 4 | 171013029 | rs189761461 | 0 (5) | - | - | - | C | T | 5,84 | 3,99 | 2,38 | 4,06 | 7,46 | 3,00 | [illegible] |
| | | | 4 | 171013032 | rs966003146 | 0.39931 (5) | - | - | - | C | A | 0,27 | 3,99 | 2,38 | 4,71 | 0,18 | 3,00 | [illegible] |
| | | | 4 | 171013117 | rs150766792 | 0.14427 (5) | - | - | - | A | G | 4,90 | 3,99 | 2,39 | 4,03 | 4,40 | 3,00 | [illegible] |
| | | | 4 | 171013410 | rs115581937 | 0.15359 (6) | *LINC024.* | - | - | T | A | 0,78 | 1,34 | 2,40 | 2,75 | 1,10 | 3,00 | [illegible] |
| | | | 4 | 171013541 | rs6830066 | 0.21221 (6) | - | - | - | G | A | 1,65 | 1,34 | 2,41 | 1,64 | 1,35 | 3,00 | [illegible] |
| | | | 4 | 171013669 | rs6830294 | 0 (6) | - | - | - | G | T | 1,63 | 3,99 | 2,42 | 2,45 | 3,47 | 3,00 | [illegible] |
| | | | 4 | 171013746 | rs193263591 | 0.4855 (6) | - | - | - | C | A | 10,94 | 1,34 | 2,42 | 2,24 | 7,12 | 3,00 | [illegible] |
| | | | 4 | 171013933 | rs72706513 | 0.18412 (7) | *LINC024.* | - | - | G | A | 0,10 | 1,34 | 2,43 | 2,20 | 0,36 | 3,00 | [illegible] |
| MAF<5% | *LINC02382* | 4:171032001-171036000 | 4 | 171032373 | rs74394400 | 0.18412 (7) | - | - | - | A | T | 4,43 | 1,34 | 4,43 | 1,90 | 5,66 | 3,00 | [illegible] |
| | | | 4 | 171032433 | rs75306363 | 0.18412 (7) | - | - | - | G | A | 0,00 | 1,34 | 4,44 | 1,00 | 0,07 | 3,00 | [illegible] |
| | | | 4 | 171032512 | rs72706528 | 0.47449 (6) | *LINC024.* | - | - | C | G | 0,00 | 1,34 | 4,46 | 0,60 | 0,03 | 3,00 | [illegible] |
| | | | 4 | 171032572 | rs1033255738 | 0.00333 (6) | - | - | - | CAAA | C | 2,89 | 1,34 | 4,47 | 0,62 | 7,11 | 3,00 | [illegible] |
| | | | 4 | 171032583 | rs538558891 | 0.26281 (6) | - | - | - | T | TA | 1,65 | 1,34 | 4,47 | 0,56 | 3,54 | 3,00 | [illegible] |
| | | | 4 | 171032717 | rs944052014 | 0.53625 (6) | - | - | - | C | T | 0,14 | 1,34 | 4,50 | 3,15 | 0,34 | 3,00 | [illegible] |
| | | | 4 | 171032893 | rs77920842 | 0.13454 (5) | - | - | - | T | G | 1,55 | 1,34 | 4,54 | 3,58 | 2,57 | 3,00 | [illegible] |
| | | | 4 | 171033168 | rs534530292 | 0 (5) | - | - | - | C | CA | 7,04 | 1,34 | 4,60 | 1,69 | 8,82 | 3,00 | [illegible] |
| | | | 4 | 171033270 | rs534530292 | 0.13454 (5) | - | - | - | T | C | 2,94 | 1,34 | 4,63 | 2,15 | 2,63 | 3,00 | [illegible] |
| | | | 4 | 171033321 | rs62344480 | 0.01689 (5) | - | - | - | A | G | 0,80 | 1,34 | 4,64 | 2,75 | 1,41 | 3,00 | [illegible] |
| | | | 4 | 171033372 | rs576454580 | 0.3882 (5) | - | - | - | CAAT | C | 1,88 | 1,34 | 4,65 | 2,43 | 4,39 | 3,00 | [illegible] |
| | | | 4 | 171033388 | rs567714339 | 0.60906 (4) | - | - | - | A | G | 8,60 | 1,34 | 4,66 | 1,97 | 9,62 | 3,00 | [illegible] |
| | | | 4 | 171034044 | rs72706530 | 0.20285 (6) | *LINC024.* | - | - | T | C | 0,27 | 3,99 | 4,82 | 1,90 | 0,51 | 3,00 | [illegible] |
| | | | 4 | 171034896 | - | - | - | - | - | G | T | 1,28 | 1,34 | 5,06 | 2,38 | 2,73 | 3,00 | [illegible] |
| | | | 4 | 171035432 | rs1276763570 | 0 (6) | - | - | - | T | C | 4,53 | 1,34 | 5,22 | 1,74 | 4,95 | 3,00 | [illegible] |
| | | | 4 | 171035553 | rs555578498 | 0.18412 (7) | - | - | - | G | C | 1,99 | 1,34 | 5,26 | 1,79 | 4,82 | 3,00 | [illegible] |
| | | | 4 | 171035565 | rs143085326 | 0.18412 (7) | - | - | - | T | C | 3,35 | 1,34 | 5,27 | 1,81 | 3,75 | 3,00 | [illegible] |
| | | | 4 | 171035681 | rs72706531 | 0.18412 (7) | *LINC024.* | - | - | T | C | 3,93 | 1,34 | 5,31 | 2,19 | 4,86 | 3,00 | [illegible] |
| | | | 4 | 171035987 | rs113727037 | 0.80555 (3a | - | - | - | C | G | 0,34 | 6,45 | 5,41 | 3,65 | 0,54 | 3,00 | [illegible] |
| MAF<5% | *LINC02382* | 4:171050001-171054000 | 4 | 171050076 | rs900248889 | 0 (6) | - | - | *MFAP3L* i | C | T | 1,23 | 1,34 | 5,68 | 3,62 | 1,45 | 3,00 | [illegible] |
| | | | 4 | 171050098 | rs921025045 | 0.36297 (6) | - | - | *MFAP3L* i | A | G | 6,35 | 1,34 | 5,68 | 3,59 | 4,21 | 3,00 | [illegible] |
| | | | 4 | 171050286 | rs1391403329 | 0.19425 (6) | - | - | *MFAP3L* i | T | C | 7,50 | 1,34 | 5,68 | 3,48 | 10,13 | 3,00 | [illegible] |
| | | | 4 | 171050444 | rs1167744827 | 0.18412 (7) | - | - | *MFAP3L* i | A | C | 1,14 | 1,34 | 5,68 | 1,84 | 0,71 | 3,00 | [illegible] |
| | | | 4 | 171050617 | rs775599629 | 0.18412 (7) | - | - | *MFAP3L* i | G | T | 1,27 | 1,34 | 5,69 | 2,26 | 1,93 | 3,00 | [illegible] |
| | | | 4 | 171050785 | rs1410696517 | 0.26281 (6) | - | - | *MFAP3L* i | A | G | 4,64 | 1,34 | 5,69 | 1,58 | 5,31 | 3,00 | [illegible] |
| | | | 4 | 171050794 | rs185217809 | 0.18412 (7) | - | - | *MFAP3L* i | A | G | 1,84 | 1,34 | 5,69 | 1,96 | 1,68 | 3,00 | [illegible] |
| | | | 4 | 171051003 | rs189659831 | 0.18412 (7) | - | - | *MFAP3L* i | C | G | 3,30 | 1,34 | 5,70 | 3,13 | 6,93 | 3,00 | [illegible] |
| | | | 4 | 171051479 | rs1340673092 | 0.00083 (6) | - | - | *MFAP3L* i | C | A | 4,52 | 6,00 | 5,72 | 3,73 | 6,49 | 3,00 | [illegible] |
| | | | 4 | 171051522 | rs188083265 | 0.18412 (7) | - | - | *MFAP3L* i | G | T | 1,17 | 5,42 | 5,72 | 3,26 | 1,94 | 3,00 | [illegible] |
| | | | 4 | 171051582 | rs1266177810 | 0.22365 (6) | - | - | *MFAP3L* i | C | A | 1,92 | 5,42 | 5,72 | 2,43 | 2,15 | 3,00 | [illegible] |
| | | | 4 | 171051652 | - | - | - | - | *MFAP3L* i | C | G | 0,06 | 3,99 | 5,72 | 2,33 | 0,07 | 3,00 | [illegible] |
| | | | 4 | 171051707 | rs72706543 | 0.18412 (7) | *LINC024.* | - | *MFAP3L* i | A | G | 5,29 | 3,99 | 5,73 | 2,25 | 4,35 | 3,00 | [illegible] |
| | | | 4 | 171051944 | rs1732356346 | - | - | - | - | C | T | 0,76 | 1,34 | 5,74 | 5,90 | 0,59 | 3,00 | [illegible] |
| | | | 4 | 171052320 | rs190544213 | 0.18412 (7) | - | - | - | A | G | 1,39 | 1,34 | 5,77 | 1,49 | 1,01 | 3,00 | [illegible] |
| | | | 4 | 171052349 | rs181630684 | 0.18412 (7) | - | - | - | A | G | 2,83 | 1,34 | 5,77 | 1,50 | 4,25 | 3,00 | [illegible] |
| | | | 4 | 171052366 | rs1233421270 | 0.18412 (7) | - | - | - | A | C | 0,93 | 1,34 | 5,77 | 1,31 | 0,76 | 3,00 | [illegible] |
| | | | 4 | 171052541 | rs186858647 | 0.42619 (6) | - | - | - | G | A | 1,87 | 1,34 | 5,79 | 1,58 | 4,74 | 3,00 | [illegible] |
| | | | 4 | 171052914 | rs144892300 | 0.13454 (5) | - | - | - | G | T | 1,72 | 1,34 | 5,83 | 2,51 | 4,79 | 3,00 | [illegible] |
| | | | 4 | 171053023 | rs72706546 | 0.18412 (7) | *LINC024.* | - | - | T | C | 4,44 | 1,34 | 5,84 | 2,20 | 5,58 | 3,00 | [illegible] |
| | | | 4 | 171053088 | rs1213651322 | 0.15359 (6) | - | - | - | A | C | 7,86 | 1,34 | 5,85 | 1,11 | 9,16 | 3,00 | [illegible] |
| | | | 4 | 171053108 | - | - | - | - | - | A | C | 1,31 | 1,34 | 5,85 | 0,85 | 0,95 | 3,00 | [illegible] |
| | | | 4 | 171053374 | rs761114109 | 0.18412 (7) | - | - | - | CAT | C | 3,39 | 1,34 | 5,88 | 3,58 | 8,42 | 3,00 | [illegible] |
| | | | 4 | 171053559 | rs9996746 | 0.13454 (5) | - | - | - | T | C | 4,55 | 1,34 | 5,91 | 2,08 | 4,17 | 3,00 | [illegible] |
| | | | 4 | 171053570 | rs10027596 | 0.13454 (5) | - | - | - | C | A | 1,45 | 1,34 | 5,91 | 2,45 | 3,22 | 3,00 | [illegible] |

| | | | | | | | | | | | | | | | | | | | | | | | | | | | | | | | | | | | | | | | | | | | | | | | | | | | | | | | | | | | | | | | | | | | | | | | |
|---|---|---|---|---|---|---|---|---|---|---|---|---|---|---|---|---|---|---|---|---|---|---|---|---|---|---|---|---|---|---|---|---|---|---|---|---|---|---|---|---|---|---|---|---|---|---|---|---|---|---|---|---|---|---|---|---|---|---|---|---|---|---|---|---|---|---|---|---|---|---|---|
| | | | 4 | 171053694 | rs553652839 | 0.28786 (6) | - | - | - | AG | A | 2,54 | 3,99 | 5,92 | 3,11 | 7,23 | 3,00 | 2,97 | 1,97 | 1 % | -0,157 | 1,064 | 8,83E-01 | - | 7 | 1 % | -1,122 | 1,532 | 4,64E-01 | - | 6 | 1 % | -1,068 | 2,304 | 6,43E-01 | - | 6 | 0,7 % | -0,517 | 2,579 | 1,82E-01 | ≤3 | 48 | 0,8 % | -0,355 | 2,034 | 4,70E-01 | ≤3 | 47 | 0,8 % | -0,757 | 1,160 | 3,80E-01 | ≤3 | 46 | 0,7 % | -0,073 | 0,080 | 3,64E-01 | 0,7 % | -0,060 | 0,083 | 4,65E-01 | 0,7 % | -0,077 | 0,087 | 3,74E-01 | 0,7 % | 0,061 | 0,160 | 7,02E-01 |
| | | | 4 | 171053760 | rs182466405 | 0.43933 (5) | - | - | - | G | T | 0,67 | 3,99 | 5,93 | 2,56 | 0,89 | 3,00 | 2,97 | 1,29 | 0 % | -1,327 | 2,348 | 5,72E-01 | - | ≤3 | 0 % | -1,211 | 2,695 | 6,53E-01 | - | ≤3 | 0 % | -1,075 | 4,191 | 7,98E-01 | - | ≤3 | - | - | - | - | - | - | - | - | - | - | - | - | - | - | - | - | - | - | - | - | - | - | - | - | - | - | - | - | - | - | - | - | - | - |
| | | | 4 | 171053881 | rs72706549 | 0.13454 (5) | *LINC024.* | - | - | G | A | 0,68 | 1,34 | 5,95 | 3,63 | 0,90 | 3,00 | 2,97 | 1,08 | 1 % | 2,765 | 1,008 | 6,09E-03 | - | 7 | 0 % | 1,955 | 1,772 | 2,70E-01 | - | 4 | 0 % | 6,209 | 2,697 | 2,13E-02 | - | 4 | 0,7 % | -0,010 | 2,755 | 9,78E-01 | - | 62 | 0,7 % | -0,688 | 2,104 | 1,48E-01 | - | 57 | 0,8 % | -0,593 | 1,215 | 4,71E-01 | - | 57 | 0,7 % | 0,099 | 0,075 | 1,87E-01 | 0,7 % | 0,096 | 0,077 | 2,14E-01 | 0,7 % | 0,094 | 0,081 | 2,45E-01 | 0,7 % | -0,001 | 0,151 | 9,94E-01 |
| MAF<5% | *LINC02382* | 4:171070001-171074000 | 4 | 171070042 | rs72706601 | 0.60906 (4) | *LINC024.* | - | - | G | T | 1,42 | 6,00 | 3,91 | 5,95 | 4,75 | 3,00 | 2,97 | 0,24 | 1 % | 2,765 | 1,008 | 6,09E-03 | - | 7 | 0 % | 1,955 | 1,772 | 2,70E-01 | - | 4 | 0 % | 6,209 | 2,697 | 2,13E-02 | - | 4 | 0,7 % | -0,010 | 2,755 | 9,78E-01 | - | 62 | 0,7 % | -0,688 | 2,104 | 1,48E-01 | - | 57 | 0,8 % | -0,593 | 1,215 | 4,71E-01 | - | 57 | 0,7 % | 0,098 | 0,075 | 1,88E-01 | 0,7 % | 0,096 | 0,077 | 2,16E-01 | 0,7 % | 0,093 | 0,081 | 2,47E-01 | 0,7 % | -0,001 | 0,151 | 9,96E-01 |
| | | | 4 | 171070158 | rs72706602 | 0.60906 (4) | *LINC024.* | - | - | G | A | 2,55 | 3,99 | 3,89 | 4,06 | 4,27 | 3,00 | 2,97 | 0,47 | 1 % | 2,765 | 1,008 | 6,09E-03 | - | 7 | 0 % | 1,955 | 1,772 | 2,70E-01 | - | 4 | 0 % | 6,209 | 2,697 | 2,13E-02 | - | 4 | 0,7 % | -0,010 | 2,755 | 9,78E-01 | - | 62 | 0,7 % | -0,688 | 2,104 | 1,48E-01 | - | 57 | 0,8 % | -0,593 | 1,215 | 4,71E-01 | - | 57 | 0,7 % | 0,098 | 0,075 | 1,88E-01 | 0,7 % | 0,096 | 0,077 | 2,16E-01 | 0,7 % | 0,093 | 0,081 | 2,47E-01 | 0,7 % | -0,001 | 0,151 | 9,96E-01 |
| | | | 4 | 171070413 | rs72708403 | 0.13454 (5) | *LINC024.* | - | - | C | G | 3,66 | 3,99 | 3,85 | 3,93 | 6,91 | 3,00 | 2,97 | 0,39 | 1 % | 2,765 | 1,008 | 6,09E-03 | - | 7 | 0 % | 1,955 | 1,772 | 2,70E-01 | - | 4 | 0 % | 6,209 | 2,697 | 2,13E-02 | - | 4 | 0,7 % | -0,010 | 2,755 | 9,78E-01 | - | 62 | 0,7 % | -0,688 | 2,104 | 1,48E-01 | - | 57 | 0,8 % | -0,593 | 1,215 | 4,71E-01 | - | 57 | 0,7 % | 0,098 | 0,075 | 1,88E-01 | 0,7 % | 0,096 | 0,077 | 2,16E-01 | 0,7 % | 0,093 | 0,081 | 2,47E-01 | 0,7 % | -0,001 | 0,151 | 9,96E-01 |
| | | | 4 | 171070487 | rs72708404 | 0.1694 (5) | *LINC024.* | - | - | G | A | 0,94 | 5,42 | 3,84 | 4,35 | 0,85 | 3,00 | 2,97 | 0,41 | 1 % | 2,765 | 1,008 | 6,09E-03 | - | 7 | 0 % | 1,955 | 1,772 | 2,70E-01 | - | 4 | 0 % | 6,209 | 2,697 | 2,13E-02 | - | 4 | 0,7 % | -0,010 | 2,755 | 9,78E-01 | - | 62 | 0,7 % | -0,688 | 2,104 | 1,48E-01 | - | 57 | 0,8 % | -0,593 | 1,215 | 4,71E-01 | - | 57 | 0,7 % | 0,098 | 0,075 | 1,88E-01 | 0,7 % | 0,096 | 0,077 | 2,16E-01 | 0,7 % | 0,093 | 0,081 | 2,47E-01 | 0,7 % | -0,001 | 0,151 | 9,96E-01 |
| | | | 4 | 171070531 | rs72708406 | 0.13454 (5) | *LINC024.* | - | - | C | T | 1,83 | 5,42 | 3,83 | 3,49 | 3,01 | 3,00 | 2,97 | 0,44 | 1 % | 2,765 | 1,008 | 6,09E-03 | - | 7 | 0 % | 1,955 | 1,772 | 2,70E-01 | - | 4 | 0 % | 6,209 | 2,697 | 2,13E-02 | - | 4 | 0,7 % | -0,010 | 2,755 | 9,78E-01 | - | 62 | 0,7 % | -0,688 | 2,104 | 1,48E-01 | - | 57 | 0,8 % | -0,593 | 1,215 | 4,71E-01 | - | 57 | 0,7 % | 0,098 | 0,075 | 1,88E-01 | 0,7 % | 0,096 | 0,077 | 2,16E-01 | 0,7 % | 0,093 | 0,081 | 2,47E-01 | 0,7 % | -0,001 | 0,151 | 9,96E-01 |
| | | | 4 | 171070710 | rs72708407 | 0.18412 (7) | *LINC024.* | - | - | A | C | 3,05 | 1,34 | 3,81 | 1,10 | 3,83 | 3,00 | 2,97 | 0,59 | 1 % | 2,765 | 1,008 | 6,09E-03 | - | 7 | 0 % | 1,955 | 1,772 | 2,70E-01 | - | 4 | 0 % | 6,209 | 2,697 | 2,13E-02 | - | 4 | 0,7 % | -0,010 | 2,755 | 9,78E-01 | - | 62 | 0,7 % | -0,688 | 2,104 | 1,48E-01 | - | 57 | 0,8 % | -0,593 | 1,215 | 4,71E-01 | - | 57 | 0,7 % | 0,098 | 0,075 | 1,88E-01 | 0,7 % | 0,096 | 0,077 | 2,16E-01 | 0,7 % | 0,093 | 0,081 | 2,47E-01 | 0,7 % | -0,001 | 0,151 | 9,96E-01 |
| | | | 4 | 171070838 | rs554176589 | 0.18412 (7) | - | - | - | T | C | 2,49 | 1,34 | 3,79 | 2,25 | 3,50 | 3,00 | 2,97 | 0,34 | 0 % | -1,258 | 2,521 | 6,18E-01 | - | ≤3 | 0 % | -1,129 | 3,261 | 7,29E-01 | - | ≤3 | 0 % | -1,077 | 4,192 | 7,97E-01 | - | ≤3 | - | - | - | - | - | - | - | - | - | - | - | - | - | - | - | - | - | - | - | - | - | - | - | - | - | - | - | - | - | - | - | - | - | - |
| | | | 4 | 171070840 | rs143518320 | 0.18412 (7) | - | - | - | G | T | 0,47 | 1,34 | 3,79 | 2,25 | 0,96 | 3,00 | 2,97 | 0,36 | 0 % | 3,202 | 2,176 | 1,41E-01 | - | ≤3 | 0 % | 4,543 | 2,430 | 6,15E-02 | - | ≤3 | - | - | - | - | - | - | - | - | - | - | - | - | - | - | - | - | - | - | - | - | - | - | - | - | - | - | - | - | - | - | - | - | - | - | - | - | - | - | - | - |
| | | | 4 | 171070869 | rs150955954 | 0.18412 (7) | *LINC024.* | - | - | G | C | 0,81 | 1,34 | 3,78 | 3,48 | 3,00 | 3,00 | 2,97 | 0,30 | 1 % | 2,765 | 1,008 | 6,09E-03 | - | 7 | 0 % | 1,955 | 1,772 | 2,70E-01 | - | 4 | 0 % | 6,209 | 2,697 | 2,13E-02 | - | 4 | 0,7 % | -0,010 | 2,755 | 9,78E-01 | - | 62 | 0,7 % | -0,688 | 2,104 | 1,48E-01 | - | 57 | 0,8 % | -0,593 | 1,215 | 4,71E-01 | - | 57 | 0,7 % | 0,098 | 0,075 | 1,88E-01 | 0,7 % | 0,096 | 0,077 | 2,16E-01 | 0,7 % | 0,093 | 0,081 | 2,47E-01 | 0,7 % | -0,001 | 0,151 | 9,96E-01 |
| | | | 4 | 171070909 | rs188961487 | 0.18412 (7) | - | - | - | A | T | 0,81 | 1,34 | 3,78 | 2,74 | 0,69 | 3,00 | 2,97 | 0,34 | 0 % | -1,254 | 1,841 | 4,96E-01 | - | ≤3 | 0 % | -1,151 | 2,239 | 6,07E-01 | - | ≤3 | 0 % | -1,062 | 3,263 | 7,45E-01 | - | ≤3 | 0,1 % | 0,329 | 0,952 | 7,54E-01 | - | 7 | 0,1 % | 0,947 | 0,756 | 4,74E-01 | - | 7 | 0,1 % | -0,294 | 0,432 | 8,99E-01 | - | 7 | 0,1 % | -0,214 | 0,183 | 2,43E-01 | 0,1 % | -0,259 | 0,189 | 1,70E-01 | 0,1 % | -0,293 | 0,198 | 1,39E-01 | 0,1 % | 0,178 | 0,354 | 6,15E-01 |
| | | | 4 | 171070966 | rs562532534 | 0.18412 (7) | - | - | - | C | A | 3,55 | 1,34 | 3,77 | 2,57 | 5,75 | 3,00 | 2,97 | 0,52 | 0 % | -1,244 | 1,922 | 5,17E-01 | - | ≤3 | 0 % | -1,123 | 2,478 | 6,50E-01 | - | ≤3 | 0 % | -1,069 | 3,395 | 7,53E-01 | - | ≤3 | 0,1 % | -1,047 | 0,851 | 3,73E-01 | - | 6 | 0,1 % | -0,909 | 0,678 | 5,38E-01 | - | 6 | 0,1 % | -0,775 | 0,392 | 7,61E-01 | - | 6 | 0,1 % | 0,032 | 0,258 | 9,03E-01 | 0,1 % | 0,100 | 0,266 | 7,06E-01 | 0,1 % | 0,122 | 0,277 | 6,58E-01 | 0,1 % | -0,610 | 0,531 | 2,51E-01 |
| | | | 4 | 171070982 | rs191804760 | 0.50466 (6) | - | - | - | T | C | 3,71 | 1,34 | 3,77 | 2,84 | 3,94 | 3,00 | 2,97 | 0,51 | 1 % | -0,475 | 0,694 | 4,93E-01 | - | 13 | 1 % | -0,553 | 0,862 | 5,21E-01 | - | 12 | 1 % | 0,221 | 1,176 | 8,51E-01 | - | 12 | 0,8 % | 0,231 | 2,695 | 5,34E-01 | - | 61 | 0,8 % | 1,120 | 2,130 | 1,71E-02 | - | 61 | 0,8 % | -0,738 | 1,165 | 3,90E-01 | - | 54 | 0,8 % | 0,038 | 0,075 | 6,09E-01 | 0,8 % | 0,029 | 0,078 | 7,14E-01 | 0,8 % | 0,062 | 0,081 | 4,49E-01 | 0,8 % | -0,049 | 0,147 | 7,38E-01 |
| | | | 4 | 171071002 | rs115203131 | 0.18412 (7) | *LINC024.* | - | - | C | G | 1,45 | 1,34 | 3,76 | 3,69 | 3,39 | 3,00 | 2,97 | 0,47 | 1 % | 2,765 | 1,008 | 6,09E-03 | - | 7 | 0 % | 1,955 | 1,772 | 2,70E-01 | - | 4 | 0 % | 6,209 | 2,697 | 2,13E-02 | - | 4 | 0,7 % | -0,010 | 2,755 | 9,78E-01 | - | 62 | 0,7 % | -0,688 | 2,104 | 1,48E-01 | - | 57 | 0,8 % | -0,593 | 1,215 | 4,71E-01 | - | 57 | 0,7 % | 0,098 | 0,075 | 1,88E-01 | 0,7 % | 0,096 | 0,077 | 2,16E-01 | 0,7 % | 0,093 | 0,081 | 2,47E-01 | 0,7 % | -0,001 | 0,151 | 9,96E-01 |
| | | | 4 | 171071112 | rs72708409 | 0.3297 (5) | *LINC024.* | - | - | C | G | 1,66 | 1,34 | 3,75 | 2,94 | 3,39 | 3,00 | 2,97 | 0,48 | 1 % | 2,765 | 1,008 | 6,09E-03 | - | 7 | 0 % | 1,955 | 1,772 | 2,70E-01 | - | 4 | 0 % | 6,209 | 2,697 | 2,13E-02 | - | 4 | 0,7 % | -0,010 | 2,755 | 9,78E-01 | - | 62 | 0,7 % | -0,688 | 2,104 | 1,48E-01 | - | 57 | 0,8 % | -0,593 | 1,215 | 4,71E-01 | - | 57 | 0,7 % | 0,098 | 0,075 | 1,88E-01 | 0,7 % | 0,096 | 0,077 | 2,16E-01 | 0,7 % | 0,093 | 0,081 | 2,47E-01 | 0,7 % | -0,001 | 0,151 | 9,96E-01 |
| | | | 4 | 171071146 | rs72708410 | 0.13454 (5) | *LINC024.* | - | - | A | G | 1,71 | 1,34 | 3,74 | 3,91 | 0,89 | 3,00 | 2,97 | 0,42 | 1 % | 2,765 | 1,008 | 6,09E-03 | - | 7 | 0 % | 1,955 | 1,772 | 2,70E-01 | - | 4 | 0 % | 6,209 | 2,697 | 2,13E-02 | - | 4 | 0,7 % | -0,010 | 2,755 | 9,78E-01 | - | 62 | 0,7 % | -0,688 | 2,104 | 1,48E-01 | - | 57 | 0,8 % | -0,593 | 1,215 | 4,71E-01 | - | 57 | 0,7 % | 0,098 | 0,075 | 1,88E-01 | 0,7 % | 0,096 | 0,077 | 2,16E-01 | 0,7 % | 0,093 | 0,081 | 2,47E-01 | 0,7 % | -0,001 | 0,151 | 9,96E-01 |
| | | | 4 | 171071172 | rs186843742 | 0.13454 (5) | - | - | - | T | A | 3,25 | 1,34 | 3,74 | 3,54 | 3,42 | 3,00 | 2,97 | 0,51 | 0 % | -1,245 | 1,861 | 5,04E-01 | - | ≤3 | 0 % | -1,141 | 2,266 | 6,15E-01 | - | ≤3 | 0 % | -1,066 | 3,301 | 7,47E-01 | - | ≤3 | 0,2 % | -0,662 | 1,268 | 4,01E-01 | - | 12 | 0,2 % | 0,109 | 1,010 | 9,12E-01 | - | 12 | 0,2 % | -1,095 | 0,561 | 5,39E-01 | - | 11 | 0,2 % | 0,216 | 0,151 | 1,51E-01 | 0,2 % | 0,250 | 0,156 | 1,08E-01 | 0,2 % | 0,304 | 0,162 | 6,12E-02 | 0,2 % | -0,108 | 0,304 | 7,22E-01 |
| | | | 4 | 171071211 | rs72708411 | 0.13454 (5) | *LINC024.* | - | - | A | C | 4,02 | 1,34 | 3,73 | 2,40 | 4,24 | 3,00 | 2,97 | 0,35 | 1 % | 2,765 | 1,008 | 6,09E-03 | - | 7 | 0 % | 1,955 | 1,772 | 2,70E-01 | - | 4 | 0 % | 6,209 | 2,697 | 2,13E-02 | - | 4 | 0,7 % | -0,010 | 2,755 | 9,78E-01 | - | 62 | 0,7 % | -0,688 | 2,104 | 1,48E-01 | - | 57 | 0,8 % | -0,593 | 1,215 | 4,71E-01 | - | 57 | 0,7 % | 0,098 | 0,075 | 1,88E-01 | 0,7 % | 0,096 | 0,077 | 2,16E-01 | 0,7 % | 0,093 | 0,081 | 2,47E-01 | 0,7 % | -0,001 | 0,151 | 9,96E-01 |
| | | | 4 | 171071376 | rs112285566 | 0.18347 (6) | - | - | - | G | A | 0,96 | 1,34 | 3,71 | 1,94 | 0,96 | 3,00 | 2,97 | 0,43 | 1 % | 0,276 | 0,648 | 6,71E-01 | - | 16 | 1 % | 0,031 | 0,810 | 9,69E-01 | - | 14 | 1 % | 0,459 | 1,261 | 7,16E-01 | - | 13 | 1,0 % | -0,257 | 2,975 | 4,44E-01 | - | 73 | 1,0 % | 0,083 | 2,337 | 8,46E-01 | - | 71 | 0,9 % | -0,793 | 1,312 | 2,98E-01 | - | 67 | 0,9 % | 0,017 | 0,068 | 7,98E-01 | 0,9 % | -0,015 | 0,070 | 8,30E-01 | 0,9 % | -0,011 | 0,074 | 8,84E-01 | 0,9 % | 0,171 | 0,136 | 2,07E-01 |
| | | | 4 | 171071411 | rs772212036 | 0.00889 (6) | - | - | - | C | G | 0,84 | 1,34 | 3,71 | 2,03 | 0,69 | 3,00 | 2,97 | 0,35 | 0 % | -1,301 | 2,399 | 5,88E-01 | - | ≤3 | 0 % | -1,183 | 2,827 | 6,76E-01 | - | ≤3 | 0 % | -1,067 | 4,396 | 8,08E-01 | - | ≤3 | - | - | - | - | - | - | - | - | - | - | - | - | - | - | - | - | - | - | - | - | - | - | - | - | - | - | - | - | - | - | - | - | - | - |
| | | | 4 | 171071684 | rs7673717 | 0.18412 (7) | - | - | - | G | A | 1,13 | 3,99 | 3,67 | 1,22 | 0,92 | 3,00 | 2,97 | 0,52 | 4 % | 1,834 | 0,379 | 1,34E-06 | ≤3 | 48 | 4 % | 2,601 | 0,487 | 9,51E-08 | ≤3 | 42 | 3 % | 0,572 | 0,923 | 5,36E-01 | - | 30 | 4,9 % | -0,161 | 6,737 | 2,78E-01 | 7 | 375 | 5,0 % | -0,069 | 5,259 | 7,18E-01 | 7 | 360 | 5,0 % | -0,004 | 2,987 | 9,91E-01 | 7 | 349 | 4,7 % | 0,027 | 0,030 | 3,55E-01 | 4,7 % | 0,043 | 0,031 | 1,61E-01 | 4,7 % | 0,021 | 0,032 | 5,03E-01 | 4,7 % | 0,030 | 0,059 | 6,11E-01 |
| | | | 4 | 171071927 | rs568221847 | 0.00167 (6) | - | - | - | T | G | 1,42 | 1,34 | 3,63 | 1,37 | 0,92 | 3,00 | 2,97 | 0,54 | 1 % | 0,357 | 0,751 | 6,34E-01 | - | 11 | 1 % | -0,081 | 1,047 | 9,38E-01 | - | 9 | 1 % | 0,833 | 1,392 | 5,49E-01 | - | 9 | 0,6 % | -0,757 | 2,404 | 6,89E-02 | - | 47 | 0,7 % | -0,639 | 1,914 | 2,21E-01 | - | 47 | 0,7 % | -0,530 | 1,105 | 5,58E-01 | - | 47 | 0,7 % | -0,089 | 0,077 | 2,48E-01 | 0,7 % | -0,091 | 0,080 | 2,58E-01 | 0,7 % | -0,049 | 0,084 | 5,60E-01 | 0,7 % | -0,317 | 0,152 | 3,69E-02 |
| | | | 4 | 171071943 | rs7674265 | 0.18412 (7) | - | - | - | G | A | 0,50 | 1,34 | 3,63 | 1,24 | 0,29 | 3,00 | 2,97 | 0,40 | 4 % | 1,834 | 0,379 | 1,34E-06 | ≤3 | 48 | 4 % | 2,601 | 0,487 | 9,51E-08 | ≤3 | 42 | 3 % | 0,572 | 0,923 | 5,36E-01 | - | 30 | 4,9 % | -0,161 | 6,737 | 2,78E-01 | 7 | 375 | 5,0 % | -0,069 | 5,259 | 7,18E-01 | 7 | 360 | 5,0 % | -0,004 | 2,987 | 9,91E-01 | 7 | 349 | 4,7 % | 0,027 | 0,030 | 3,55E-01 | 4,7 % | 0,043 | 0,031 | 1,61E-01 | 4,7 % | 0,021 | 0,032 | 5,03E-01 | 4,7 % | 0,030 | 0,059 | 6,11E-01 |
| | | | 4 | 171071979 | rs114811062 | 0.32023 (6) | - | - | - | G | A | 0,77 | 1,34 | 3,63 | 2,68 | 0,91 | 3,00 | 2,97 | 0,37 | 5 % | -0,831 | 0,365 | 2,28E-02 | ≤3 | 51 | 5 % | -0,664 | 0,446 | 1,36E-01 | ≤3 | 50 | 5 % | -1,155 | 0,663 | 8,16E-02 | ≤3 | 47 | 4,8 % | 0,141 | 6,279 | 3,77E-01 | 5 | 327 | 4,9 % | 0,343 | 4,879 | 9,47E-02 | 5 | 311 | 4,7 % | -0,508 | 2,717 | 1,68E-01 | ≤3 | 298 | 4,5 % | -0,049 | 0,031 | 1,13E-01 | 4,5 % | -0,053 | 0,032 | 9,47E-02 | 4,5 % | -0,045 | 0,034 | 1,81E-01 | 4,5 % | -0,018 | 0,062 | 7,69E-01 |
| | | | 4 | 171072020 | rs28887169 | 0.13454 (5) | - | - | - | G | T | 1,66 | 1,34 | 3,62 | 2,17 | 4,25 | 3,00 | 2,97 | 0,38 | 4 % | 1,834 | 0,379 | 1,34E-06 | ≤3 | 48 | 4 % | 2,601 | 0,487 | 9,51E-08 | ≤3 | 42 | 3 % | 0,572 | 0,923 | 5,36E-01 | - | 30 | 4,9 % | -0,161 | 6,737 | 2,78E-01 | 7 | 375 | 5,0 % | -0,069 | 5,259 | 7,18E-01 | 7 | 360 | 5,0 % | -0,004 | 2,987 | 9,91E-01 | 7 | 349 | 4,7 % | 0,027 | 0,030 | 3,55E-01 | 4,7 % | 0,043 | 0,031 | 1,61E-01 | 4,7 % | 0,021 | 0,032 | 5,03E-01 | 4,7 % | 0,030 | 0,059 | 6,11E-01 |
| | | | 4 | 171072073 | rs1486735113 | 0.13454 (5) | - | - | - | C | G | 8,03 | 1,34 | 3,61 | 3,29 | 9,56 | 3,00 | 2,97 | 0,80 | 0 % | -1,351 | 2,301 | 5,57E-01 | - | ≤3 | 0 % | -1,193 | 2,766 | 6,66E-01 | - | ≤3 | 0 % | -1,099 | 3,645 | 7,63E-01 | - | ≤3 | - | - | - | - | - | - | - | - | - | - | - | - | - | - | - | - | - | - | - | - | - | - | - | - | - | - | - | - | - | - | - | - | - | - |
| | | | 4 | 171072369 | rs117014432 | 0.13454 (5) | - | - | - | C | T | 8,06 | 1,34 | 3,57 | 2,74 | 10,13 | 3,00 | 2,97 | 0,39 | 1 % | 1,809 | 1,023 | 7,70E-02 | - | 6 | 1 % | 3,326 | 1,230 | 6,85E-03 | - | 6 | 0 % | -1,076 | 2,792 | 7,00E-01 | - | ≤3 | 0,3 % | -0,435 | 1,650 | 4,73E-01 | - | 22 | 0,3 % | 0,054 | 1,313 | 9,44E-01 | - | 22 | 0,3 % | -0,617 | 0,742 | 6,47E-01 | - | 21 | 0,2 % | 0,141 | 0,128 | 2,69E-01 | 0,2 % | 0,132 | 0,132 | 3,17E-01 | 0,2 % | 0,205 | 0,139 | 1,39E-01 | 0,2 % | 0,051 | 0,249 | 8,37E-01 |
| | | | 4 | 171072671 | rs112263741 | 0.27853 (6) | - | - | - | A | G | 1,92 | 1,34 | 3,54 | 0,78 | 1,77 | 3,00 | 2,97 | 0,32 | 3 % | 2,100 | 0,433 | 1,25E-06 | ≤3 | 36 | 3 % | 2,963 | 0,562 | 1,36E-07 | ≤3 | 31 | 2 % | 1,368 | 1,113 | 2,19E-01 | - | 22 | 3,3 % | 0,087 | 5,540 | 6,30E-01 | ≤3 | 256 | 3,3 % | 0,101 | 4,293 | 6,64E-01 | ≤3 | 242 | 3,3 % | 0,366 | 2,436 | 3,72E-01 | ≤3 | 234 | 3,3 % | -0,0002 | 0,035 | 9,95E-01 | 3,3 % | 0,012 | 0,036 | 7,38E-01 | 3,3 % | -0,005 | 0,038 | 8,96E-01 | 3,3 % | 0,011 | 0,070 | 8,74E-01 |
| | | | 4 | 171072680 | rs187224116 | 0.18412 (7) | - | - | - | C | T | 0,22 | 1,34 | 3,54 | 0,71 | 0,63 | 3,00 | 2,97 | 0,33 | 0 % | -1,489 | 2,183 | 4,95E-01 | - | ≤3 | 0 % | -1,249 | 2,645 | 6,37E-01 | - | ≤3 | 0 % | -1,164 | 3,272 | 7,22E-01 | - | ≤3 | 0,1 % | -0,970 | 0,917 | 3,74E-01 | - | 6 | 0,1 % | -0,805 | 0,730 | 5,57E-01 | - | 6 | 0,1 % | -0,690 | 0,422 | 7,71E-01 | - | 6 | 0,1 % | -0,061 | 0,193 | 7,53E-01 | 0,1 % | -0,043 | 0,201 | 8,32E-01 | 0,1 % | -0,079 | 0,210 | 7,08E-01 | 0,1 % | -0,217 | 0,375 | 5,63E-01 |
| | | | 4 | 171072692 | rs920321548 | 0.27792 (6) | - | - | - | T | C | 10,04 | 1,34 | 3,53 | 0,80 | 13,30 | 3,00 | 2,97 | 0,32 | 0 % | 10,752 | 3,444 | 1,80E-03 | - | ≤3 | 0 % | 18,322 | 4,403 | 3,17E-05 | - | ≤3 | - | - | - | - | - | - | - | - | - | - | - | - | - | - | - | - | - | - | - | - | - | - | - | - | - | - | - | - | - | - | - | - | - | - | - | - | - | - | - | - |
| | | | 4 | 171072838 | rs182790955 | 0.18412 (7) | - | - | - | T | C | 3,65 | 1,34 | 3,52 | 0,72 | 3,85 | 3,00 | 2,97 | 0,18 | 3 % | 0,817 | 0,468 | 8,12E-02 | - | 32 | 3 % | 1,004 | 0,611 | 1,00E-01 | - | 28 | 3 % | 1,261 | 0,897 | 1,60E-01 | - | 25 | 2,1 % | -0,073 | 4,561 | 7,38E-01 | ≤3 | 167 | 2,1 % | -0,472 | 3,516 | 9,73E-02 | ≤3 | 156 | 2,2 % | 0,164 | 2,029 | 7,40E-01 | ≤3 | 156 | 2,3 % | 0,039 | 0,042 | 3,58E-01 | 2,3 % | 0,054 | 0,043 | 2,12E-01 | 2,3 % | 0,053 | 0,045 | 2,40E-01 | 2,3 % | 0,017 | 0,083 | 8,36E-01 |
| | | | 4 | 171072874 | rs751170265 | 0.18412 (7) | - | - | - | C | T | 2,50 | 1,34 | 3,51 | 0,79 | 3,46 | 3,00 | 2,97 | 0,19 | 0 % | -1,257 | 1,891 | 5,06E-01 | - | ≤3 | 0 % | -1,150 | 2,348 | 6,25E-01 | - | ≤3 | 0 % | -1,075 | 3,211 | 7,38E-01 | - | ≤3 | 0,1 % | 0,740 | 1,105 | 4,13E-01 | - | 10 | 0,1 % | 0,981 | 0,834 | 4,13E-01 | - | 9 | 0,1 % | -0,695 | 0,454 | 7,52E-01 | - | 8 | 0,2 % | 0,041 | 0,144 | 7,77E-01 | 0,2 % | 0,105 | 0,148 | 4,80E-01 | 0,2 % | 0,057 | 0,155 | 7,15E-01 | 0,2 % | 0,183 | 0,285 | 5,22E-01 |
| | | | 4 | 171073135 | rs549726823 | 0.13454 (5) | - | - | - | G | GT | 3,04 | 1,34 | 3,48 | 0,62 | 3,81 | 3,00 | 2,97 | 0,19 | 0 % | -1,285 | 2,441 | 5,99E-01 | - | ≤3 | 0 % | -1,152 | 3,035 | 7,04E-01 | - | ≤3 | 0 % | -1,081 | 4,048 | 7,89E-01 | - | ≤3 | 0,1 % | 0,791 | 0,864 | 4,95E-01 | - | 6 | 0,1 % | -0,418 | 0,628 | 7,93E-01 | - | 5 | 0,1 % | -0,303 | 0,363 | 9,12E-01 | - | 5 | 0,1 % | 0,019 | 0,205 | 9,27E-01 | 0,1 % | 0,089 | 0,212 | 6,74E-01 | 0,1 % | 0,172 | 0,221 | 4,36E-01 | 0,1 % | -0,460 | 0,434 | 2,89E-01 |
| | | | 4 | 171073180 | rs114541370 | 0.13454 (5) | *LINC024.* | - | - | A | G | 2,64 | 1,34 | 3,47 | 1,35 | 3,24 | 3,00 | 2,97 | 0,17 | 1 % | 2,765 | 1,008 | 6,09E-03 | - | 7 | 0 % | 1,955 | 1,772 | 2,70E-01 | - | 4 | 0 % | 6,209 | 2,697 | 2,13E-02 | - | 4 | 0,7 % | -0,010 | 2,755 | 9,78E-01 | - | 62 | 0,7 % | -0,688 | 2,104 | 1,48E-01 | - | 57 | 0,8 % | -0,593 | 1,215 | 4,71E-01 | - | 57 | 0,7 % | 0,098 | 0,075 | 1,88E-01 | 0,7 % | 0,096 | 0,077 | 2,16E-01 | 0,7 % | 0,093 | 0,081 | 2,47E-01 | 0,7 % | -0,001 | 0,151 | 9,95E-01 |
| | | | 4 | 171073275 | rs188150427 | 0.73862 (3a | - | - | - | A | T | 1,28 | 1,34 | 3,46 | 1,20 | 3,33 | 3,00 | 2,97 | 0,06 | 3 % | 0,817 | 0,468 | 8,12E-02 | - | 32 | 3 % | 1,004 | 0,611 | 1,00E-01 | - | 28 | 3 % | 1,261 | 0,897 | 1,60E-01 | - | 25 | 2,2 % | -0,075 | 4,561 | 7,33E-01 | ≤3 | 167 | 2,1 % | -0,474 | 3,516 | 9,54E-02 | ≤3 | 156 | 2,2 % | 0,168 | 2,028 | 7,34E-01 | ≤3 | 156 | 2,3 % | 0,036 | 0,042 | 3,97E-01 | 2,3 % | 0,050 | 0,043 | 2,52E-01 | 2,3 % | 0,049 | 0,045 | 2,79E-01 | 2,3 % | 0,019 | 0,083 | 8,20E-01 |
| | | | 4 | 171073291 | rs72708412 | 0.60906 (4) | *LINC024.* | - | - | C | T | 0,28 | 3,99 | 3,46 | 1,48 | 0,63 | 3,00 | 2,97 | 0,07 | 1 % | 2,765 | 1,008 | 6,09E-03 | - | 7 | 0 % | 1,955 | 1,772 | 2,70E-01 | - | 4 | 0 % | 6,209 | 2,697 | 2,13E-02 | - | 4 | 0,7 % | -0,010 | 2,755 | 9,78E-01 | - | 62 | 0,7 % | -0,688 | 2,104 | 1,48E-01 | - | 57 | 0,8 % | -0,593 | 1,215 | 4,71E-01 | - | 57 | 0,7 % | 0,098 | 0,075 | 1,88E-01 | 0,7 % | 0,096 | 0,077 | 2,16E-01 | 0,7 % | 0,093 | 0,081 | 2,47E-01 | 0,7 % | -0,001 | 0,151 | 9,95E-01 |
| | | | 4 | 171073292 | rs146667288 | 0.60906 (4) | - | - | - | G | A | 0,11 | 3,99 | 3,46 | 1,47 | 0,14 | 3,00 | 2,97 | 0,07 | 0 % | 0,371 | 1,328 | 7,80E-01 | - | ≤3 | 0 % | 1,070 | 1,530 | 4,84E-01 | - | ≤3 | 0 % | -1,106 | 2,769 | 6,89E-01 | - | ≤3 | 0,1 % | -1,090 | 1,114 | 2,25E-01 | - | 10 | 0,1 % | -0,940 | 0,887 | 4,04E-01 | - | 10 | 0,1 % | -0,878 | 0,511 | 6,53E-01 | - | 10 | 0,1 % | 0,292 | 0,198 | 1,41E-01 | 0,1 % | 0,371 | 0,206 | 7,09E-02 | 0,1 % | 0,488 | 0,215 | 2,30E-02 | 0,1 % | -0,263 | 0,395 | 5,06E-01 |
| | | | 4 | 171073324 | rs72708414 | 0.60906 (4) | *LINC024.* | - | - | G | T | 0,43 | 3,99 | 3,46 | 1,48 | 3,00 | 3,00 | 2,97 | 0,08 | 1 % | 2,765 | 1,008 | 6,09E-03 | - | 7 | 0 % | 1,955 | 1,772 | 2,70E-01 | - | 4 | 0 % | 6,209 | 2,697 | 2,13E-02 | - | 4 | 0,7 % | -0,010 | 2,755 | 9,78E-01 | - | 62 | 0,7 % | -0,688 | 2,104 | 1,48E-01 | - | 57 | 0,8 % | -0,593 | 1,215 | 4,71E-01 | - | 57 | 0,7 % | 0,098 | 0,075 | 1,88E-01 | 0,7 % | 0,096 | 0,077 | 2,16E-01 | 0,7 % | 0,093 | 0,081 | 2,47E-01 | 0,7 % | -0,001 | 0,151 | 9,95E-01 |
| | | | 4 | 171073509 | rs182602760 | 0.32977 (3a | - | - | - | C | T | 0,70 | 3,99 | 3,43 | 1,38 | 0,70 | 3,00 | 2,97 | 0,29 | 0 % | -1,207 | 1,989 | 5,44E-01 | - | ≤3 | 0 % | -1,162 | 2,236 | 6,03E-01 | - | ≤3 | 0 % | -1,052 | 4,135 | 7,99E-01 | - | ≤3 | - | - | - | - | - | - | - | - | - | - | - | - | - | - | - | - | - | - | - | - | - | - | - | - | - | - | - | - | - | - | - | - | - | - |
| | | | 4 | 171073541 | rs148166479 | 0.86 (3b) | *LINC024.* | - | - | G | A | 0,58 | 3,99 | 3,43 | 1,37 | 0,73 | 3,00 | 2,97 | 0,37 | 1 % | 2,765 | 1,008 | 6,09E-03 | - | 7 | 0 % | 1,955 | 1,772 | 2,70E-01 | - | 4 | 0 % | 6,209 | 2,697 | 2,13E-02 | - | 4 | 0,7 % | -0,010 | 2,755 | 9,78E-01 | - | 62 | 0,7 % | -0,688 | 2,104 | 1,48E-01 | - | 57 | 0,8 % | -0,593 | 1,215 | 4,71E-01 | - | 57 | 0,7 % | 0,098 | 0,075 | 1,88E-01 | 0,7 % | 0,096 | 0,077 | 2,16E-01 | 0,7 % | 0,093 | 0,081 | 2,47E-01 | 0,7 % | -0,001 | 0,151 | 9,95E-01 |
| | | | 4 | 171073579 | rs562527026 | 1 (5) | *LINC024.* | - | - | T | TC | 4,90 | 1,34 | 3,43 | 1,91 | 5,81 | 3,00 | 2,97 | 0,37 | 1 % | 2,765 | 1,008 | 6,09E-03 | - | 7 | 0 % | 1,955 | 1,772 | 2,70E-01 | - | 4 | 0 % | 6,209 | 2,697 | 2,13E-02 | - | 4 | 0,8 % | 0,029 | 2,758 | 9,35E-01 | - | 62 | 0,7 % | -0,684 | 2,106 | 1,50E-01 | - | 57 | 0,8 % | -0,593 | 1,216 | 4,71E-01 | - | 57 | 0,7 % | 0,111 | 0,075 | 1,38E-01 | 0,7 % | 0,110 | 0,077 | 1,56E-01 | 0,7 % | 0,104 | 0,081 | 1,95E-01 | 0,7 % | 0,009 | 0,151 | 9,54E-01 |
| | | | 4 | 171073660 | rs72708415 | 0.94917 (5) | *LINC024.* | - | - | T | G | 3,30 | 1,34 | 3,42 | 0,76 | 4,79 | 3,00 | 2,97 | 0,45 | 1 % | 2,765 | 1,008 | 6,09E-03 | - | 7 | 0 % | 1,955 | 1,772 | 2,70E-01 | - | 4 | 0 % | 6,209 | 2,697 | 2,13E-02 | - | 4 | 0,7 % | -0,010 | 2,755 | 9,78E-01 | - | 62 | 0,7 % | -0,688 | 2,104 | 1,48E-01 | - | 57 | 0,8 % | -0,593 | 1,215 | 4,71E-01 | - | 57 | 0,7 % | 0,098 | 0,075 | 1,88E-01 | 0,7 % | 0,096 | 0,077 | 2,16E-01 | 0,7 % | 0,093 | 0,081 | 2,47E-01 | 0,7 % | -0,001 | 0,151 | 9,95E-01 |
| | | | 4 | 171073706 | rs72708416 | 1 (5) | *LINC024.* | - | - | T | A | 1,37 | 1,34 | 3,41 | 0,87 | 1,61 | 3,00 | 2,97 | 0,36 | 1 % | 2,765 | 1,008 | 6,09E-03 | - | 7 | 0 % | 1,955 | 1,772 | 2,70E-01 | - | 4 | 0 % | 6,209 | 2,697 | 2,13E-02 | - | 4 | 0,7 % | -0,010 | 2,755 | 9,78E-01 | - | 62 | 0,7 % | -0,688 | 2,104 | 1,48E-01 | - | 57 | 0,8 % | -0,593 | 1,215 | 4,71E-01 | - | 57 | 0,7 % | 0,098 | 0,075 | 1,88E-01 | 0,7 % | 0,096 | 0,077 | 2,16E-01 | 0,7 % | 0,093 | 0,081 | 2,47E-01 | 0,7 % | -0,001 | 0,151 | 9,95E-01 |
| | | | 4 | 171073757 | rs564623572 | 0.58955 (5) | - | - | - | C | T | 3,05 | 1,34 | 3,40 | 0,96 | 4,07 | 3,00 | 2,97 | 0,27 | 1 % | -0,648 | 0,815 | 4,27E-01 | - | 11 | 1 % | -0,196 | 1,003 | 8,45E-01 | - | 11 | 1 % | -1,106 | 1,524 | 4,68E-01 | - | 10 | 0,9 % | 0,178 | 2,953 | 6,00E-01 | ≤3 | 67 | 0,9 % | -0,128 | 2,268 | 7,72E-01 | ≤3 | 62 | 0,9 % | 0,344 | 1,299 | 6,55E-01 | ≤3 | 61 | 0,8 % | -0,035 | 0,071 | 6,22E-01 | 0,8 % | -0,068 | 0,073 | 3,55E-01 | 0,8 % | -0,082 | 0,077 | 2,87E-01 | 0,8 % | 0,068 | 0,141 | 6,31E-01 |
| | | | 4 | 171073767 | rs778905995 | 0.79101 (5) | - | - | - | A | G | 2,12 | 1,34 | 3,40 | 1,17 | 1,90 | 3,00 | 2,97 | 0,35 | 0 % | -1,298 | 1,778 | 4,66E-01 | - | ≤3 | 0 % | -1,205 | 2,064 | 5,59E-01 | - | ≤3 | 0 % | -1,073 | 3,153 | 7,34E-01 | - | ≤3 | 0,1 % | 0,539 | 0,870 | 6,39E-01 | - | 6 | 0,1 % | 1,953 | 0,693 | 1,76E-01 | - | 6 | 0,1 % | -0,937 | 0,364 | 7,33E-01 | - | 5 | 0,1 % | -0,252 | 0,199 | 2,07E-01 | 0,1 % | -0,255 | 0,207 | 2,18E-01 | 0,1 % | -0,249 | 0,216 | 2,51E-01 | 0,1 % | -0,453 | 0,391 | 2,47E-01 |
| | | | 4 | 171073804 | rs74623553 | 0.32 (5) | - | - | - | A | G | 0,58 | 1,34 | 3,40 | 0,92 | 0,79 | 3,00 | 2,97 | 0,64 | 3 % | 0,817 | 0,468 | 8,12E-02 | - | 32 | 3 % | 1,004 | 0,611 | 1,00E-01 | - | 28 | 3 % | 1,261 | 0,897 | 1,60E-01 | - | 25 | 2,2 % | -0,075 | 4,561 | 7,33E-01 | ≤3 | 167 | 2,1 % | -0,474 | 3,516 | 9,54E-02 | ≤3 | 156 | 2,2 % | 0,168 | 2,028 | 7,34E-01 | ≤3 | 156 | 2,3 % | 0,036 | 0,042 | 3,97E-01 | 2,3 % | 0,050 | 0,043 | 2,52E-01 | 2,3 % | 0,049 | 0,045 | 2,79E-01 | 2,3 % | 0,019 | 0,083 | 8,20E-01 |
| MAF<5% | *LINC02382* | 4:171078001-171082000 | 4 | 171078290 | rs564702927 | 0.15078 (6) | - | - | - | A | G | 5,97 | 1,34 | 3,02 | 1,73 | 4,19 | 3,00 | 2,97 | 4,17 | 1 % | 0,648 | 1,011 | 5,21E-01 | - | 6 | 0 % | 0,498 | 1,315 | 7,05E-01 | - | 5 | 0 % | -1,071 | 2,523 | 6,71E-01 | - | 4 | 0,4 % | -0,349 | 1,985 | 4,88E-01 | - | 32 | 0,4 % | -0,366 | 1,553 | 5,70E-01 | - | 31 | 0,4 % | 1,021 | 0,897 | 3,60E-01 | - | 31 | 0,4 % | -0,071 | 0,103 | 4,87E-01 | 0,4 % | -0,039 | 0,106 | 7,14E-01 | 0,4 % | -0,044 | 0,111 | 6,90E-01 | 0,4 % | 0,052 | 0,206 | 8,02E-01 |
| | | | 4 | 171078453 | rs72708442 | 0.2714 (5) | *LINC024.* | - | - | G | A | 1,51 | 1,34 | 3,02 | 3,47 | 2,71 | 3,00 | 2,97 | 4,31 | 1 % | 2,765 | 1,008 | 6,09E-03 | - | 7 | 0 % | 1,955 | 1,772 | 2,70E-01 | - | 4 | 0 % | 6,209 | 2,697 | 2,13E-02 | - | 4 | 0,7 % | -0,022 | 2,757 | 9,52E-01 | - | 62 | 0,7 % | -0,699 | 2,106 | 1,41E-01 | - | 57 | 0,8 % | -0,604 | 1,216 | 4,63E-01 | - | 57 | 0,7 % | 0,109 | 0,074 | 1,45E-01 | 0,7 % | 0,108 | 0,077 | 1,61E-01 | 0,7 % | 0,101 | 0,080 | 2,06E-01 | 0,7 % | 0,032 | 0,150 | 8,32E-01 |
| | | | 4 | 171078467 | - | - | - | - | - | C | T | 0,49 | 1,34 | 3,02 | 2,95 | 0,44 | 3,00 | 2,97 | 4,02 | 0 % | 4,754 | 2,456 | 5,29E-02 | - | ≤3 | 0 % | 8,736 | 3,142 | 5,43E-03 | - | ≤3 | - | - | - | - | - | - | - | - | - | - | - | - | - | - | - | - | - | - | - | - | - | - | - | - | - | - | - | - | - | - | - | - | - | - | - | - | - | - | - | - |
| | | | 4 | 171078729 | rs72708444 | 0.13454 (5) | *LINC024.* | - | - | G | T | 11,11 | 5,42 | 3,01 | 5,37 | 9,70 | 3,00 | 2,97 | 2,11 | 1 % | 2,765 | 1,008 | 6,09E-03 | - | 7 | 0 % | 1,955 | 1,772 | 2,70E-01 | - | 4 | 0 % | 6,209 | 2,697 | 2,13E-02 | - | 4 | 0,7 % | -0,022 | 2,758 | 9,51E-01 | - | 62 | 0,7 % | -0,699 | 2,106 | 1,41E-01 | - | 57 | 0,8 % | -0,604 | 1,216 | 4,63E-01 | - | 57 | 0,7 % | 0,109 | 0,074 | 1,45E-01 | 0,7 % | 0,108 | 0,077 | 1,61E-01 | 0,7 % | 0,101 | 0,080 | 2,06E-01 | 0,7 % | 0,032 | 0,150 | 8,32E-01 |
| | | | 4 | 171078809 | rs72708445 | 0.13454 (5) | *LINC024.* | - | - | C | T | 3,83 | 3,99 | 3,01 | 2,26 | 6,02 | 3,00 | 2,97 | 3,44 | 1 % | 2,765 | 1,008 | 6,09E-03 | - | 7 | 0 % | 1,955 | 1,772 | 2,70E-01 | - | 4 | 0 % | 6,209 | 2,697 | 2,13E-02 | - | 4 | 0,7 % | -0,023 | 2,758 | 9,50E-01 | - | 62 | 0,7 % | -0,700 | 2,106 | 1,41E-01 | - | 57 | 0,8 % | -0,604 | 1,216 | 4,62E-01 | - | 57 | 0,7 % | 0,109 | 0,074 | 1,44E-01 | 0,7 % | 0,108 | 0,077 | 1,61E-01 | 0,7 % | 0,101 | 0,080 | 2,06E-01 | 0,7 % | 0,032 | 0,150 | 8,32E-01 |
| | | | 4 | 171078895 | rs183300218 | 0.23618 (6) | - | - | - | G | T | 8,20 | 3,99 | 3,00 | 3,80 | 8,52 | 3,00 | 2,97 | 1,68 | 0 % | 6,637 | 2,798 | 1,77E-02 | - | ≤3 | 0 % | 9,429 | 3,250 | 3,72E-03 | - | ≤3 | - | - | - | - | - | - | 0,1 % | 1,637 | 1,179 | 5,36E-02 | - | 11 | 0,1 % | 1,061 | 0,851 | 3,67E-01 | - | 9 | 0,1 % | -0,746 | 0,465 | 7,29E-01 | - | 8 | 0,1 % | -0,248 | 0,224 | 2,69E-01 | 0,1 % | -0,324 | 0,233 | 1,64E-01 | 0,1 % | -0,239 | 0,244 | 3,28E-01 | 0,1 % | -0,326 | 0,417 | 4,35E-01 |
| | | | 4 | 171079293 | rs1373947513 | 0.17627 (5) | - | - | - | A | G | 6,09 | 1,34 | 3,00 | 2,69 | 5,50 | 3,00 | 2,97 | 1,53 | 0 % | -1,314 | 2,378 | 5,81E-01 | - | ≤3 | 0 % | -1,151 | 3,054 | 7,06E-01 | - | ≤3 | 0 % | -1,097 | 3,733 | 7,69E-01 | - | ≤3 | - | - | - | - | - | - | - | - | - | - | - | - | - | - | - | - | - | - | - | - | - | - | - | - | - | - | - | - | - | - | - | - | - | - |
| | | | 4 | 171079328 | rs72708446 | 0.03 (5) | *LINC024.* | - | - | C | G | 5,56 | 3,99 | 2,99 | 3,21 | 9,81 | 3,00 | 2,97 | 1,58 | 1 % | 2,765 | 1,008 | 6,09E-03 | - | 7 | 0 % | 1,955 | 1,772 | 2,70E-01 | - | 4 | 0 % | 6,209 | 2,697 | 2,13E-02 | - | 4 | 0,8 % | -0,024 | 2,759 | 9,47E-01 | - | 62 | 0,7 % | -0,700 | 2,107 | 1,40E-01 | - | 57 | 0,8 % | -0,605 | 1,217 | 4,62E-01 | - | 57 | 0,7 % | 0,109 | 0,074 | 1,44E-01 | 0,7 % | 0,108 | 0,077 | 1,61E-01 | 0,7 % | 0,102 | 0,080 | 2,06E-01 | 0,7 % | 0,032 | 0,150 | 8,31E-01 |
| | | | 4 | 171079466 | rs529624103 | 0.36782 (6) | *LINC024.* | - | - | G | GTA | 2,52 | 3,99 | 2,99 | 1,99 | 4,45 | 3,00 | 2,97 | 2,10 | 1 % | 2,765 | 1,008 | 6,09E-03 | - | 7 | 0 % | 1,955 | 1,772 | 2,70E-01 | - | 4 | 0 % | 6,209 | 2,697 | 2,13E-02 | - | 4 | 0,8 % | -0,024 | 2,758 | 9,48E-01 | - | 62 | 0,7 % | -0,700 | 2,107 | 1,40E-01 | - | 57 | 0,8 % | -0,605 | 1,217 | 4,62E-01 | - | 57 | 0,7 % | 0,109 | 0,074 | 1,44E-01 | 0,7 % | 0,108 | 0,077 | 1,61E-01 | 0,7 % | 0,102 | 0,080 | 2,06E-01 | 0,7 % | 0,032 | 0,150 | 8,31E-01 |
| | | | 4 | 171079487 | rs564734422 | 0.55602 (6) | - | - | - | G | A | 3,67 | 3,99 | 2,99 | 1,62 | 4,56 | 3,00 | 2,97 | 2,26 | 0 % | -1,317 | 2,356 | 5,76E-01 | - | ≤3 | 0 % | -1,187 | 2,785 | 6,70E-01 | - | ≤3 | 0 % | -1,074 | 4,099 | 7,93E-01 | - | ≤3 | - | - | - | - | - | - | - | - | - | - | - | - | - | - | - | - | - | - | - | - | - | - | - | - | - | - | - | - | - | - | - | - | - | - |
| | | | 4 | 171079680 | - | - | - | - | - | G | T | 0,97 | 3,99 | 2,99 | 2,69 | 1,52 | 3,00 | 2,97 | 1,85 | 0 % | -1,188 | 2,792 | 6,70E-01 | - | ≤3 | 0 % | -1,123 | 3,301 | 7,34E-01 | - | ≤3 | 0 % | -1,044 | 5,470 | 8,49E-01 | - | ≤3 | - | - | - | - | - | - | - | - | - | - | - | - | - | - | - | - | - | - | - | - | - | - | - | - | - | - | - | - | - | - | - | - | - | - |
| | | | 4 | 171079876 | rs548276922 | 0.38564 (6) | - | - | - | A | AAA | 0,46 | 3,99 | 2,98 | 1,76 | 0,96 | 3,00 | 2,97 | 3,14 | 1 % | 2,765 | 1,008 | 6,09E-03 | - | 7 | 0 % | 1,955 | 1,772 | 2,70E-01 | - | 4 | 0 % | 6,209 | 2,697 | 2,13E-02 | - | 4 | - | - | - | - | - | - | - | - | - | - | - | - | - | - | - | - | - | - | - | - | - | - | - | - | - | - | - | - | - | - | - | - | - | - |
| | | | 4 | 171079915 | rs567891732 | 0.18412 (7) | - | - | - | T | C | 2,22 | 1,34 | 2,98 | 1,65 | 1,42 | 3,00 | 2,97 | 3,11 | 0 % | -1,195 | 2,903 | 6,81E-01 | - | ≤3 | 0 % | -1,095 | 4,052 | 7,87E-01 | - | ≤3 | 0 % | -1,056 | 5,846 | 8,57E-01 | - | ≤3 | 0,2 % | 0,688 | 1,159 | 4,25E-01 | - | 11 | 0,2 % | 0,766 | 0,879 | 5,01E-01 | - | 10 | 0,1 % | -0,689 | 0,482 | 7,40E-01 | - | 9 | 0,1 % | -0,064 | 0,186 | 7,32E-01 | 0,1 % | -0,027 | 0,192 | 8,87E-01 | 0,1 % | -0,058 | 0,201 | 7,72E-01 | 0,1 % | -0,381 | 0,362 | 2,93E-01 |
| | | | 4 | 171079937 | rs977866951 | 0.18412 (7) | - | - | - | A | G | 4,65 | 1,34 | 2,98 | 1,67 | 3,34 | 3,00 | 2,97 | 2,77 | 0 % | -1,203 | 2,764 | 6,63E-01 | - | ≤3 | 0 % | -1,123 | 3,400 | 7,41E-01 | - | ≤3 | 0 % | -1,070 | 4,668 | 8,19E-01 | - | ≤3 | 0,1 % | -0,679 | 1,114 | 4,49E-01 | - | 10 | 0,1 % | -0,570 | 0,887 | 6,13E-01 | - | 10 | 0,1 % | -0,421 | 0,513 | 8,29E-01 | - | 10 | 0,1 % | -0,074 | 0,221 | 7,36E-01 | 0,1 % | -0,197 | 0,228 | 3,89E-01 | 0,1 % | -0,311 | 0,239 | 1,93E-01 | 0,1 % | 0,598 | 0,429 | 1,64E-01 |
| | | | 4 | 171080113 | rs17056439 | 0.18412 (7) | - | - | - | C | T | 0,59 | 5,42 | 2,98 | 1,87 | 0,56 | 3,00 | 2,97 | 2,87 | 5 % | 1,671 | 0,370 | 6,30E-06 | ≤3 | 52 | 5 % | 2,388 | 0,474 | 4,77E-07 | ≤3 | 46 | 4 % | 0,420 | 0,884 | 6,35E-01 | - | 34 | 5,0 % | -0,165 | 6,768 | 2,64E-01 | 7 | 380 | 5,0 % | -0,072 | 5,284 | 7,03E-01 | 7 | 365 | 5,1 % | -0,007 | 3,001 | 9,84E-01 | 7 | 354 | 4,8 % | 0,025 | 0,030 | 4,05E-01 | 4,8 % | 0,042 | 0,031 | 1,72E-01 | 4,8 % | 0,019 | 0,032 | 5,44E-01 | 4,8 % | 0,022 | 0,059 | 7,08E-01 |
| | | | 4 | 171080360 | rs116541898 | 0.18412 (7) | - | - | - | G | A | 12,22 | 1,34 | 2,97 | 0,81 | 12,83 | 3,00 | 2,97 | 1,71 | 0 % | 0,465 | 1,310 | 7,22E-01 | - | 4 | 0 % | 1,238 | 1,548 | 4,24E-01 | - | 4 | 0 % | -1,069 | 2,702 | 6,93E-01 | - | ≤3 | 0,5 % | 0,276 | 2,123 | 5,57E-01 | ≤3 | 32 | 0,5 % | 0,460 | 1,644 | 4,50E-01 | ≤3 | 30 | 0,5 % | 1,111 | 0,934 | 2,99E-01 | ≤3 | 29 | 0,5 % | 0,205 | 0,087 | 1,87E-02 | 0,5 % | 0,207 | 0,090 | 2,16E-02 | 0,5 % | 0,193 | 0,094 | 4,01E-02 | 0,5 % | 0,183 | 0,179 | 3,07E-01 |
| | | | 4 | 171080367 | rs1508514 | 0.18412 (7) | - | - | - | C | T | 8,67 | 1,34 | 2,97 | 0,91 | 6,65 | 3,00 | 2,97 | 1,86 | 5 % | 1,671 | 0,370 | 6,30E-06 | ≤3 | 52 | 5 % | 2,388 | 0,474 | 4,77E-07 | ≤3 | 46 | 4 % | 0,420 | 0,884 | 6,35E-01 | - | 34 | 5,0 % | -0,165 | 6,767 | 2,64E-01 | 7 | 380 | 5,0 % | -0,072 | 5,283 | 7,03E-01 | 7 | 365 | 5,1 % | -0,006 | 3,001 | 9,85E-01 | 7 | 354 | 4,8 % | 0,025 | 0,030 | 4,06E-01 | 4,8 % | 0,042 | 0,031 | 1,72E-01 | 4,8 % | 0,019 | 0,032 | 5,44E-01 | 4,8 % | 0,022 | 0,059 | 7,09E-01 |
| | | | 4 | 171080589 | - | - | - | - | - | T | C | 6,68 | 1,34 | 2,97 | 1,07 | 6,75 | 3,00 | 2,97 | 2,58 | 0 % | -1,136 | 3,143 | 7,18E-01 | - | ≤3 | 0 % | -1,076 | 4,053 | 7,91E-01 | - | ≤3 | 0 % | -1,038 | 5,827 | 8,59E-01 | - | ≤3 | - | - | - | - | - | - | - | - | - | - | - | - | - | - | - | - | - | - | - | - | - | - | - | - | - | - | - | - | - | - | - | - | - | - |
| | | | 4 | 171080753 | rs1848431 | 0 (5) | - | - | - | T | A | 0,29 | 1,34 | 2,97 | 3,72 | 0,53 | 3,00 | 2,97 | 2,02 | 5 % | 1,671 | 0,370 | 6,30E-06 | ≤3 | 52 | 5 % | 2,388 | 0,474 | 4,77E-07 | ≤3 | 46 | 4 % | 0,420 | 0,884 | 6,35E-01 | - | 34 | 5,0 % | -0,165 | 6,767 | 2,64E-01 | 7 | 380 | 5,0 % | -0,072 | 5,283 | 7,04E-01 | 7 | 365 | 5,1 % | -0,006 | 3,001 | 9,85E-01 | 7 | 354 | 4,8 % | 0,025 | 0,030 | 4,06E-01 | 4,8 % | 0,042 | 0,031 | 1,72E-01 | 4,8 % | 0,019 | 0,032 | 5,43E-01 | 4,8 % | 0,022 | 0,059 | 7,10E-01 |
| | | | 4 | 171080962 | rs9684852 | 0 (6) | - | - | - | A | G | 0,11 | 1,34 | 2,96 | 1,34 | 0,17 | 3,00 | 2,97 | 1,21 | 1 % | 2,765 | 1,008 | 6,09E-03 | - | 7 | 0 % | 1,955 | 1,772 | 2,70E-01 | - | 4 | 0 % | 6,209 | 2,697 | 2,13E-02 | - | 4 | 0,8 % | -0,026 | 2,760 | 9,43E-01 | - | 62 | 0,7 % | -0,702 | 2,108 | 1,39E-01 | - | 57 | 0,8 % | -0,606 | 1,217 | 4,60E-01 | - | 57 | 0,7 % | 0,109 | 0,074 | 1,44E-01 | 0,7 % | 0,108 | 0,077 | 1,61E-01 | 0,7 % | 0,102 | 0,080 | 2,06E-01 | 0,7 % | 0,032 | 0,150 | 8,31E-01 |
| | | | 4 | 171080970 | rs9684968 | 0 (6) | - | - | - | T | A | 0,23 | 1,34 | 2,96 | 1,22 | 0,94 | 3,00 | 2,97 | 1,14 | 1 % | 2,765 | 1,008 | 6,09E-03 | - | 7 | 0 % | 1,955 | 1,772 | 2,70E-01 | - | 4 | 0 % | 6,209 | 2,697 | 2,13E-02 | - | 4 | - | - | - | - | - | - | - | - | - | - | - | - | - | - | - | - | - | - | - | - | - | - | - | - | - | - | - | - | - | - | - | - | - | - |
| | | | 4 | 171081207 | rs566183872 | 0.20016 (6) | - | - | - | G | GT | 3,24 | 1,34 | 2,96 | 1,72 | 4,75 | 3,00 | 2,97 | 2,56 | 3 % | 1,184 | 0,429 | 5,74E-03 | - | 39 | 3 % | 1,117 | 0,581 | 5,44E-02 | - | 32 | 3 % | 1,776 | 0,856 | 3,81E-02 | - | 29 | - | - | - | - | - | - | - | - | - | - | - | - | - | - | - | - | - | - | - | - | - | - | - | - | - | - | - | - | - | - | - | - | - | - |
| | | | 4 | 171081364 | rs76996950 | 0.18412 (7) | - | - | - | A | G | 3,20 | 1,34 | 2,96 | 4,01 | 3,87 | 3,00 | 2,97 | 1,54 | 3 % | 0,817 | 0,468 | 8,12E-02 | - | 32 | 3 % | 1,004 | 0,611 | 1,00E-01 | - | 28 | 3 % | 1,261 | 0,897 | 1,60E-01 | - | 25 | 2,2 % | -0,071 | 4,559 | 7,45E-01 | ≤3 | 167 | 2,1 % | -0,474 | 3,514 | 9,56E-02 | ≤3 | 156 | 2,2 % | 0,168 | 2,027 | 7,33E-01 | ≤3 | 156 | 2,3 % | 0,035 | 0,042 | 4,06E-01 | 2,3 % | 0,049 | 0,043 | 2,57E-01 | 2,3 % | 0,049 | 0,045 | 2,84E-01 | 2,3 % | 0,018 | 0,083 | 8,33E-01 |
| | | | 4 | 171081380 | rs12643672 | 0.18412 (7) | - | - | - | C | T | 2,91 | 1,34 | 2,96 | 4,05 | 2,69 | 3,00 | 2,97 | 1,62 | 5 % | 1,671 | 0,370 | 6,30E-06 | ≤3 | 52 | 5 % | 2,388 | 0,474 | 4,77E-07 | ≤3 | 46 | 4 % | 0,420 | 0,884 | 6,35E-01 | - | 34 | 5,0 % | -0,165 | 6,767 | 2,64E-01 | 7 | 380 | 5,0 % | -0,072 | 5,283 | 7,04E-01 | 7 | 365 | 5,1 % | -0,006 | 3,001 | 9,85E-01 | 7 | 354 | 4,8 % | 0,025 | 0,030 | 4,06E-01 | 4,8 % | 0,042 | 0,031 | 1,72E-01 | 4,8 % | 0,019 | 0,032 | 5,43E-01 | 4,8 % | 0,022 | 0,059 | 7,10E-01 |
| | | | 4 | 171081723 | rs4555589 | 0.13454 (5) | - | - | - | G | A | 1,91 | 4,89 | 2,95 | 1,49 | 2,74 | 3,00 | 2,97 | 3,48 | 5 % | 1,671 | 0,370 | 6,30E-06 | ≤3 | 52 | 5 % | 2,388 | 0,474 | 4,77E-07 | ≤3 | 46 | 4 % | 0,420 | 0,884 | 6,35E-01 | - | 34 | 5,0 % | -0,165 | 6,767 | 2,64E-01 | 7 | 380 | 5,0 % | -0,072 | 5,283 | 7,04E-01 | 7 | 365 | 5,1 % | -0,006 | 3,001 | 9,85E-01 | 7 | 354 | 4,8 % | 0,025 | 0,030 | 4,07E-01 | 4,8 % | 0,042 | 0,031 | 1,72E-01 | 4,8 % | 0,019 | 0,032 | 5,44E-01 | 4,8 % | 0,022 | 0,059 | 7,10E-01 |
| MAF<5% | *ARHGEF37, LOC105378224* | 5:149692001-149696000 | 5 | 149692421 | rs1351095191 | 0.13454 (5) | - | - | - | C | T | 2,42 | 1,34 | 8,72 | 3,32 | 1,08 | 3,00 | 2,97 | 2,40 | 0 % | 4,499 | 2,407 | 6,16E-02 | - | ≤3 | - | - | - | - | - | - | - | - | - | - | - | - | - | - | - | - | - | - | - | - | - | - | - | - | - | - | - | - | - | - | 0,0 % | -0,604 | 0,376 | 1,09E-01 | 0,0 % | -0,558 | 0,390 | 1,52E-01 | 0,0 % | -0,501 | 0,407 | 2,19E-01 | 0,0 % | -1,054 | 0,739 | 1,54E-01 |
| | | | 5 | 149692610 | rs139875661 | 0.13454 (5) | - | - | - | C | T | 3,36 | 1,34 | 8,90 | 6,16 | 3,30 | 3,00 | 2,97 | 1,16 | 0 % | -1,666 | 2,076 | 4,22E-01 | - | ≤3 | 0 % | -1,442 | 2,245 | 5,21E-01 | - | ≤3 | 0 % | -1,200 | 2,972 | 6,86E-01 | - | ≤3 | - | - | - | - | - | - | - | - | - | - | - | - | - | - | - | - | - | - | - | - | - | - | - | - | - | - | - | - | - | - | - | - | - | - |
| | | | 5 | 149693137 | rs78481200 | 0.13454 (5) | *ARHGEF.* | *PCYOX1L,* | - | C | T | 3,84 | 1,34 | 9,49 | 5,96 | 4,13 | 3,00 | 2,97 | 1,57 | 2 % | -0,368 | 0,564 | 5,14E-01 | - | 22 | 2 % | -0,250 | 0,693 | 7,18E-01 | - | 21 | 2 % | -1,123 | 1,068 | 2,93E-01 | - | 19 | 2,0 % | 0,641 | 4,301 | 5,82E-03 | - | 156 | 1,9 % | 0,390 | 3,260 | 2,04E-01 | - | 141 | 1,9 % | -0,130 | 1,833 | 8,12E-01 | - | 133 | 2,0 % | -0,012 | 0,045 | 7,94E-01 | 2,0 % | -0,003 | 0,047 | 9,50E-01 | 2,0 % | -0,007 | 0,049 | 8,82E-01 | 2,0 % | -0,050 | 0,090 | 5,78E-01 |
| | | | 5 | 149693489 | rs142606152 | 0.13454 (5) | - | - | - | A | T | 5,13 | 6,00 | 9,96 | 7,30 | 6,36 | 3,00 | 2,97 | 1,16 | 0 % | -1,399 | 2,242 | 5,33E-01 | - | ≤3 | 0 % | -1,267 | 2,504 | 6,13E-01 | - | ≤3 | 0 % | -1,088 | 3,945 | 7,83E-01 | - | ≤3 | - | - | - | - | - | - | - | - | - | - | - | - | - | - | - | - | - | - | - | - | - | - | - | - | - | - | - | - | - | - | - | - | - | - |
| | | | 5 | 149693493 | rs146817331 | 0.13454 (5) | - | *SYNPO,SH* | - | A | G | 3,42 | 6,00 | 9,96 | 7,48 | 3,00 | 3,00 | 2,97 | 1,15 | 0 % | 1,578 | 1,698 | 3,53E-01 | - | ≤3 | 0 % | 3,058 | 2,057 | 1,37E-01 | - | ≤3 | 0 % | -1,069 | 4,315 | 8,04E-01 | - | ≤3 | 0,2 % | -0,837 | 1,516 | 2,04E-01 | - | 18 | 0,2 % | -0,692 | 1,207 | 4,04E-01 | - | 18 | 0,2 % | -0,724 | 0,694 | 6,15E-01 | - | 18 | 0,3 % | -0,066 | 0,114 | 5,64E-01 | 0,3 % | -0,048 | 0,117 | 6,80E-01 | 0,3 % | -0,090 | 0,123 | 4,62E-01 | 0,3 % | 0,138 | 0,229 | 5,46E-01 |
| | | | 5 | 149693610 | rs1183208132 | 0.55134 (3a | - | - | - | CA | C | 0,41 | 6,00 | 10,14 | 7,97 | 1,47 | 3,00 | 2,97 | 2,97 | 0 % | -1,182 | 2,020 | 5,58E-01 | - | ≤3 | 0 % | -1,116 | 2,467 | 6,51E-01 | - | ≤3 | 0 % | -1,050 | 3,814 | 7,83E-01 | - | ≤3 | - | - | - | - | - | - | - | - | - | - | - | - | - | - | - | - | - | - | - | - | - | - | - | - | - | - | - | - | - | - | - | - | - | - |
| | | | 5 | 149693837 | rs187241258 | - | - | - | - | T | C | 3,07 | 3,99 | 10,49 | 6,72 | 1,91 | 3,00 | 2,97 | 1,33 | 0 % | 3,202 | 2,176 | 1,41E-01 | - | ≤3 | 0 % | 4,543 | 2,430 | 6,15E-02 | - | ≤3 | - | - | - | - | - | - | - | - | - | - | - | - | - | - | - | - | - | - | - | - | - | - | - | - | - | - | - | - | - | - | - | - | - | - | - | - | - | - | - | - |
| | | | 5 | 149693942 | rs114559837 | 0.60906 (4) | - | - | - | G | A | 0,10 | 6,45 | 10,67 | 7,43 | 0,20 | 3,00 | 2,97 | 0,73 | 0 % | 1,503 | 1,201 | 2,11E-01 | - | 4 | 0 % | 2,898 | 1,438 | 4,38E-02 | - | 4 | 0 % | -1,072 | 3,430 | 7,55E-01 | - | ≤3 | 0,5 % | 0,035 | 2,263 | 9,36E-01 | ≤3 | 34 | 0,5 % | -0,532 | 1,690 | 3,69E-01 | ≤3 | 33 | 0,5 % | 1,079 | 0,991 | 2,85E-01 | ≤3 | 34 | 0,5 % | 0,127 | 0,087 | 1,46E-01 | 0,5 % | 0,132 | 0,090 | 1,43E-01 | 0,5 % | 0,121 | 0,094 | 1,99E-01 | 0,5 % | 0,157 | 0,175 | 3,70E-01 |
| | | | 5 | 149694214 | rs75117515 | 0.68144 (2b | - | - | - | C | T | 21,63 | 12,30 | 11,20 | 7,78 | 24,17 | 3,00 | 2,97 | 1,04 | 1 % | -0,347 | 0,707 | 6,23E-01 | - | 14 | 1 % | -0,458 | 0,878 | 6,02E-01 | - | 13 | 1 % | 0,309 | 1,205 | 7,98E-01 | - | 13 | 1,1 % | -0,494 | 3,289 | 1,04E-01 | - | 89 | 1,2 % | -0,592 | 2,574 | 1,28E-01 | - | 86 | 1,2 % | -0,691 | 1,479 | 3,07E-01 | - | 85 | 1,1 % | -0,012 | 0,061 | 8,44E-01 | 1,1 % | -0,041 | 0,063 | 5,18E-01 | 1,1 % | -0,015 | 0,066 | 8,19E-01 | 1,1 % | 0,091 | 0,121 | 4,53E-01 |
| | | | 5 | 149694483 | rs569293635 | 0.60906 (4) | - | - | - | AG | A | 1,54 | 6,88 | 11,82 | 6,02 | 3,03 | 3,00 | 2,97 | 1,60 | 0 % | 2,056 | 1,845 | 2,65E-01 | - | ≤3 | 0 % | 3,649 | 2,203 | 9,76E-02 | - | ≤3 | 0 % | -1,137 | 3,255 | 7,27E-01 | - | ≤3 | 0,7 % | -0,047 | 2,643 | 9,02E-01 | ≤3 | 53 | 0,7 % | -0,119 | 2,048 | 8,07E-01 | ≤3 | 50 | 0,8 % | 1,576 | 1,187 | 6,13E-02 | ≤3 | 50 | 0,9 % | 0,007 | 0,069 | 9,19E-01 | 0,9 % | -0,022 | 0,071 | 7,58E-01 | 0,9 % | -0,050 | 0,074 | 5,00E-01 | 0,9 % | 0,221 | 0,137 | 1,07E-01 |
| | | | 5 | 149694857 | rs184797567 | 0 (5) | - | - | - | A | T | 6,05 | 8,29 | 13,03 | 3,69 | 5,91 | 3,00 | 2,97 | 0,51 | 1 % | 0,698 | 0,724 | 3,35E-01 | - | 12 | 1 % | 0,480 | 0,931 | 6,06E-01 | - | 10 | 1 % | -1,143 | 1,453 | 4,31E-01 | - | 8 | 0,5 % | -0,130 | 2,044 | 7,90E-01 | - | 34 | 0,5 % | 0,690 | 1,627 | 2,61E-01 | - | 34 | 0,5 % | -0,733 | 0,899 | 5,10E-01 | - | 31 | 0,4 % | -0,134 | 0,096 | 1,62E-01 | 0,4 % | -0,125 | 0,099 | 2,07E-01 | 0,4 % | -0,111 | 0,104 | 2,86E-01 | 0,4 % | -0,110 | 0,191 | 5,66E-01 |
| | | | 5 | 149694893 | rs527817444 | 0.38825 (5) | - | - | - | C | A | 1,69 | 8,88 | 13,19 | 4,40 | 1,83 | 3,00 | 2,97 | 0,43 | 0 % | -1,192 | 2,775 | 6,68E-01 | - | ≤3 | 0 % | -1,112 | 3,442 | 7,47E-01 | - | ≤3 | 0 % | -1,052 | 5,044 | 8,35E-01 | - | ≤3 | - | - | - | - | - | - | - | - | - | - | - | - | - | - | - | - | - | - | - | - | - | - | - | - | - | - | - | - | - | - | - | - | - | - |
| | | | 5 | 149694960 | rs941743265 | 0.13454 (5) | - | - | - | G | T | 11,32 | 8,29 | 13,50 | 6,40 | 11,84 | 3,00 | 2,97 | 0,47 | 0 % | 4,606 | 2,431 | 5,81E-02 | - | ≤3 | 0 % | 7,012 | 2,864 | 1,44E-02 | - | ≤3 | - | - | - | - | - | - | - | - | - | - | - | - | - | - | - | - | - | - | - | - | - | - | - | - | - | - | - | - | - | - | - | - | - | - | - | - | - | - | - | - |
| | | | 5 | 149694979 | rs189261927 | 0.13454 (5) | - | - | - | C | T | 0,39 | 7,64 | 13,60 | 7,53 | 0,40 | 3,00 | 2,97 | 0,49 | 0 % | -1,213 | 1,986 | 5,41E-01 | - | ≤3 | 0 % | -1,109 | 2,560 | 6,65E-01 | - | ≤3 | 0 % | -1,059 | 3,601 | 7,69E-01 | - | ≤3 | 0,2 % | 0,585 | 1,328 | 4,37E-01 | - | 14 | 0,2 % | 0,591 | 1,020 | 5,46E-01 | - | 13 | 0,2 % | 3,982 | 0,589 | 1,89E-02 | - | 13 | 0,2 % | 0,239 | 0,176 | 1,76E-01 | 0,2 % | 0,115 | 0,183 | 5,28E-01 | 0,2 % | -0,020 | 0,192 | 9,19E-01 | 0,2 % | 0,808 | 0,343 | 1,83E-02 |
| | | | 5 | 149694997 | rs71586138 | 0.13454 (5) | - | - | - | C | G | 1,86 | 7,64 | 13,69 | 7,12 | 1,75 | 3,00 | 2,97 | 0,54 | 1 % | -0,199 | 1,030 | 8,47E-01 | - | 7 | 1 % | 0,414 | 1,257 | 7,42E-01 | - | 7 | 1 % | -1,067 | 2,072 | 6,06E-01 | - | 6 | 1,4 % | 0,470 | 3,629 | 8,79E-02 | ≤3 | 104 | 1,4 % | 0,512 | 2,782 | 1,54E-01 | ≤3 | 96 | 1,3 % | 0,571 | 1,566 | 3,71E-01 | ≤3 | 91 | 1,4 % | 0,022 | 0,054 | 6,82E-01 | 1,4 % | 0,024 | 0,056 | 6,61E-01 | 1,4 % | 0,027 | 0,058 | 6,41E-01 | 1,4 % | 0,053 | 0,107 | 6,20E-01 |
| | | | 5 | 149695239 | rs550269426 | 0 (5) | - | - | - | T | G | 17,17 | 6,88 | 15,21 | 4,70 | 21,33 | 3,00 | 2,97 | 0,35 | 0 % | 0,310 | 1,295 | 8,11E-01 | - | 4 | 0 % | -1,119 | 2,088 | 5,92E-01 | - | ≤3 | 0 % | -1,064 | 3,097 | 7,31E-01 | - | ≤3 | 0,4 % | -0,940 | 1,924 | 7,06E-02 | - | 30 | 0,4 % | -0,779 | 1,532 | 2,33E-01 | - | 30 | 0,4 % | -0,681 | 0,885 | 5,47E-01 | - | 30 | 0,3 % | -0,067 | 0,114 | 5,57E-01 | 0,3 % | -0,018 | 0,118 | 8,77E-01 | 0,3 % | -0,005 | 0,123 | 9,70E-01 | 0,3 % | -0,349 | 0,225 | 1,21E-01 |
| | | | 5 | 149695451 | rs536041352 | 0.00167 (6) | - | - | - | G | A | 0,96 | 3,99 | 17,40 | 2,31 | 1,37 | 3,00 | 2,97 | 0,69 | 0 % | -1,121 | 3,290 | 7,33E-01 | - | ≤3 | 0 % | -1,066 | 4,323 | 8,05E-01 | - | ≤3 | 0 % | -1,030 | 6,580 | 8,76E-01 | - | ≤3 | - | - | - | - | - | - | - | - | - | - | - | - | - | - | - | - | - | - | 0,1 % | 0,115 | 0,223 | 6,05E-01 | 0,1 % | 0,160 | 0,230 | 4,86E-01 | 0,1 % | 0,034 | 0,242 | 8,88E-01 | 0,1 % | 0,349 | 0,451 | 4,40E-01 |
| | | | 5 | 149695510 | rs767685573 | 0.31452 (6) | - | - | - | A | G | 0,25 | 1,34 | 18,31 | 3,61 | 0,18 | 3,00 | 2,97 | 0,56 | 0 % | -1,266 | 1,829 | 4,89E-01 | - | ≤3 | 0 % | -1,151 | 2,195 | 6,00E-01 | - | ≤3 | 0 % | -1,070 | 3,243 | 7,41E-01 | - | ≤3 | 0,1 % | 1,523 | 0,847 | 1,97E-01 | - | 7 | 0,1 % | -0,727 | 0,600 | 6,63E-01 | - | 6 | 0,1 % | 5,056 | 0,354 | 7,35E-02 | - | 6 | 0,1 % | 0,099 | 0,231 | 6,67E-01 | 0,1 % | 0,139 | 0,239 | 5,60E-01 | 0,1 % | 0,106 | 0,250 | 6,72E-01 | 0,1 % | -0,050 | 0,471 | 9,15E-01 |
| | | | 5 | 149695837 | rs150739746 | 0.13454 (5) | - | - | - | A | G | 0,86 | 3,99 | 25,05 | 9,78 | 0,28 | 3,00 | 2,97 | 0,42 | 0 % | -1,136 | 3,153 | 7,19E-01 | - | ≤3 | 0 % | -1,069 | 4,258 | 8,02E-01 | - | ≤3 | 0 % | -1,039 | 5,788 | 8,58E-01 | - | ≤3 | 0,1 % | 0,218 | 0,944 | 8,37E-01 | - | 7 | 0,1 % | 1,440 | 0,751 | 2,79E-01 | - | 7 | 0,1 % | -0,830 | 0,402 | 7,39E-01 | - | 6 | 0,1 % | 0,074 | 0,235 | 7,52E-01 | 0,1 % | 0,179 | 0,243 | 4,62E-01 | 0,1 % | 0,209 | 0,254 | 4,12E-01 | 0,1 % | -0,021 | 0,467 | 9,63E-01 |
| | | | 5 | 149695899 | rs79355049 | 0.13454 (5) | *CSNK1A1* | *CTB-13H5* | - | A | C | 8,01 | 3,99 | 25,64 | 9,56 | 7,85 | 3,00 | 2,97 | 0,71 | 2 % | 2,966 | 0,544 | 4,94E-08 | ≤3 | 20 | 2 % | 4,381 | 0,733 | 2,23E-09 | ≤3 | 16 | 1 % | 3,778 | 1,560 | 1,55E-02 | - | 10 | 3,2 % | 0,152 | 5,372 | 4,15E-01 | ≤3 | 239 | 3,1 % | -0,206 | 4,120 | 3,95E-01 | ≤3 | 221 | 3,1 % | 0,301 | 2,359 | 4,78E-01 | ≤3 | 217 | 3,0 % | -0,028 | 0,037 | 4,52E-01 | 3,0 % | -0,029 | 0,039 | 4,46E-01 | 3,0 % | -0,016 | 0,040 | 6,84E-01 | 3,0 % | -0,033 | 0,075 | 6,64E-01 |

**MUTAT=Mutation density aPC**
**microRNA=microRNA aPC**
**PROT=Protein function aPC**
**CONS=Conservation aPC**
**EPIGEN=Epigenetics aPC**
**TSS/TES=Distance to TSS or TES aPC**
**TF=Transcription factor aPC**
**CADD=Combined annotation dependent depletion**

**?=Not found in the data base (GTEx and YUE lab)**

**RegulomeDB** = probability score (ranking)

| | |
|---|---|
| 1a | eQTL + TF binding + matched TF motif + matched DNase Footprint + DNase peak |
| 1b | eQTL + TF binding + any motif + DNase Footprint + DNase peak |
| 1c | eQTL + TF binding + matched TF motif + DNase peak |
| 1d | eQTL + TF binding + any motif + DNase peak |
| 1e | eQTL + TF binding + matched TF motif |
| 1f | eQTL + TF binding / DNase peak |
| 2a | TF binding + matched TF motif + matched DNase Footprint + DNase peak |
| 2b | TF binding + any motif + DNase Footprint + DNase peak |
| 2c | TF binding + matched TF motif + DNase peak |
| 3a | TF binding + any motif + DNase peak |
| 3b | TF binding + matched TF motif |
| 4 | TF binding + DNase peak |
| 5 | TF binding or DNase peak |
| 6 | Motif hit |
| 7 | Other |

**Supplementary Table 20:** Suggestively significant enhancers (STAAR-O) with replication in T1D

| | Enhancer | Gene | Discovery Stroke (WGS: T1D specific) | | | Discovery Ischemic stroke (WGS: T1D specific) | | | Discovery Hemorrhagic stroke (WGS: T1D specific) | | | Replication GWAS Stroke (T1D specific) | | | Replication GWAS Ischemic stroke (T1D specific) | | | Replication GWAS Hemorrhagic stroke (T1D specific) | | |
|---|---|---|---|---|---|---|---|---|---|---|---|---|---|---|---|---|---|---|---|---|
| | | | N | CMAC | *p* -value | N | CMAC | *p* -value | N | CMAC | *p* -value | N | CMAC | *p* -value | N | CMAC | *p* -value | N | CMAC | *p* -value |
| **MAF<5%** | 9: 71179769-71180074 | *TRPM3* | 3 | 5 | 1,34E-05 | - | - | - | - | - | - | - | - | - | - | - | - | - | - | - |
| | 11: 27694286-27694608 | *BDNF, BDNF-AS* | 2 | 8 | 6,21E-06 | 2 | 6 | 1,01E-06 | - | - | - | - | - | - | - | - | - | - | - | - |
| **MAF<1%** | 9: 71179769-71180074 | *TRPM3* | 3 | 5 | 1,34E-05 | - | - | - | - | - | - | - | - | - | - | - | - | - | - | - |
| | 11: 27694286-27694608 | *BDNF, BDNF-AS* | 2 | 8 | 6,21E-06 | 2 | 6 | 1,01E-06 | - | - | - | - | - | - | - | - | - | - | - | - |
| | 5:58190438-58190534 | *LOC105378983* | 2 | 5 | 2,69E-05 | - | - | - | - | - | - | - | - | - | - | - | - | - | - | - |

MAF<5% (significance $p < 2.1\times10^{-6}$, suggestive $p < 2.1\times10^{-5}$), MAF<1% (significance $p < 2.7\times10^{-6}$, suggestive $p < 2.7\times10^{-5}$)

**Supplementary Table 21:** Variants in the discovered enhancers (STAAR-O) with replication in T1D (FinnDiane) and general population (FinnGen)

*Note: Stroke subtype analyses may have included additional variants (i.e. passed MAF threshold in smaller data)*

| MODEL | CLOSEST GENE | CHR | POS | rsnumber | ALT | REF | RegulomeD | ChIP-seq/PWI | GTEx ( | eQTLGen ( | PCHi-C ( | PHRED SCALE SCORE MUTAT | microRN | PROT | CONS | EPIGEN | TSS/TES | TF | CADD |
|---|---|---|---|---|---|---|---|---|---|---|---|---|---|---|---|---|---|---|---|
| MAF 1% & 5% | *TRPM3* | 9 | 71180071 | rs2077314064 | T | TGAGA | - | - | - | - | - | 6,82 | 2,97 | 3,00 | 6,87 | 0,71 | 0,20 | 1,34 | 1,24 |
| | | 9 | 71180003 | rs989199629 | C | G | 0.1841 (7) | -/- | - | - | - | 5,59 | 2,97 | 3,00 | 2,65 | 1,71 | 0,20 | 1,34 | 2,61 |
| | | 9 | 71180047 | rs1587812218 | CTTTTT | C | - | - | - | - | - | 7,25 | 2,97 | 3,00 | 6,12 | 1,05 | 0,20 | 1,34 | 1,31 |
| MAF 1% & | *BDNF, BDNF-AS* | 11 | 27694337 | rs1010405514 | C | T | 0.1841 (7) | -/- | - | - | *BDNF* ir | 12,43 | 2,97 | 3,00 | 6,59 | 6,08 | 7,82 | 1,34 | 7,21 |
| | | 11 | 27694503 | rs72881243 | A | G | 0.1841 (7) | -/- | - | *LGR4, KIF1* | *BDNF* ir | 11,69 | 2,97 | 3,00 | 0,08 | 12,50 | 7,94 | 1,34 | 0,02 |
| MAF 1% | *LOC105378983* | 5 | 58190493 | rs912651902 | A | G | 0.2440 (3a) | *MAFK/RBPJ* | - | - | *GAPT* ir | 2,66 | 2,97 | 3,00 | 19,85 | 9,13 | 2,79 | 7,29 | 0,03 |
| | | 5 | 58190534 | rs558946071 | T | C | 0.6091 (4) | *MAFK/-* | - | - | *GAPT* ir | 2,35 | 2,97 | 3,00 | 2,99 | 8,56 | 2,79 | 8,01 | 3,87 |

**Discovery cohort FinnDiane WGS using score test (T1D specific)**

| rsnumber | Stroke MAF | Effect | SE | p-value | HOM | HET | Ischemic stroke MAF | Effect | SE | p-value | HOM | HET | Hemorrhagic stroke MAF | Effect | SE | p-value | HOM | HET |
|---|---|---|---|---|---|---|---|---|---|---|---|---|---|---|---|---|---|---|
| rs2077314064 | 0 % | 8,445 | 1,791 | 2,40E-06 | - | ≤3 | 0 % | 20,193 | 4,610 | 1,19E-05 | - | ≤3 | 0 % | 41,854 | 6,549 | 1,65E-10 | - | ≤3 |
| rs989199629 | 0 % | -1,175 | 2,859 | 6,81E-01 | - | ≤3 | 0 % | -1,117 | 3,364 | 7,40E-01 | - | ≤3 | 0 % | -1,039 | 5,714 | 8,56E-01 | - | ≤3 |
| rs1587812218 | 0 % | -1,122 | 3,274 | 7,32E-01 | - | ≤3 | 0 % | -1,070 | 4,178 | 7,98E-01 | - | ≤3 | 0 % | -1,029 | 6,585 | 8,76E-01 | - | ≤3 |
| rs1010405514 | 0 % | 5,560 | 1,853 | 2,69E-03 | - | ≤3 | 0 % | 16,782 | 4,225 | 7,12E-05 | - | ≤3 | - | - | - | - | - | - |
| rs72881243 | 1 % | 4,006 | 1,144 | 4,63E-04 | - | 6 | 0 % | 5,457 | 1,484 | 2,36E-04 | - | 5 | 0 % | 12,545 | 3,686 | 6,65E-04 | - | ≤3 |
| rs912651902 | 0 % | 4,623 | 2,433 | 5,74E-02 | - | ≤3 | 0 % | 7,922 | 3,014 | 8,57E-03 | - | ≤3 | - | - | - | - | - | - |
| rs558946071 | 0 % | 6,129 | 1,559 | 8,46E-05 | - | 4 | 0 % | 7,307 | 2,896 | 1,16E-02 | - | ≤3 | 0 % | 13,164 | 3,771 | 4,82E-04 | - | ≤3 |

**FinnDiane GWAS replication using score test (T1D specific)**

| rsnumber | Stroke MAF | Effect | SE | p-value | HOM | HET | Ischemic stroke MAF | Effect | SE | p-value | HOM | HET | Hemorrhagic stroke MAF | Effect | SE | p-value | HOM | HET |
|---|---|---|---|---|---|---|---|---|---|---|---|---|---|---|---|---|---|---|
| rs2077314064 | - | - | - | - | - | - | - | - | - | - | - | - | - | - | - | - | - | - |
| rs989199629 | - | - | - | - | - | - | - | - | - | - | - | - | - | - | - | - | - | - |
| rs1587812218 | - | - | - | - | - | - | - | - | - | - | - | - | - | - | - | - | - | - |
| rs1010405514 | - | - | - | - | - | - | - | - | - | - | - | - | - | - | - | - | - | - |
| rs72881243 | 0,8 % | -0,092 | 0,358 | 7,97E-01 | - | 64 | 0,9 % | 0,132 | 0,457 | 7,73E-01 | - | 1488 | 0,8 % | -0,598 | 0,810 | 4,60E-01 | - | 59 |
| rs912651902 | - | - | - | - | - | - | - | - | - | - | - | - | - | - | - | - | - | - |
| rs558946071 | 0,7 % | 0,547 | 0,371 | 1,40E-01 | - | 60 | 0,7 % | -0,153 | 0,496 | 7,58E-01 | - | 53 | 0,7 % | 0,488 | 0,866 | 5,73E-01 | - | 52 |

**FinnGen GWAS replication (general population)**

| rsnumber | Stroke MAF | Effect | SE | p-value | Ischemic stroke MAF | Effect | SE | p-value | Hemorrhagic stroke MAF | Effect | SE | p-value | Stroke, excluding SAH MAF | Effect | SE | p-value |
|---|---|---|---|---|---|---|---|---|---|---|---|---|---|---|---|---|
| rs2077314064 | - | - | - | - | - | - | - | - | - | - | - | - | - | - | - | - |
| rs989199629 | - | - | - | - | - | - | - | - | - | - | - | - | - | - | - | - |
| rs1587812218 | - | - | - | - | - | - | - | - | - | - | - | - | - | - | - | - |
| rs1010405514 | - | - | - | - | - | - | - | - | - | - | - | - | - | - | - | - |
| rs72881243 | 0,8 % | 0,030 | 0,070 | 6,73E-01 | 0,8 % | 0,070 | 0,076 | 3,54E-01 | 0,8 % | -0,087 | 0,140 | 5,36E-01 | 0,8 % | 0,075 | 0,072 | 2,98E-01 |
| rs912651902 | - | - | - | - | - | - | - | - | - | - | - | - | - | - | - | - |
| rs558946071 | 0,7 % | -0,010 | 0,077 | 8,96E-01 | 0,7 % | -0,024 | 0,084 | 7,70E-01 | 0,7 % | -0,020 | 0,152 | 8,95E-01 | 0,7 % | -0,035 | 0,080 | 6,60E-01 |

**MUTAT**=Mutation density aPC
**microRNA**=microRNA aPC
**PROT**=Protein function aPC
**CONS**=Conservation aPC
**EPIGEN**=Epigenetics aPC
**TSS/TES**=Distance to TSS or TES aPC
**TF**=Transcription factor aPC
**CADD**=Combined annotation dependent depletion

?=Not found in the data base (GTEx and YUE lab)

**RegulomeDB** = probability score (ranking)

| | |
|---|---|
| 1a | eQTL + TF binding + matched TF motif + matched DNase Footprint + DNase peak |
| 1b | eQTL + TF binding + any motif + DNase Footprint + DNase peak |
| 1c | eQTL + TF binding + matched TF motif + DNase peak |
| 1d | eQTL + TF binding + any motif + DNase peak |
| 1e | eQTL + TF binding + matched TF motif |
| 1f | eQTL + TF binding / DNase peak |
| 2a | TF binding + matched TF motif + matched DNase Footprint + DNase peak |
| 2b | TF binding + any motif + DNase Footprint + DNase peak |
| 2c | TF binding + matched TF motif + DNase peak |
| 3a | TF binding + any motif + DNase peak |
| 3b | TF binding + matched TF motif |
| 4 | TF binding + DNase peak |
| 5 | TF binding or DNase peak |
| 6 | Motif hit |
| 7 | Other |

**ChIP-seq (RegulomeDB)**: Highest probability transcription factor

**Supplementary Table 22:** Suggestively significant ($p<6\times10^{-6}$) promoters (STAAR-O) with replication in T1D

| | CAGE peak | Promoter region | Gene* | Discovery Stroke (WGS: T1D specific) | | | Discovery Ischemic stroke (WGS: T1D specific) | | | Discovery Hemorrhagic stroke (WGS: T1D specific) | | | Replication GWAS Stroke (T1D specific) | | | Replication GWAS Ischemic stroke (T1D specific) | | | Replication GWAS Hemorrhagic stroke (T1D specific) | | |
|---|---|---|---|---|---|---|---|---|---|---|---|---|---|---|---|---|---|---|---|---|---|
| | | | | N | CMAC | *p*-value | N | CMAC | *p*-value | N | CMAC | *p*-value | N | CMAC | *p*-value | N | CMAC | *p*-value | N | CMAC | *p*-value |
| **MAF<1%** | chr2:85327938-85327958(-) | 2:85327939-85328939 | *TGOLN2* | 4 | 9 | 5,60E-06 | 4 | 5 | 4,71E-07 | - | - | - | 4 | 74 | 0,439 | 4 | 72 | 0,552 | 4 | 72 | 0,563 |
| | chr2:85327963-85327996(-) | 2:85327964-85328964 | *TGOLN2* | 4 | 9 | 5,60E-06 | 4 | 5 | 4,71E-07 | - | - | - | 4 | 74 | 0,439 | 4 | 72 | 0,552 | 4 | 72 | 0,563 |
| | chr21:44425577-44425612(-) | 21:44425578-44426578 | *TRPM2-AS* | 13 | 33 | 5,78E-06 | 13 | 28 | 9,67E-05 | 13 | 22 | 0,0573 | 8 | 105 | 0,053 | 8 | 102 | 0,176 | 8 | 103 | 0,656 |

*Gene = gene upstream/downstream the transcription start site (TSS)

**Supplementary Table 23:** Variants in the discovered promoters (STAAR-O) with replication in T1D (FinnDiane) and general population (FinnGen)

*Note: Stroke subtype analyses may have included additional variants (i.e. passed MAF threshold in smaller data)*

| MODEL | GENE | CHR | POS | rsnumber | RegulomeDB | ChIP-seq/PWM | GTEx (multi | eQTLGen | REF | ALT | PHRED SCALE SCORE: CADD | TF | TSS/TES | EPIGEN | CONS | PROT | microRN/ | MUTAT |
|---|---|---|---|---|---|---|---|---|---|---|---|---|---|---|---|---|---|---|
| MAF 1% | *TGOLN2* | 2 | 85327996 | rs371742011 | 0.7050 (4) | *POLR2A/-* | - | - | C | T | 17,56 | 35,79 | 21,30 | 33,01 | 12,65 | 3,00 | 2,97 | 0,67 |
| | | 2 | 85328016 | rs376006909 | 0.7050 (4) | *POLR2A/-* | - | - | G | T | 0,79 | 36,22 | 21,87 | 32,47 | 0,07 | 3,00 | 2,97 | 0,67 |
| | | 2 | 85328462 | rs1182214256 | 0.4945 (3a) | *POLR2A/RFX5,R* | - | - | C | T | 10,12 | 25,20 | 17,57 | 31,06 | 6,25 | 3,00 | 2,97 | 1,35 |
| | | 2 | 85328585 | rs542060093 | 1.0000 (2c) | *POLR2A/GABPA,* | - | - | C | T | 8,59 | 17,88 | 13,93 | 21,88 | 4,55 | 3,00 | 2,97 | 2,64 |
| MAF 1% & 5% | *TRPM2-AS* | 21 | 44425645 | rs148758503 | 0.7650 (2b) | *POLR2A/ATF4,C* | - | - | G | A | 0,66 | 20,72 | 13,15 | 21,90 | 0,23 | 3,00 | 2,97 | 0,42 |
| | | 21 | 44425661 | rs753589764 | 0.6091 (4) | *POLR2A/-* | - | - | T | C | 5,49 | 20,90 | 12,80 | 22,03 | 1,34 | 3,00 | 2,97 | 0,47 |
| | | 21 | 44425712 | rs547070705 | 0.6091 (4) | *POLR2A/-* | - | - | GCAGG | G | 19,09 | 20,62 | 12,03 | 21,81 | 25,48 | 3,00 | 2,97 | 0,43 |
| | | 21 | 44425768 | rs201007554 | 0.6091 (4) | *POLR2A/-* | - | - | C | A | 17,01 | 19,97 | 11,48 | 21,44 | 12,89 | 3,00 | 2,97 | 0,50 |
| | | 21 | 44425924 | rs762428 | 0.7266 (2b) | *POLR2A/CTF1,N* | *TRPM2* in w | *TRPM2,C* | T | C | - | - | - | - | - | - | - | - |
| | | 21 | 44426004 | rs554907943 | 0.6377 (2b) | *POLR2A/ESR1,N* | - | - | C | T | 6,34 | 12,52 | 10,17 | 20,89 | 5,04 | 3,00 | 2,97 | 1,29 |
| | | 21 | 44426126 | rs1279586198 | 0.7050 (4) | *GABPA/-* | - | - | A | C | 0,77 | 9,33 | 9,74 | 17,60 | 0,28 | 3,00 | 2,97 | 0,47 |
| | | 21 | 44426139 | rs369327546 | 0.9992 (3a) | *GABPA/RXRA* | - | - | A | G | 1,70 | 9,33 | 9,70 | 17,77 | 0,52 | 3,00 | 2,97 | 0,60 |
| | | 21 | 44426167 | rs567417562 | 0.5849 (2b) | *GABPA/PPARA,F* | - | - | C | T | 0,81 | 9,95 | 9,62 | 17,14 | 0,32 | 3,00 | 2,97 | 0,66 |
| | | 21 | 44426322 | rs182018557 | 0.4125 (5) | *-/ETV6* | - | - | G | A | 0,50 | 8,01 | 8,75 | 17,60 | 0,67 | 3,00 | 2,97 | 1,60 |
| | | 21 | 44426323 | rs750539366 | 0.0000 (5) | *-/ETV6* | - | - | C | T | 1,03 | 8,01 | 8,75 | 17,65 | 0,77 | 3,00 | 2,97 | 1,60 |
| | | 21 | 44426417 | rs532316203 | 0.0000 (5) | *-/PLAG1* | - | - | C | T | 4,97 | 6,45 | 8,57 | 16,95 | 5,66 | 3,00 | 2,97 | 1,41 |
| | | 21 | 44426511 | rs543984554 | 0.3034 (5) | *-/SPDEF* | - | - | C | T | 1,92 | 8,01 | 8,41 | 16,43 | 0,41 | 3,00 | 2,97 | 1,28 |

**Discovery cohort FinnDiane WGS using score test (T1D specific)**

| rsnumber | Stroke: MAF | Effect | SE | p-value | HOM | HET | Ischemic stroke: MAF | Effect | SE | p-value | HOM | HET | Hemorrhagic stroke: MAF | Effect | SE | p-value | HOM | HET |
|---|---|---|---|---|---|---|---|---|---|---|---|---|---|---|---|---|---|---|
| rs371742011 | 0 % | 5,039 | 1,800 | 5,12E-03 | - | ≤3 | 0 % | 17,918 | 4,357 | 3,92E-05 | - | ≤3 | 0 % | 9,918 | 3,325 | 2,86E-03 | - | ≤3 |
| rs376006909 | 0 % | 3,425 | 1,533 | 2,54E-02 | - | ≤3 | 0 % | 3,336 | 2,121 | 1,16E-01 | - | ≤3 | 0 % | -1,089 | 3,799 | 7,74E-01 | - | ≤3 |
| rs1182214256 | 0 % | 8,356 | 2,181 | 1,27E-04 | - | ≤3 | 0 % | 16,048 | 4,138 | 1,05E-04 | - | ≤3 | - | - | - | - | - | - |
| rs542060093 | 0 % | 5,039 | 1,800 | 5,12E-03 | - | ≤3 | 0 % | 17,918 | 4,357 | 3,92E-05 | - | ≤3 | 0 % | 9,918 | 3,325 | 2,86E-03 | - | ≤3 |
| rs148758503 | 0 % | -1,238 | 1,360 | 3,63E-01 | - | 4 | 0 % | -1,135 | 1,698 | 5,04E-01 | - | 4 | 0 % | -1,071 | 2,379 | 6,53E-01 | - | 4 |
| rs753589764 | 0 % | 6,188 | 1,365 | 5,80E-06 | - | 4 | 0 % | 20,195 | 4,610 | 1,19E-05 | - | ≤3 | 0 % | 35,718 | 6,062 | 3,82E-09 | - | ≤3 |
| rs547070705 | 0 % | -1,141 | 3,100 | 7,13E-01 | - | ≤3 | 0 % | -1,074 | 4,091 | 7,93E-01 | - | ≤3 | 0 % | -1,040 | 5,652 | 8,54E-01 | - | ≤3 |
| rs201007554 | 0 % | -1,140 | 3,182 | 7,20E-01 | - | ≤3 | 0 % | -1,087 | 3,954 | 7,83E-01 | - | ≤3 | 0 % | -1,044 | 5,850 | 8,58E-01 | - | ≤3 |
| rs762428 | 0 % | 0,375 | 1,288 | 7,71E-01 | - | 4 | 0 % | -1,132 | 1,947 | 5,61E-01 | - | ≤3 | 0 % | -1,068 | 2,719 | 6,94E-01 | - | ≤3 |
| rs554907943 | 0 % | -1,136 | 3,163 | 7,20E-01 | - | ≤3 | 0 % | -1,092 | 3,751 | 7,71E-01 | - | ≤3 | 0 % | -1,029 | 6,759 | 8,79E-01 | - | ≤3 |
| rs1279586198 | 0 % | 3,071 | 1,472 | 3,69E-02 | - | ≤3 | 0 % | 5,435 | 1,813 | 2,73E-03 | - | ≤3 | 0 % | -1,048 | 5,182 | 8,40E-01 | - | ≤3 |
| rs369327546 | 0 % | 5,013 | 1,250 | 6,05E-05 | - | 5 | 0 % | 7,499 | 1,697 | 9,87E-06 | - | 4 | 0 % | 21,488 | 4,746 | 5,96E-06 | - | ≤3 |
| rs567417562 | 0 % | 0,437 | 1,318 | 7,40E-01 | - | 4 | 0 % | 1,156 | 1,535 | 4,51E-01 | - | 4 | 0 % | -1,071 | 2,888 | 7,11E-01 | - | ≤3 |
| rs182018557 | 0 % | 1,173 | 1,566 | 4,54E-01 | - | ≤3 | 0 % | 2,108 | 1,816 | 2,46E-01 | - | ≤3 | 0 % | -1,038 | 4,448 | 8,15E-01 | - | ≤3 |
| rs750539366 | 0 % | -1,164 | 2,963 | 6,94E-01 | - | ≤3 | 0 % | -1,126 | 3,339 | 7,36E-01 | - | ≤3 | 0 % | -1,038 | 6,119 | 8,65E-01 | - | ≤3 |
| rs532316203 | 0 % | -1,279 | 2,447 | 6,01E-01 | - | ≤3 | 0 % | -1,153 | 3,009 | 7,02E-01 | - | ≤3 | 0 % | -1,071 | 4,226 | 8,00E-01 | - | ≤3 |
| rs543984554 | 0 % | -1,407 | 2,266 | 5,35E-01 | - | ≤3 | 0 % | -1,228 | 2,715 | 6,51E-01 | - | ≤3 | 0 % | -1,155 | 3,326 | 7,28E-01 | - | ≤3 |

**FinnDiane GWAS replication using score test (T1D specific)**

| rsnumber | Stroke: MAF | Effect | SE | p-value | HOM | HET | Ischemic stroke: MAF | Effect | SE | p-value | HOM | HET | Hemorrhagic stroke: MAF | Effect | SE | p-value | HOM | HET |
|---|---|---|---|---|---|---|---|---|---|---|---|---|---|---|---|---|---|---|
| rs371742011 | 0,1 % | -1,112 | 0,968 | 2,81E-01 | - | 8 | 0,1 % | -0,977 | 0,771 | 4,51E-01 | - | 8 | 0,1 % | -0,772 | 0,446 | 7,31E-01 | - | 8 |
| rs376006909 | 0,2 % | -1,149 | 1,315 | 1,31E-01 | - | 14 | 0,2 % | -0,987 | 1,046 | 3,02E-01 | - | 14 | 0,2 % | -0,871 | 0,605 | 5,98E-01 | - | 14 |
| rs1182214256 | 0,5 % | -0,152 | 2,165 | 7,43E-01 | - | 42 | 0,5 % | -0,376 | 1,678 | 5,29E-01 | - | 40 | 0,5 % | 1,163 | 0,977 | 2,56E-01 | - | 41 |
| rs542060093 | 0,1 % | -1,112 | 0,968 | 2,81E-01 | - | 8 | 0,1 % | -0,977 | 0,771 | 4,51E-01 | - | 8 | 0,1 % | -0,772 | 0,446 | 7,31E-01 | - | 8 |
| rs148758503 | 0,3 % | -0,512 | 1,525 | 4,35E-01 | - | 19 | 0,3 % | -0,706 | 1,182 | 4,04E-01 | - | 18 | 0,3 % | 2,306 | 0,701 | 1,06E-01 | - | 19 |
| rs753589764 | 0,2 % | -1,009 | 1,184 | 2,32E-01 | - | 11 | 0,2 % | -0,879 | 0,943 | 4,07E-01 | - | 11 | 0,2 % | -0,737 | 0,545 | 6,88E-01 | - | 11 |
| rs547070705 | 0,2 % | -1,201 | 1,203 | 1,49E-01 | - | 11 | 0,2 % | -1,037 | 0,958 | 3,21E-01 | - | 11 | 0,2 % | -0,869 | 0,554 | 6,30E-01 | - | 11 |
| rs201007554 | - | - | - | - | - | - | - | - | - | - | - | - | - | - | - | - | - | - |
| rs762428 | 0,3 % | -0,051 | 3,031 | 8,78E-01 | - | 25 | 0,3 % | 2,589 | #N/A | 1 | - | 23 | 0,4 % | 1,488 | 0,869 | 1,96E-01 | - | 24 |
| rs554907943 | 0,0 % | -0,731 | 0,474 | 7,29E-01 | - | ≤3 | 0,0 % | 0,309 | 0,378 | 9,07E-01 | - | ≤3 | 0,0 % | -1,335 | 0,216 | 7,73E-01 | - | ≤3 |
| rs1279586198 | - | - | - | - | - | - | - | - | - | - | - | - | - | - | - | - | - | - |
| rs369327546 | - | - | - | - | - | - | - | - | - | - | - | - | - | - | - | - | - | - |
| rs567417562 | 0,1 % | -0,872 | 1,114 | 3,31E-01 | - | 10 | 0,1 % | -0,768 | 0,887 | 4,96E-01 | - | 10 | 0,1 % | -0,722 | 0,513 | 7,11E-01 | - | 10 |
| rs182018557 | - | - | - | - | - | - | - | - | - | - | - | - | - | - | - | - | - | - |
| rs750539366 | - | - | - | - | - | - | - | - | - | - | - | - | - | - | - | - | - | - |
| rs532316203 | 0,1 % | -0,646 | 1,112 | 4,73E-01 | - | 10 | 0,1 % | -0,421 | 0,885 | 7,10E-01 | - | 10 | 0,1 % | -0,573 | 0,512 | 7,70E-01 | - | 10 |
| rs543984554 | 0,2 % | -0,127 | 1,340 | 8,64E-01 | - | 14 | 0,2 % | 0,139 | 1,058 | 8,83E-01 | - | 14 | 0,2 % | 1,056 | 0,594 | 5,31E-01 | - | 13 |

**FinnGen GWAS replication (general population)**

| rsnumber | Stroke: MAF | Effect | SE | p-value | Ischemic stroke: MAF | Effect | SE | p-value | Hemorrhagic stroke: MAF | Effect | SE | p-value | Stroke, excluding SAH: MAF | Effect | SE | p-value |
|---|---|---|---|---|---|---|---|---|---|---|---|---|---|---|---|---|
| rs371742011 | 0,2 % | -0,102 | 0,148 | 4,90E-01 | 0,2 % | -0,057 | 0,153 | 7,09E-01 | 0,2 % | -0,086 | 0,161 | 5,91E-01 | 0,2 % | -0,377 | 0,290 | 1,94E-01 |
| rs376006909 | 0,1 % | 0,022 | 0,164 | 8,92E-01 | 0,1 % | 0,057 | 0,169 | 7,38E-01 | 0,1 % | 0,041 | 0,177 | 8,16E-01 | 0,1 % | 0,323 | 0,343 | 3,46E-01 |
| rs1182214256 | 0,6 % | -0,115 | 0,088 | 1,90E-01 | 0,6 % | -0,074 | 0,091 | 4,14E-01 | 0,6 % | -0,044 | 0,095 | 6,46E-01 | 0,6 % | -0,178 | 0,174 | 3,06E-01 |
| rs542060093 | 0,2 % | -0,102 | 0,148 | 4,90E-01 | 0,2 % | -0,057 | 0,153 | 7,10E-01 | 0,2 % | -0,086 | 0,161 | 5,91E-01 | 0,2 % | -0,377 | 0,290 | 1,94E-01 |
| rs148758503 | 0,2 % | -0,157 | 0,147 | 2,84E-01 | 0,2 % | -0,151 | 0,151 | 3,19E-01 | 0,2 % | -0,209 | 0,158 | 1,85E-01 | 0,2 % | -0,038 | 0,292 | 8,96E-01 |
| rs753589764 | 0,1 % | -0,387 | 0,228 | 8,98E-02 | 0,1 % | -0,367 | 0,236 | 1,20E-01 | 0,1 % | -0,330 | 0,248 | 1,82E-01 | 0,1 % | -0,402 | 0,440 | 3,61E-01 |
| rs547070705 | 0,1 % | 0,060 | 0,195 | 7,56E-01 | 0,1 % | 0,087 | 0,202 | 6,65E-01 | 0,1 % | 0,083 | 0,212 | 6,95E-01 | 0,1 % | -0,181 | 0,397 | 6,48E-01 |
| rs201007554 | - | - | - | - | - | - | - | - | - | - | - | - | - | - | - | - |
| rs762428 | 0,4 % | -0,006 | 0,098 | 9,51E-01 | 0,4 % | 0,006 | 0,101 | 9,56E-01 | 0,4 % | 0,047 | 0,106 | 6,57E-01 | 0,4 % | -0,153 | 0,195 | 4,33E-01 |
| rs554907943 | 0,1 % | 0,349 | 0,220 | 1,12E-01 | 0,1 % | 0,394 | 0,226 | 8,10E-02 | 0,1 % | 0,486 | 0,235 | 3,82E-02 | 0,1 % | -0,190 | 0,469 | 6,86E-01 |
| rs1279586198 | - | - | - | - | - | - | - | - | - | - | - | - | - | - | - | - |
| rs369327546 | - | - | - | - | - | - | - | - | - | - | - | - | - | - | - | - |
| rs567417562 | 0,2 % | -0,064 | 0,137 | 6,41E-01 | 0,2 % | -0,019 | 0,141 | 8,91E-01 | 0,2 % | -0,065 | 0,148 | 6,59E-01 | 0,2 % | -0,087 | 0,278 | 7,56E-01 |
| rs182018557 | 0,0 % | -0,745 | 0,476 | 1,17E-01 | 0,0 % | -0,689 | 0,494 | 1,63E-01 | 0,0 % | -0,596 | 0,516 | 2,49E-01 | 0,0 % | -1,274 | 0,945 | 1,78E-01 |
| rs750539366 | - | - | - | - | - | - | - | - | - | - | - | - | - | - | - | - |
| rs532316203 | 0,1 % | -0,035 | 0,228 | 8,77E-01 | 0,1 % | -0,062 | 0,235 | 7,92E-01 | 0,1 % | -0,011 | 0,245 | 9,64E-01 | 0,1 % | -0,350 | 0,483 | 4,68E-01 |
| rs543984554 | 0,2 % | 0,002 | 0,155 | 9,89E-01 | 0,2 % | -0,031 | 0,160 | 8,48E-01 | 0,2 % | -0,043 | 0,167 | 7,96E-01 | 0,2 % | 0,323 | 0,312 | 3,00E-01 |

**MUTAT**=Mutation density aPC
**microRNA**=microRNA aPC
**PROT**=Protein function aPC
**CONS**=Conservation aPC
**EPIGEN**=Epigenetics aPC
**TSS/TES**=Distance to TSS or TES aPC
**TF**=Transcription factor aPC
**CADD**=Combined annotation dependent depletion

**RegulomeDB** = probability score (ranking)

| | |
|---|---|
| 1a | eQTL + TF binding + matched TF motif + matched DNase Footprint + DNase peak |
| 1b | eQTL + TF binding + any motif + DNase Footprint + DNase peak |
| 1c | eQTL + TF binding + matched TF motif + DNase peak |
| 1d | eQTL + TF binding + any motif + DNase peak |
| 1e | eQTL + TF binding + matched TF motif |
| 1f | eQTL + TF binding / DNase peak |
| 2a | TF binding + matched TF motif + matched DNase Footprint + DNase peak |
| 2b | TF binding + any motif + DNase Footprint + DNase peak |
| 2c | TF binding + matched TF motif + DNase peak |
| 3a | TF binding + any motif + DNase peak |
| 3b | TF binding + matched TF motif |
| 4 | TF binding + DNase peak |
| 5 | TF binding or DNase peak |
| 6 | Motif hit |
| 7 | Other |

**ChIP-seq (RegulomeDB)**: Highest probability transcription factor
